# Multi-ancestry, trans-generational GWAS meta-analysis of gestational diabetes and glycaemic traits during pregnancy reveals limited evidence of pregnancy-specific genetic effects

**DOI:** 10.1101/2025.08.19.25333735

**Authors:** Caroline Brito Nunes, Valentina Rukins, Aminata H. Cisse, Frédérique White, Nancy McBride, Alan Kuang, Catherine Allard, Justiina Ronkainen, Alice Hughes, Amanda Elliott, Gudmar Thorleifsson, Marc Vaudel, Triin Laisk, Yuqin Gu, Amel Lamri, Li Chen, Johanna Tuhkanen, Jari Lahti, Lucinda Calas, Manon Muntaner, Ville Karhunen, Jaewon Choi, Anni Heiskala, Gad Hatem, Nicolas Fragoso-Bargas, Sheryl L. Rifas-Shiman, Guillermo Paz-Lopez, Sara E. Stinson, Bishwajit Bhowmik, Emma Ahlqvist, “Estonian Biobank Research Team”, “Genes & Health Research Team”, Jean-Francois Deleuze, Johan G. Eriksson, Jongseok Park, Koon Teo, Katri Räikkönen, Kari Stefansson, Maria Molina-Vega, Padmaja Subbarao, Robin N. Beaumont, Stefan Johansson, Tiinamaija Tuomi, Torben Hansen, Line Engelbrechtsen, Sonsoles Morcillo, Emily Oken, Elisabeth Quigstad, Kåre I. Birkeland, Marja Vääräsmäki, Andrew T. Hattersley, Sylvain Sebert, Graham A Hitman, Soo Heon Kwak, Marjo-Riitta Järvelin, Rashmi B Prasad, Barbara Heude, Aline Meirhaeghe, Luigi Bouchard, Pierre-Étienne Jacques, Hannele Laivuori, Kok Hian Tan, Sonia S. Anand, David A. van Heel, Siyang Liu, Pål R. Njølstad, Valgerdur Steinthorsdottir, Elisabeth Widén, Elina Keikkala, Denise M Scholtens, William L. Lowe, Sarah Finer, Andrew P Morris, Reedik Mägi, Julia Zöllner, Maria Carolina Borges, Deborah A. Lawlor, Marie-France Hivert, Rachel M. Freathy, David M Evans, Gunn-Helen Moen

## Abstract

Gestational diabetes mellitus (GDM) affects ∼14% of pregnancies and is linked to adverse pregnancy outcomes and increased maternal type 2 diabetes mellitus (T2DM) risk. The GenDiP Consortium conducted trans-generational, multi-ancestry genome-wide association study meta-analyses of GDM and pregnancy glycemic traits in up to 38,305 GDM cases and 776,145 controls. We identified 37 GDM-associated loci (19 novel) and five novel loci for glycemic traits, all operating through the maternal genome. Most GDM loci overlapped with T2DM and non-pregnant glycemic traits, with limited evidence for pregnancy-specific effects. *MTNR1B* showed pregnancy-enhanced effects on 2-hour glucose, potentially mediated by interaction with *GPR61*, a novel GDM locus, suggesting a gestation-specific melatonin-glucose signalling axis. We also observed ancestry-specific effects at the fasting glucose locus *ABCB11*, with opposite directions in European and East Asian populations. Our findings provide new insights into the genetic architecture of GDM and highlight the need for larger, ancestrally diverse studies.

---

Gestational diabetes mellitus (GDM) affects ∼14.0% of pregnancies worldwide, and poses risks to both maternal and fetal health^1,2^. Prevalence varies across regions, with the highest rates in Middle Eastern and North African countries (27.6%) and the lowest in North America and the Caribbean (7.1%). This variation is influenced by population characteristics (e.g. ancestry and obesity rates), and differences in screening strategies and diagnostic criteria^3^.

Pregnancy induces metabolic changes essential for fetal development, including increased insulin resistance, heightened insulin secretion, and fluctuations in glucose levels^4–10^. While fasting glucose (FG) levels typically decline in the first trimester before stabilising, postprandial glucose levels tend to rise due to insulin resistance^4,8,10^. GDM arises when these adaptations are disrupted - either by excessive insulin resistance and/or insufficient insulin secretion - leading to maternal hyperglycemia^5,11,12^. While maternal glucose crosses the placenta, maternal insulin does not. In response, elevated maternal glucose levels trigger fetal insulin secretion, contributing to fetal overgrowth and potentially adverse outcomes such as obstructed labour, preterm delivery, operative delivery, stillbirth and neonatal hypoglycaemia^13^.

A previous diagnosis of GDM carries an 8-10-fold higher risk of type 2 diabetes mellitus (T2DM)^13–17^. GDM and T2DM share genetic and lifestyle risk factors^18–22^, with recent genome-wide association studies (GWASs) reporting a strong genetic correlation between the two conditions (rG ∼ 0.70), and only a few genome-wide significant loci unique to GDM^21,23^. However, while large-scale multi-ancestry GWAS of T2DM have identified hundreds of loci and included millions of individuals^24–26^, GDM GWAS have been small in comparison^21,23,27–31^. The disparity in sample size between GDM and T2DM meta-analyses limits attempts to understand whether GDM and T2DM represent the same condition (i.e. the physiological stress of pregnancy unmasking a predisposition to T2DM) or the extent to which there are distinct genetic determinants of both.

The largest and most recent GDM GWASs to date include the FinnGen study (12,332 cases; 131,109 controls; European ancestry)^23^ and the MONN study (12,024 cases; 67,845 controls; East Asian ancestry)^31^, which together identified 27 loci associated with GDM. Both these large GWASs, however, were limited to single ancestral populations, unlike the previous Genetics of Diabetes in Pregnancy (GenDiP) GWAS (5,485 cases, 347,856 controls)^21^ and this current GWAS.

Further insights into the genetic aetiology of GDM may come from GWAS of glycemic traits during pregnancy, such as FG, 1-hour (1hG) and 2-hour (2hG) post-oral glucose tolerance test (OGTT) glucose levels and glycated haemoglobin (HbA1c)^30–35^. These traits capture variation in maternal glucose regulation and may reveal loci influencing glycemic physiology not detectable through binary GDM analyses. A recent GWAS in East Asian mothers (up to 85,856 individuals) identified distinct genetic architectures for fasting versus post-OGTT glucose regulation, suggesting pregnancy-specific effects at several loci ^30,31^. However, this study was limited to East Asians, and critical gaps remain in determining whether glycemic trait regulation during pregnancy differs across populations.

In addition to maternal genetic contributions, recent studies have highlighted the role of fetal genetics in shaping the prenatal environment independently of the maternal genome. While it is expected that fetal genetic variants influence fetal traits such as birth weight and placental weight, emerging evidence suggests that the fetal genome may also influence maternal conditions, such as pre-eclampsia, gestational hypertension, and gestational duration - even after accounting for maternal genetics^36–43^. Studies also support a potential role of fetal imprinted genes in modulating maternal insulin resistance and fasting responses via placental hormone signalling^44,45^. However, studies often fail to account for the correlation between maternal and fetal genotypes, limiting conclusions about independent fetal effects on GDM.

To address these gaps, we performed the largest multi-ancestry and trans-generational GWAS meta-analyses of GDM and glycemic traits during pregnancy to date (**Fig. 1**). We further performed downstream analyses with our two largest ancestry groups - Europeans and East Asians - to elucidate the degree to which GDM and T2DM represent the same or distinct conditions and whether genetic effects on glycemic traits differ across population ns.

**Figure 1.**
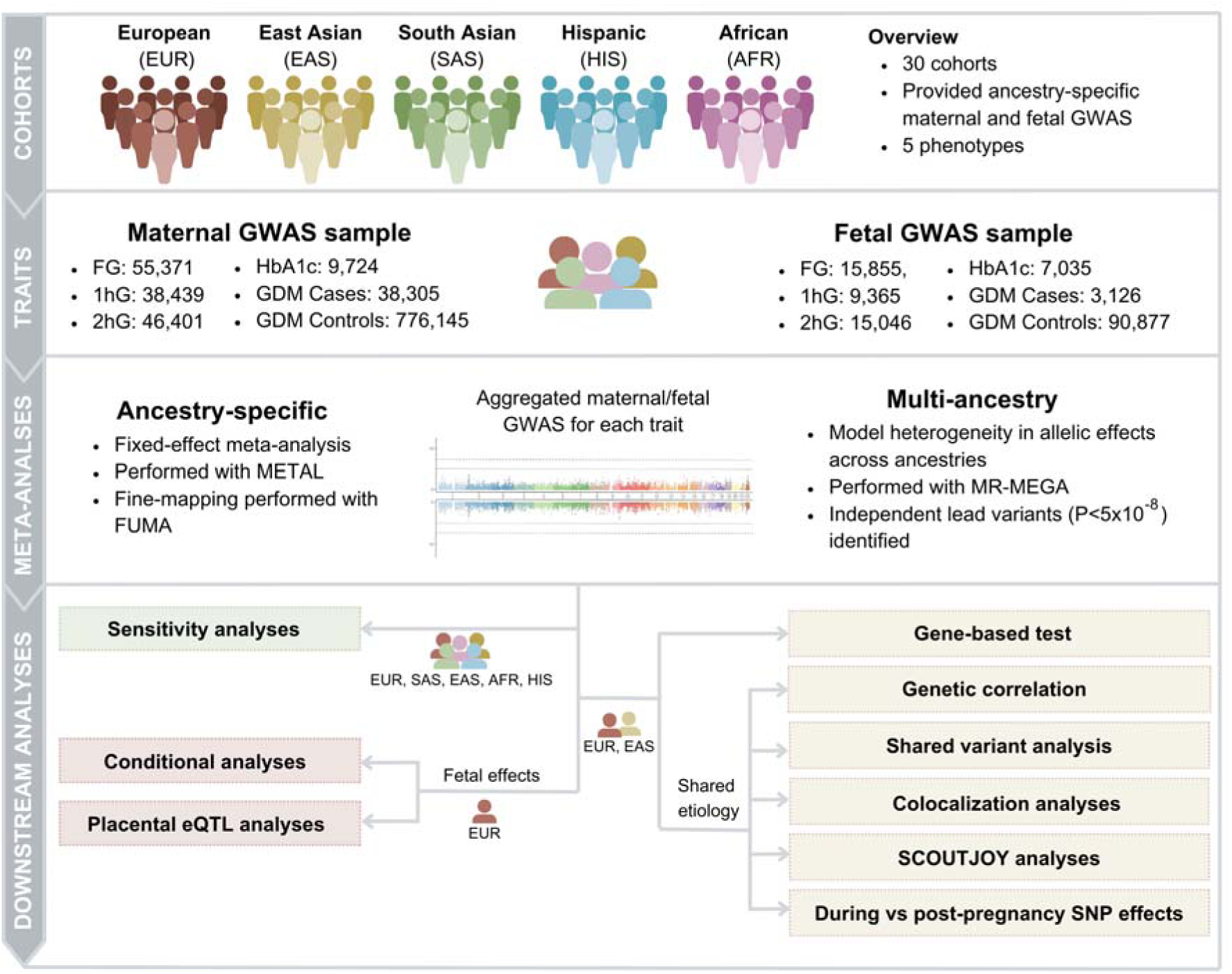
Study overview. Summary of the data resources, study sample and analyses conducted in this study. GDM; Gestational Diabetes Mellitus. FG; Fasting glucose. 1hG; 1-hour glucose after oral-glucose tolerance test (OGTT). 2hG; 2-hour glucose after OGTT. AFR: African. EAS; East Asian. EUR; European. SAS; South Asian. HIS; Hispanic.

## Results

### Discovery of maternal and fetal genetic associations in multi-ancestry meta-analyses

Our GWAS meta-analysis included data from up to thirty cohorts across European (EUR), East Asian (EAS), South Asian (SAS), admixed Africans (AFR), and admixed Hispanics (HIS) populations (**Supplementary Tables 1-3**). We conducted both ancestry-specific and multi-ancestry meta-analyses of maternal and fetal GWAS for five traits: i) GDM diagnosis (N_maternal_=38,305 cases and 776,145 controls; N_fetal_=3,126 cases and 90,877 controls), ii) FG (N_maternal_=55,371; N_fetal_=15,855), iii) 1hG post-OGTT (N_maternal_=38,439; N_fetal_=9,365), iv) 2hG post-OGTT (N_maternal_=46,401; N_fetal_=15,046) and v) HbA1c (N_maternal_=9,724; N_fetal_=7,035).

In the multi-ancestry maternal GWAS meta-analyses, we identified 37 loci associated with GDM at genome-wide significance (P<5×10^-8^), including 19 novel associations (**Fig. 2**; **Table 1**; **Supplementary Table 4**, Manhattan plot: **Supplementary Figure 1**; Plots of the axes of genetic variation separating cohorts: **Supplementary Figure 2**; Locus zoom plots: **Supplementary Figures 3-11**). We also detected several maternal loci associated with glycemic traits during pregnancy: 13 for FG, nine for 1hG, eight for 2hG, and four for HbA1c (**Supplementary Table 5;** Manhattan plots: **Supplementary Figures 12-15**; Plots of the axes of genetic variation separating cohorts: **Supplementary Figures 16-19**; Locus zoom plots: **Supplementary Figures 20-27**). Of these, five glycemic trait-locus associations were novel (*RBM47* and *IRAG2* in the FG analyses; *TTC27* in 1hG; *TCF7L2* and *ABCC8* in 2hG). Notably, several loci identified overlapped between GDM and glycemic traits (**Fig. 2**). No additional loci were identified in the ancestry-specific meta-analysis.

**Figure 2.**
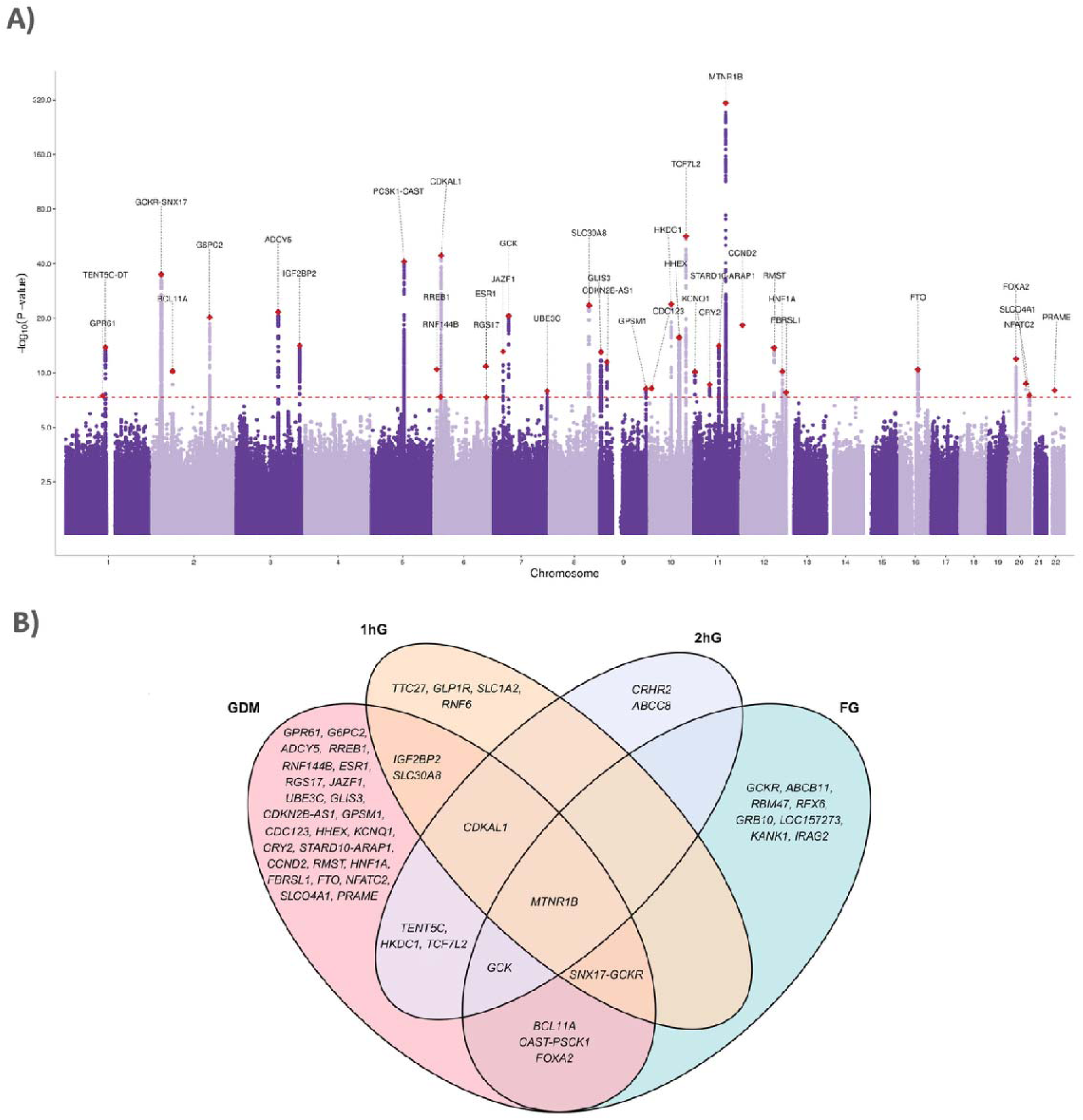
Summary of the associations for the maternal multi-ancestry GDM GWAS meta-analyses. (A) Manhattan plot for the maternal multi-ancestral GDM meta-analysis (38,305 cases and 776,145 controls), with putative gene names labelled at genome-wide significant loci. (B) Venn diagram illustrating the genetic overlap between GDM and glycemic traits reaching genome-wide significance in the maternal multi-ancestry GWAS meta-analyses. Several loci showed evidence of shared genetic architecture between GDM and glycemic traits. *MTNR1B* variants were associated with all four traits. *CDKAL1* showed overlap with GDM, 1hG, and 2hG, while shared signals at *BCL11A*, *CAST-PCSK1*, and *FOXA2* (GDM and FG), as well as *IGF2BP2* and *SLC30A8* (GDM and 1hG) were also observed. Additional loci with trait overlap included *GCK* (GDM, FG, 2hG), *SNX17-GCKR* (GDM, FG, 1hG), as well as *TCF7L2*, *HKDC1* and *TENT5C* (GDM and 2hG). Sample sizes for the GWAS of glycemic traits were as follows: FG (N=55,371), 1hG (N=38,439), and 2hG (N=46,401). GDM; Gestational Diabetes Mellitus. FG; Fasting glucose. 1hG; 1-hour glucose after oral-glucose tolerance test (OGTT). 2hG; 2-hour glucose after oral-glucose tolerance test (OGTT).

**Table 1.** Summary of the independent lead variants identified in the trans-ancestry maternal meta-analysis of GDM. The association P-value indicates the association between the variant and GDM after accounting for heterogeneity correlated with ancestry. The ancestry P-values were obtained from a test of heterogeneity in effect sizes across ancestries, whereas the residual P-values were obtained from a test of residual heterogeneity assessing the degree of variation in the data that is not captured by the principal coordinates. Asterisks indicate novel GDM loci. For the columns with the results from follow-up analyses, dashes (-) indicate variants that could not be investigated as they were not present in the GWASs used in the analysis. GDM; Gestational Diabetes Mellitus. T2DM; Type 2 Diabetes Mellitus. AFR; African. EAS; East Asian. EUR; European. SAS; South Asian. HIS; Hispanic.

| Lead Variant | CHR | POS | Putative gene | EA | NEA | EAF | Trans-ancestry meta-analysis |  |  | Significant in the ancestry-specific meta-analysis (P<0.05) | Class assigned in shared variant analyses |  |
| --- | --- | --- | --- | --- | --- | --- | --- | --- | --- | --- | --- | --- |
|  |  |  |  |  |  |  | Association P-value | Ancestry P-value | Residual P-value |  | EUR | EAS |
| rs41279738 | 1 | 109539929 | *GPR61* * | G | T | 0.959 | $3.40 \times 10^{-8}$ | $1.88 \times 10^{-2}$ | $6.58 \times 10^{-1}$ | EUR | Unspecified | - |
| rs1975283 | 1 | 117599936 | *TENT5C* * | A | C | 0.329 | $1.46 \times 10^{-14}$ | $1.45 \times 10^{-1}$ | $7.00 \times 10^{-1}$ | EUR, EAS | GDM | GDM |
| rs4665972 | 2 | 27375230 | *SNX17-GCKR* | C | T | 0.372 | $1.15 \times 10^{-35}$ | $8.82 \times 10^{-2}$ | $1.09 \times 10^{-2}$ | EUR, EAS, HIS, AFR | GDM | Unspecified |
| rs560887 | 2 | 168906638 | *G6PC2* | C | T | 0.182 | $6.60 \times 10^{-11}$ | $4.83 \times 10^{-4}$ | $1.62 \times 10^{-1}$ | EUR | GDM | Unspecified |
| rs5831573 | 2 | 60303772 | *BCL11A* * | GC | G | 0.299 | $6.28 \times 10^{-21}$ | $3.48 \times 10^{-2}$ | $8.21 \times 10^{-2}$ | EUR | - | Unspecified |
| rs72964564 | 3 | 123335923 | *ADCY5* | A | C | 0.791 | $2.31 \times 10^{-22}$ | $8.86 \times 10^{-1}$ | $6.52 \times 10^{-1}$ | EUR, SAS | Unspecified | Unspecified |
| rs113851927 | 3 | 185819848 | *IGF2BP2* * | GT | G | 0.313 | $8.13 \times 10^{-15}$ | $6.94 \times 10^{-3}$ | $9.86 \times 10^{-1}$ | EUR, EAS, SAS | - | Unspecified |
| rs10036439 | 5 | 96374664 | *CAST-PSCK1* | C | G | 0.699 | $9.06 \times 10^{-42}$ | $2.51 \times 10^{-6}$ | $1.02 \times 10^{-2}$ | EUR, EAS, AFR | GDM | GDM |
| rs9379084 | 6 | 7231610 | *RREB1* * | G | A | 0.893 | $3.51 \times 10^{-11}$ | $1.64 \times 10^{-2}$ | $7.37 \times 10^{-1}$ | EUR, SAS, AFR | T2DM | T2DM |
| rs142406759 | 6 | 18474036 | *RNF144B* * | G | GA | 0.020 | $4.24 \times 10^{-8}$ | $1.60 \times 10^{-6}$ | $2.21 \times 10^{-1}$ | EUR, HIS | - | - |
| rs9348441 | 6 | 20680447 | *CDKAL1* | T | A | 0.718 | $4.86 \times 10^{-45}$ | $2.80 \times 10^{-7}$ | $3.83 \times 10^{-2}$ | EUR, EAS, SAS, HIS | T2DM | T2DM |
| rs537224022 | 6 | 151805650 | *ESR1* | G | C | 0.011 | $1.43 \times 10^{-11}$ | $6.20 \times 10^{-1}$ | $8.99 \times 10^{-1}$ | EUR | GDM | - |
| rs7767938 | 6 | 153046478 | *RGS17\** | C | T | 0.743 | $4.65 \times 10^{-8}$ | $1.33 \times 10^{-1}$ | $9.00 \times 10^{-1}$ | EUR, EAS, SAS | Unspecified | Unspecified |
| rs849134 | 7 | 28156603 | *JAZF1\** | G | A | 0.477 | $7.81 \times 10^{-14}$ | $3.99 \times 10^{-1}$ | $1.09 \times 10^{-1}$ | EUR, EAS, SAS | T2DM | Unspecified |
| rs6975024 | 7 | 44192287 | *GCK* | T | C | 0.140 | $2.46 \times 10^{-21}$ | $1.51 \times 10^{-1}$ | $8.13 \times 10^{-2}$ | EUR, EAS, SAS, AFR | GDM | Unspecified |
| rs1182412 | 7 | 157267283 | *UBE3C\** | G | A | 0.171 | $1.13 \times 10^{-8}$ | $6.16 \times 10^{-2}$ | $1.08 \times 10^{-1}$ | EUR | Unspecified | Unspecified |
| rs11558471 | 8 | 117173494 | *SLC30A8* | G | A | 0.346 | $3.15 \times 10^{-24}$ | $1.44 \times 10^{-5}$ | $8.54 \times 10^{-1}$ | EUR | T2DM | T2DM |
| rs1574285 | 9 | 4283137 | *GLIS3* | T | G | 0.423 | $9.00 \times 10^{-14}$ | $1.21 \times 10^{-1}$ | $1.88 \times 10^{-2}$ | EUR, EAS, SAS | Unspecified | T2DM |
| rs7018475 | 9 | 22137686 | *CDKN2B-AS1* | T | G | 0.276 | $3.56 \times 10^{-12}$ | $9.02 \times 10^{-2}$ | $4.00 \times 10^{-2}$ | EUR | T2DM | Unspecified |
| rs28642213 | 9 | 136353630 | *GPSM1\** | G | A | 0.257 | $6.81 \times 10^{-9}$ | $4.25 \times 10^{-1}$ | $8.53 \times 10^{-1}$ | EUR, SAS | T2DM | T2DM |
| rs11257655 | 10 | 12265895 | *CDC123\** | T | C | 0.241 | $6.30 \times 10^{-9}$ | $2.10 \times 10^{-3}$ | $6.05 \times 10^{-1}$ | EUR, EAS, SAS | T2DM | T2DM |
| rs10762264 | 10 | 69217077 | *HKDC1* | G | A | 0.308 | $1.38 \times 10^{-24}$ | $4.56 \times 10^{-2}$ | $1.87 \times 10^{-5}$ | EUR, EAS, SAS | GDM | Unspecified |
| rs11187141 | 10 | 92707388 | *HHEX* | A | T | 0.600 | $2.28 \times 10^{-16}$ | $1.20 \times 10^{-2}$ | $1.50 \times 10^{-1}$ | EUR, EAS, SAS | T2DM | T2DM |
| rs7903146 | 10 | 112998590 | *TCF7L2* | T | C | 0.247 | $1.95 \times 10^{-57}$ | $6.96 \times 10^{-1}$ | $5.42 \times 10^{-2}$ | EUR, EAS, SAS | T2DM | T2DM |
| rs2299620 | 11 | 2837065 | *KCNQ1\** | T | C | 0.952 | $7.85 \times 10^{-11}$ | $9.04 \times 10^{-1}$ | $9.59 \times 10^{-1}$ | EUR, EAS | T2DM | T2DM |
| rs7933420 | 11 | 45875246 | *CRY2\** | A | T | 0.528 | $2.51 \times 10^{-9}$ | $3.07 \times 10^{-1}$ | $2.71 \times 10^{-1}$ | EUR | Unspecified | Unspecified |
| rs613937 | 11 | 72763794 | *STARD10-ARAP1\** | G | A | 0.204 | $8.59 \times 10^{-15}$ | $6.36 \times 10^{-2}$ | $6.90 \times 10^{-1}$ | EUR, EAS | T2DM | Unspecified |
| rs10830963 | 11 | 92975544 | *MTNR1B* | G | C | 0.304 | $5.00 \times 10^{-324}$ | $4.21 \times 10^{-15}$ | $1.42 \times 10^{-6}$ | EUR, EAS, SAS, HIS | GDM | GDM |
| rs76895963 | 12 | 4275678 | *CCND2* | T | G | 0.978 | $5.32 \times 10^{-19}$ | $9.16 \times 10^{-1}$ | $1.24 \times 10^{-1}$ | EUR | T2DM | - |
| rs77864822 | 12 | 97454997 | *RMST* | A | G | 0.923 | $1.80 \times 10^{-14}$ | $1.06 \times 10^{-2}$ | $1.30 \times 10^{-1}$ | EUR | GDM | Unspecified |
| rs7970695 | 12 | 120985573 | *HNF1A* * | A | G | 0.393 | $6.70 \times 10^{-11}$ | $8.38 \times 10^{-2}$ | $3.45 \times 10^{-1}$ | EUR, SAS | Unspecified | Unspecified |
| rs3751297 | 12 | 132511579 | *FBRSL1* | G | A | 0.709 | $1.61 \times 10^{-8}$ | $3.61 \times 10^{-1}$ | $3.64 \times 10^{-3}$ | EUR, SAS | Unspecified | Unspecified |
| rs55872725 | 16 | 53775211 | *FTO* * | C | T | 0.596 | $3.77 \times 10^{-11}$ | $9.72 \times 10^{-1}$ | $2.05 \times 10^{-1}$ | EUR, SAS | T2DM | T2DM |
| rs11087387 | 20 | 22568541 | *FOXA2* | T | C | 0.953 | $1.22 \times 10^{-12}$ | $3.54 \times 10^{-2}$ | $6.11 \times 10^{-1}$ | EUR, EAS | GDM | GDM |
| rs6021276 | 20 | 51538847 | *NFATC2* * | C | T | 0.373 | $1.94 \times 10^{-9}$ | $1.64 \times 10^{-1}$ | $9.84 \times 10^{-2}$ | EUR, SAS, AFR | GDM | Unspecified |
| rs6122141 | 20 | 62642365 | *SLCO4A1* * | C | G | 0.401 | $3.08 \times 10^{-8}$ | $1.99 \times 10^{-3}$ | $4.53 \times 10^{-1}$ | EUR, EAS | Unspecified | GDM |
| rs77090130 | 22 | 22542471 | *PRAME* | A | G | 0.014 | $9.84 \times 10^{-9}$ | $5.72 \times 10^{-9}$ | $2.67 \times 10^{-1}$ | None | Unspecified | - |

Fetal GWAS meta-analyses identified four loci associated with GDM and/or glycemic traits (Manhattan plots: **Supplementary Figures 28-32**; Plots of the axes of genetic variation separating cohorts: **Supplementary Figures 33-37)**. However, DINGO analyses conditioning on maternal genotype suggested all signals were maternally driven, likely reflecting correlations with maternal genotypes rather than independent fetal effects (**Supplementary Figures 38-45**).

In EUR, observed SNP heritability estimates ranged from 0.014 ± 0.002 (liability SNP h^2^ = 0.108 ± 0.014) for GDM to 0.1767 ± 0.085 for HbA1c, with intermediate values for FG (0.092 ± 0.025), 1hG (SNP h² = 0.056 ± 0.058), and 2hG (SNP h² = 0.076 ± 0.039) (**Supplementary Table 6**). EAS populations showed higher SNP heritability for GDM (observed SNP h² = 0.053 ± 0.021; liability SNP h^2^ = 0.119 ± 0.047) and post-load glucose traits (1hG: SNP h² = 0.144 ± 0.025; 2hG: SNP h² = 0.112 ± 0.023) compared to EUR, while FG heritability was similar (SNP h² = 0.075 ± 0.017) (**Supplementary Table 6**).

### Ancestry-specific variation in allelic effects

Our maternal multi-ancestry meta-analyses revealed significant heterogeneity in allelic effects across populations (P_ancestry_<0.05) (**Supplementary Tables 4 and 5**), with several lead variants exhibiting discordant direction and/or magnitude of effect across ancestries (**Supplementary Figures 46-55**). This was captured by the meta-regression used in MR-MEGA, which partitions heterogeneity associated with population differences (represented by principal components (PCs)) and residual sources of variation (heterogeneity not captured by PCs). These differences were most evident between the two largest ancestry groups in our study - EAS and EUR - though effect estimates in the remaining populations in our analysis were accompanied by wide confidence intervals.

For GDM, notable differences in effect estimates between EUR and EAS were observed for *IGF2BP2* (lead variant rs113851927), *CDKAL1* (rs9348441), *SLC30A8* (rs11558471), *SLCO4A1* (rs6122141) and *RMST* (rs77864822) (**Supplementary Figure 46**). Similar differences were observed for glycemic traits, including variants at *GCK* (rs730497), *LOC157273* (rs4841132), *IRAG2* (rs1683163), and *MTNR1B* (rs3847554) associated with maternal FG as well as *MTNR1B* (rs10765572) and *HKDC1* (rs5030937) detected in the maternal 1hG and 2hG analyses, respectively (**Supplementary Figures 47-49**). A striking example of ancestry-specific divergence was observed at *ABCB11* (rs2685806), where the effect on maternal FG was in opposite directions across EUR (β= -0.047, 95% CI: -0.058 to - 0.035) and EAS (β= 0.066, 95% CI: 0.051 to 0.080) populations (**Supplementary Figures 47** and 51).

### Residual heterogeneity and sensitivity analyses

We observed significant residual heterogeneity (P_residual_<0.05) for several variants associated with glycemic traits and GDM in the multi-ancestry maternal meta-analysis (**Table 1**; **Supplementary Tables 4 and 5**). This suggests that the PCs included as covariates in the meta-regression do not fully explain between-study variability, potentially due to differences in GDM screening/diagnostic criteria, OGTT load, the gestational age at which glycemic traits were measured, or other differences in the populations included.

To investigate potential sources of this variation, we conducted a sensitivity analysis, comparing cohorts based on the GDM ascertainment method to assess the contribution to residual variation in allelic effects detected in the GDM analyses. Out of 40 contributing datasets, 26 employed universal OGTT testing for GDM diagnosis (N_cases_=6,770, N_controls_=37,992), while 14 relied on risk-based testing or self-report (N_cases_=31,605, N_controls_=738,083). Among the 37 variants identified in the maternal multi-ancestry meta-analysis of GDM, only one lead variant (rs4665972 at the *GCKR-SNX17* locus) exhibited significant variability in the effect estimates between the testing approaches (P_sensitivity_=3.96×10^-4^) (**Supplementary Table 7, Supplementary Figure 56**). After accounting for the testing strategy, no significant residual heterogeneity was observed for this locus (P_residual_>0.05).

We then explored whether body mass index (BMI) could explain the opposite-direction effects observed across ancestries for maternal FG lead variant rs2685806. However, sensitivity analysis revealed no evidence for BMI-correlated heterogeneity in the allelic effects at this locus (P=0.78).

### Identification of additional genes and masking effects through gene-based testing

Given the modest SNP-based heritability estimates for GDM and glycemic traits during pregnancy, we employed gene-based association tests in EUR and EAS to capture additional signals not detected by single-variant analyses (**Supplementary Tables 8 and 9**). Although there was strong overlap between the genes identified, mBAT-combo identified additional genes not detected in the multi-ancestry GWAS meta-analyses (**Supplementary Figures 57-66; Supplementary Tables 8 and 9**).

In the EUR GDM analysis, novel genes detected included *TYR*, *SIX3*, *HLA-DQA2*, and *PLEKHA7*, all showing evidence of masking effects (i.e. significant in mBAT but not in fastBAT analyses after multiple test correction where α=0.05/n genes). Other novel GDM-associated genes detected in the EUR analysis were *KCNJ11*, *HMGA2*, *DNAJC27*, *SLC38A11*, *SLC9B1*, *FOXP4*, and *LMO1*. In EAS, novel associations included *EXOSC8* (associated with both GDM and 2hG), and *SHOX2*, *S100P*, *VPS13C*, and *MNX1* identified in the 1hG analysis, with *VPS13C* showing evidence of masking effects. For 2hG, mBAT-combo further detected *MNX1* and *HHEX* in EAS.

### Genetic correlations with T2DM, glycemic traits and pregnancy complications

To understand the shared genetic architecture between glycemic traits during and outside of pregnancy and assess the genetic overlap between these traits in pregnancy and pregnancy complications, we performed genetic correlation analyses. Our analyses revealed strong genetic correlations between diabetes-related traits in EUR (**Figure 3; Supplementary Table 10; Supplementary Figure 67**), with GDM and T2DM showing high genetic correlation (rG = 0.77, 95% CI = 0.70–0.84). GDM showed significant positive genetic correlations with most glycemic traits measured both during and outside of pregnancy, with rG ranging from 0.31 to 0.87. Notably, pregnancy measures of glycemic traits demonstrated moderate to strong genetic correlations with their counterparts outside of pregnancy. These patterns were similar, though less precise in EAS, with wider confidence intervals (**Supplementary Figure 68**). In EUR, GDM also showed positive genetic correlations with adverse pregnancy outcomes, including gestational hypertension (rG = 0.32, 95% CI = 0.23-0.42) and pre- eclampsia (rG = 0.32, 95% CI = 0.19-0.44) (**Figure 3**).

**Figure 3:**
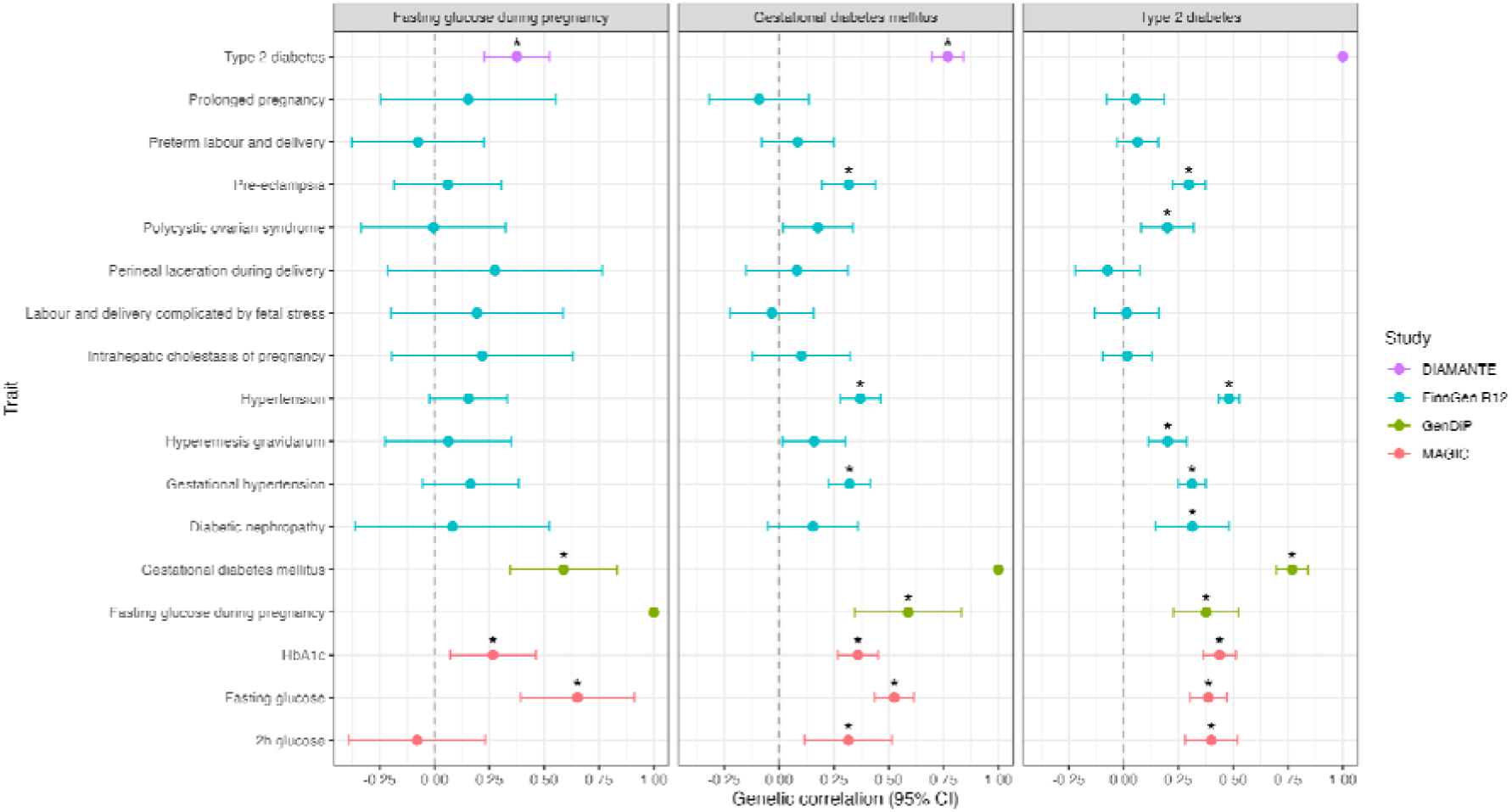
Genetic correlation analyses in Europeans. Panels display trait-specific genetic correlations for fasting glucose during pregnancy, GDM and T2DM. Each panel shows correlations with: (i) T2DM (purple; DIAMANTE consortium^71^), (ii) hypertension, adverse pregnancy outcomes and women’s reproductive health (blue; FinnGen R12^72^), (iii) pregnancy-specific glycemic traits (green; from our European GWAS meta-analyses), and (iv) glycemic trait measures outside of pregnancy (red; MAGIC consortium^46^). Genetic correlation estimates (rG) are presented as forest plots with 95% confidence intervals. Asterisks indicate statistically significant associations after false discovery rate correction (FDR < 0.05). The vertical dashed line at rG = 0 represents no genetic correlation. GDM; Gestational Diabetes Mellitus. T2DM; Type 2 Diabetes Mellitus.

### Heterogeneity in genetic effects between GDM and T2DM

We used SCOUTJOY to assess the degree of heterogeneity in genetic effects between GDM and T2DM, and between glycaemic traits measured during and outside of pregnancy using ancestry-specific effect estimates.

A significant positive relationship in genetic effects was observed between GDM and T2DM before and after outlier removal in both EUR and EAS, using either sex-combined or female-only T2DM GWAS data (**Supplementary Figures 69-72**; **Supplementary Table 11**). Despite this overall concordance, several variants exhibited significant heterogeneity in effect sizes: 16 and three outlier SNPs were identified in EUR and EAS, respectively, when using sex-combined T2DM, and 12 and three outliers when using the female-only T2DM data.

We also observed significant positive relationships between inside and outside pregnancy for FG and 2hG traits, although the strength of associations with 2hG was smaller in magnitude (**Supplementary Figure 73; Supplementary Table 11**). Similarly to the GDM analysis, we detected outlier SNPs: four (EUR) and two (EAS) in FG analyses, and two (EUR) and one (EAS) in 2hG analyses.

### Classification of GDM-associated loci into GDM- or T2DM-predominant effects

To characterise the heterogeneity in genetic effects observed between GDM and T2DM in the SCOUTJOY analyses, we classified each GDM-associated lead variant based on whether its genetic effect was stronger for GDM or T2DM in both EAS and EUR populations. The classifications were largely consistent across ancestries; however, several variants could not be classified in the EAS analyses due to imprecise effect estimates arising from smaller GWAS sample sizes (**Supplementary Table 12; Supplementary Figure 74-75**).

Across both ancestries, 12 GDM lead variants showed strong evidence of GDM-predominant effects (*TENT5C, SNX17-GCKR, G6PC2, CAST-PCSK1, ESR1, GCK, HKDC1, MTNR1B, RMST, FOXA2*, *NFATC2* and *SLCO4A1*), while 14 variants were predominantly associated with T2DM (**Figure 4**). Of the remaining variants, 10 could not be confidently classified in either population, and one (at the *RNF144B* locus) was not available in the EAS or EUR T2DM GWAS (**Figure 4**).

**Figure 4.**
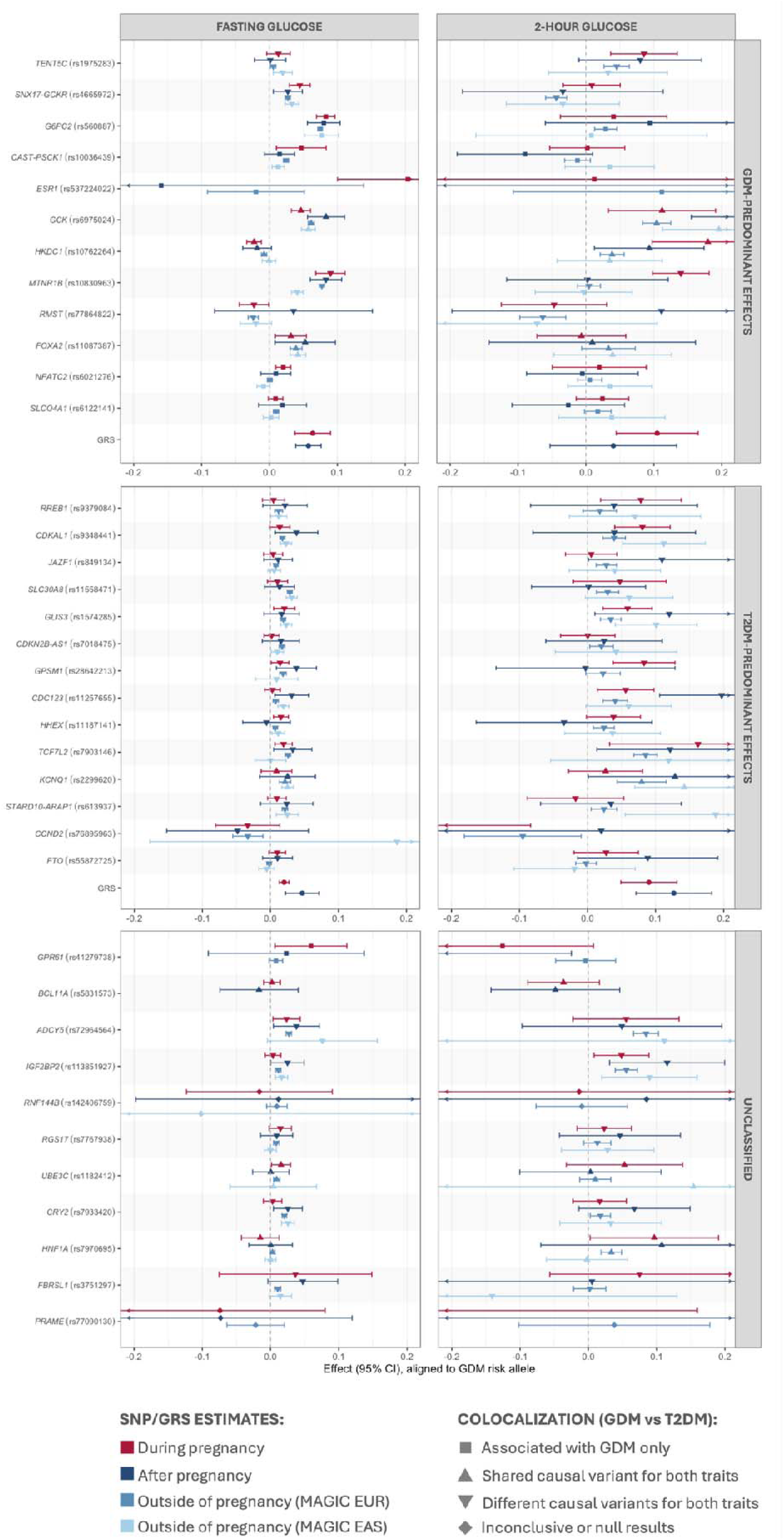
SNP and GRS associations with fasting and 2-hour glucose for GDM-associated loci. This figure presents the 37 lead SNPs identified in the maternal multi-ancestry GDM GWAS meta-analyses. Panels are vertically arranged by classification of SNPs into GDM-predominant, T2DM-predominant, and unclassified variants, based on the combined results from shared variant analyses in EUR and EAS. For each SNP, glucose effect estimates are plotted in the following order: (i) during pregnancy (N=11,117 for FG and 9,850 for 2hG), (ii) after pregnancy in previously pregnant individuals (N=14,028 for FG and 4,307 for 2hG), (iii) outside of pregnancy in individuals of European ancestry (EUR) from the MAGIC consortium (N=200,622 for FG and 63,396 for 2hG), and (iv) outside of pregnancy in individuals of East Asian ancestry (EAS) from the MAGIC consortium (N=35,619 for FG and for 8,509 2hG). The effects on fasting glucose are shown in the left panel and 2-hour glucose in the right panel, aligned to the GDM risk-increasing allele. Effect estimates during pregnancy are shown in dark red, while those after or outside of pregnancy are indicated in blue shades. Shape annotations reflect colocalization results for GDM and T2DM: squares indicate SNPs only associated with GDM, upward-pointing triangles indicate a shared causal variant between GDM and T2DM, downward-pointing triangles reflect distinct causal variants for GDM and T2DM, and diamonds represent inconclusive or null colocalization results. In general, classifications were based on the overall pattern across populations. If colocalization suggested a shared causal variant in one ancestry and was inconclusive in the other, the locus was still classified as having a shared causal variant. In the GDM- and T2DM-predominant panels, we also show glucose effects for genetic risk scores (GRS) constructed from GDM- or T2DM-predominant SNPs, respectively, with the effects of the GDM-predominant GRS (11 SNPs) and the T2DM-predominant GRS (13 SNPs) classified in EUR evaluated during and after pregnancy adjusted for principal components and maternal age. SNP and GRS effects during and after pregnancy were derived from our analyses investigating the effects of GDM-lead SNPs on glucose levels in pregnancy and postpartum, including up to ten cohorts of multiple ancestries, while estimates outside of pregnancy were obtained from the MAGIC consortium for EUR and EAS populations^46^. Arrows at the ends of confidence intervals indicate that the intervals extend beyond the plotted axis limits. GDM; Gestational Diabetes Mellitus. T2DM; Type 2 Diabetes Mellitus. SNP; Single Nucleotide Polymorphism. EUR; Europeans. EAS; East Asians. GRS; Genetic Risk Score. FG; Fasting glucose. 2hG; 2-hour glucose after oral-glucose tolerance test (OGTT).

### Colocalization analysis

We then conducted colocalization analyses to assess whether loci associated with GDM and glycemic traits during pregnancy share causal variants with T2DM and glycemic traits outside of pregnancy. Among the 37 genetic variants associated with GDM, colocalization analysis using data from EUR mothers revealed that six SNPs were associated only with GDM (*GPR61*, *G6PC2*, *CAST-PCSK1*, *ESR1*, *NFATC2*, and *SLCO4A1*), seven SNPs were associated with both GDM and T2DM but harboured distinct causal variants, 22 SNPs showed evidence of a shared causal variants with T2DM (**Figure 4**, **Supplementary Table 13; Supplementary Figures 76-112**). GDM and T2DM colocalization results were generally consistent between populations, although in EAS analyses, several loci showed inconclusive results or indicated that variants were not strongly associated with either trait.

Colocalization analyses of lead SNPs from multi-ancestry meta-analyses of glycemic traits in pregnancy - 13 for FG, eight for 2hG, and four for HbA1c - identified nine loci with shared causal variants for glycemic traits during and outside pregnancy in at least one ancestry (**Supplementary Figures 113-121**). Of these, three FG loci (*GCKR, GCK, MTNR1B*) colocalized with FG outside of pregnancy in both EUR and EAS datasets, while six loci colocalized with glycemic traits outside of pregnancy in only one ancestry: *RFX6* (FG) and *TCF7L2* (2hG) in EUR; *KANK1, FOXA2* (FG), *CDKAL1* (2hG), and *HBS1L* (HbA1c) in EAS. *ABCB11* (FG) and *GCK* (2hG) during and outside of pregnancy showed different causal variants in EUR and EAS, respectively, with inconclusive results in the other populations (**Supplementary Figures 122-123)**. Five loci showed associations only during pregnancy: *ABCC8* (2hG) in both ancestries; *RBM47* (FG) in EUR analyses; and *IRAG2*, *TENT5C*, and *CRHR2* (2hG) in EAS analyses (**Supplementary Figures 124-128**). Four loci showed ancestry-specific differences: *CAST-PCSK1*, *GRB10* (FG), and *HKDC1* (2hG) shared causal variants in EUR but were only associated with the trait during pregnancy in EAS; *LOC157273* (FG) showed distinct causal variants in EUR and associations only in pregnancy in EAS (**Supplementary Figures 129-132**). Results for the remaining variants were inconclusive in both ancestries (**Supplementary Figures 133-137**).

### Associations of GDM-associated lead SNPs with glucose levels during and after pregnancy

To further explore whether GDM-associated variants reflect pregnancy-specific disruption of glucose regulation, we compared genetic risk score (GRS) associations with FG and 2hG levels measured during and after pregnancy. We analysed data from across six multi-ancestry cohorts, including women with glucose measurements during pregnancy (N=11,117 for FG and 9,850 for 2hG) and post-pregnancy (N=14,028 for FG and 4,307 for 2hG) (**Supplementary Table 14**).

A GRS including the 37 variants associated with GDM (All-GRS) and a partitioned GRS including 11 SNPs showing GDM-predominant effects in EUR (based on shared variant analyses - **Supplementary Figure 74**; G-GRS) showed similar associations with FG levels during pregnancy and post-pregnancy. In contrast, the partitioned T2DM-predominant GRS (T-GRS; 13 SNPs based on shared variant analyses in EUR - **Supplementary Figure 74**) showed weaker associations with FG during pregnancy than post-pregnancy (**Supplementary Figure 138; Supplementary Tables 15-17**). The All-GRS and T-GRS were strongly and consistently associated with 2hG levels during pregnancy and post-pregnancy. We observed positive associations for the G-GRS with 2hG during pregnancy, and while the post-pregnancy was seemingly weaker with 95% CI crossing the null, CIs overlapped (**Supplementary Figure 139; Supplementary Tables 15-17**). The results remained consistent when adjusting for maternal age, restricting to EUR only, or only including women with glucose measurements both during and post-pregnancy and when weighting the GRSs by the T2DM effect estimates (**Supplementary Table 16**).

Leave-one-out analyses suggested that no individual study influenced the between-study heterogeneity, except for the removal of i) HAPO (AFR), which strengthened the G-GRS association with 2hG during pregnancy and removed evidence of heterogeneity, and ii) HAPO (HIS) reduced the G-GRS association with 2hG post-pregnancy and removed evidence of heterogeneity (**Supplementary Table 17**). Analyses of individual SNPs suggested greater effects on 2hG during pregnancy than post-pregnancy for the *MTNR1B* and *HKDC1* loci in class G, but power was limited (**Figure 4**).

### Expression quantitative trait locus (eQTL) Associations in the Placenta

As placental gene regulation may mediate maternal-fetal glucose dynamics, we examined whether GDM- and glycemic trait-associated variants identified in the multi-ancestry analyses were associated with placental gene expression from the Gen3G cohort (placenta biopsies from the maternal-facing side). In total, 570 SNP-gene pairs were tested, involving 408 genes within 500 Kb of at least one lead SNP. Linear regression analysis, adjusting for offspring sex, maternal genetic PCs, expression PCs and offspring genotype, revealed 16 maternal eQTLs, involving 14 SNPs and 15 genes (**Supplementary Table 18**). The strongest maternal signals included rs9379084 (RNA transcript: *SSR1*; P=2.00×10^-3^), rs4869273 (*ELL2*, P=5.80×10^-3^), rs4389513 (*SENP2*, P=6.30×10^-3^), and rs2299620 (*CDKN1C*, P=8.50×10^-3^). Of note, rs2299620 was also an eQTL for placental *KCNQ1* expression with opposite direction depending on whether the SNP was maternal versus offspring origin. None of the associations surpassed the multiple testing correction threshold (FDR < 0.05).

## Discussion

Our multi-ancestry GWAS meta-analyses identified 37 independent genome-wide significant variants associated with GDM, including 19 novel loci, as well as multiple genetic variants associated with glycemic traits during pregnancy. We found no evidence of fetal-specific genetic effects for any of the traits analysed. While most loci overlapped with T2DM and glycemic traits outside of pregnancy, a few loci showed stronger associations with GDM and context-dependent modulation of genetic effects in pregnancy.

A key strength of our study was the inclusion of diverse populations, which enabled the detection of ancestry-specific differences in allelic effects at several GDM- and glycemic trait-associated loci. A striking example is *ABCB11* (rs2685806), where the lead variant showed opposite directions of association in EUR (β=-0.047, SE= 0.006) and EAS (β=0.066, SE= 0.007) for FG during pregnancy - a pattern also observed for the same variant in FG outside of pregnancy after adjusting for body mass index (BMI), although not explicitly discussed by the authors^46^. *ABCB11* encodes the bile salt export pump (BSEP), a key transporter in hepatic bile acid secretion^47,48^. Variants in this gene have been previously implicated in cholestatic liver diseases, including intrahepatic cholestasis of pregnancy (ICP)^49,50^ - a condition which disrupts bile acid homeostasis and can impact glucose metabolism. Sensitivity analyses for FG incorporating mean cohort-level BMI as a meta-regression covariate found no evidence that BMI could account for this difference between ancestries, and imputation artifacts were unlikely given the use of distinct reference panels across cohorts. The consistency of ancestry-specific effects across cohorts also supported a genuine ancestry-related difference, perhaps reflecting differences in linkage disequilibrium (LD) structure, where the same SNP tags different causal variants across populations. Consistent with this, colocalization in EUR suggested distinct causal variants for FG during versus outside pregnancy, whereas, although not definitive, EAS analysis suggested a shared causal variant (PP=0.68). Nevertheless, population-specific gene-environment or gene-gene interactions - arising due to different environmental exposures, dietary patterns, or genetic backgrounds - could also explain these population differences.

We also observed that SNP heritability estimates for GDM and pregnancy-related glycemic traits remained modest across ancestries (e.g., EUR liability-scale SNP h² for GDM = 0.108 ± 0.014). This may be explained by differences in screening strategies and GDM diagnosis criteria across the cohorts. However, another explanation is the presence of masking effects, where LD between causal variants with opposing or reinforcing effects attenuates marginal SNP associations^51^. Supporting this, our gene-based tests identified multiple genes in EAS and EUR showing evidence of masking effects and not detected in our standard GWAS, suggesting heritability may reside partly in loci with complex LD patterns or multiple weakly associated variants, which are poorly captured by traditional GWAS and LDSC regression.

To elucidate the genetic architecture of GDM and its relationship with T2DM, we examined loci with GDM-predominant effects, integrating findings from several of our analyses (**Table 2**). Unlike previous studies that variably labelled loci as “pregnancy-specific” often without robust comparison to non-pregnant contexts, our study classifies GDM-predominant loci into five categories reflecting more precise biological interpretations: (i) loci influencing diabetes and glucose metabolism exclusively during pregnancy, (ii) glycemic trait loci with no T2DM effect, (iii) glycemic trait loci with distinct causal variants affecting GDM and T2DM, (iv) loci sharing causal variants with T2DM but with effects modified by pregnancy status, and (v) glycemic trait loci sharing causal variants with T2DM, but with evidence of effect heterogeneity across testing approaches.

**Table 2.** Classification of GDM-associated variants showing GDM-predominant effects. This table summarises the classification of lead GDM-associated variants that exhibited GDM-predominant effects based on shared variant analyses in at least one ancestry. For each locus, we report the lead gene and rsID, individual SNP effects on fasting and 2-hour glucose (from Figure 4), whether the variant was flagged as an outlier in SCOUTJOY (in either Europeans or East Asians), and whether it showed heterogeneity in allelic effects attributable to GDM diagnostic testing (from sensitivity analyses). We further indicate whether each variant reached genome-wide significance for T2DM or glycemic traits outside pregnancy (as reported in the GWAS Catalog) and present colocalization results for (i) GDM vs T2DM and (ii) glycemic traits during vs outside of pregnancy. Colocalization analyses were conducted separately in European and East Asian ancestries using lead SNPs for fasting glucose (FG) or 2-hour glucose (2hG), which may differ from the GDM lead variant; loci not detected in glycemic trait analyses were excluded from colocalization results. GDM-predominant variants were classified into categories reflecting the underlying genetic architecture: (1) **Pregnancy-specific locus** – Loci with no evidence of genome-wide association with T2DM or glycemic traits outside pregnancy (in GWAS Catalog), supported by distinct colocalization, SCOUTJOY outlier status and SNP estimates show some suggestive trend where effects on glycemic traits are greater during pregnancy. (2) **Glycemic trait loci with no T2DM effect** – Loci with no genome-wide association with T2DM, but associated with other glycemic traits outside of pregnancy (in GWAS-Catalog), supported by distinct colocalization patterns (distinct causal variants or associations with GDM only), SCOUTJOY outlier status, and SNP estimates show some suggestive trend where effects on FG and 2hG are i) greater during pregnancy or ii) confidence intervals are wide so it is possible they have greater effects during pregnancy; (3) **Glycemic trait loci with distinct causal variants affecting GDM and T2DM** – Loci at glycemic trait regions with distinct causal variants for GDM vs T2DM, SCOUTJOY outlier status and associated with glycemic traits outside of pregnancy; (4) **Loci sharing causal variants with T2DM but with effects modified by pregnancy status** – Loci at glycemic trait regions with shared causal variants between GDM and T2DM, SCOUTJOY outlier status, suggestive evidence of stronger effects in pregnancy and associated with glycemic traits outside of pregnancy and; (5) **Glycemic trait loci sharing causal variants with T2DM, but with evidence of effect heterogeneity across testing approaches** – Loci with shared causal variants between GDM and T2DM, SCOUTJOY outlier status, associated with glycemic traits outside of pregnancy and showing evidence of heterogeneity in allelic effects due to GDM diagnostic testing. GDM, Gestational Diabetes Mellitus; T2DM, Type 2 Diabetes Mellitus; FG, fasting glucose; 2hG, 2-hour glucose.

| Gene<br>(rsID) | Individual SNP effects | Outlier in<br>SCOUTJOY | Heterogeneity in<br>effect due to<br>testing<br>approaches | Genome-wide associations |  | Colocalization analyses |  | Classification |
| --- | --- | --- | --- | --- | --- | --- | --- | --- |
|  |  |  |  | T2DM | Glycemic traits<br>outside of<br>pregnancy | GDM vs T2DM | Glycemic trait during vs<br>outside of pregnancy |  |
| *TENT5C*<br>(rs1975283) | No clear pattern observed. | Yes | No | Yes | HbA1c, T2DM,<br>and random blood<br>glucose. | EUR: share causal<br>variant; EAS:<br>inconclusive results. | EUR: association with 2hG<br>only outside of pregnancy;<br>EAS: association with 2hG<br>only during pregnancy. | Unclassified. |
| *GCKR*<br>(rs4665972) | Suggestive evidence of a greater<br>effect in FG during pregnancy;<br>Suggestive evidence of a negative<br>effect on 2hG outside of<br>pregnancy, with the pregnancy<br>estimate crossing the null. | Yes | Yes | Yes | FG, random<br>glucose, T2DM,<br>HbA1c, fasting<br>insulin, 2hG. | EUR and EAS: share a<br>causal variant. | EUR and EAS: shared causal<br>variants for FG during and<br>outside of pregnancy. | Glycemic trait<br>loci sharing causal<br>variants with<br>T2DM, but with<br>evidence of effect<br>heterogeneity<br>across testing<br>approaches. |
| *G6PC2*<br>(rs560887) | No clear pattern observed. | Yes | No | No | FG, HbA1c,<br>HOMA-B. | EUR: associated with<br>GDM only; EAS:<br>inconclusive results. | - | Glycemic trait<br>loci with no<br>T2DM effect. |
| *PCSK1*<br>(rs10036439) | No clear pattern observed. | Yes | No | No | Fasting proinsulin,<br>FG. | EUR: associated with<br>GDM only; EAS:<br>inconclusive results. | EUR: shared causal variant<br>for FG during and outside of<br>pregnancy; EAS: only<br>associated with FG during<br>pregnancy. | Glycemic trait<br>loci with no<br>T2DM effect. |
| *ESR1*<br>(rs537224022) | Suggestive evidence of a stronger<br>effect in FG during pregnancy;<br>No clear pattern observed for<br>2hG. | Yes | No | Yes | HbA1c and T2DM. | EUR: associated with<br>GDM only; EAS:<br>inconclusive results. | - | Unclassified. |
| *GCK*<br>(rs6975024) | Suggestive evidence of a lower<br>effect in FG during pregnancy;<br>Suggestive evidence of a stronger<br>effect in 2hG during pregnancy. | Yes | No | Yes | Random glucose,<br>FG, HbA1c, 2hG,<br>HOMA-B, T2DM. | EUR: different causal<br>variants; EAS:<br>inconclusive results. | EUR and EAS: shared causal<br>variants for FG during and<br>outside of pregnancy. EUR:<br>associated only with 2hG<br>outside of pregnancy; EAS:<br>different causal variants. | Glycemic trait<br>loci with distinct<br>causal variants<br>affecting GDM<br>and T2DM. |
| *HKDC1*<br>(rs10762264) | CIs in FG overlap; Stronger effect in 2hG during pregnancy. | Yes | No | No | HbA1c. | EUR: different causal variants*; EAS: associated with T2DM only**. | EUR: shared causal variants for FG during and outside of pregnancy; EAS: association with 2hG only in pregnancy. | Glycemic trait loci with no T2DM effect. |
| *MTNR1B*<br>(rs10830963) | CIs in FG overlap; Stronger effect in 2hG during pregnancy. | Yes | No | Yes | HbA1c, FG, T2DM, HOMA-B, random glucose. | EUR and EAS: share a causal variant. | EUR and EAS: shared causal variants for FG during and outside of pregnancy. EUR: associated only with 2hG during pregnancy; EAS: inconclusive results. | Loci sharing causal variants with T2DM but with effects modified by pregnancy status. |
| *RMST*<br>(rs77864822) | No clear pattern observed. | No | No | Yes | HbA1c, random glucose, FG, T2DM. | EUR: share causal variant; EAS: associated with T2DM only**. | - | Unclassified. |
| *FOXA2*<br>(rs11087387) | No clear pattern observed. | Yes | No | No | FG, HbA1c. | EUR: different causal variants*; EAS: associated with T2DM only**. | EUR: associated with FG only outside of pregnancy; EAS: shared causal variant for FG during and outside of pregnancy. | Glycemic trait loci with no T2DM effect. |
| *NFATC2*<br>(rs6021276) | Suggestive evidence of greater effect on FG during pregnancy; No clear pattern observed for 2hG. | Yes | No | Yes | FG, HbA1c, T2DM. | EUR: associated with GDM only; EAS: inconclusive results. | - | Unclassified. |
| *SLCO4A1*<br>(rs6122141) | No clear pattern observed. | Yes | No | Yes | FG, T2DM. | EUR and EAS: associated with GDM only**. | - | Unclassified. |

\*Colocalization supports H<sub>0</sub> (distinct causal variants) as both traits have regional signals, but the top SNPs and regional patterns differ for the two traits.
\*\*EAS GDM GWAS yielded a weak regional signal due to small sample size, so colocalization with the larger T2DM GWAS supports association with T2DM only (H<sub>0</sub>), likely due to limited GDM power rather than absence of a true effect.

No loci met the criteria for pregnancy-specific effects (Category i), suggesting we have limited evidence of variants acting exclusively during pregnancy without influencing glycemia outside gestation. Consistent with this, our GDM-predominant GRS showed similar associations with FG and 2hG during and after pregnancy, reinforcing limited, if existing, pregnancy[specific genetic effects. However, we identified four GDM loci - *G6PC2*, *CAST- PCSK1*, *HKDC1*, and *FOXA2* - that colocalized only with GDM or harboured distinct causal variants from those affecting T2DM (Category ii). These loci lacked genome-wide associations with T2DM despite associations with other glycemic traits outside of pregnancy. Although individual SNP confidence intervals were wide, two loci showed suggestive trends toward stronger effects on glycemic traits during pregnancy: *CAST-PCSK1* in FG and *HKDC1* in 2hG. This pattern suggests that the effects of these variants may be amplified by the metabolic demands of pregnancy or may reflect differences in diagnostic thresholds between GDM and T2DM, contributing to their apparent pregnancy[specificity. Additionally, for one locus, *GCK*, we observed evidence of distinct causal variants for GDM and T2DM (Category iii). Although this gene is strongly associated with glycemic traits outside of pregnancy, our colocalization results suggest locus-level pleiotropy and potentially separate regulatory mechanisms for GDM and T2DM.

We also observed evidence of pregnancy-dependent effect modification at *MTNR1B*, a well-established locus associated with glycemic traits outside of pregnancy (Category[iv). While the causal variant is shared between GDM and T2DM, *MTNR1B* shows markedly stronger associations with 2hG during pregnancy. This pregnancy-amplified effect may be explained by its regulatory interaction with *GPR61* - a novel locus detected in our maternal multi-ancestry GDM meta-analysis - via receptor heteromerization^47^. Although *GPR61* remained unclassified in the shared variant analyses, it did not colocalize with T2DM or show genome-wide associations with glycemic traits outside of pregnancy. However, *GPR61* showed genome-wide significant associations in GWAS of anthropometric traits^53–55^, lipid biomarkers^56–58^, and metabolic syndrome^59^ - pointing to a broader metabolic role. Altogether, these findings support the existence of a gestation-specific melatonin-glucose signalling axis, where MTNR1B-GPR61 interaction potentially modulates the effect of *MTNR1B* during pregnancy.

Similar to *MTNR1B*, *GCKR* is known to influence glycemic traits outside of gestation and was shown to share a causal variant between GDM and T2DM. Although individual SNP effects appeared to indicate a potentially stronger effect in FG during pregnancy, consistent with pregnancy-dependent effect modification, our sensitivity analyses revealed substantial heterogeneity in GDM effect estimates depending on glucose testing approach (i.e. universal vs risk-based) (Category v). This suggests that clinical practice variations could account for differences in effects for this locus and reinforces the importance of standardised testing and diagnostic criteria for GDM.

Finally, several GDM-predominant loci (*TENT5C, ESR1, RMST, NFATC2, SLCO4A1*) remained unclassified due to incomplete evidence. Overall, evidence suggests that GDM-predominant loci represent a spectrum of genetic effects shaped by context, locus-level pleiotropy, and testing variability rather than uniformly “pregnancy-specific” mechanisms.

In addition to GDM-predominant loci, we identified several GDM loci with stronger T2DM effects. Colocalization analyses indicated distinct causal variants for GDM and T2DM at all but one locus - *KCNQ1*. Located in an imprinted region, *KCNQ1* harbours a likely shared causal variant for GDM and T2DM and has been previously implicated in parent-of-origin effects, where maternally inherited risk alleles show stronger effects on diabetes-related traits^60^. In our eQTL analyses, rs2299620 exhibited allele-origin-dependent effects, with maternal alleles reducing *KCNQ1* expression while fetal alleles increasing it, consistent with imprinting mechanisms and prior evidence linking *KCNQ1* methylation with maternal insulin sensitivity^61^. This interplay of opposing maternal and fetal regulatory effects at this locus may modulate its influence during pregnancy, potentially attenuating GDM effects and contributing to its stronger apparent effect in T2DM.

Our study has several limitations. First, while MR-MEGA enabled the integration of multi-ancestry genetic data, some signals (e.g. rs77090130) may reflect cohort-specific differences rather than true biological signals, as indicated by substantial heterogeneity and lack of significance in ancestry-specific analyses. Second, sample size imbalance - GDM cases predominantly EUR and glycemic traits largely powered by EAS cohorts - limited ancestry-specific discovery, stratified analyses and interpretation. Third, colocalization results should be interpreted cautiously as differences between EUR and EAS may partly reflect disparities in sample size and power, and the assumption of a single causal variant per locus may not hold in complex regions with multiple independent signals. Phenotypic heterogeneity across cohorts due to varying GDM diagnostic criteria may also have contributed to reduced power and heterogeneity in the analyses. Interpretation of HbA1c results was further limited by small sample size, pregnancy-related changes in red cells turnover, and heritability in glycation. Finally, fetal GWAS sample sizes were small, and the lack of observed fetal-specific effects may reflect limited power.

In conclusion, this multi-ancestry study provides the most comprehensive genomic analysis of GDM and pregnancy glycemic traits to date, identifying novel loci and uncovering both shared and distinct components with T2DM. Our findings demonstrate that GDM largely shares its genetic architecture with T2DM, with limited evidence for pregnancy-specific loci, while revealing ancestry-related differences in allelic effects and glycemic regulation during pregnancy. These findings underscore the importance of integrating diverse statistical approaches to refine locus interpretation and highlight the need for larger, ancestrally representative studies to guide future precision medicine approaches.

## Methods

### Study cohorts

This was a multi-ancestry meta-analysis that included thirty cohorts of EUR, EAS, SAS, AFR, and HIS ancestries. A detailed overview of cohorts can be found in **Supplementary Table 1**. All studies were approved by the relevant institutional ethics review boards.

### Traits analysed

GWAS analyses were conducted for maternal genotype data (i.e. maternal genotypes and maternal GDM diagnosis/glucose measurement) and fetal genotype data (i.e. offspring genotypes and maternal GDM diagnosis/glucose measurement). When available, individual cohorts provided GWAS on *i)* Fasting glucose (FG; mmol/l), *ii)* 1-hour post-OGTT glucose (1hG; mmol/l), *iii)* 2-hour post-OGTT glucose (2hG; mmol/l), *iv)* HbA1c (%), and *v)* GDM diagnosis (self-reported, ICD code or calculated from OGTT). Cohort contributions, phenotype definitions and individual cohort covariate adjustments can be found in **Supplementary Tables 1-3**.

### Individual cohort analyses

Individuals were genotyped in each participating study/cohort, and stringent quality control measures were applied to both samples and SNPs. Sample data were pre-phased and subsequently imputed using reference panels appropriate to each cohort’s ancestry and design. Associations between genetic variants and both glycaemic traits and GDM were assessed using additive models based on the minor allele dosage and employing a regression framework involving principal components and study-specific covariates (such as maternal age, assessment centre, etc). Studies addressed population structure concerns by either excluding related samples and incorporating PCs as additional covariates in the regression model or by including a random effect for a genetic relatedness matrix within a mixed model. Study-level quality control, pre-phasing, imputation, and GWAS analysis procedure (including covariate and analysis tools) can be found in **Supplementary Table 3**.

### Quality control of GWAS

Prior to the meta-analysis and subsequent downstream analyses, quality control was performed by the GenDiP analysts, excluding SNPs exhibiting poor imputation quality (R^2^<0.3 or info score<0.4) and/or with a minor allele count <10. If required, genetic data were transformed to align with the NCBI’s Genome Reference Consortium human build 38 (GRCh38) using SNPTracker^62^, providing a standardised reference for subsequent analyses. Further, for the multi-ancestry analyses, odds ratios (ORs) and 95% confidence intervals (CIs) were calculated from the beta coefficients and standard errors provided by the original GWAS summary statistics.

### Meta-analyses

Ancestry-specific meta-analyses were performed using METAL (v 2017)^63^, aggregating either maternal or fetal GWAS summary statistics using a fixed-effect approach with inverse-variance weights to obtain effect size estimates within each of the five broad ancestry groups.

The multi-ancestry fetal and maternal GWAS meta-analyses were performed using MR-MEGA (v 0.2)^64^. Three PCs that defined axes of genetic variation were derived and employed as covariates, distinguishing GWAS within the five ancestry groups (i.e. AFR, SAS, EAS, HIS, and EUR).

After performing the multi-ancestry and ancestry-specific meta-analyses, we removed variants that were not present in at least half of the total meta-analysis sample size and variants with a MAF of less than 1%. Manhattan plots illustrating multi-ancestry GWAS results were generated using *topr* R package (v 2.0.2)^65^. Novel variants were defined as those not previously reported in the literature and/or located >1Mb away from any previously reported variant.

### Multi-ancestry lead variants and functional mapping

From our multi-ancestry meta-analysis, independent lead variants were identified by first filtering for variants reaching genome-wide significance (P < 5×10^-8^) and then sorting these variants by chromosome and genomic position. Variants were clustered into genomic loci using a 1-megabase (Mb) proximity threshold, and within each locus, we selected the variant with the smallest P-value as the lead variant. Locus zoom plots were generated for genomic regions surrounding lead variants using *locuszoomr* R package (v0.3.8) to visualise association signals and define locus boundaries^66^. Gene track information for *Homo sapiens* (GRCh38, Ensembl v113) was obtained through *AnnotationHub* R package (v3.14.0). In case no protein-coding genes were present within ±250 kb of the lead variant, available long non-coding RNAs and microRNAs were visualised instead. Putative genes were assigned to each genomic locus based on positional proximity to lead variants.

### DINGO analyses

Maternal and offspring genetic effects on GDM, FG, 1hG, and 2hG were further analysed using the Direct and INdirect effects analysis of Genetic lOci (DINGO) software^67^. DINGO employs *i)* conditional maternal analyses (adjusted for fetal genotype), *ii)* conditional fetal analyses (adjusted for maternal genotype). Effective sample overlap between offspring and maternal GWAS was calculated using bivariate linkage disequilibrium score (LDSC) regression^68^. As LDSC regression does not perform well with small samples, DINGO was only performed with the EUR GWAS meta-analyses, as both the maternal and fetal sample sizes were the largest when compared to the other ancestry groups; However, the EUR HbA1c GWAS were too small to be included. For GDM, a binary outcome, genetic effects were transformed to the liability scale using the linear approximation before implementing DINGO^69^.

### GDM sensitivity analyses

To characterise the impact of GDM diagnostic criteria on the heterogeneity of allelic effects, we conducted a sensitivity analysis using cohort-level summary statistics of maternal GDM from 40 datasets, corresponding to 28 cohorts. We performed a meta-regression analysis using MR-MEGA (v0.2), where we employed universal diagnostic testing status (binary variable, i.e. use of universal blood-based diagnostic test) as an additional covariate alongside three principal components to account for diversity between ancestry groups. The universal diagnostic testing status was determined according to the phenotype ascertainment provided by contributing studies (**Supplementary Table 1)**. We then examined the extent of heterogeneity arising from phenotype definition among lead variants identified in the multi-ancestry meta-analysis of maternal GDM. A likelihood-ratio test with one degree of freedom was used to assess the goodness of the extended model over the baseline model. The level of significance was set at 0.05/37=1.35×10^-3^, taking into account the number of tested lead variants.

Additionally, we characterised the impact of BMI on rs2685806 (*ABCB11* locus), the lead variant from the maternal FG multi-ancestry meta-analysis exhibiting opposite-direction effects in EAS and EUR populations. We extended the baseline model containing three ancestry-specific PCs in MR-MEGA by adding the mean BMI reported for 29 cohorts. Similarly to the GDM diagnostic criteria sensitivity analysis, the effect of BMI was assessed via a one-degree-of-freedom likelihood-ratio test.

### Heritability and genetic correlations

The LD-score regression software^68^ (https://github.com/bulik/ldsc) implemented in the Complex-Traits Genetics Virtual Lab^70^ (CTG-VL, https://vl.genoma.io/) was used to estimate SNP heritability (h^2^) of GDM and glycemic traits during pregnancy in both the EUR and EAS populations. We then assessed pairwise genetic correlations (rG) of our traits with publicly available summary statistics for T2DM (DIAMANTE consortium)^71^, hypertension (FinnGen R12 for EUR^72^ and KoGES for EAS^73^), and glycemic traits outside of pregnancy^46^ (MAGIC). For the EUR analysis, we additionally looked at 8 complications of pregnancy and delivery, as well as diabetic nephropathy and polycystic ovarian syndrome, using the FinnGen publicly available dataset. The overview of traits is provided in **Supplementary Table 10**. Summary statistics were filtered to the common, well-imputed SNPs from the HapMap 3 reference panel, and variants from the HLA region were removed. Pre-computed 1000 Genomes Project EUR and EAS LD scores available on the CTG-VL platform were used. Statistical significance was corrected for a false discovery rate of 5% using *p.adjust* function in R (v4.4.2). Observed-scale SNP h^2^ of GDM and corresponding SE were transformed to the liability scale^74^, where EUR and EAS population prevalences were presumed to be 12.3% and 14.7%^1^.

### Gene-based test

Gene-based analyses were performed using mBAT-combo^51^ implemented in GCTA (v 1.94.3)^75^ to identify putative genes associated with GDM, FG, 1hG, and 2hG. mBAT-combo assesses gene-trait associations by combining mBAT and fastBAT approaches using the Cauchy combination method, enhancing the detection of gene-level associations even in the presence of masking effects (e.g. SNPs with the same direction of effect but in negative LD). These analyses were conducted separately for both EUR and EAS using population-specific LD reference panels from 1000G and against the recommended gene list (GENCODE v40). The default settings were used, with mBAT-combo assigning SNPs to genes if they fell within 50kb upstream or downstream of the gene boundaries and allowing an allele frequency difference of 0.2 between the GWAS summary and LD reference dataset. A genome-wide significant threshold was used to identify genes significantly associated with the traits analysed (α =0.05/n genes; α =2.66×10^-6^ for EUR; α =2.75×10^-6^ for EAS).

### Heterogeneity in genetic effect estimates across traits

Significant Cross-Trait Outliers and Trends in Joint York regression (SCOUTJOY) was employed to assess the relationship and compare the heterogeneity of genetic effects among glycaemic traits and diabetes-associated loci^23^. This method extends the techniques introduced in MR-PRESSO to address heterogeneity detection by considering sample overlap and estimation error in both GWAS datasets^76^. In this approach, estimators for the York regression with a fixed intercept were employed to estimate the slope of the average relationship of the variant’s effect sizes for two phenotypes. This is followed by a global heterogeneity test based on the overall goodness of fit of the York regression model to the observed distribution of effect size estimates (i.e. testing if the observed effect sizes across top hits follow a single uniform relationship). Outlier variants were detected if they presented larger than expected York regression residuals. *SCOUTJOY* was implemented in R (v0.0.5).

The lead SNPs from the multi-ancestry GWAS meta-analyses were used for the analyses comparing the heterogeneity in genetic effects between *i)* GDM and T2DM (sex-combined sample^25^ and female-only sample^77,78^ unadjusted for BMI), *ii)* FG during and outside of pregnancy (sex-combined)^46^, and *iii)* 2hG values during and outside of pregnancy (sex-combined)^46^. These analyses were conducted in both EUR and EAS; utilising SNPs identified in the multi-ancestry meta-analysis but with ancestry-specific effect sizes and standard errors (obtained from our ancestry-specific meta-analyses). Lead SNPs not present in the GWAS summary statistics downloaded or in the meta-analyses of glycemic traits conducted were excluded from the analyses. If the SNPs were not available in the datasets, we attempted to identify proxies; however, some SNPs were not present in the 1000G reference panel used in LDproxy^79^, had no variants in high LD, or did not have a proxy that reached genome-wide significance in the multi-ancestry GWAS (and was therefore not considered a suitable variant). Intercepts of the univariate and bivariate LDSC regression analyses required for SCOUTJOY were calculated before the analyses.

### Shared variant analysis

To explore whether lead SNPs identified in the multi-ancestry GDM GWAS meta-analysis exhibit genetic effects more strongly associated with T2DM or GDM, we applied the *linemodels* R-package (v.03)^80^ to summary statistics from both the GDM GWAS (Multi-ancestry: 38,241 cases and 775,169 controls; EUR: 29,625 cases and 736,101 controls; EAS: 4,064 cases and 22,276) and the T2DM GWAS in females (EUR: 30,053 cases and 434,336 controls^77^; EAS: 27,370 cases and 135,055 controls^78^). These analyses were run separately using EAS or EUR-specific GWAS. Although lead SNPs from the multi-ancestry GDM analysis were initially selected, comparisons used ancestry-specific effect estimates and standard errors; GDM SNPs absent from the T2DM dataset were excluded. The analyses ultimately included 34 GDM lead SNPs and 73 T2DM lead SNPs in the EUR analyses and 32 GDM lead SNPs and 40 T2DM lead SNPs in the EAS analyses.

Using bivariate effect sizes, SNPs were classified into two groups based on line models fitted with an Expectation-Maximization (EM) algorithm as implemented in Elliot et al., 2024^23^, producing slope estimates of 1.747 for one group (designated class T) and 0.2497 for the other (designated class G) in EUR and 1.082 for class T and 0.1443 for class G in EAS. Scale parameters for both groups were set to 0.2, while correlation parameters that defined the permissible deviation from the line were set to 0.99. Membership probabilities for each SNP in the two classes were calculated by a Bayesian classifier, assuming equal prior probability for each class. Due to the non-overlapping samples in the two GWAS datasets, the correlation of effect size estimates was set to 0. Variants were assigned to class T or G based on the highest posterior probability; those with a posterior probability ≥0.95 were labelled as "GDM" or "T2DM," while those below this threshold were categorised as "Unspecified."

### Colocalization analysis

We conducted genetic colocalization analyses using the *coloc* R package^81^ to determine whether there were shared causal variants between GDM and T2DM for each locus identified in the maternal multi-ancestry GDM GWAS meta-analysis. These analyses were restricted to the maternal meta-analysis from participants of EUR and EAS ancestry due to sample size and availability of matching ancestry summary statistics. Additionally, we performed these analyses assessing the presence of shared variants for continuous traits - specifically FG, 2hG, and HbA1c - during and outside of pregnancy. Summary data for pregnancy-related traits were derived from our meta-analyses, while publicly available data from the DIAMANTE (T2DM^71^) and MAGIC (FG, 2hG, and HbA1c^46^) consortia were used for non-pregnancy glycemic traits.

For each analysis, we extracted summary data for the relevant traits for SNPs within a +/- 500kb window from the multi-ancestry GWAS meta-analyses lead SNPs. We assumed a prior probability that any SNP in the region is associated with trait 1 only (in this case, GDM) (p1) of 1 × 10^-4^, trait 2 only (in this case, T2DM) (p2) of 1 × 10^-4^, or both traits (p12) 1 × 10^-5^. *Coloc* computes all possible configurations of causal variants for each of two traits and uses a Bayesian approach to calculate support for each causal model (H_0_: no association; H_1_: association with trait 1 only; H_2_: association with trait 2 only; H_3_: association with both traits owing to distinct causal variants; H_4_: association with both traits owing to a shared causal variant). We consider a posterior probability of association (PPA) for H_4_ > 0.7 to provide strong evidence for colocalization and PPA for H_3_ > 0.5 as evidence against colocalization, PPA for H_1_ > 0.5 as evidence for association with the pregnancy trait only, and PPA for H_2_ > 0.5 as evidence for association with the non-pregnancy trait only.

### Associations of GDM-associated lead SNPs with glucose levels during and after pregnancy

To test whether GDM-associated variants exert pregnancy-specific or consistent effects on glucose regulation, we compared the associations of genetic risk scores (GRS) with FG and 2hG levels measured both during and after pregnancy. We analysed data from 10 datasets, corresponding to six cohorts (some including multiple ancestries): EFSOCH (EUR), Gen3G (EUR), FinnGeDi (EUR), ALSPAC (EUR), HAPO (EUR, AFR, EAS, SAS, HIS), and ELGH (SAS). We analysed a total of 11,117 women with FG and 9,850 with 2hG levels during pregnancy (measured at a mean of between 24-28.4 weeks of gestation, mean maternal age ranged from 25.7 to 31.2 years at pregnancy) and 14,028 women with FG and 4,307 women with 2hG levels outside of pregnancy, with mean maternal ages across cohorts ranging from 27.0 to 47.4 years (**Supplementary Table 14**). Women were included if they did not have a diagnosis of diabetes (type 1 or 2) prior to pregnancy.

We analysed individual SNP associations but also grouped SNPs into GRSs to increase power. GRSs were based on the GDM-predominant (class G; n= 11 SNPs) and T2DM- predominant (class T; n=13 SNPs) SNP classifications derived from the shared variant analyses, and an overall GRS composed of all GDM variants identified in this study (n=37 SNPs), which additionally incorporated the 13 unclassified SNPs (G-GRS, T-GRS, and All- GRS, respectively). An individual’s GRS was calculated as the sum of the genotype dosages for each GDM increasing risk allele, weighted by its corresponding beta value (effect size, log-odds of GDM) from the European-ancestry GWAS, given in **Supplementary Table 4**.

Associations were first estimated separately within each cohort using appropriate linear regression models, adjusting for the first 4 or 5 PCs and any study-specific covariables such as genotyping batches. Results from individual cohorts were combined using an inverse-variance weighted random-effects meta-analysis. We additionally analysed associations of individual SNPs and performed sensitivity analyses (i) including only women with glucose measurements both during and after pregnancy, (ii) conducting a leave-one-out cohort meta-analysis to assess heterogeneity, (iii) performing the meta-analysis in women of EUR ancestry only, and (iv) weighting genetic scores by log-odds of T2DM (given in **Supplementary Table 12**).

### eQTL Associations in the Placenta

#### Placental Sample Collection and RNA Sequencing

We used data from the Gen3G cohort to explore associations with placental expression data. Placentas were processed within 30 minutes of delivery following standard protocols previously described in Hivert et al. (2024)^82^. A 1 cm³ sample was excised from the maternal-facing side of the placenta, approximately 5 cm from the umbilical cord insertion (if transverse). The samples were preserved in RNAlater™ solution at 4°C for 24 hours before being stored at -80°C until RNA extraction. Total RNA was extracted using the mirVana™ Paris™ Kit, and library preparation was performed with the Illumina TruSeq Stranded mRNA Sample Preparation Kit using 250 ng of RNA per sample. RNA sequencing was conducted on an Illumina HiSeq 4000 platform, generating 101-bp paired-end reads. Sequencing reads were processed following the Genotype-Tissue Expression (GTEx) V8 pipeline^83^, with alignment performed using the hg38 reference genome and gene quantification based on GENCODE V30 annotations, using RNASeQC v2.3.6. The Gen3G placental RNA-seq dataset is available on dbGaP^84^.

### Gene Expression Normalisation and Covariate Adjustment

To process gene expression data for quantitative trait locus (eQTL) discovery, we followed the approach used by the GTEx project by applying rimmed Mean of M-values (TMM) normalisation to expression data from 58,011 genes in a subset of 375 European-ancestry samples with both placental RNA-seq data and maternal genotype information available^85^. Genes with low abundance were filtered out, retaining 20,578 genes with a minimum read count of six and transcript per million (TPM) values > 0.1 in at least 20% of samples^85^. An inverse normal transformation was then applied to each gene across samples^85^. To account for unmeasured sources of variation, we performed principal component analysis (PCA) on the normalised expression data. The first seven PCs, which primarily captured variance attributable to placenta cell-type composition as estimated through cell deconvolution, were retained as covariates.

### eQTL Analysis

eQTL associations were evaluated within a 500 Kb window upstream and downstream of each lead SNP (1 Mb total). Linear regression models were used to test associations between each SNP and nearby genes, with gene expression as the dependent variable and the following covariates: offspring sex, five maternal genetic PCs, and the seven expression PCs. To assess the potential contribution of offspring genotype to eQTL effects, we repeated the analysis in a subset of 267 samples with available offspring genotypes (from whole genome sequencing). An eQTL was considered under maternal genotype control if the maternal SNP-eQTL association had a p-value < 0.05 in the model adjusting for offspring genotype and if the term for the offspring genotype SNP-eQTL in the adjusted model was not significant (p-value > 0.05).

## Data availability

GWAS summary statistics are available via the University of Queensland eSpace repository: maternal multi-ancestry GWAS (https://espace.library.uq.edu.au/view/UQ:0aa6ba4), maternal ancestry-specific GWAS (EUR/EAS; https://espace.library.uq.edu.au/view/UQ:2e6c470) and DINGO analyses (https://espace.library.uq.edu.au/view/UQ:ed470a2).

## Code availability

Genetic data were transformed to align GRCh38 using SNPTracker (http://pmglab.top/snptracker/).

Multi-ancestry meta-analyses were conducted with MR-MEGA v0.2 (https://genomics.ut.ee/en/tools). Ancestry-specific meta-analyses were performed with METAL v2017 (https://github.com/statgen/METAL).

mBAT-combo (https://github.com/Share-AL-work/mBAT.combo) was implemented in GCTA v1.94.3 (https://yanglab.westlake.edu.cn/software/gcta/#Overview).

Maternal and fetal conditional analyses were conducted using the DINGO software (https://github.com/danielldhwang/DINGO) implemented in the CTG-VL platform (https://vl.genoma.io/).

Functional mapping and annotation of ancestry-specific signals was performed with FUMA v1.7.0 (https://fuma.ctglab.nl/).

Genetic correlation and heritability estimates were performed using LDSC v1.0.1 (https://github.com/bulik/ldsc) in the CTG-VL platform (https://vl.genoma.io/).

Comparison of heterogeneity of cross-trait genetic effects was performed using SCOUTJOY v0.0.5 (https://github.com/aelliott08/SCOUTJOY). Classification of variants based on their bivariate effect size was carried out using Linemodels 0.2.0 (https://github.com/mjpirinen/linemodels).

Colocalization analyses were performed with the coloc package implemented in R (https://github.com/chr1swallace/coloc).

eQTL analyses were carried out following GTEx steps for normalisation ( https://github.com/broadinstitute/gtex-pipeline/tree/master/qtl) and using custom R scripts to test associations (https://github.com/whitef19/conditional-eQTL)

Meta-analyses of lead SNPs were conducted in R using the implemented package “metafor” (https://wviechtb.github.io/metafor/)

## Supporting information

Cohort funding, approval and acknowledgents

## Author contributions

CBN, VR, AHC and FW contributed equally to this work. CBN and GHM conceived the study. CBN, VR, AHC, FW, NMB, AH, SF, APM, RM, JZ, MCB, DAL, MFH, RMF, DME, GHM designed the study and were a part of the analysis and writing group of this paper. CBN, VR, AHC, FW, NMB and MCB performed the meta-analysis and/or follow-up analysis. CBN, VR, AHC, FW, NMB, AK, CA, JR, AE, GT, MV, TL, YG, AL, LC, JT, JL, LC, MM, VK, JC, AH, GH, NFB, SLRS, GPL, SES, BB, EA, JFD, JGE, JP, KT, KR, KS, MMV, PS, RNB, SJ, TT, TH, LE, SM, EO, EQ, KIB, MV, ATH, SS, GAH, SHK, MRJ, RBP, BH, AM, LB, PEJ, HL, KHT, SSA, DAvH, SL, PRN, VS, EW, EK, DMS, WLL, SF, RM, JZ, MCB, DAL, MFH, RMF, DME and GHM provided cohort level analysis and/or contributed to gathering, processing and curating data. MFH, RMF, DME, and GHM jointly supervised the project. All authors had access to all the data in the study. All authors contributed to the review and the editing of the paper. All authors approved the paper before submission.

## References

1. Wang, H. et al. IDF Diabetes Atlas: Estimation of Global and Regional Gestational Diabetes Mellitus Prevalence for 2021 by International Association of Diabetes in Pregnancy Study Group’s Criteria. Diabetes Res Clin Pract 183, 109050 (2022).

2. McIntyre, H. D. et al. Gestational diabetes mellitus. Nat Rev Dis Primers 5, 47 (2019).

3. Wang, H. et al. IDF Diabetes Atlas: Estimation of Global and Regional Gestational Diabetes Mellitus Prevalence for 2021 by International Association of Diabetes in Pregnancy Study Group’s Criteria. Diabetes Res Clin Pract 183, 109050 (2022).

4. Mills, J. L. et al. Physiological reduction in fasting plasma glucose concentration in the first trimester of normal pregnancy: the diabetes in early pregnancy study. Metabolism 47, 1140–1144 (1998).

5. Buchanan, T. A., Metzger, B. E., Freinkel, N. & Bergman, R. N. Insulin sensitivity and B-cell responsiveness to glucose during late pregnancy in lean and moderately obese women with normal glucose tolerance or mild gestational diabetes. Am J Obstet Gynecol 162, 1008–1014 (1990).

6. Sorenson, R. L. & Brelje, T. C. Adaptation of islets of Langerhans to pregnancy: beta-cell growth, enhanced insulin secretion and the role of lactogenic hormones. Horm Metab Res 29, 301–307 (1997).

7. Metzger, B. E., Phelps, R. L., Freinkel, N. & Navickas, I. A. Effects of Gestational Diabetes on Diurnal Profiles of Plasma Glucose, Lipids, and Individual Amino Acids. Diabetes Care 3, 402– 409 (1980).

8. Frøslie, K. F. et al. Shape information in repeated glucose curves during pregnancy provided significant physiological information for neonatal outcomes. PLoS One 9, e90798 (2014).

9. Catalano, P. M., Roman-Drago, N. M., Amini, S. B. & Sims, E. A. Longitudinal changes in body composition and energy balance in lean women with normal and abnormal glucose tolerance during pregnancy. Am J Obstet Gynecol 179, 156–165 (1998).

10. Dratver, M. A. B. et al. Longitudinal changes in glucose during pregnancy in women with gestational diabetes risk factors. Diabetologia 65, 541–551 (2022).

11. Butler, A. E. et al. Adaptive changes in pancreatic beta cell fractional area and beta cell turnover in human pregnancy. Diabetologia 53, 2167–2176 (2010).

12. Powe, C. E. et al. Heterogeneous Contribution of Insulin Sensitivity and Secretion Defects to Gestational Diabetes Mellitus. Diabetes Care 39, 1052–1055 (2016).

13. Song, C. et al. Long-term risk of diabetes in women at varying durations after gestational diabetes: a systematic review and meta-analysis with more than 2 million women. Obes Rev 19, 421–429 (2018).

14. Li, Z. et al. Incidence Rate of Type 2 Diabetes Mellitus after Gestational Diabetes Mellitus: A Systematic Review and Meta-Analysis of 170,139 Women. J Diabetes Res 2020, 3076463 (2020).

15. Dennison, R. A. et al. The absolute and relative risk of type 2 diabetes after gestational diabetes: A systematic review and meta-analysis of 129 studies. Diabetes Res Clin Pract 171, 108625 (2021).

16. Herath, H., Herath, R. & Wickremasinghe, R. Gestational diabetes mellitus and risk of type 2 diabetes 10 years after the index pregnancy in Sri Lankan women-A community based retrospective cohort study. PLoS One 12, e0179647 (2017).

17. Vounzoulaki, E. et al. Progression to type 2 diabetes in women with a known history of gestational diabetes: systematic review and meta-analysis. BMJ 369, m1361 (2020).

18. Gadve, S. S. et al. Risk of Developing Type 2 Diabetes Mellitus in South Asian Women with History of Gestational Diabetes Mellitus: A Systematic Review and Meta-Analysis. Indian Journal of Endocrinology and Metabolism 25, 176 (2021).

19. Dennison, R. A. et al. The absolute and relative risk of type 2 diabetes after gestational diabetes: A systematic review and meta-analysis of 129 studies. Diabetes Research and Clinical Practice 171, 108625 (2021).

20. Monod, C. et al. Prevalence of gestational diabetes mellitus in women with a family history of type 2 diabetes in first- and second-degree relatives. Acta Diabetol 60, 345–351 (2023).

21. Pervjakova, N. et al. Multi-ancestry genome-wide association study of gestational diabetes mellitus highlights genetic links with type 2 diabetes. Hum Mol Genet 31, 3377–3391 (2022).

22. Borges, M. C. et al. Integrating multiple lines of evidence to assess the effects of maternal BMI on pregnancy and perinatal outcomes. BMC Med 22, 32 (2024).

23. Elliott, A. et al. Distinct and shared genetic architectures of gestational diabetes mellitus and type 2 diabetes. Nat Genet 56, 377–382 (2024).

24. Vujkovic, M. et al. Discovery of 318 new risk loci for type 2 diabetes and related vascular outcomes among 1.4 million participants in a multi-ancestry meta-analysis. Nat Genet 52, 680–691 (2020).

25. Mahajan, A. et al. Multi-ancestry genetic study of type 2 diabetes highlights the power of diverse populations for discovery and translation. Nat Genet 54, 560–572 (2022).

26. Suzuki, K. et al. Multi-ancestry genome-wide study in >2.5 million individuals reveals heterogeneity in mechanistic pathways of type 2 diabetes and complications. medRxiv 2023.03.31.23287839 (2023) doi:10.1101/2023.03.31.23287839.

27. Kwak, S. H. et al. A genome-wide association study of gestational diabetes mellitus in Korean women. Diabetes 61, 531–541 (2012).

28. Yue, S. et al. Genome-wide analysis study of gestational diabetes mellitus and related pathogenic factors in a Chinese Han population. BMC Pregnancy Childbirth 23, 856 (2023).

29. Wu, N.-N. et al. A genome-wide association study of gestational diabetes mellitus in Chinese women. J Matern Fetal Neonatal Med 34, 1557–1564 (2021).

30. Zhen, J. et al. Genome-wide association and Mendelian randomisation analysis among 30,699 Chinese pregnant women identifies novel genetic and molecular risk factors for gestational diabetes and glycaemic traits. Diabetologia 67, 703–713 (2024).

31. Gu, Y. et al. Genetic architecture and risk prediction of gestational diabetes mellitus in Chinese pregnancies. Nat Commun 16, 4178 (2025).

32. Zhu, H. et al. Novel insights into the genetic architecture of pregnancy glycemic traits from 14,744 Chinese maternities. Cell Genomics 4, (2024).

33. Lowe, W. L., Kuang, A., Hayes, M. G., Hivert, M.-F. & Scholtens, D. M. Genetics of glucose homeostasis in pregnancy and postpartum. Diabetologia 67, 2726–2739 (2024).

34. Kuang, A., Hivert, M.-F., Hayes, M. G., Lowe, W. L. & Scholtens, D. M. Multi-ancestry genome-wide association analyses: a comparison of meta- and mega-analyses in the Hyperglycemia and Adverse Pregnancy Outcome (HAPO) study. BMC Genomics 26, 65 (2025).

35. Hayes, M. G. et al. Identification of HKDC1 and BACE2 as genes influencing glycemic traits during pregnancy through genome-wide association studies. Diabetes 62, 3282–3291 (2013).

36. Warrington, N. M. et al. Maternal and fetal genetic effects on birth weight and their relevance to cardio-metabolic risk factors. Nat Genet 51, 804–814 (2019).

37. Beaumont, R. N. et al. Genome-wide association study of offspring birth weight in 86L577 women identifies five novel loci and highlights maternal genetic effects that are independent of fetal genetics. Hum Mol Genet 27, 742–756 (2018).

38. Juliusdottir, T. et al. Distinction between the effects of parental and fetal genomes on fetal growth. Nat Genet 53, 1135–1142 (2021).

39. Beaumont, R. N. et al. Genome-wide association study of placental weight identifies distinct and shared genetic influences between placental and fetal growth. Nat Genet 55, 1807–1819 (2023).

40. Chen, J. et al. Dissecting maternal and fetal genetic effects underlying the associations between maternal phenotypes, birth outcomes, and adult phenotypes: A mendelian-randomization and haplotype-based genetic score analysis in 10,734 mother-infant pairs. PLoS Med 17, e1003305 (2020).

41. Solé-Navais, P. et al. Genetic effects on the timing of parturition and links to fetal birth weight. Nat Genet 55, 559–567 (2023).

42. McGinnis, R. et al. Variants in the fetal genome near FLT1 are associated with risk of preeclampsia. Nat Genet 49, 1255–1260 (2017).

43. Steinthorsdottir, V. et al. Genetic predisposition to hypertension is associated with preeclampsia in European and Central Asian women. Nat Commun 11, 5976 (2020).

44. Cleaton, M. A. M. et al. Fetus-derived DLK1 is required for maternal metabolic adaptations to pregnancy and is associated with fetal growth restriction. Nat Genet 48, 1473–1480 (2016).

45. Lopez-Tello, J. et al. Fetal manipulation of maternal metabolism is a critical function of the imprinted Igf2 gene. Cell Metab 35, 1195–1208.e6 (2023).

46. Chen, J. et al. The trans-ancestral genomic architecture of glycemic traits. Nat Genet 53, 840– 860 (2021).

47. Dean, M., Rzhetsky, A. & Allikmets, R. The human ATP-binding cassette (ABC) transporter superfamily. Genome Res 11, 1156–1166 (2001).

48. Soroka, C. J. & Boyer, J. L. Biosynthesis and trafficking of the bile salt export pump, BSEP: therapeutic implications of BSEP mutations. Mol Aspects Med 37, 3–14 (2014).

49. Dixon, P. H. et al. GWAS meta-analysis of intrahepatic cholestasis of pregnancy implicates multiple hepatic genes and regulatory elements. Nat Commun 13, 4840 (2022).

50. Liu, Y. et al. Genetic study of intrahepatic cholestasis of pregnancy in Chinese women unveils East Asian etiology linked to historic HBV epidemic. Journal of Hepatology 82, 826–835 (2025).

51. Li, A. et al. mBAT-combo: A more powerful test to detect gene-trait associations from GWAS data. Am J Hum Genet 110, 30–43 (2023).

52. Oishi, A. et al. Orphan GPR61, GPR62 and GPR135 receptors and the melatonin MT2 receptor reciprocally modulate their signaling functions. Sci Rep 7, 8990 (2017).

53. Huang, J. et al. Genomics and phenomics of body mass index reveals a complex disease network. Nat Commun 13, 7973 (2022).

54. Verma, A. et al. Diversity and scale: Genetic architecture of 2068 traits in the VA Million Veteran Program. Science 385, eadj1182 (2024).

55. Shungin, D. et al. New genetic loci link adipose and insulin biology to body fat distribution. Nature 518, 187–196 (2015).

56. Sinnott-Armstrong, N. et al. Genetics of 35 blood and urine biomarkers in the UK Biobank. Nat Genet 53, 185–194 (2021).

57. Richardson, T. G. et al. Evaluating the relationship between circulating lipoprotein lipids and apolipoproteins with risk of coronary heart disease: A multivariable Mendelian randomisation analysis. PLOS Medicine 17, e1003062 (2020).

58. Koskeridis, F. et al. Pleiotropic genetic architecture and novel loci for C-reactive protein levels. Nat Commun 13, 6939 (2022).

59. Park, S. et al. Multivariate genomic analysis of 5 million people elucidates the genetic architecture of shared components of the metabolic syndrome. Nat Genet 56, 2380–2391 (2024).

60. Hanson, R. L. et al. Strong Parent-of-Origin Effects in the Association of KCNQ1 Variants With Type 2 Diabetes in American Indians. Diabetes 62, 2984–2991 (2013).

61. Hivert, M.-F. et al. Interplay of Placental DNA Methylation and Maternal Insulin Sensitivity in Pregnancy. Diabetes 69, 484–492 (2020).

62. Deng, J.-E., Sham, P. C. & Li, M.-X. SNPTracker: A Swift Tool for Comprehensive Tracking and Unifying dbSNP rs IDs and Genomic Coordinates of Massive Sequence Variants. G3 (Bethesda) 6, 205–207 (2015).

63. Willer, C. J., Li, Y. & Abecasis, G. R. METAL: fast and efficient meta-analysis of genomewide association scans. Bioinformatics 26, 2190–2191 (2010).

64. Mägi, R. et al. Trans-ethnic meta-regression of genome-wide association studies accounting for ancestry increases power for discovery and improves fine-mapping resolution. Hum Mol Genet 26, 3639–3650 (2017).

65. Juliusdottir, T. topr: an R package for viewing and annotating genetic association results. BMC Bioinformatics 24, 268 (2023).

66. Lewis, M. J. & Wang, S. locuszoomr: an R package for visualizing publication-ready regional gene locus plots. Bioinform Adv 5, vbaf006 (2025).

67. Hwang, L.-D. et al. DINGO: increasing the power of locus discovery in maternal and fetal genome-wide association studies of perinatal traits. Nat Commun 15, 9255 (2024).

68. Bulik-Sullivan, B. K. et al. LD Score regression distinguishes confounding from polygenicity in genome-wide association studies. Nat Genet 47, 291–295 (2015).

69. Wu, T. & Sham, P. C. On the Transformation of Genetic Effect Size from Logit to Liability Scale. Behav Genet 51, 215–222 (2021).

70. Cuéllar-Partida, G. et al. Complex-Traits Genetics Virtual Lab: A community-driven web platform for post-GWAS analyses. 518027 Preprint at 10.1101/518027 (2019).

71. Mahajan, A. et al. Multi-ancestry genetic study of type 2 diabetes highlights the power of diverse populations for discovery and translation. Nat Genet 54, 560–572 (2022).

72. Kurki, M. I. et al. FinnGen provides genetic insights from a well-phenotyped isolated population. Nature 613, 508–518 (2023).

73. Nam, K., Kim, J. & Lee, S. Genome-wide study on 72,298 individuals in Korean biobank data for 76 traits. Cell Genom 2, 100189 (2022).

74. Lee, S. H., Wray, N. R., Goddard, M. E. & Visscher, P. M. Estimating missing heritability for disease from genome-wide association studies. Am J Hum Genet 88, 294–305 (2011).

75. Yang, J., Lee, S. H., Goddard, M. E. & Visscher, P. M. GCTA: A Tool for Genome-wide Complex Trait Analysis. Am J Hum Genet 88, 76–82 (2011).

76. Verbanck, M., Chen, C.-Y., Neale, B. & Do, R. Detection of widespread horizontal pleiotropy in causal relationships inferred from Mendelian randomization between complex traits and diseases. Nat Genet 50, 693–698 (2018).

77. Mahajan, A. et al. Fine-mapping type 2 diabetes loci to single-variant resolution using high-density imputation and islet-specific epigenome maps. Nat Genet 50, 1505–1513 (2018).

78. Spracklen, C. N. et al. Identification of type 2 diabetes loci in 433,540 East Asian individuals. Nature 582, 240–245 (2020).

79. Machiela, M. J. & Chanock, S. J. LDlink: a web-based application for exploring population-specific haplotype structure and linking correlated alleles of possible functional variants. Bioinformatics 31, 3555–3557 (2015).

80. Pirinen, M. linemodels: clustering effects based on linear relationships. Bioinformatics 39, btad115 (2023).

81. Foley, C. N. et al. A fast and efficient colocalization algorithm for identifying shared genetic risk factors across multiple traits. Nat Commun 12, 764 (2021).

82. Hivert, M.-F. et al. Placental IGFBP1 levels during early pregnancy and the risk of insulin resistance and gestational diabetes. Nat Med 30, 1689–1695 (2024).

83. Broad Institute. broadinstitute/gtex-pipeline. Broad Institute (2018).

84. Gen3G. dbGaP Study. https://www.ncbi.nlm.nih.gov/projects/gap/cgi-bin/study.cgi?study_id=phs003151.v1.p1.

85. Robinson, M. D. & Oshlack, A. A scaling normalization method for differential expression analysis of RNA-seq data. Genome Biology 11, R25 (2010).

