## Supplementary Figures for "Multi-ancestry, trans-generational GWAS meta-analysis of gestational diabetes and glycaemic traits during pregnancy reveals limited evidence of pregnancy-specific genetic effects"

**Table of Contents**

**1.0 Maternal GWAS meta-analyses	3**

1.2 Axes of genetic variation separating GWAS of GDM in MR-MEGA	4

1.3 Locus ZoomPlots for the meta-analysis of GDM	5

1.4 Multi-ancestry GWAS meta-analyses of the glycemic traits	14

1.5 Axes of genetic variation separating GWAS of glycemic traits in MR-MEGA	18

1.6 Locus ZoomPlots for the meta-analyses of glycemic traits	22

**2.0 Fetal GWAS meta-analyses	30**

2.1 Multi-ancestry GWAS meta-analyses	30

2.2 Axes of genetic variation separating GWAS	33

2.3 Conditional analyses using DINGO	38

**3.0 Heterogeneity in allelic effects in the maternal multi-ancestry meta-analyses	42**

3.1 Ancestry-specific variation in allelic effects	42

3.2 Associations driven by study heterogeneity	48

3.3 GDM diagnostic test sensitivity analysis	52

**4.0 Gene-based test	53**

4.1 European analyses	53

4.2 East Asian analyses	57

4.3 Genetic overlap between phenotypes	61

**5.0 Genetic correlation analyses	63**

**6.0 SCOUTJOY analyses	64**

**7.0 Shared variant analyses	71**

**8.0 Colocalization analyses	73**

8.1 Genetic signal for GDM only	73

8.2 Different causal variants for GDM and T2DM	79

8.3 Shared causal variants for GDM and T2DM	86

8.4 No association with GDM or T2DM	108

8.5 Shared causal variants for glycemic traits during and outside of pregnancy	110

8.6 Different causal variants for glycemic traits during and outside of pregnancy	119

8.7 Association with glycemic traits only in pregnancy	121

8.8 Different colocalization results in EAS and EUR	126

8.9 No association with trait during or outside of pregnancy	130

**9.0 GDM GRS associations with glucose levels during and after pregnancy	135**

### **1.0 Maternal GWAS meta-analyses**

#### 1.1 Multi-ancestry GDM GWAS meta-analysis

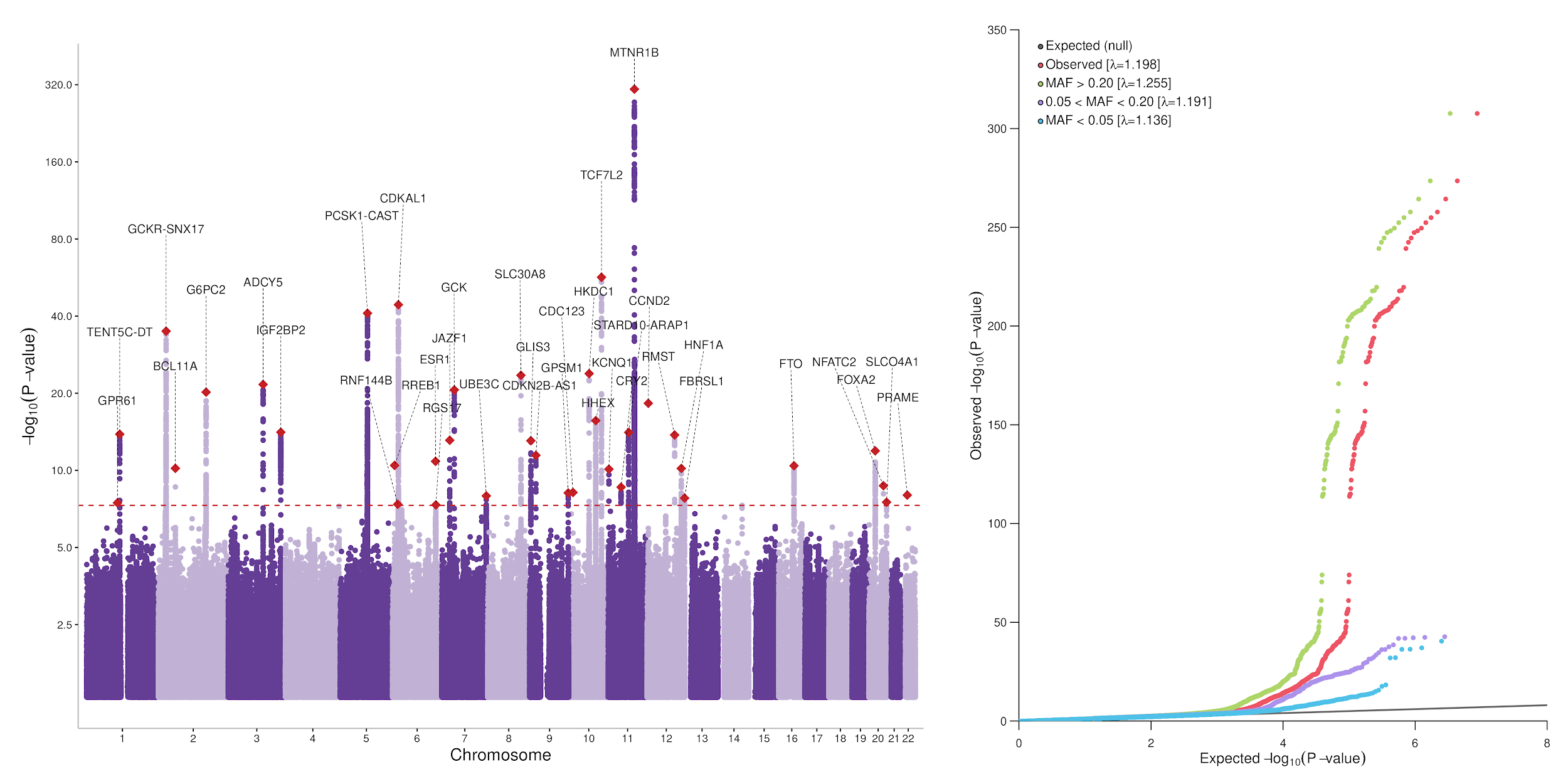

**Supplementary Figure 1.** *Manhattan plot and QQ plot of the results for the GWAS meta-analysis of maternal GDM including 38,305 cases and 776,145 controls.* Only SNPs with P-value < 0.05 are plotted on the Manhattan plot. The horizontal red dashed line represents genome-wide significance (P=5$\times$10^-8^). GDM; Gestational Diabetes Mellitus. MAF; minor allele frequency.

#### 1.2 Axes of genetic variation separating GWAS of GDM in MR-MEGA

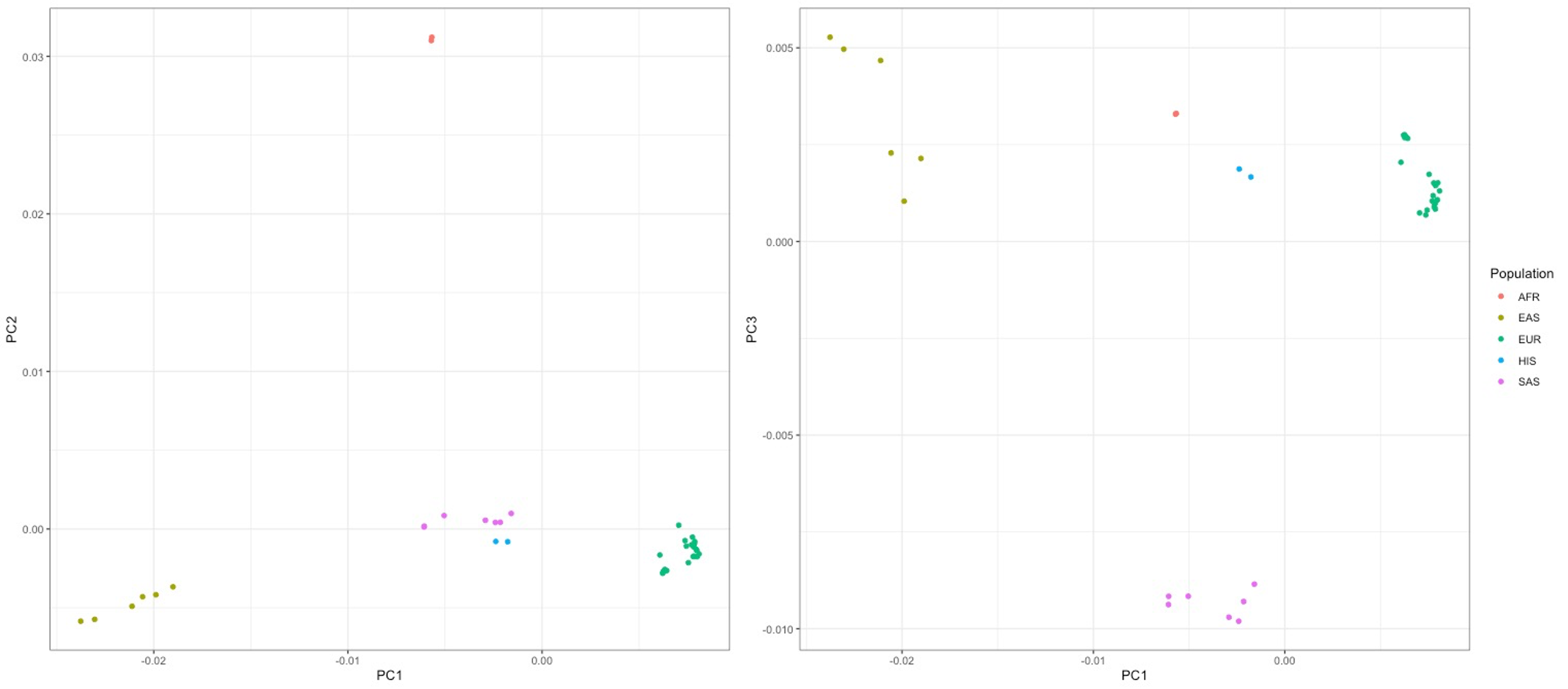

**Supplementary Figure 2.** *Axes of genetic variation separating maternal GWAS of GDM.* The first three axes of genetic variation from multi-dimensional scaling of the Euclidean distance matrix between GWAS are sufficient to separate five ancestry groups: South Asians (SAS), African American (AFR), Hispanic/Latino (HIS), East Asian (EAS) and European (EUR).

#### 1.3 Locus ZoomPlots for the meta-analysis of GDM

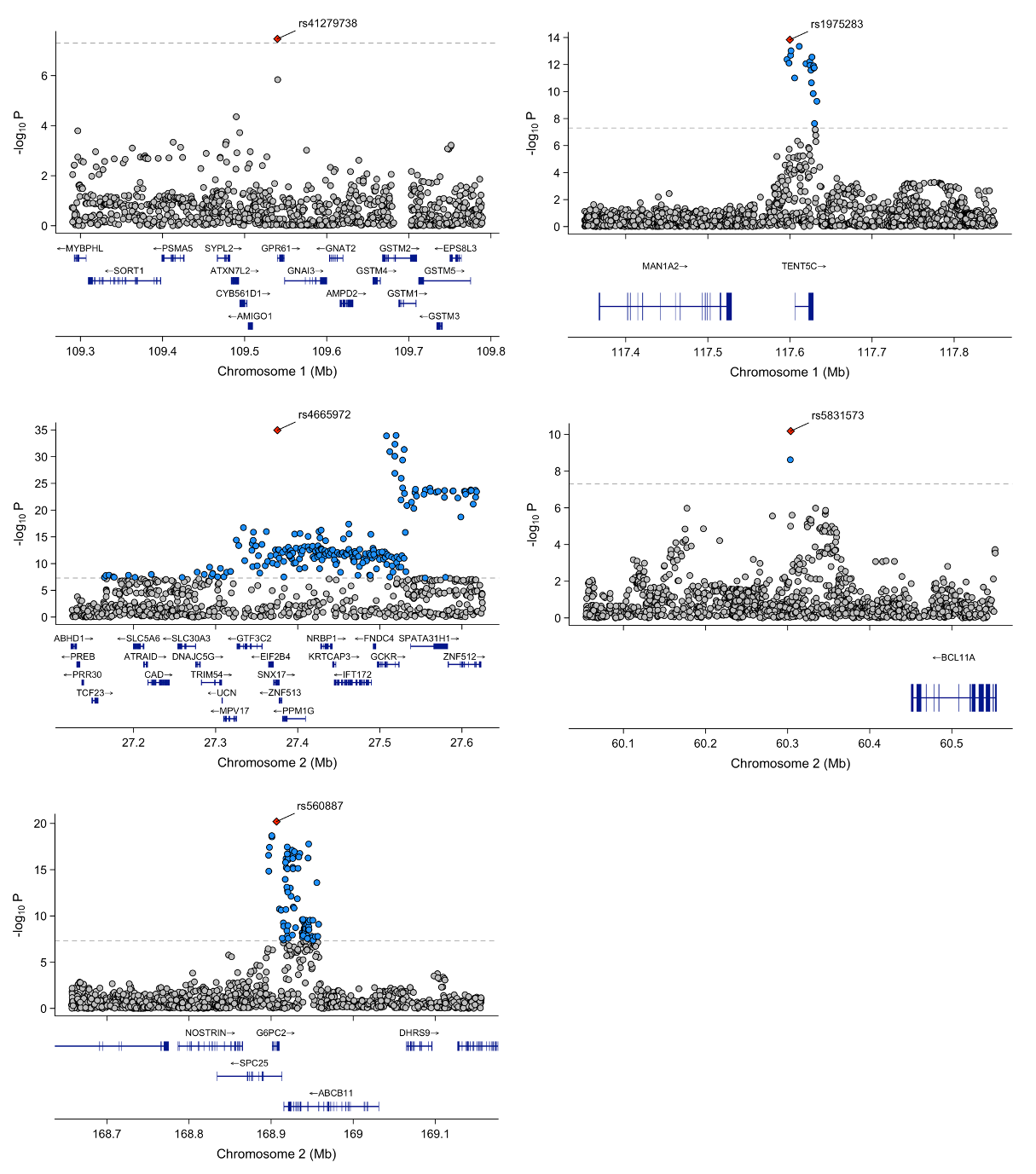

**Supplementary Figure 3.** *Locus zoom plot for the genomic region surrounding the index SNPs located on chromosomes 1 and 2, reaching genome-wide significance in the maternal multi-ancestry GWAS meta-analysis of GDM.* Each point represents a variant from the meta-analysis, with the red diamond indicating the independent lead variant. Blue variants represent nearby variants also reaching genome-wide significance in the meta-analysis. Variants were plotted with their conditional p-value (on a -log10 scale) as a function of genomic position (GRCh38). The dashed line indicates the genome-wide significance threshold. GDM: Gestational Diabetes Mellitus.

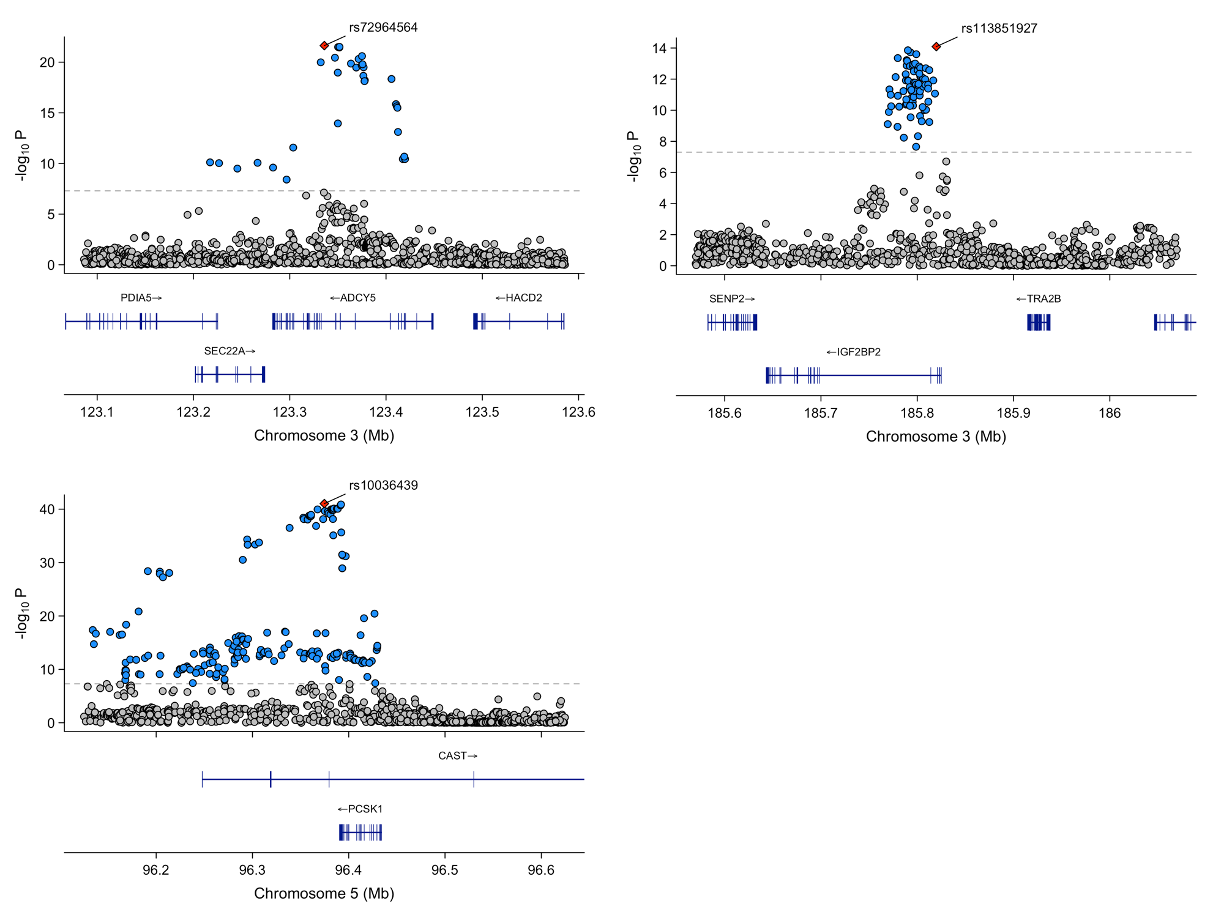

**Supplementary Figure 4.** *Locus zoom plot for the genomic region surrounding the index SNPs located on chromosomes 3 and 5, reaching genome-wide significance in the maternal multi-ancestry GWAS meta-analysis of GDM.* Each point represents a variant from the meta-analysis, with the red diamond indicating the independent lead variant. Blue variants represent nearby variants also reaching genome-wide significance in the meta-analysis. Variants were plotted with their conditional p-value (on a -log10 scale) as a function of genomic position (GRCh38). The dashed line indicates the genome-wide significance threshold. GDM: Gestational Diabetes Mellitus.

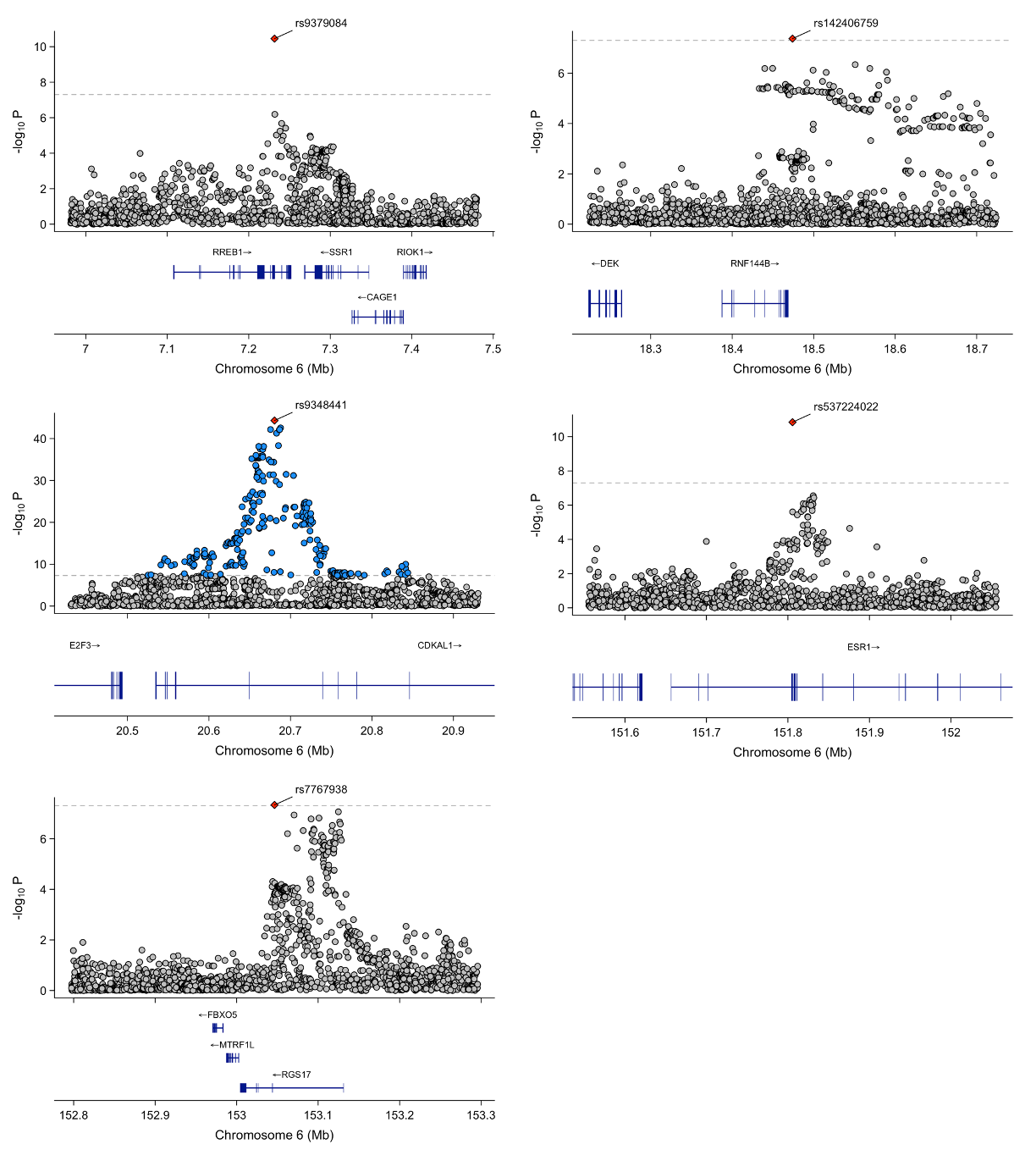

**Supplementary Figure 5.** *Locus zoom plot for the genomic region surrounding the index SNPs located on chromosome 6, reaching genome-wide significance in the maternal multi-ancestry GWAS meta-analysis of GDM.* Each point represents a variant from the meta-analysis, with the red diamond indicating the independent lead variant. Blue variants represent nearby variants also reaching genome-wide significance in the meta-analysis. Variants were plotted with their conditional p-value (on a -log10 scale) as a function of genomic position (GRCh38). The dashed line indicates the genome-wide significance threshold. GDM: Gestational Diabetes Mellitus.

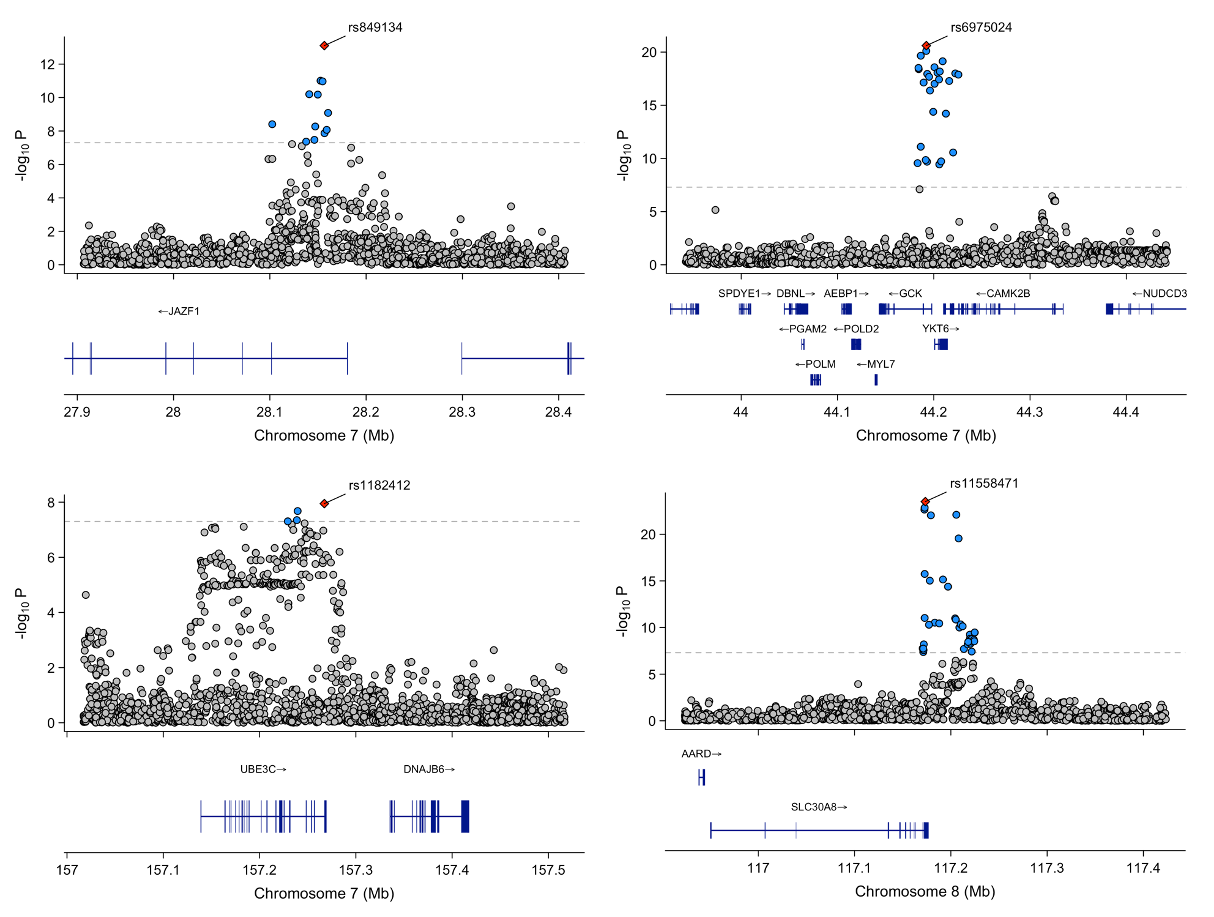

**Supplementary Figure 6.** *Locus zoom plot for the genomic region surrounding the index SNPs located on chromosomes 7 and 8, reaching genome-wide significance in the maternal multi-ancestry GWAS meta-analysis of GDM.* Each point represents a variant from the meta-analysis, with the red diamond indicating the independent lead variant. Blue variants represent nearby variants also reaching genome-wide significance in the meta-analysis. Variants were plotted with their conditional p-value (on a -log10 scale) as a function of genomic position (GRCh38). The dashed line indicates the genome-wide significance threshold. GDM: Gestational Diabetes Mellitus.

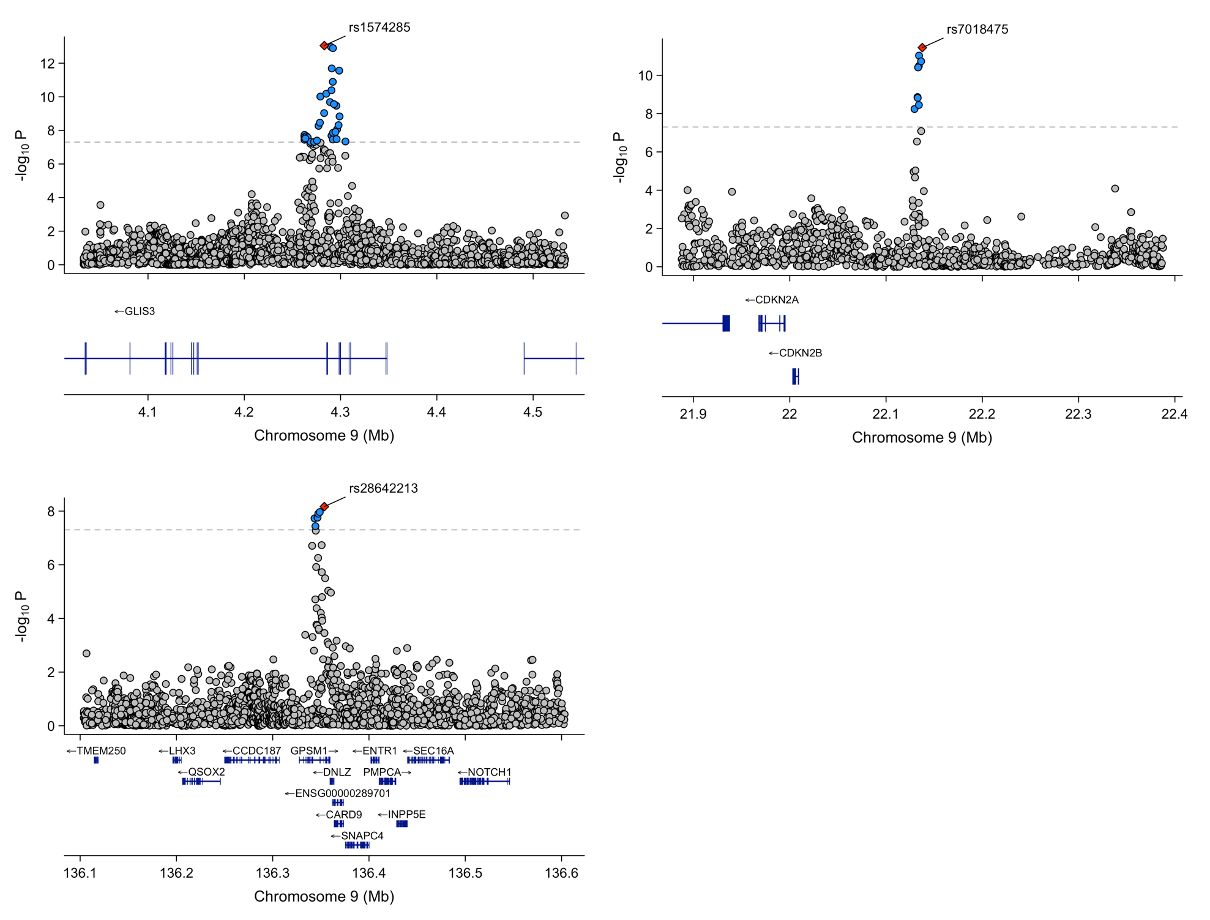

**Supplementary Figure 7.** *Locus zoom plot for the genomic region surrounding the index SNPs located on chromosome 9, reaching genome-wide significance in the maternal multi-ancestry GWAS meta-analysis of GDM.* Each point represents a variant from the meta-analysis, with the red diamond indicating the independent lead variant. Blue variants represent nearby variants also reaching genome-wide significance in the meta-analysis. Variants were plotted with their conditional p-value (on a -log10 scale) as a function of genomic position (GRCh38). The dashed line indicates the genome-wide significance threshold. GDM: Gestational Diabetes Mellitus.

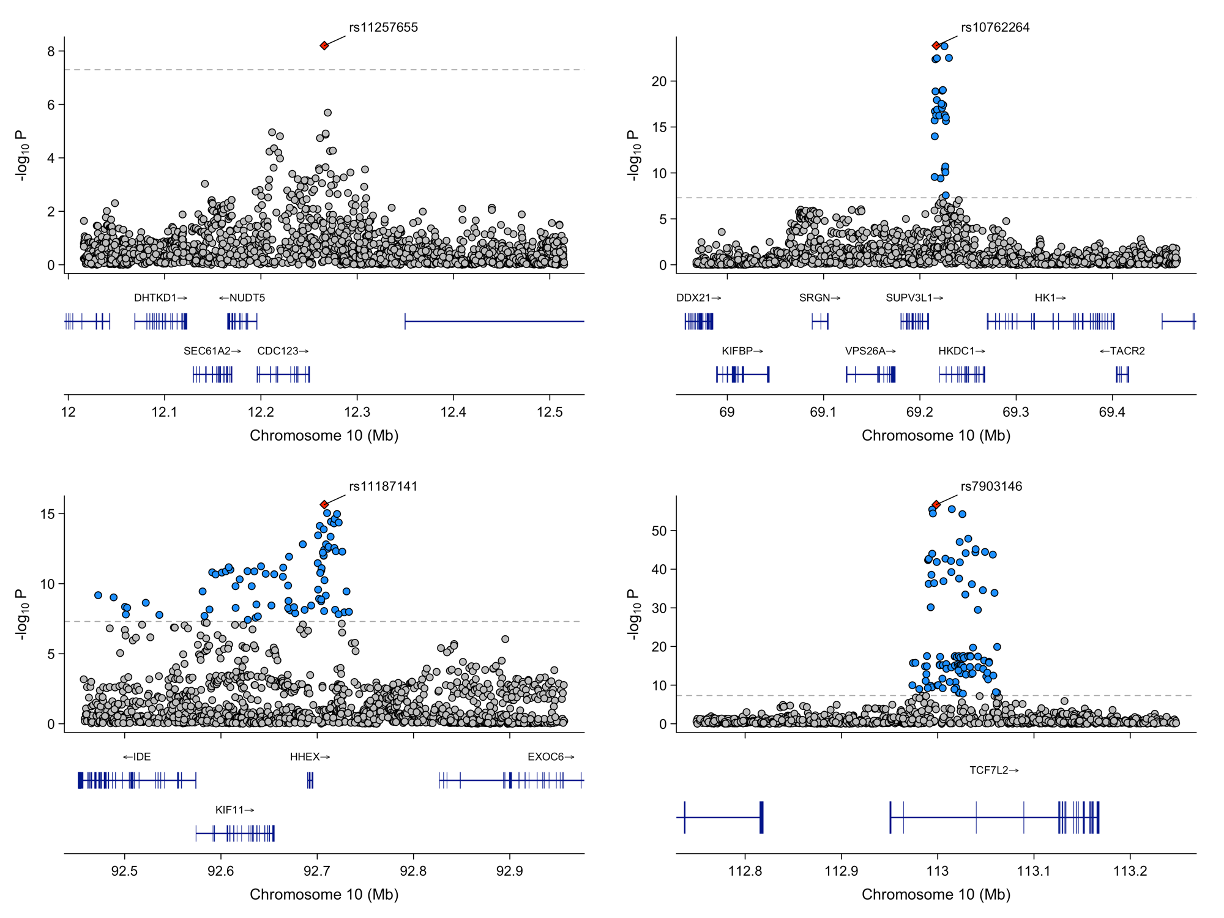

**Supplementary Figure 8.** *Locus zoom plot for the genomic region surrounding the index SNPs located on chromosome 10, reaching genome-wide significance in the maternal multi-ancestry GWAS meta-analysis of GDM.* Each point represents a variant from the meta-analysis, with the red diamond indicating the independent lead variant. Blue variants represent nearby variants also reaching genome-wide significance in the meta-analysis. Variants were plotted with their conditional p-value (on a -log10 scale) as a function of genomic position (GRCh38). The dashed line indicates the genome-wide significance threshold. GDM: Gestational Diabetes Mellitus.

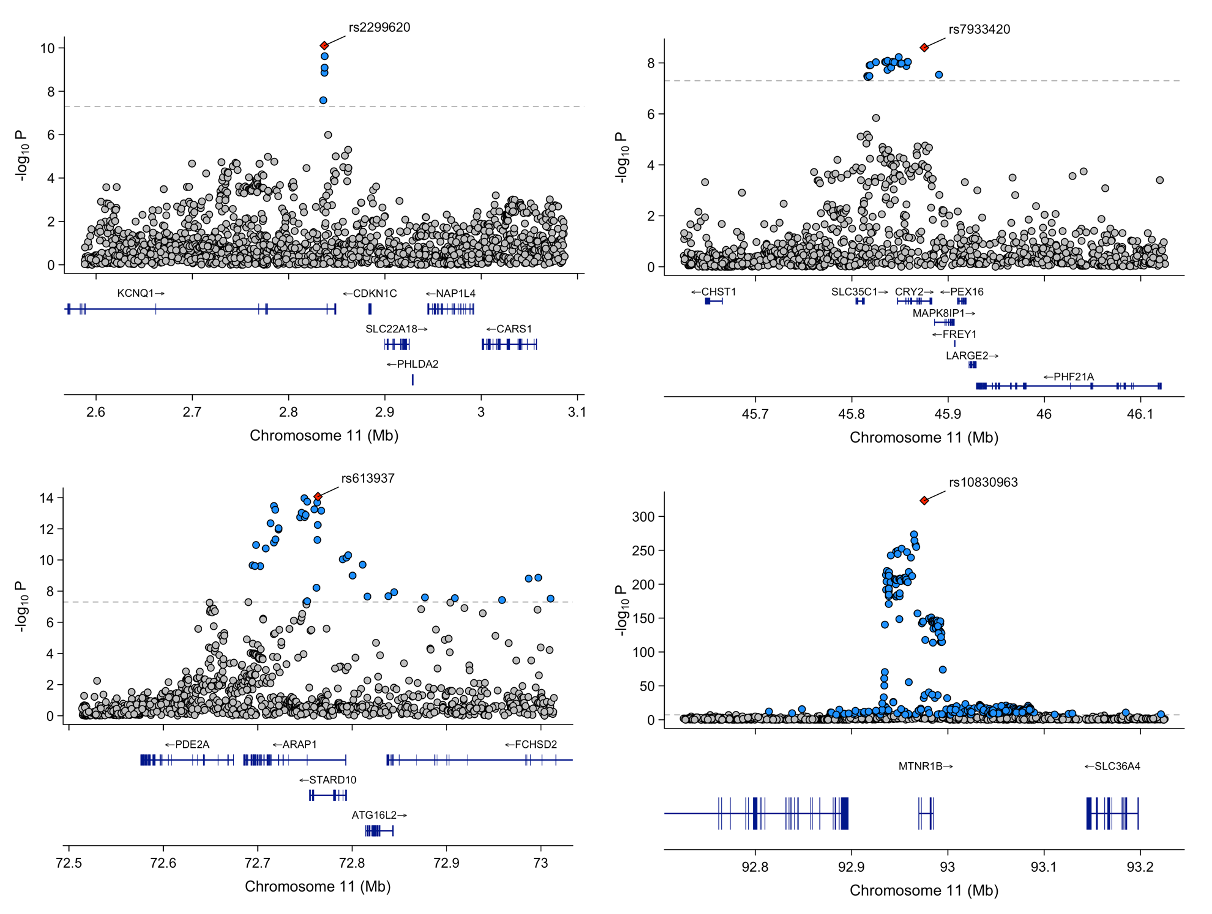

**Supplementary Figure 9.** *Locus zoom plot for the genomic region surrounding the index SNPs located on chromosome 11, reaching genome-wide significance in the maternal multi-ancestry GWAS meta-analysis of GDM.* Each point represents a variant from the meta-analysis, with the red diamond indicating the independent lead variant. Blue variants represent nearby variants also reaching genome-wide significance in the meta-analysis. Variants were plotted with their conditional p-value (on a -log10 scale) as a function of genomic position (GRCh38). The dashed line indicates the genome-wide significance threshold. GDM: Gestational Diabetes Mellitus.

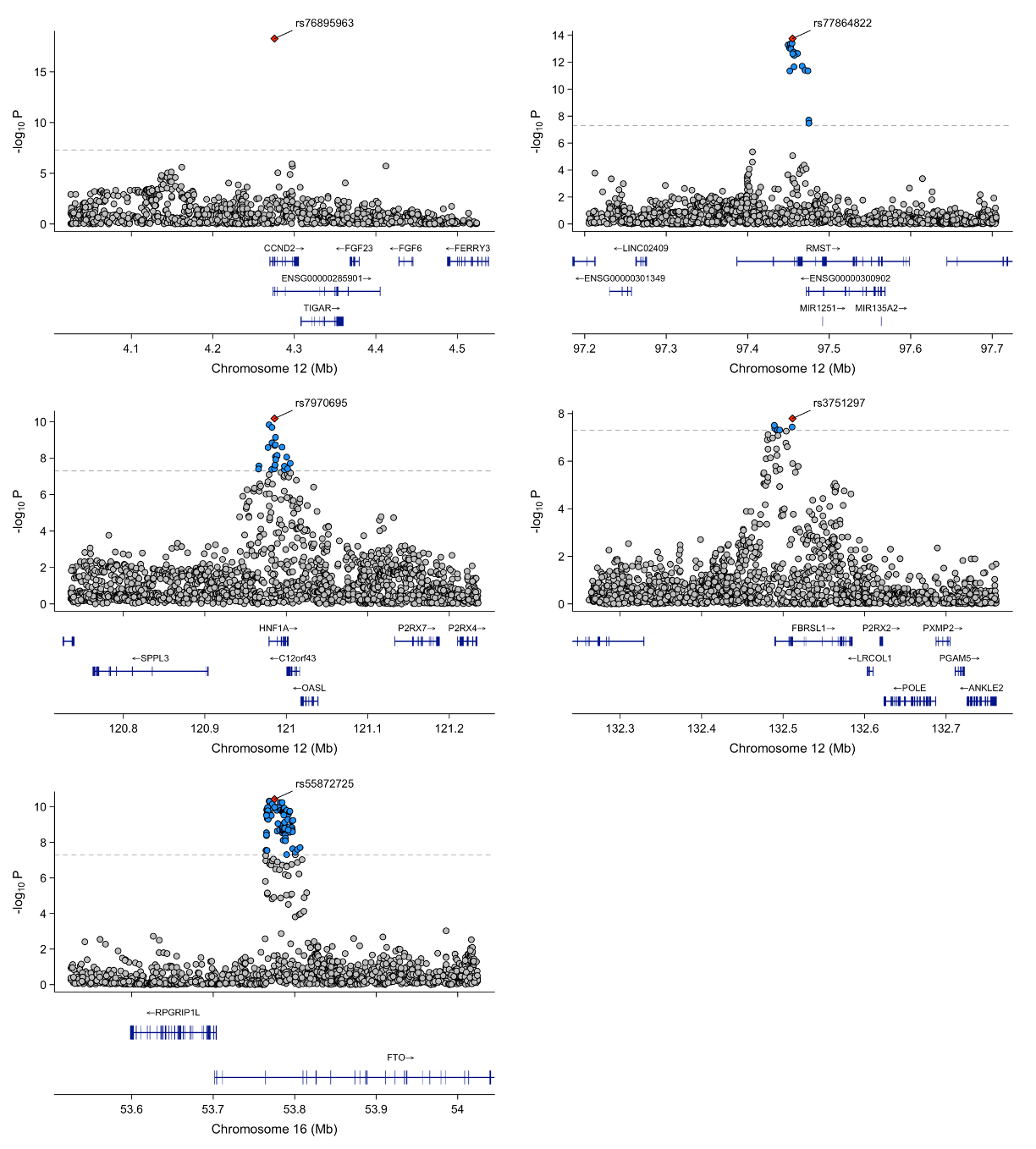

**Supplementary Figure 10.** *Locus zoom plot for the genomic region surrounding the index SNPs located on chromosomes 12 and 16, reaching genome-wide significance in the maternal multi-ancestry GWAS meta-analysis of GDM.* Each point represents a variant from the meta-analysis, with the red diamond indicating the independent lead variant. Blue variants represent nearby variants also reaching genome-wide significance in the meta-analysis. Variants were plotted with their conditional p-value (on a -log10 scale) as a function of genomic position (GRCh38). The dashed line indicates the genome-wide significance threshold. GDM: Gestational Diabetes Mellitus.

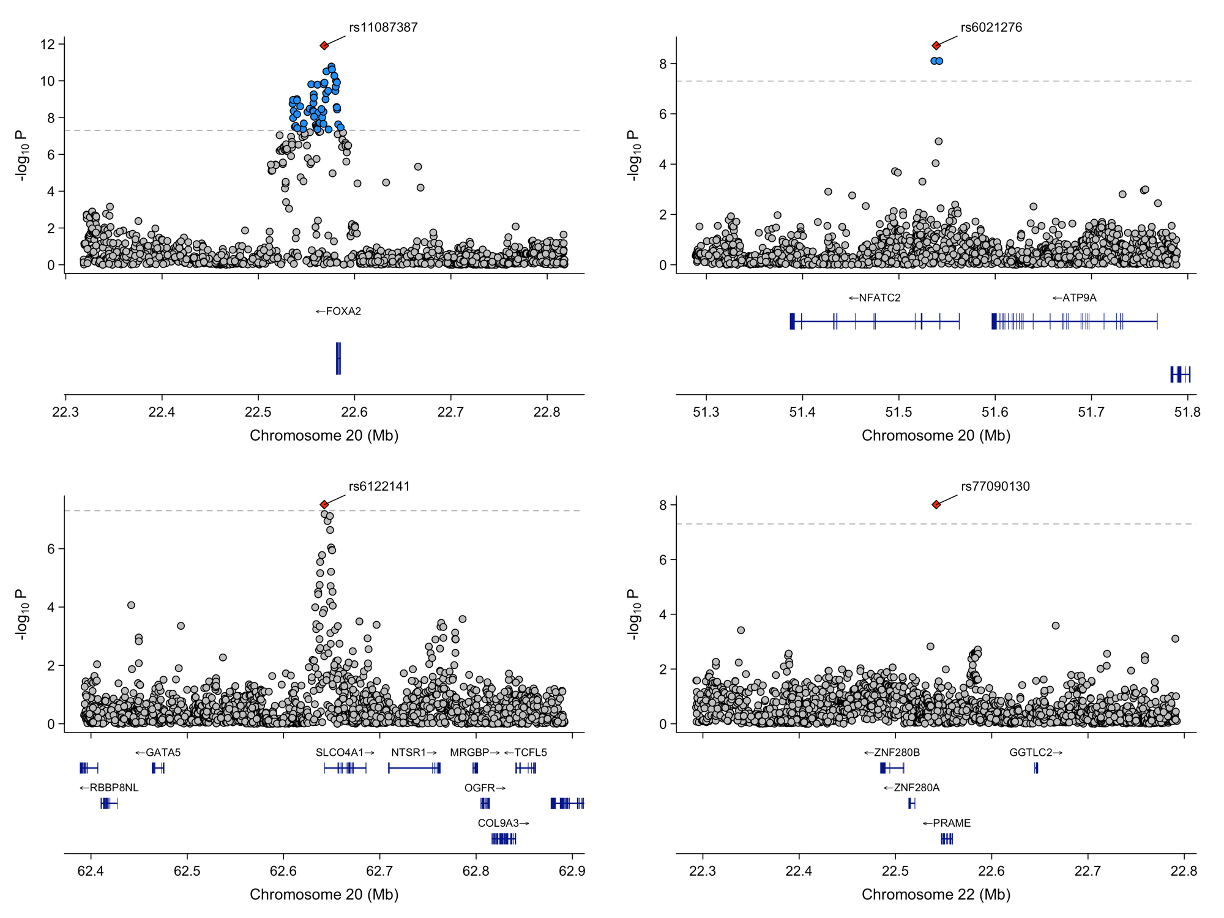

**Supplementary Figure 11.** *Locus zoom plot for the genomic region surrounding the index SNPs located on chromosomes 20 and 22, reaching genome-wide significance in the maternal multi-ancestry GWAS meta-analysis of GDM.* Each point represents a variant from the meta-analysis, with the red diamond indicating the independent lead variant. Blue variants represent nearby variants also reaching genome-wide significance in the meta-analysis. Variants were plotted with their conditional p-value (on a -log10 scale) as a function of genomic position (GRCh38). The dashed line indicates the genome-wide significance threshold. GDM: Gestational Diabetes Mellitus.

#### 1.4 Multi-ancestry GWAS meta-analyses of the glycemic traits

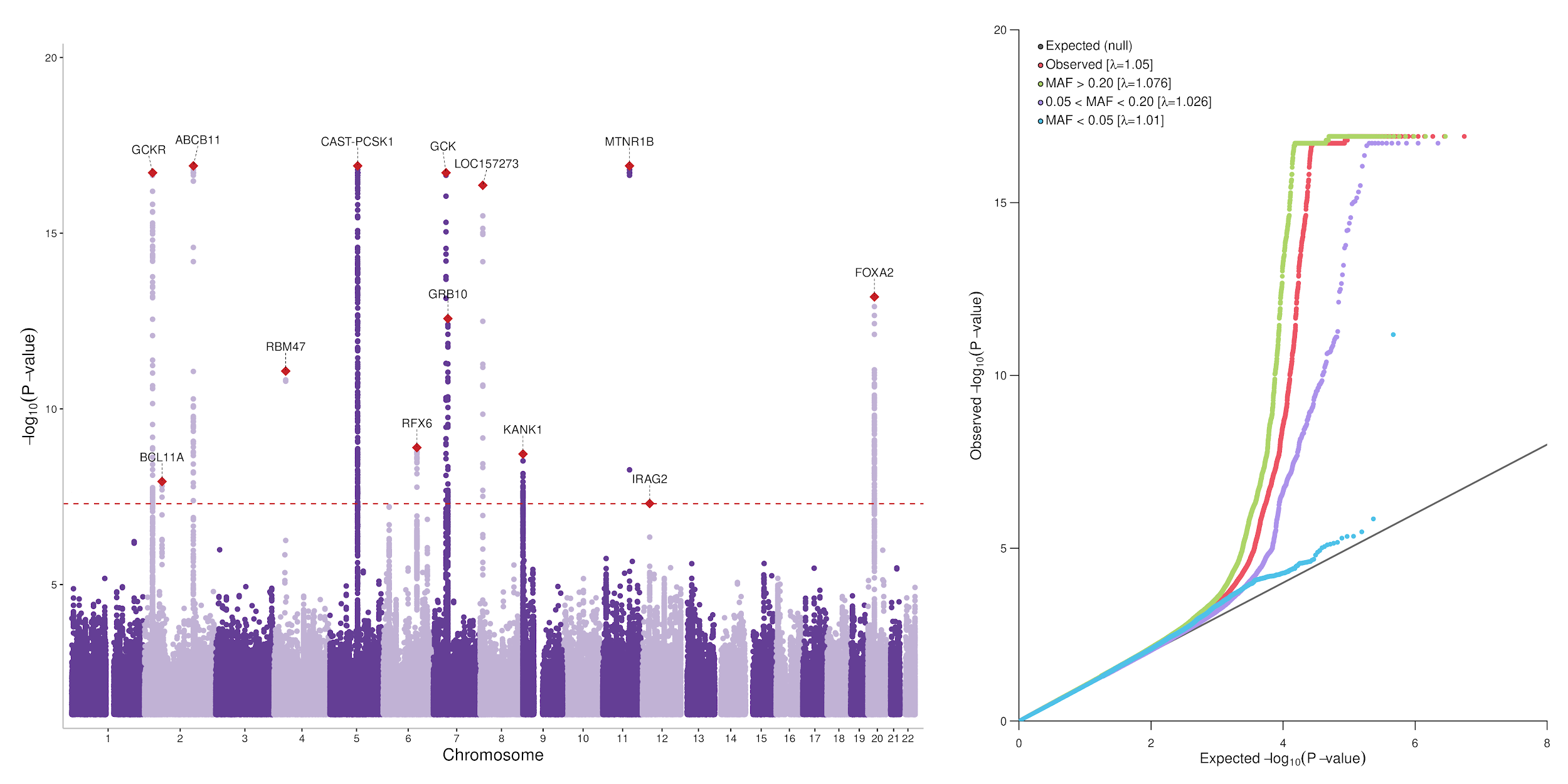

**Supplementary Figure 12.** *Manhattan plot and QQ plot of the results for the GWAS meta-analysis of maternal FG, including 55,371 pregnant women.* Only SNPs with P-value < 0.05 are plotted on the Manhattan plot. The horizontal red dashed line represents genome-wide significance (P=5$\times$10^-8^). FG; Fasting Glucose. MAF; minor allele frequency.

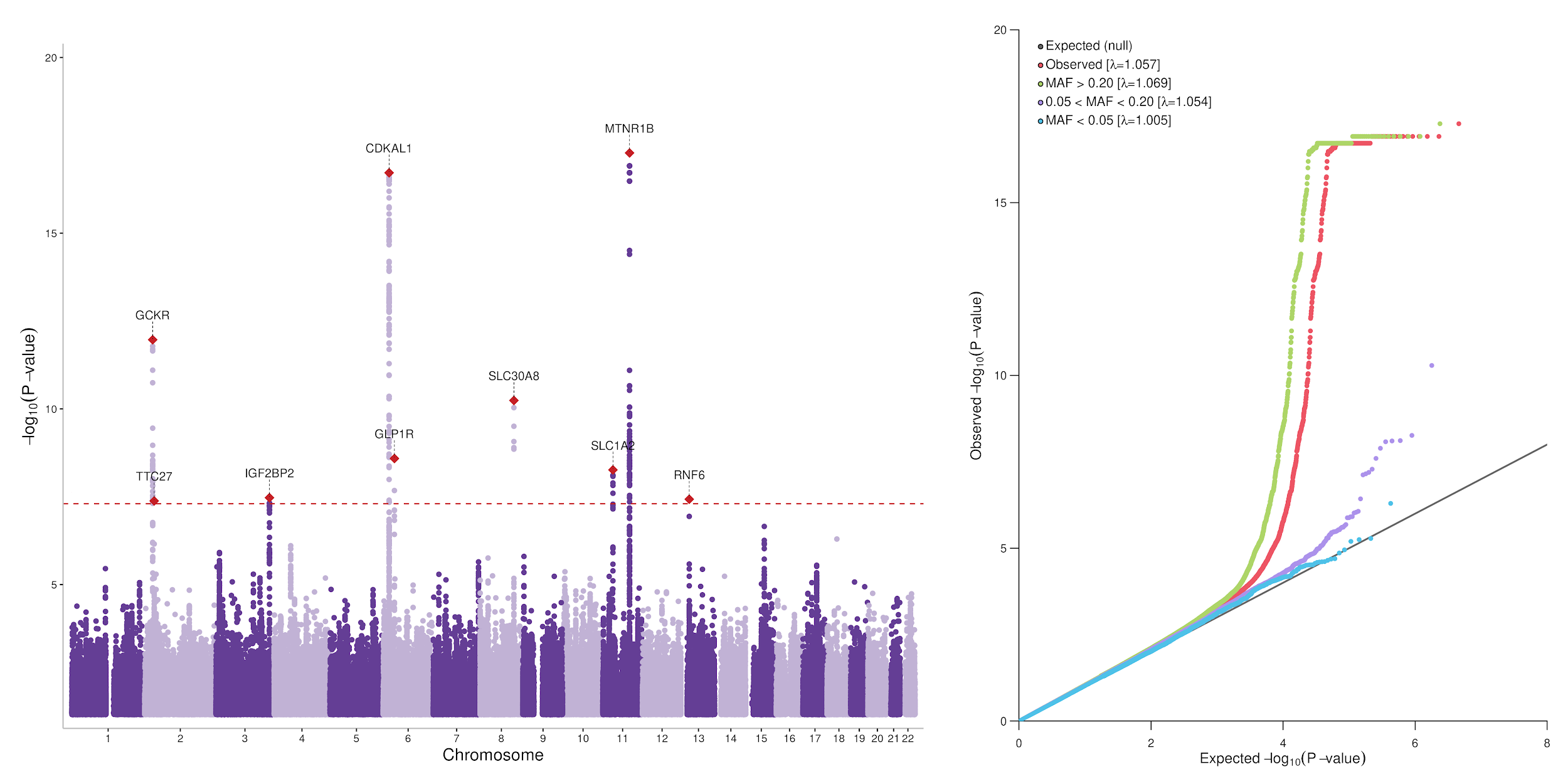

**Supplementary Figure 13.** *Manhattan plot and QQ plot of the results for the GWAS meta-analysis of maternal 1hG, including 38,439 pregnant women.* Only SNPs with P-value < 0.05 are plotted on the Manhattan plot. The horizontal red dashed line represents genome-wide significance (P=5$\times$10^-8^). 1hG; 1-hour post-oral glucose tolerance test. MAF; minor allele frequency.

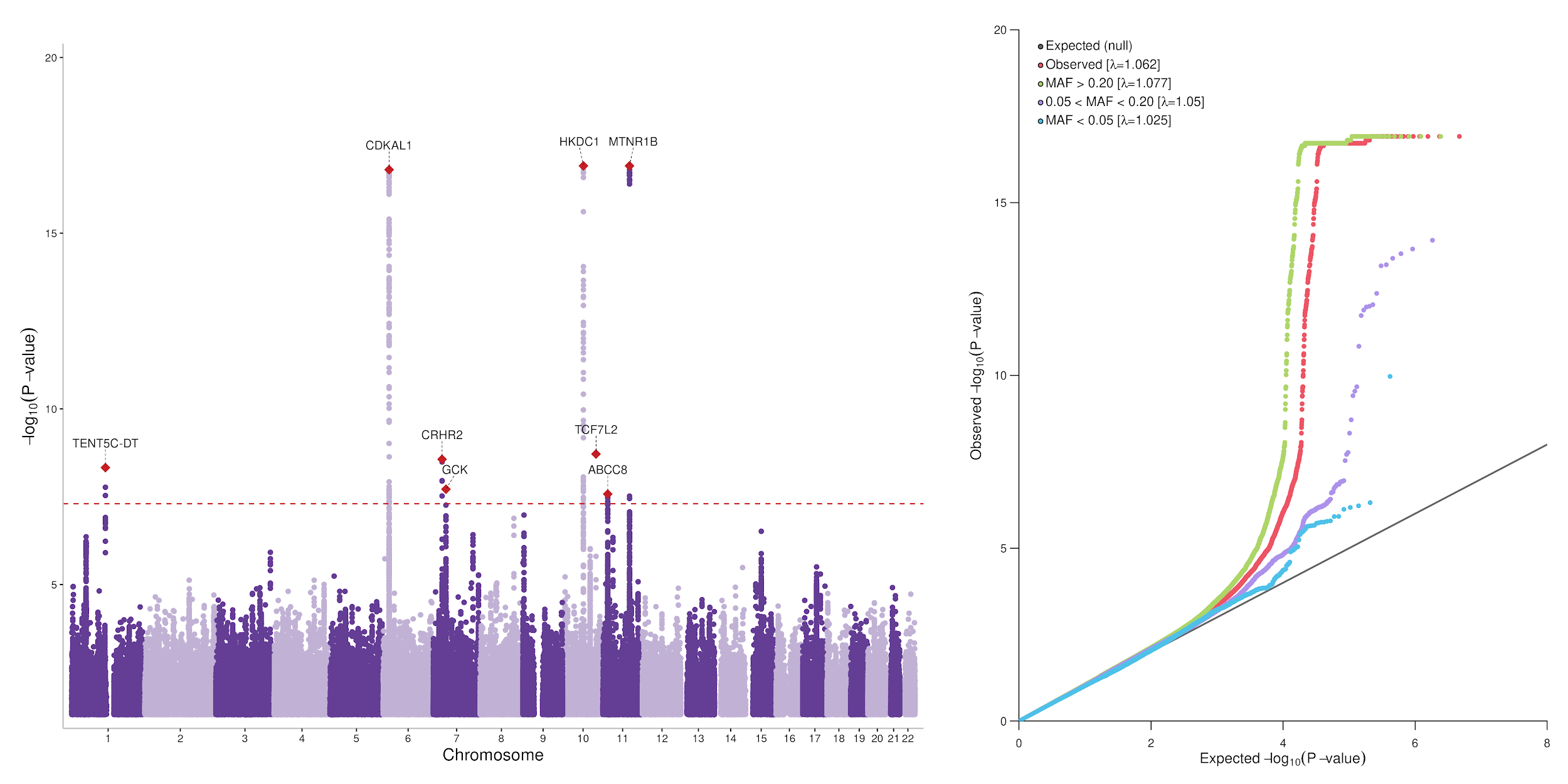

**Supplementary Figure 14.** *Manhattan plot and QQ plot of the results for the GWAS meta-analysis of maternal 2hG, including 46,401 pregnant women.* Only SNPs with P-value < 0.05 are plotted on the Manhattan plot. The horizontal red dashed line represents genome-wide significance (P=5$\times$10^-8^). 2hG; 2-hour post-oral glucose tolerance test. MAF; minor allele frequency.

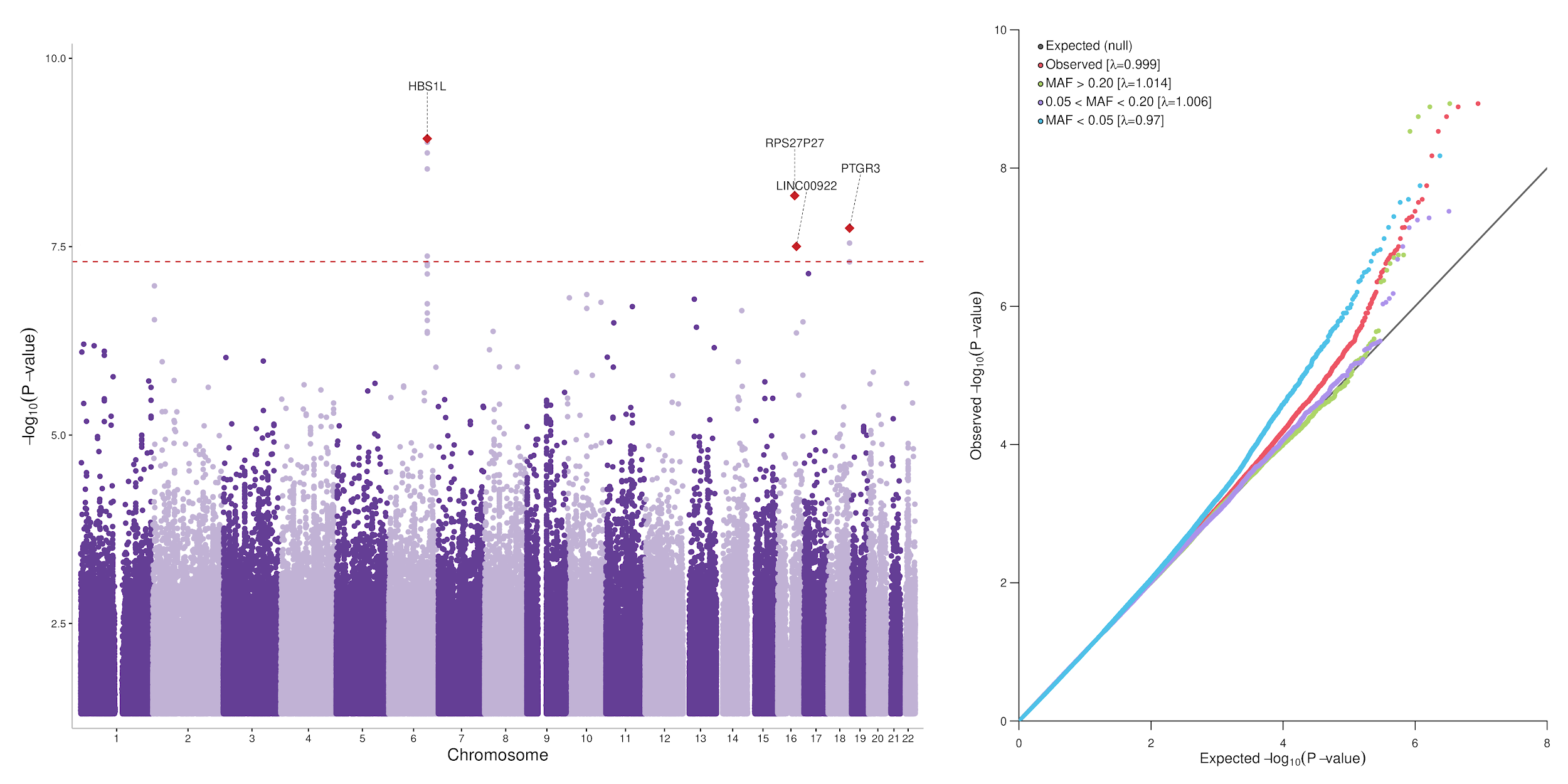

**Supplementary Figure 15.** *Manhattan plot and QQ plot of the results for the GWAS meta-analysis of maternal HbA1c, including 9,724 pregnant women.* Only SNPs with P-value < 0.05 are plotted on the Manhattan plot. The horizontal red dashed line represents genome-wide significance (P=5$\times$10^-8^). MAF; minor allele frequency.

#### 1.5 Axes of genetic variation separating GWAS of glycemic traits in MR-MEGA

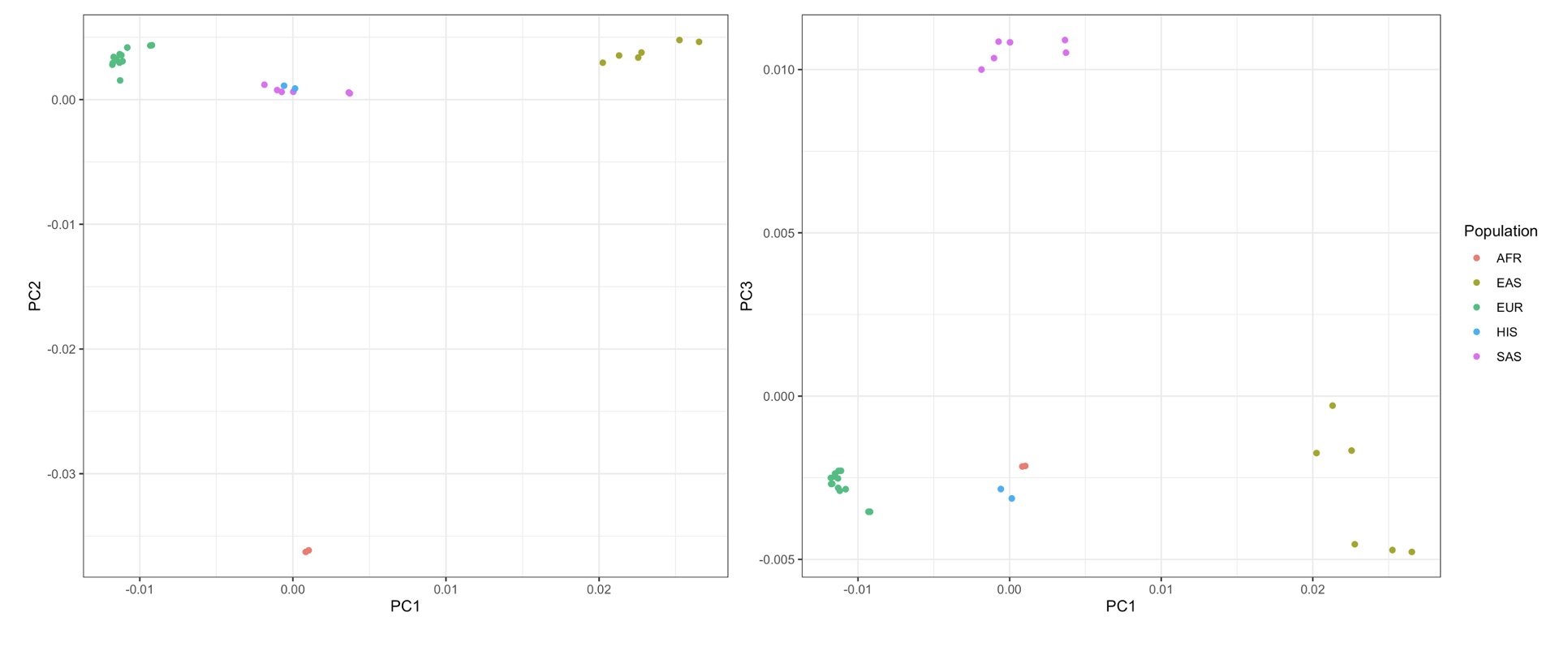
**Supplementary Figure 16.** *Axes of genetic variation separating maternal GWAS of FG.* The first three axes of genetic variation from multi-dimensional scaling of the Euclidean distance matrix between GWAS are sufficient to separate five ancestry groups: South Asians (SAS), African American (AFR), Hispanic/Latino (HIS), East Asian (EAS) and European (EUR). FG; Fasting Glucose.

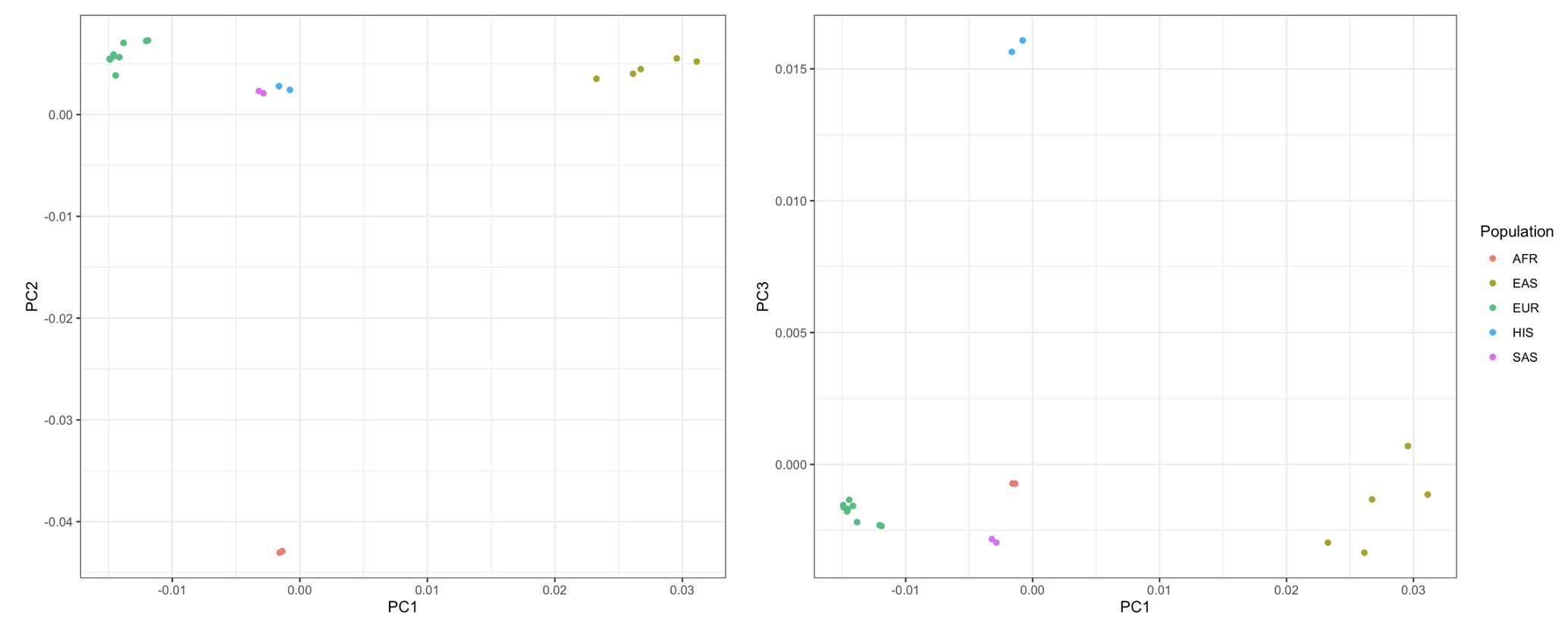

**Supplementary Figure 17.** *Axes of genetic variation separating maternal GWAS of 1hG.* The first three axes of genetic variation from multi-dimensional scaling of the Euclidean distance matrix between GWAS are sufficient to separate five ancestry groups: South Asians (SAS), African American (AFR), Hispanic/Latino (HIS), East Asian (EAS) and European (EUR). 1hG; 1-hour glucose post-oral glucose tolerance test.

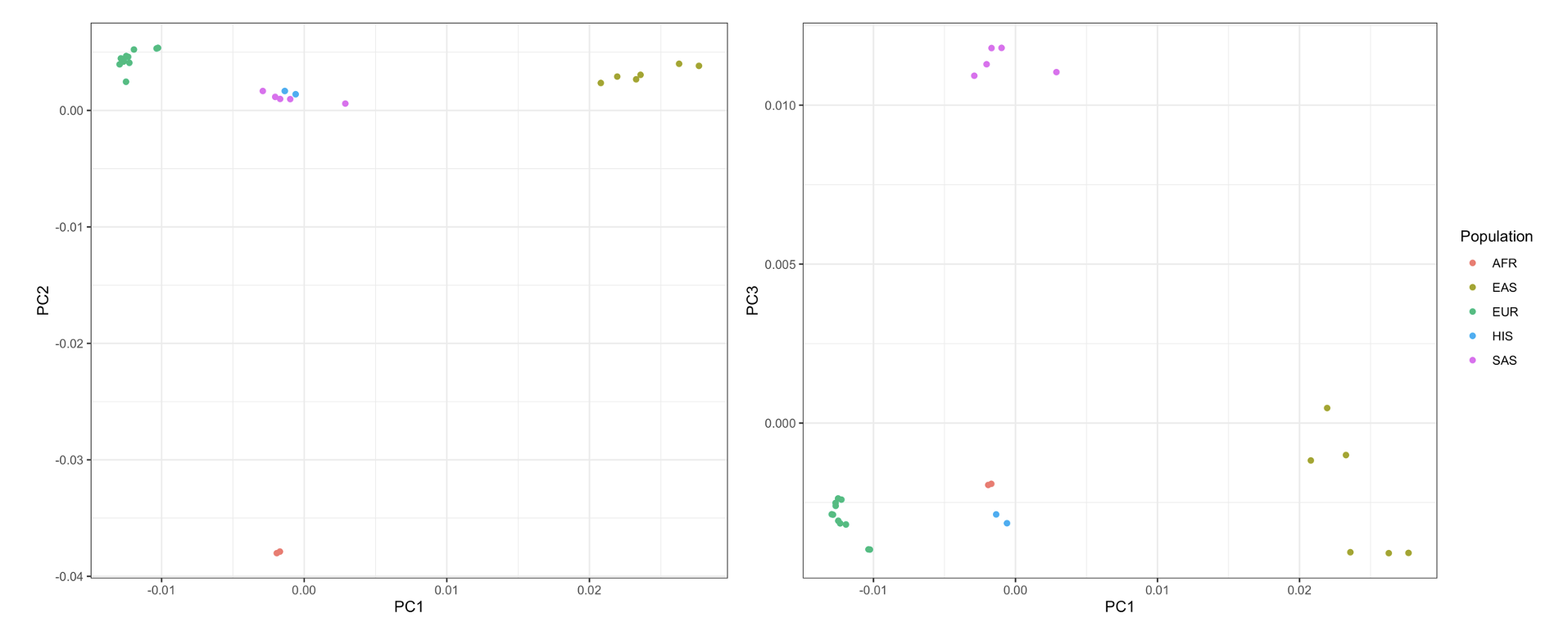

**Supplementary Figure 18.** *Axes of genetic variation separating maternal GWAS of 2hG.* The first three axes of genetic variation from multi-dimensional scaling of the Euclidean distance matrix between GWAS are sufficient to separate five ancestry groups: South Asians (SAS), African American (AFR), Hispanic/Latino (HIS), East Asian (EAS) and European (EUR). 2hG; 2-hour glucose post-oral glucose tolerance test.

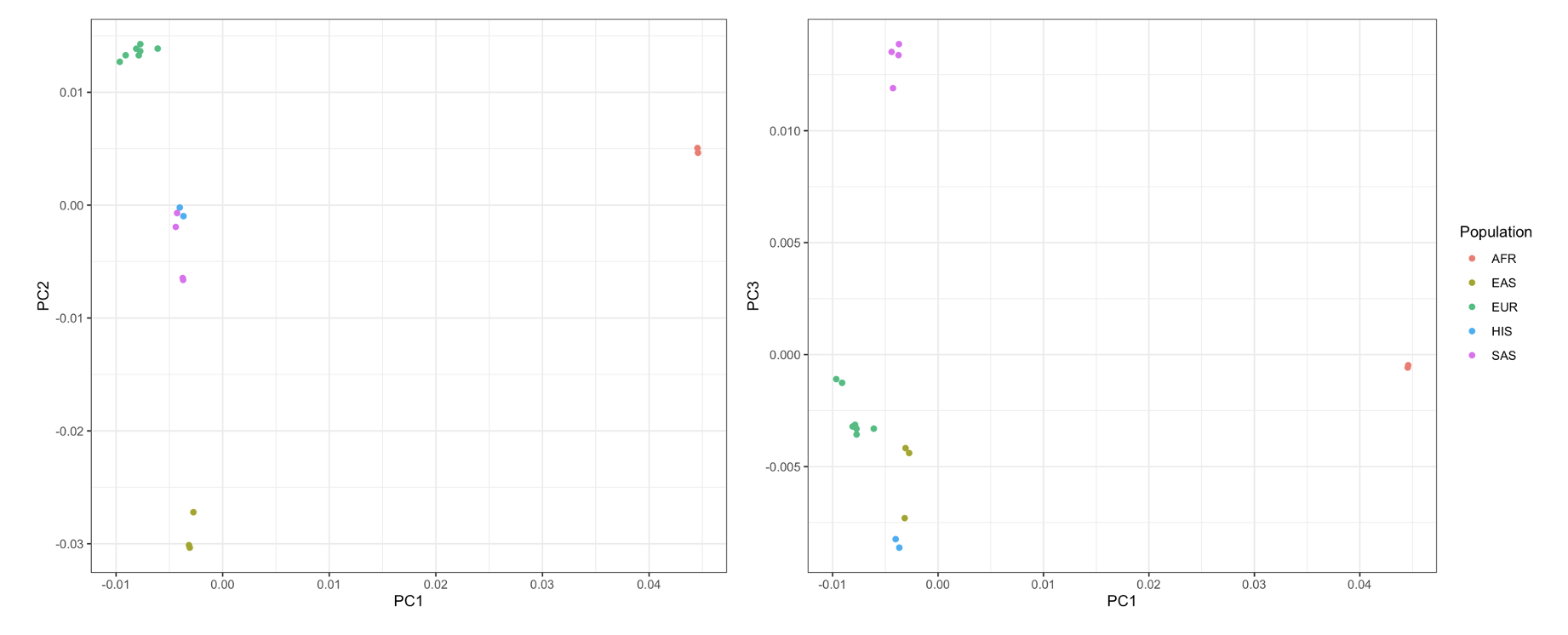
**Supplementary Figure 19.** *Axes of genetic variation separating maternal GWAS of HbA1c.* The first three axes of genetic variation from multi-dimensional scaling of the Euclidean distance matrix between GWAS are sufficient to separate five ancestry groups: South Asians (SAS), African American (AFR), Hispanic/Latino (HIS), East Asian (EAS) and European (EUR).

#### 1.6 Locus ZoomPlots for the meta-analyses of glycemic traits

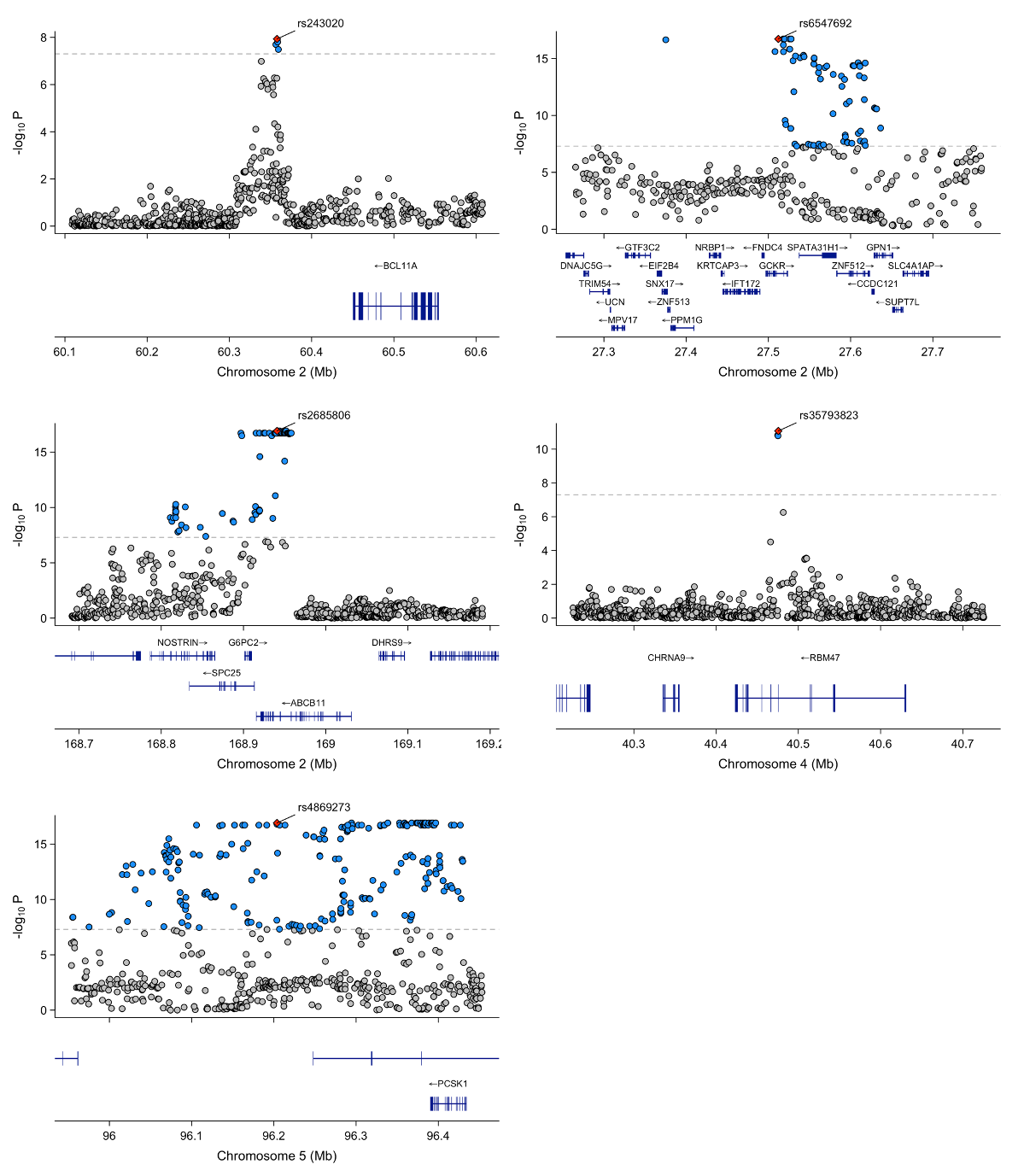

**Supplementary Figure 20.** *Locus zoom plot for the genomic region surrounding the index SNPs located on chromosomes 2, 4 and 5, reaching genome-wide significance in the maternal multi-ancestry GWAS meta-analysis of FG.* Each point represents a variant from the meta-analysis, with the red diamond indicating the independent lead variant. Blue variants represent nearby variants also reaching genome-wide significance in the meta-analysis. Variants were plotted with their conditional p-value (on a -log10 scale) as a function of genomic position (GRCh38). The dashed line indicates the genome-wide significance threshold. FG; Fasting Glucose.

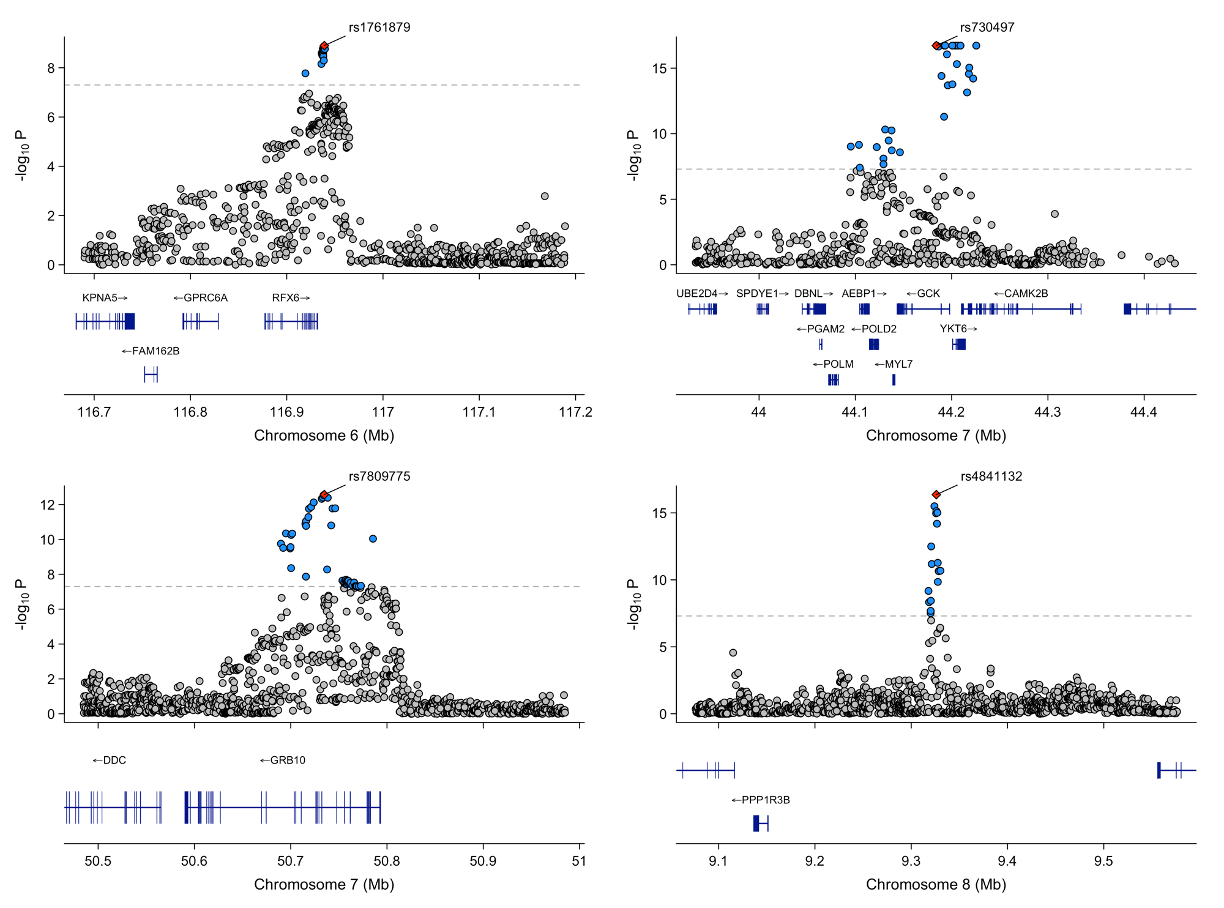

**Supplementary Figure 21.** *Locus zoom plot for the genomic region surrounding the index SNPs located on chromosomes 6, 7 and 8, reaching genome-wide significance in the maternal multi-ancestry GWAS meta-analysis of FG.* Each point represents a variant from the meta-analysis, with the red diamond indicating the independent lead variant. Blue variants represent nearby variants also reaching genome-wide significance in the meta-analysis. Variants were plotted with their conditional p-value (on a -log10 scale) as a function of genomic position (GRCh38). The dashed line indicates the genome-wide significance threshold. FG; Fasting Glucose.

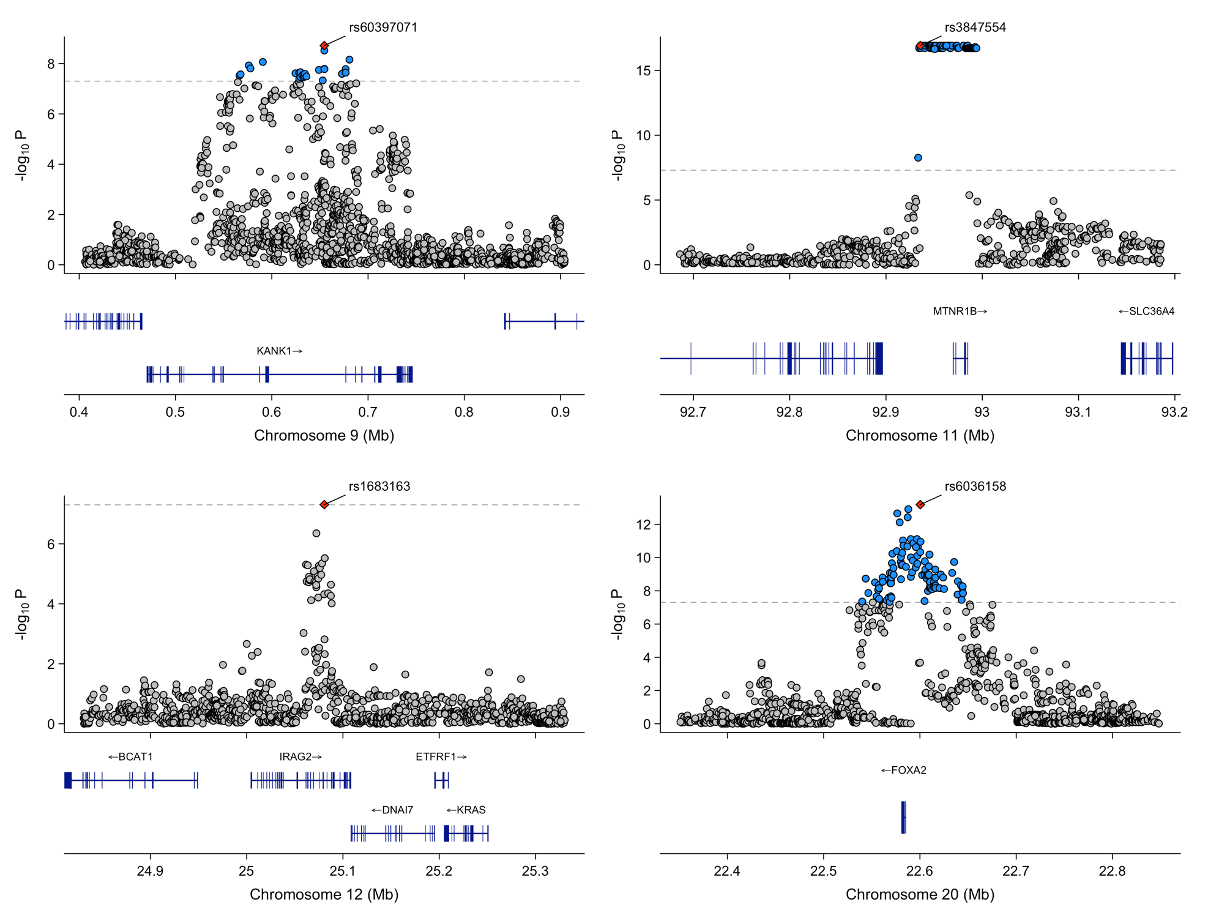

**Supplementary Figure 22.** *Locus zoom plot for the genomic region surrounding the index SNPs located on chromosomes 9, 11, 12 and 20, reaching genome-wide significance in the maternal multi-ancestry GWAS meta-analysis of FG.* Each point represents a variant from the meta-analysis, with the red diamond indicating the independent lead variant. Blue variants represent nearby variants also reaching genome-wide significance in the meta-analysis. Variants were plotted with their conditional p-value (on a -log10 scale) as a function of genomic position (GRCh38). The dashed line indicates the genome-wide significance threshold. FG; Fasting Glucose.

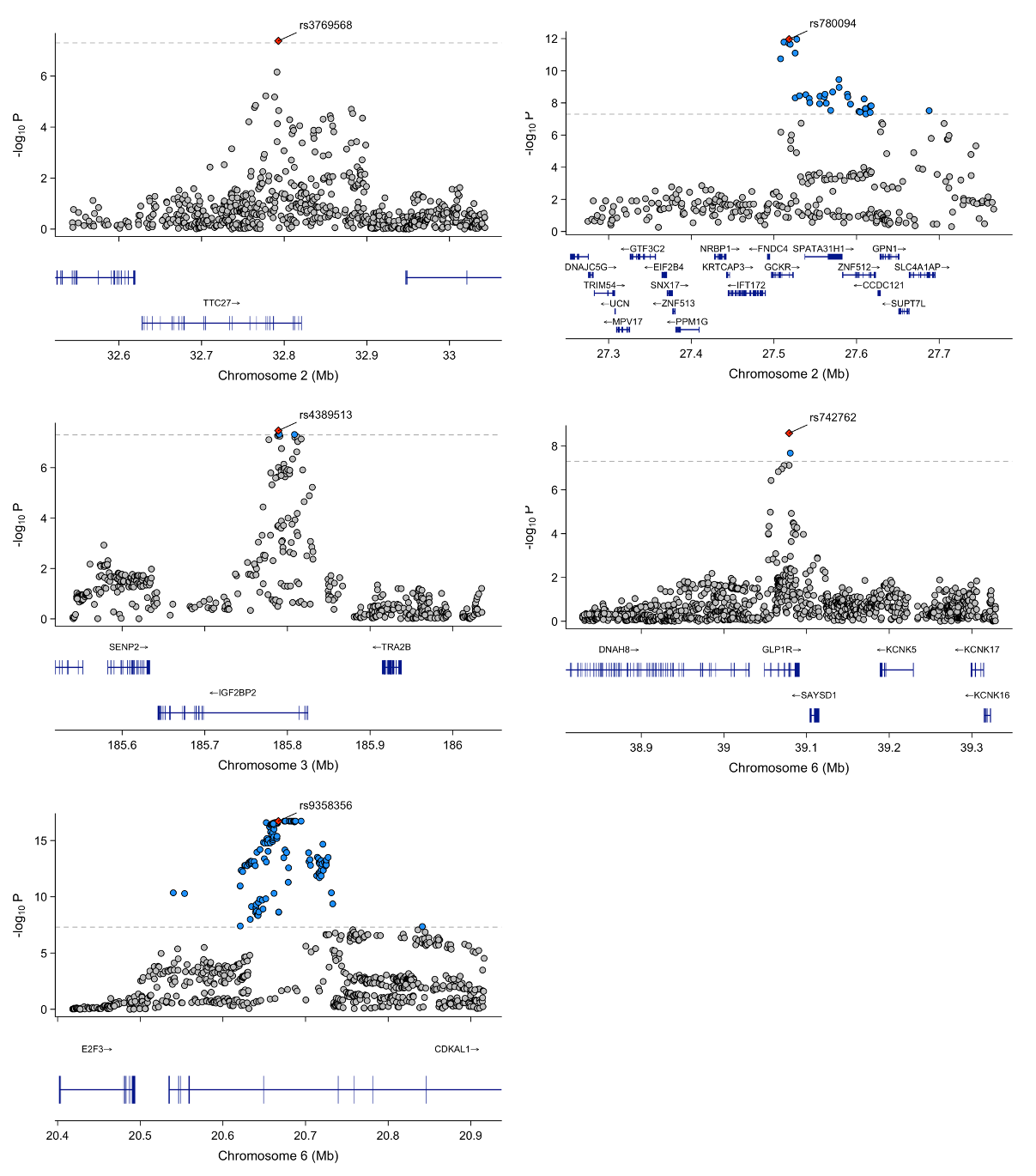

**Supplementary Figure 23.** *Locus zoom plot for the genomic region surrounding the index SNPs located on chromosomes 2, 3, and 6, reaching genome-wide significance in the maternal multi-ancestry GWAS meta-analysis of 1hG.* Each point represents a variant from the meta-analysis, with the red diamond indicating the independent lead variant. Blue variants represent nearby variants also reaching genome-wide significance in the meta-analysis. Variants were plotted with their conditional p-value (on a -log10 scale) as a function of genomic position (GRCh38). 1hG; 1-hour glucose post-oral glucose tolerance test.

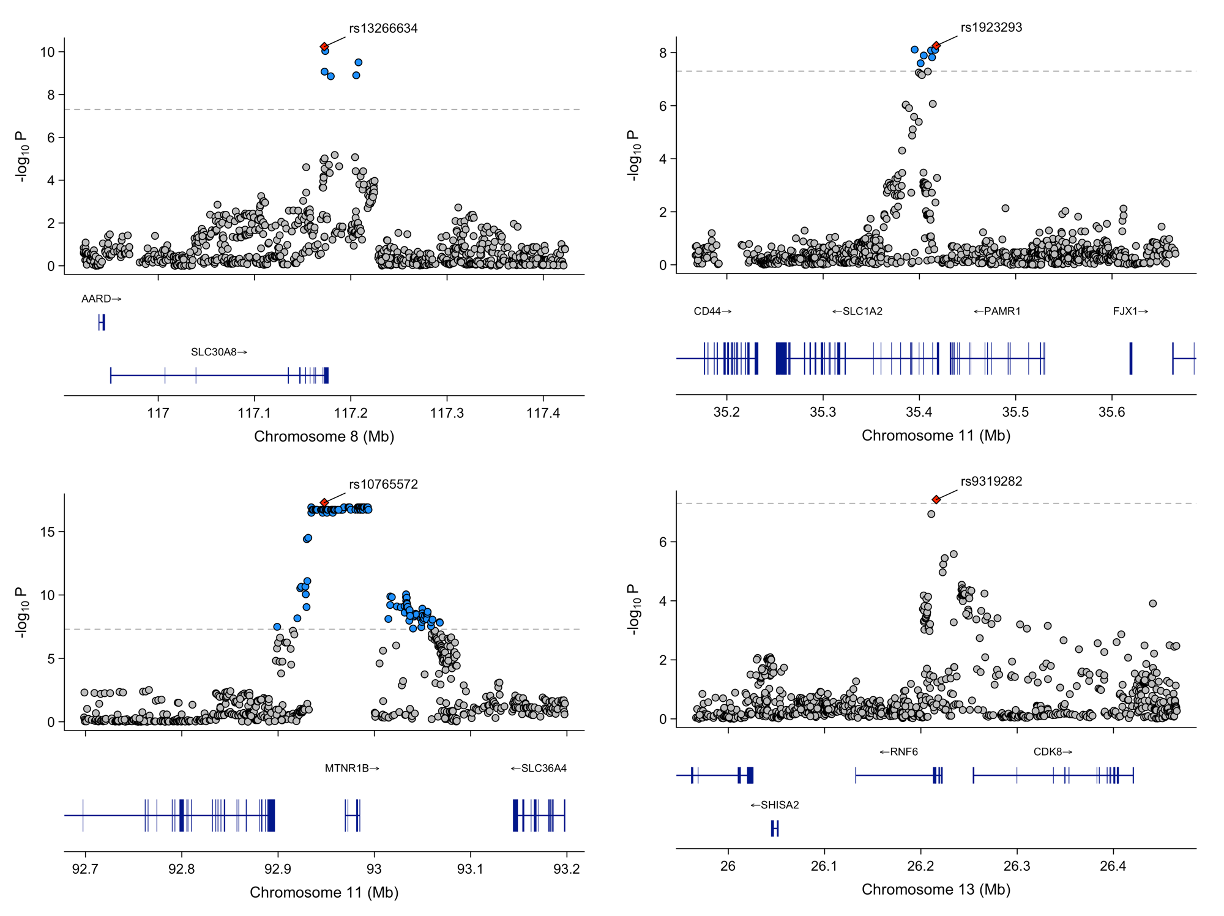

**Supplementary Figure 24.** *Locus zoom plot for the genomic region surrounding the index SNPs located on chromosomes 8, 11, and 13, reaching genome-wide significance in the maternal multi-ancestry GWAS meta-analysis of 1hG.* Each point represents a variant from the meta-analysis, with the red diamond indicating the independent lead variant. Blue variants represent nearby variants also reaching genome-wide significance in the meta-analysis. Variants were plotted with their conditional p-value (on a -log10 scale) as a function of genomic position (GRCh38). 1hG; 1-hour glucose post-oral glucose tolerance test.

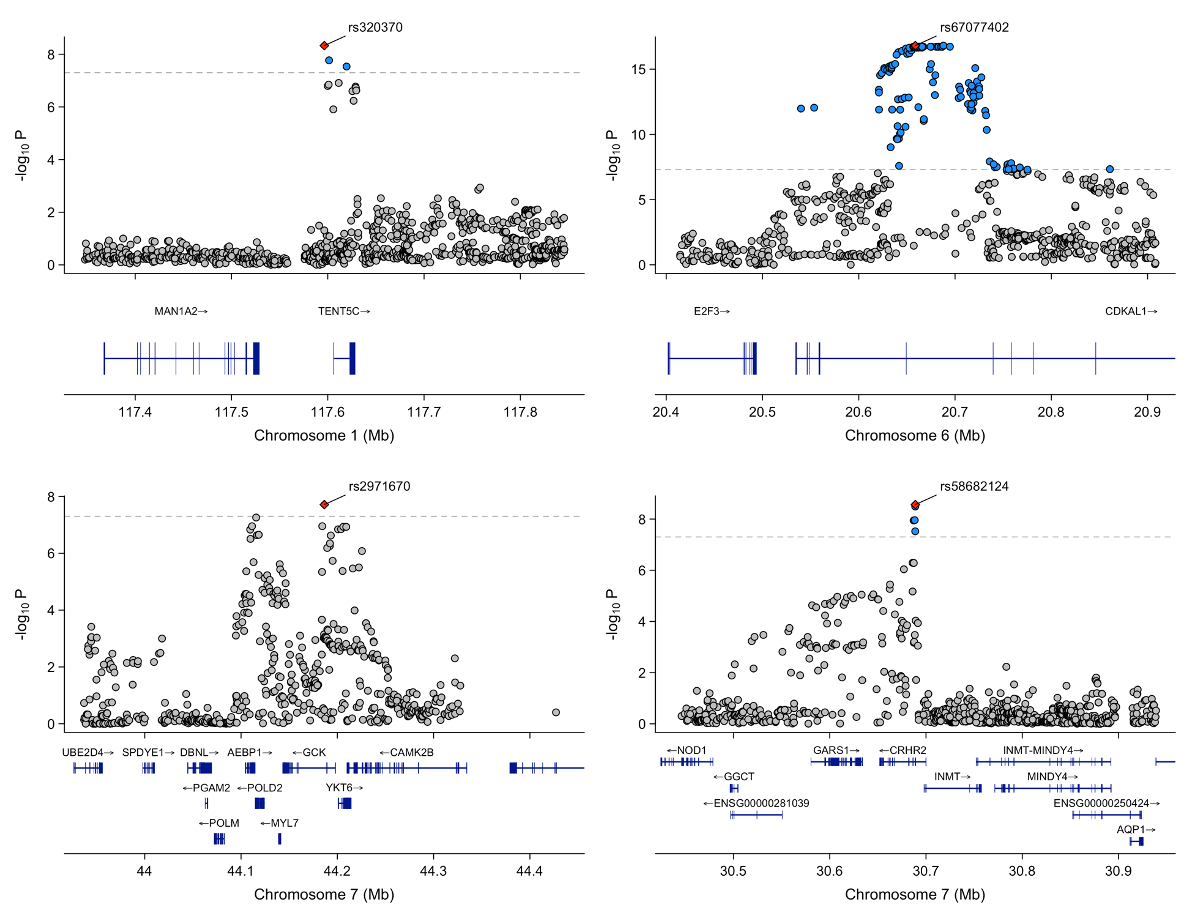

**Supplementary Figure 25.** *Locus zoom plot for the genomic region surrounding the index SNPs located on chromosomes 1, 6, and 7, reaching genome-wide significance in the maternal multi-ancestry GWAS meta-analysis of 2hG.* Each point represents a variant from the meta-analysis, with the red diamond indicating the independent lead variant. Blue variants represent nearby variants also reaching genome-wide significance in the meta-analysis. Variants were plotted with their conditional p-value (on a -log10 scale) as a function of genomic position (GRCh38). 2hG; 2-hour glucose post-oral glucose tolerance test.

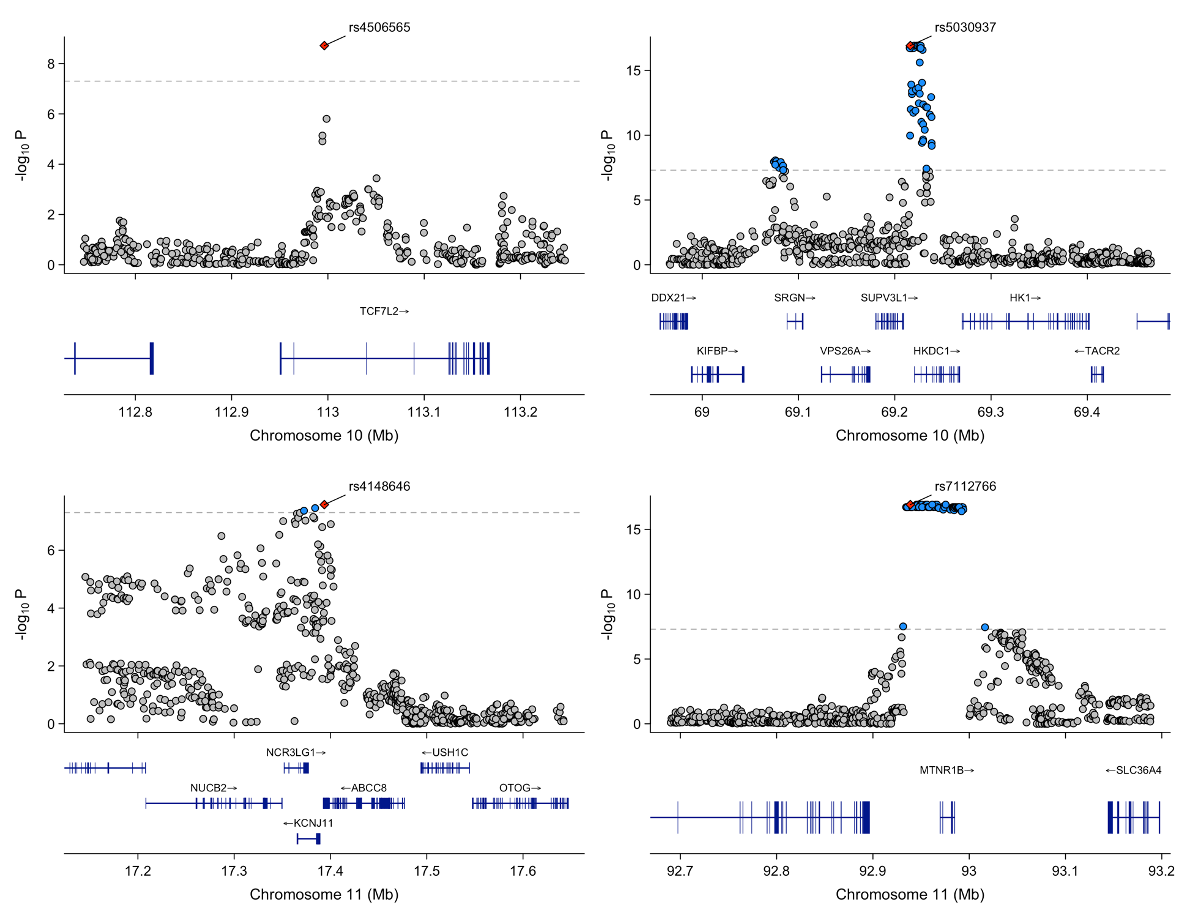

**Supplementary Figure 26.** *Locus zoom plot for the genomic region surrounding the index SNPs located on chromosomes 10 and 11, reaching genome-wide significance in the maternal multi-ancestry GWAS meta-analysis of 2hG.* Each point represents a variant from the meta-analysis, with the red diamond indicating the independent lead variant. Blue variants represent nearby variants also reaching genome-wide significance in the meta-analysis. Variants were plotted with their conditional p-value (on a -log10 scale) as a function of genomic position (GRCh38). 2hG; 2-hour glucose post-oral glucose tolerance test.

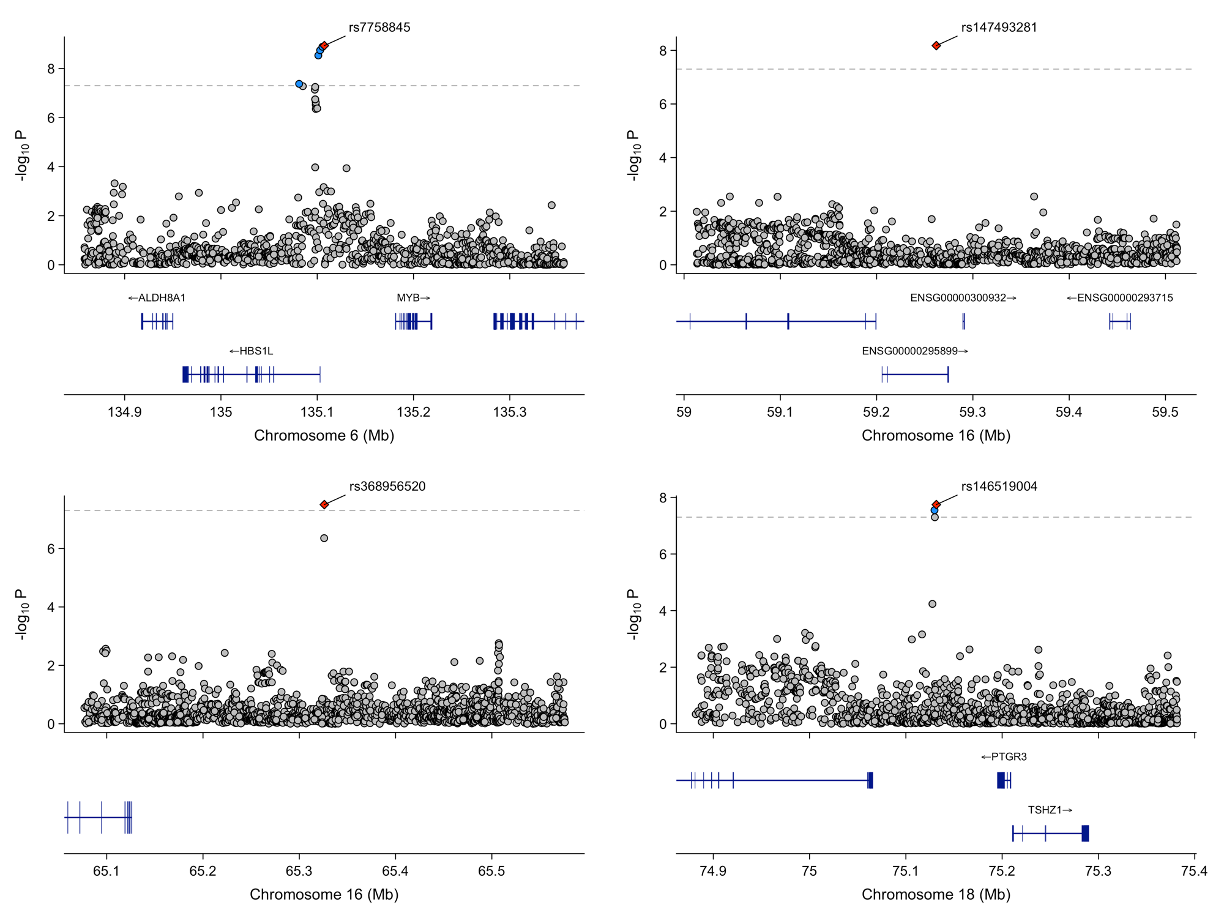

**Supplementary Figure 27.** *Locus zoom plot for the genomic region surrounding the index SNPs located on chromosomes 6, 16 and 18 reaching genome-wide significance in the maternal multi-ancestry GWAS meta-analysis of HbA1c.* Each point represents a variant from the meta-analysis, with the red diamond indicating the independent lead variant. Blue variants represent nearby variants also reaching genome-wide significance in the meta-analysis. Variants were plotted with their conditional p-value (on a -log10 scale) as a function of genomic position (GRCh38).

### **2.0 Fetal GWAS meta-analyses**

#### 2.1 Multi-ancestry GWAS meta-analyses

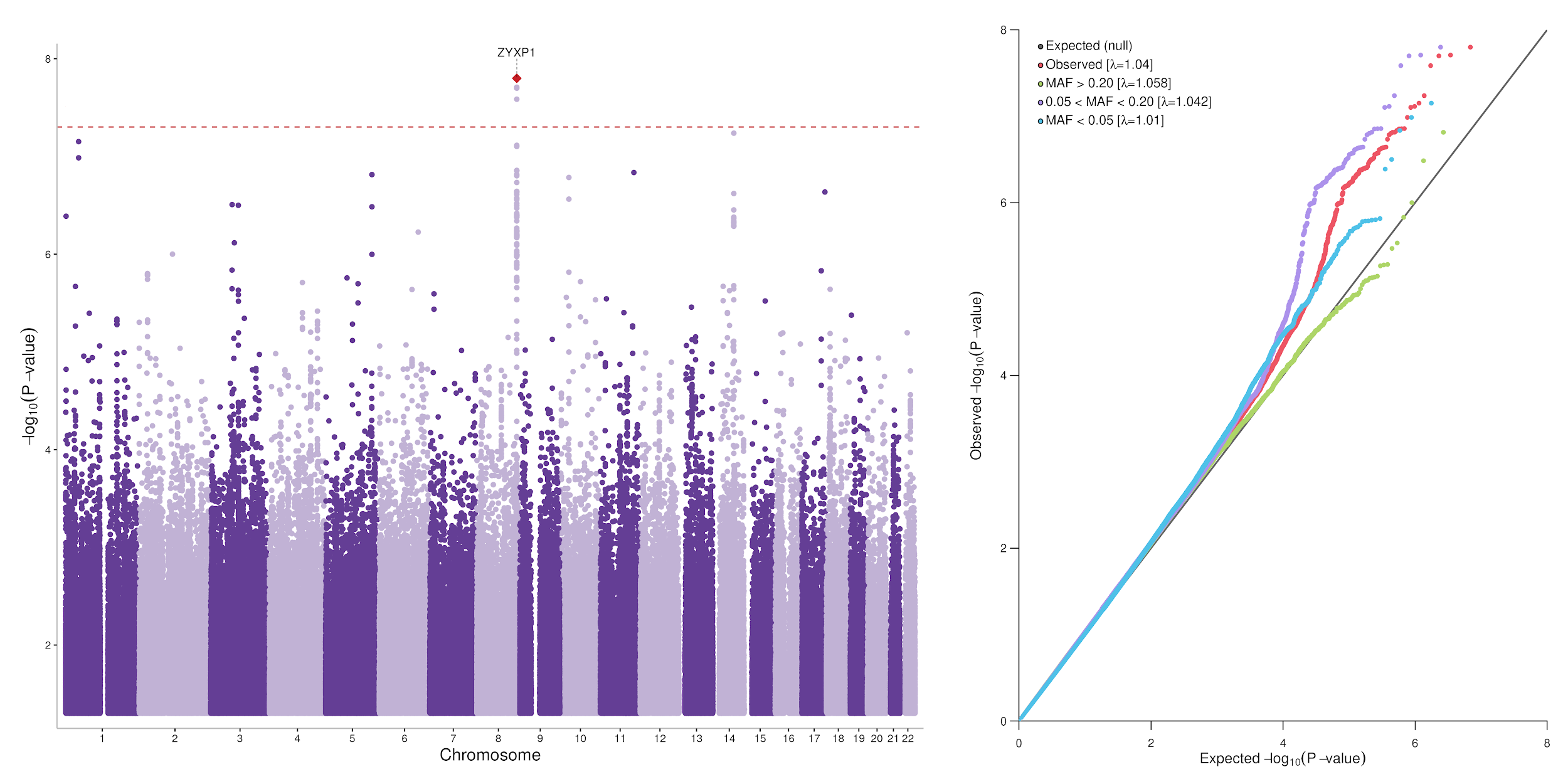

**Supplementary Figure 28.** *Manhattan plot and QQ plot of the results for the fetal GWAS meta-analysis of maternal GDM including 3,126 cases and 90,877 controls.* Only SNPs with P-value < 0.05 are plotted on the Manhattan plot. The horizontal red dashed line represents genome-wide significance (P=5×10^-8^). GDM; Gestational Diabetes Mellitus.

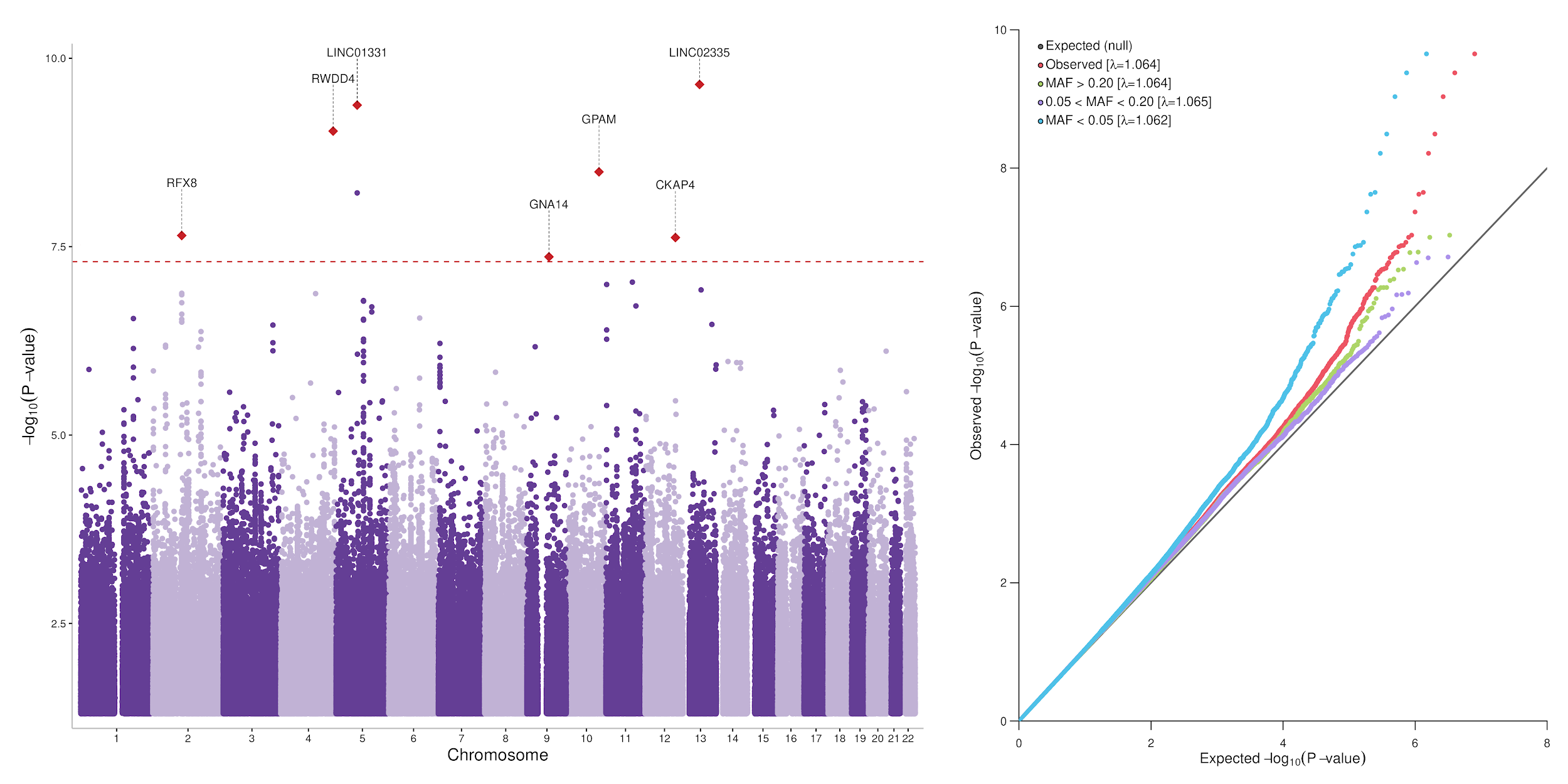

**Supplementary Figure 29.** *Manhattan plot and QQ plots of the results for the fetal GWAS meta-analysis of maternal FG including 15,855 offspring.* Only SNPs with P-value < 0.05 are plotted on the Manhattan plot. The horizontal red dashed line represents genome-wide significance (P=5×10^-8^). FG; Fasting Glucose.

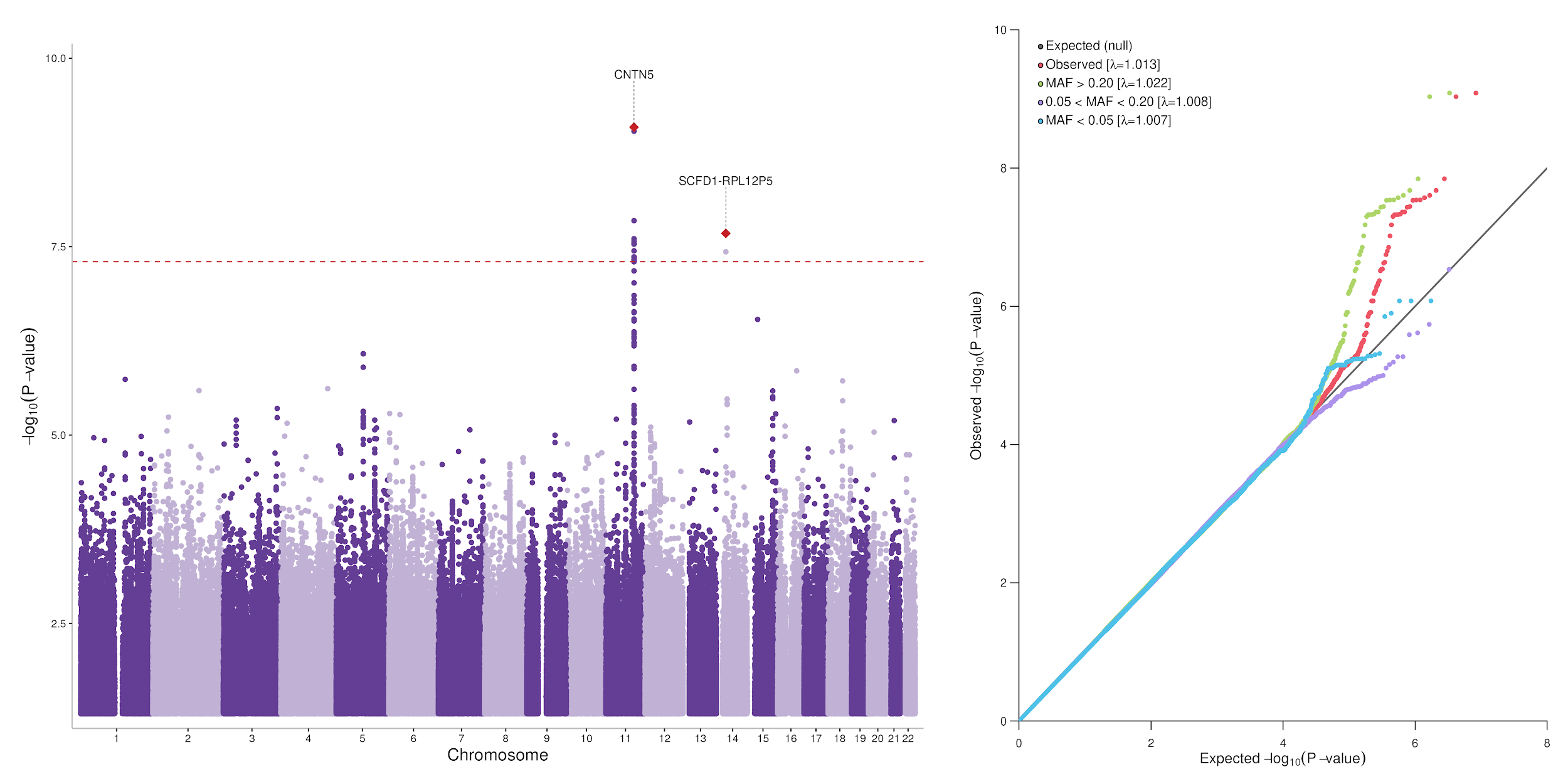

**Supplementary Figure 30.** *Manhattan plot and QQ plots of the results for the fetal GWAS meta-analysis of maternal 1hG including 9,365 offspring.* Only SNPs with P-value < 0.05 are plotted on the Manhattan plot. The horizontal red dashed line represents genome-wide significance (P=5×10^-8^). 1hG; 1-hour glucose post-oral glucose tolerance test.

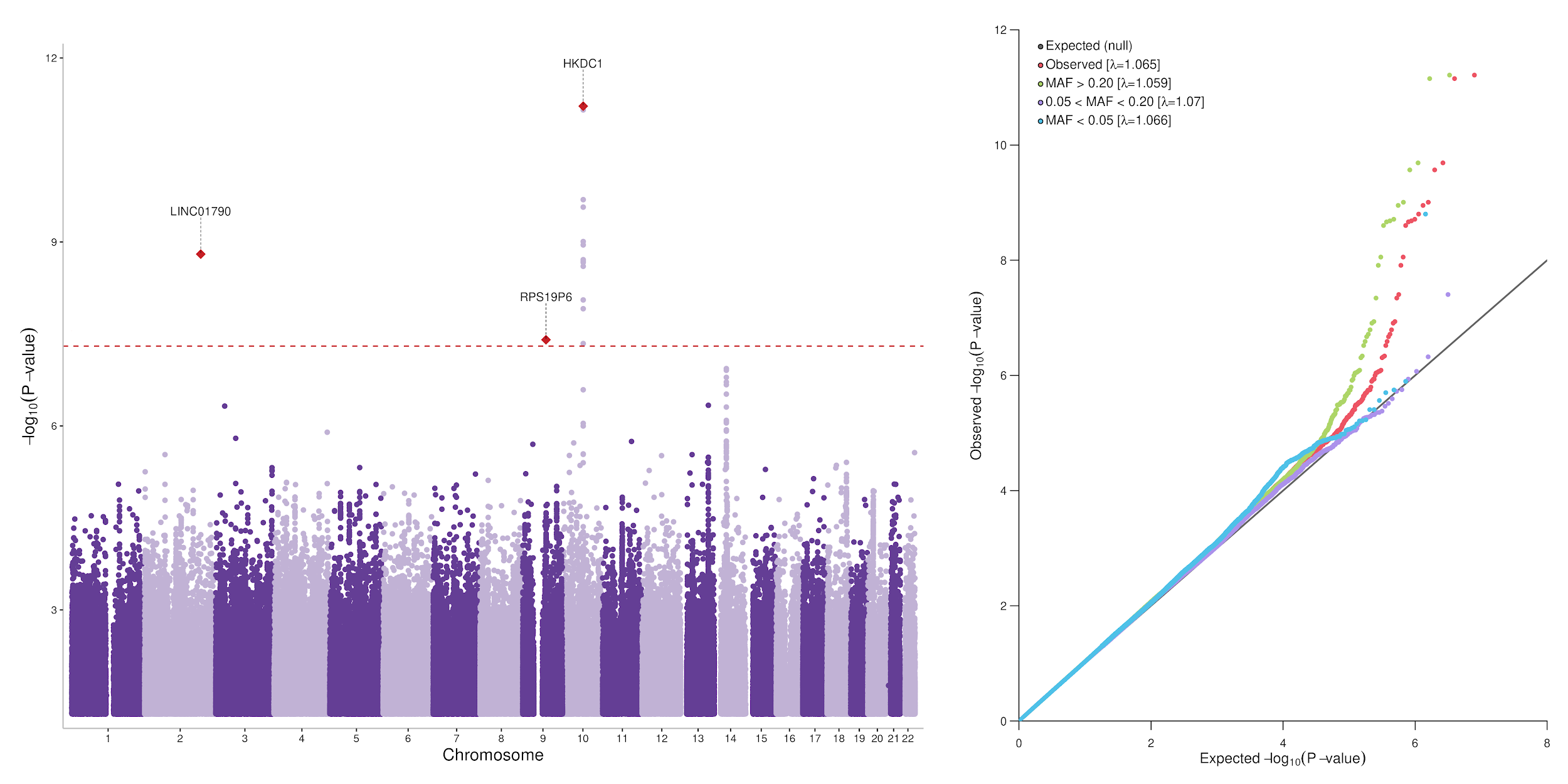

**Supplementary Figure 31.** *Manhattan plot and QQ plots of the results for the fetal GWAS meta-analysis of maternal 2hG including 15,046 pregnant women.* Only SNPs with P-value < 0.05 are plotted on the Manhattan plot. The horizontal red dashed line represents genome-wide significance (P=5×10^-8^). 2hG; 2-hour glucose post-oral glucose tolerance test.

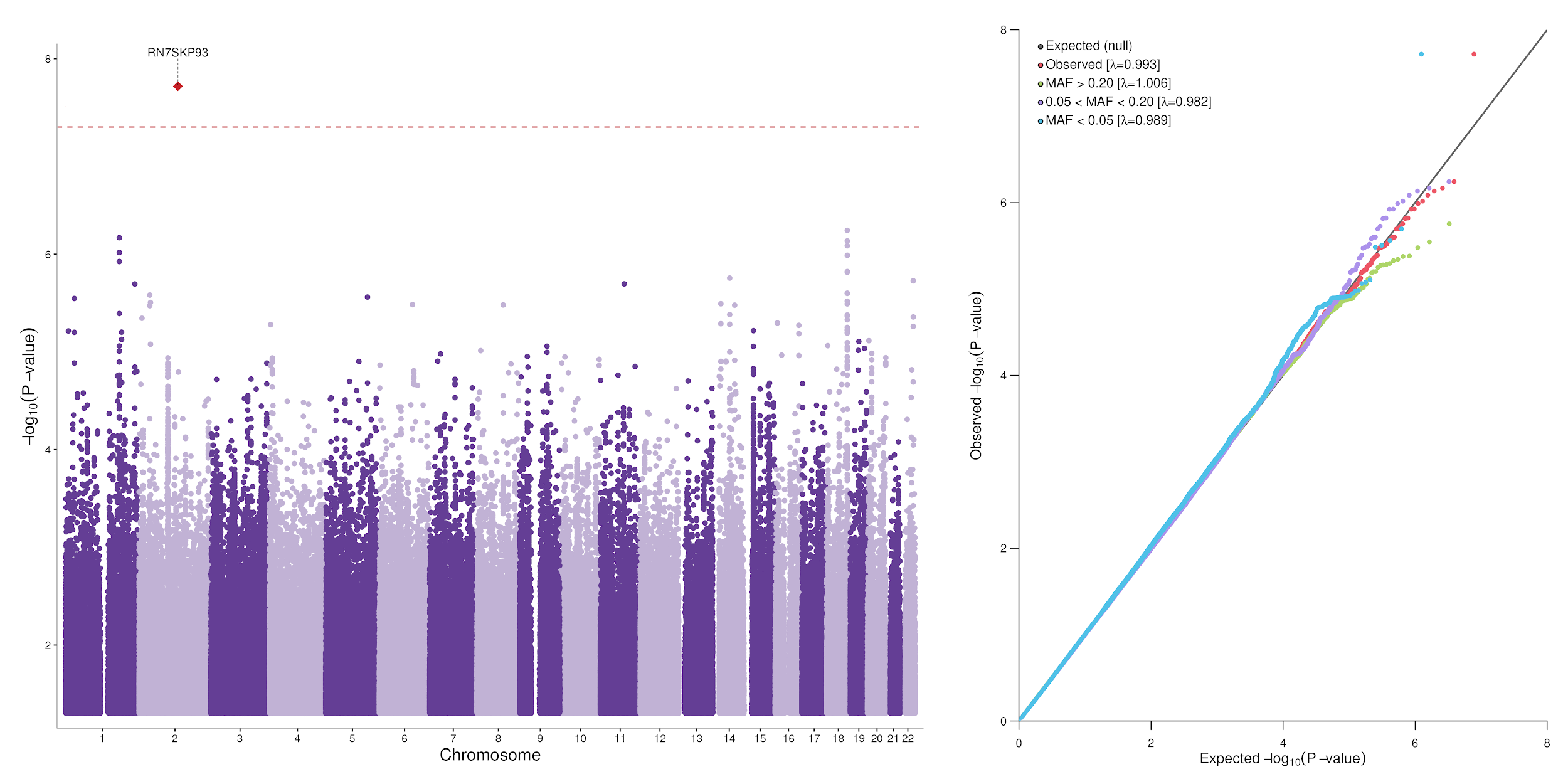

**Supplementary Figure 32.** *Manhattan plot and QQ plots of the results for the fetal GWAS meta-analysis of maternal HbA1c, including 7,035 pregnant women.* Only SNPs with P-value < 0.05 are plotted on the Manhattan plot. The horizontal red dashed line represents genome-wide significance (P=5×10^-8^).

#### 2.2 Axes of genetic variation separating GWAS

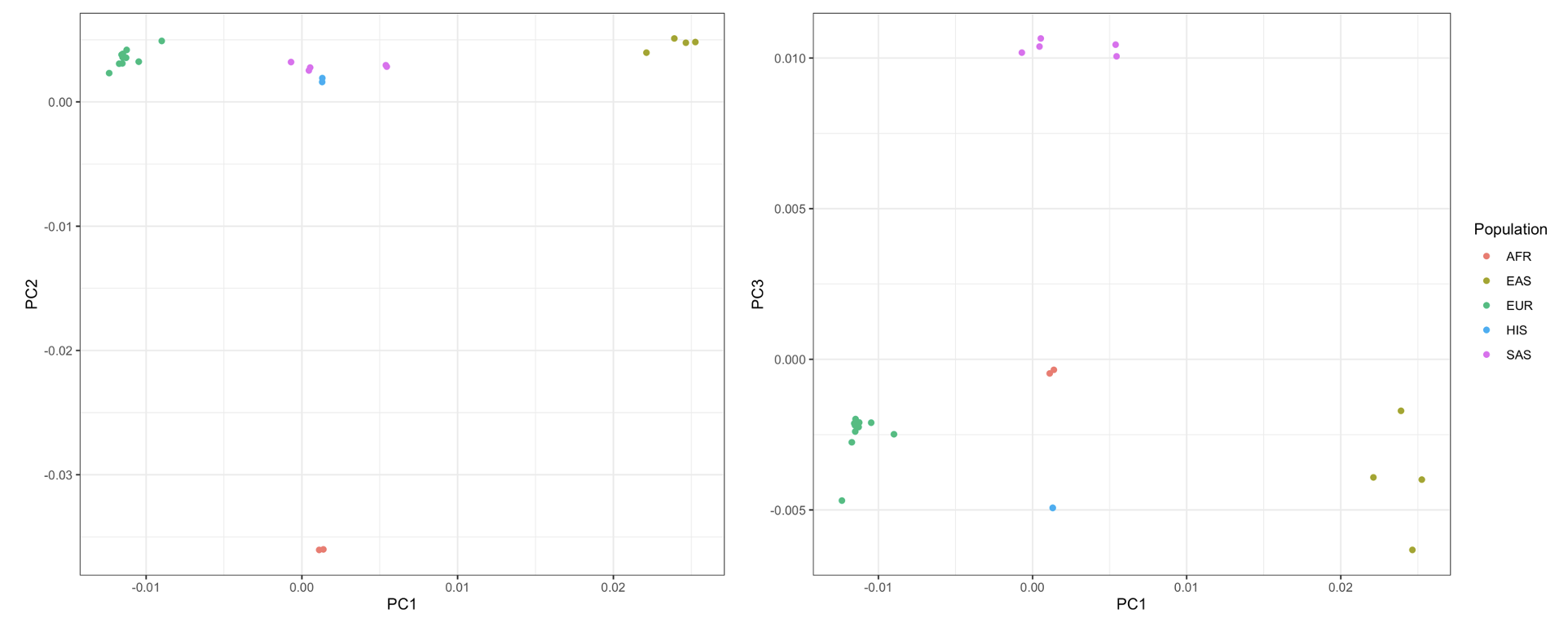

**Supplementary Figure 33.** *Axes of genetic variation separating fetal GWAS of maternal GDM.* The first two three of genetic variation from multi-dimensional scaling of the Euclidean distance matrix between GWAS are sufficient to separate five ancestry groups: South Asians (SAS), African American (AFR), Hispanic/Latino (HIS), East Asian (EAS) and European (EUR). GDM; Gestational Diabetes Mellitus.

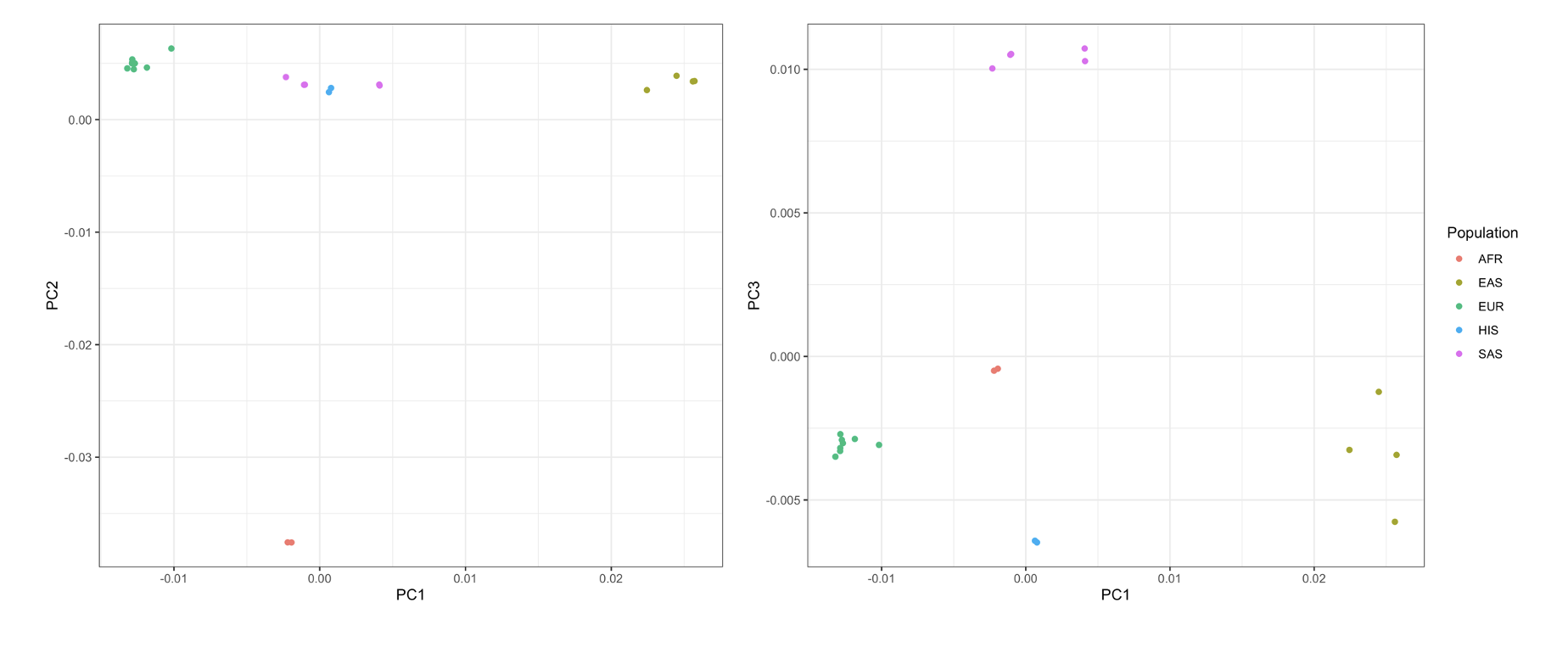

**Supplementary Figure 34.** *Axes of genetic variation separating fetal GWAS of maternal FG*. The first three axes of genetic variation from multi-dimensional scaling of the Euclidean distance matrix between GWAS are sufficient to separate five ancestry groups: South Asians (SAS), African American (AFR), Hispanic/Latino (HIS), East Asian (EAS) and European (EUR). FG: Fasting Glucose.

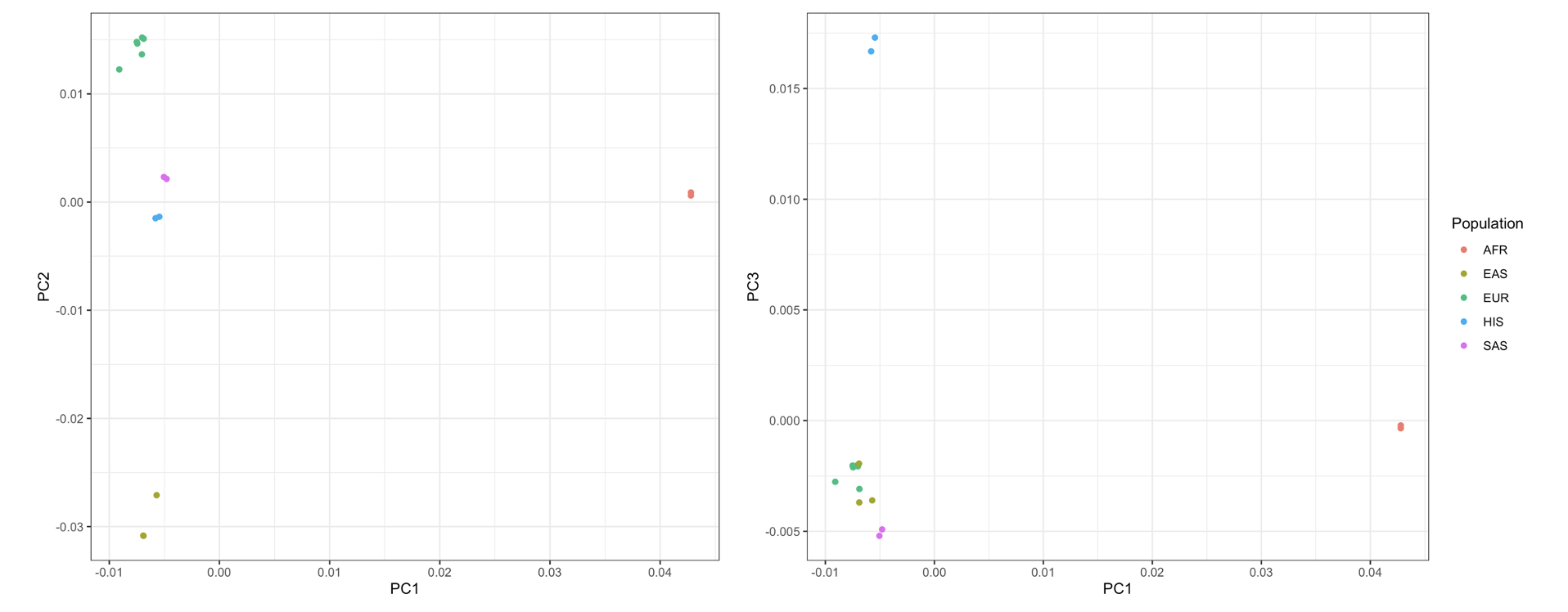
**Supplementary Figure 35.** *Axes of genetic variation separating fetal GWAS of maternal 1hG.* The first three axes of genetic variation from multi-dimensional scaling of the Euclidean distance matrix between GWAS are sufficient to separate five ancestry groups: South Asians (SAS), African American (AFR), Hispanic/Latino (HIS), East Asian (EAS) and European (EUR). 1hG; 1-hour glucose post-oral glucose tolerance test.

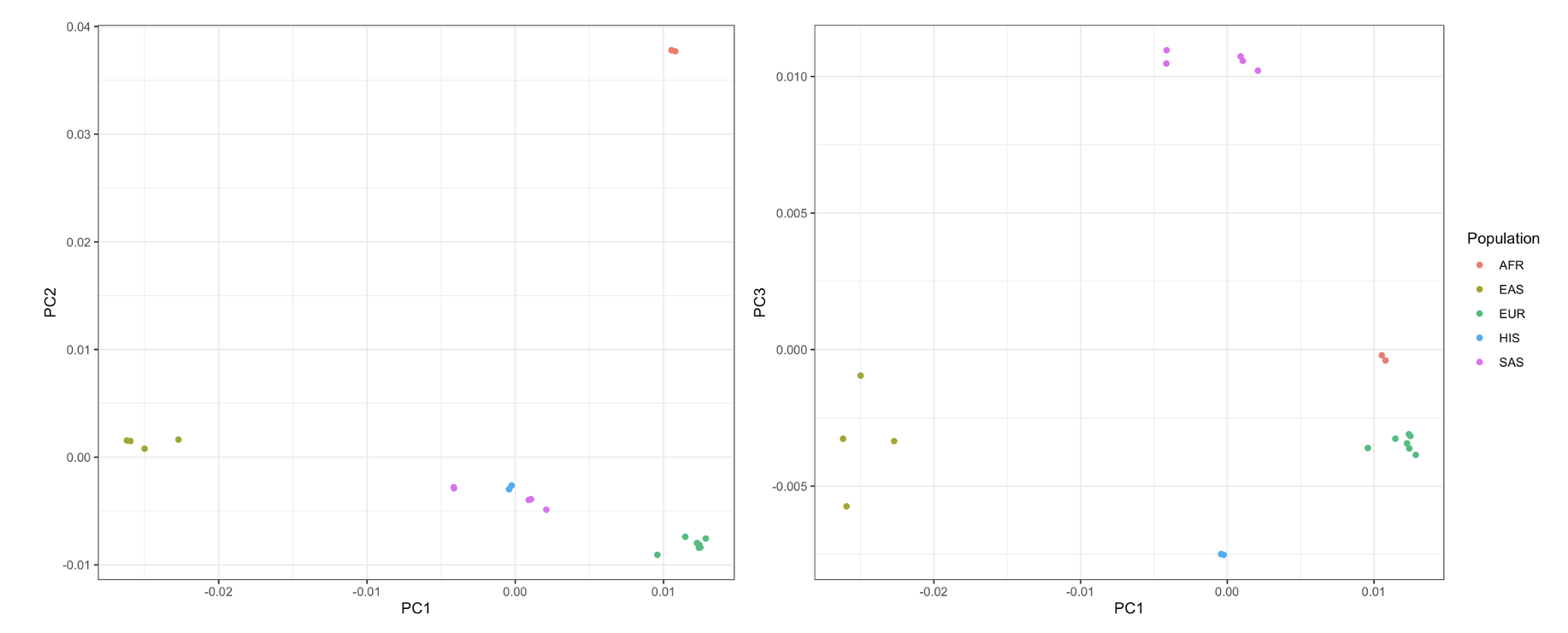
**Supplementary Figure 36.** *Axes of genetic variation separating fetal GWAS of maternal 2hG.* The first three axes of genetic variation from multi-dimensional scaling of the Euclidean distance matrix between GWAS are sufficient to separate five ancestry groups: South Asians (SAS), African American (AFR), Hispanic/Latino (HIS), East Asian (EAS) and European (EUR). 2hG; 2-hour glucose post-oral glucose tolerance test.

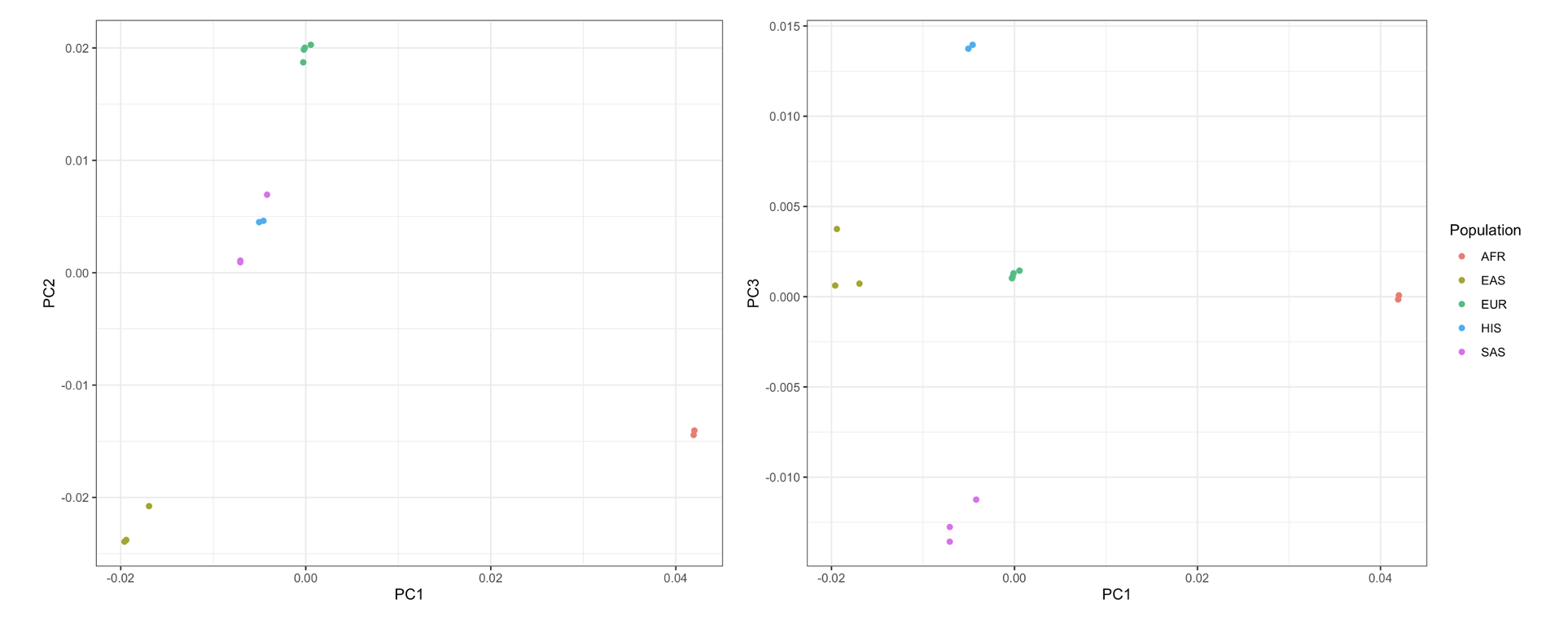
**Supplementary Figure 37.** *Axes of genetic variation separating fetal GWAS of maternal HbA1c*. The first three axes of genetic variation from multi-dimensional scaling of the Euclidean distance matrix between GWAS are sufficient to separate five ancestry groups: South Asians (SAS), African American (AFR), Hispanic/Latino (HIS), East Asian (EAS) and European (EUR).

#### 2.3 Conditional analyses using DINGO

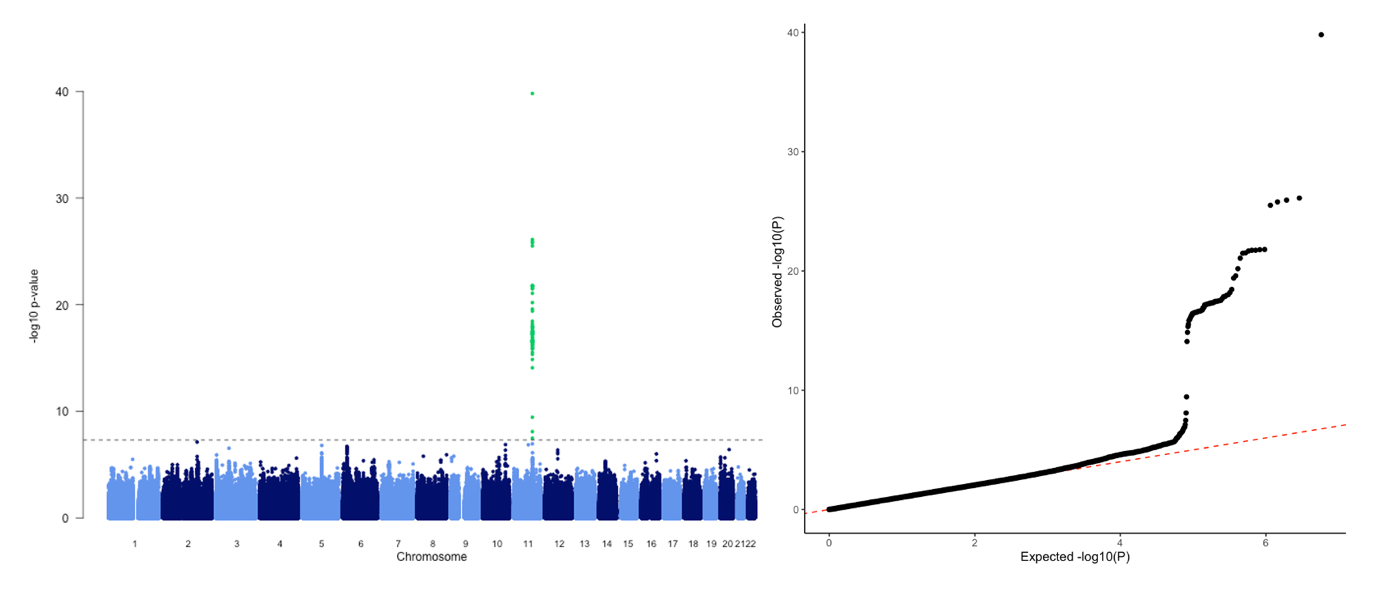

**Supplementary Figure 38.** *Manhattan plot (left) and QQ plot (right) of the results for the maternal adjusted meta-analysis of gestational diabetes mellitus (GDM) conducted using DINGO.* Plot shows maternal effects after adjusting for fetal genotype. Calculated λ=1.047.

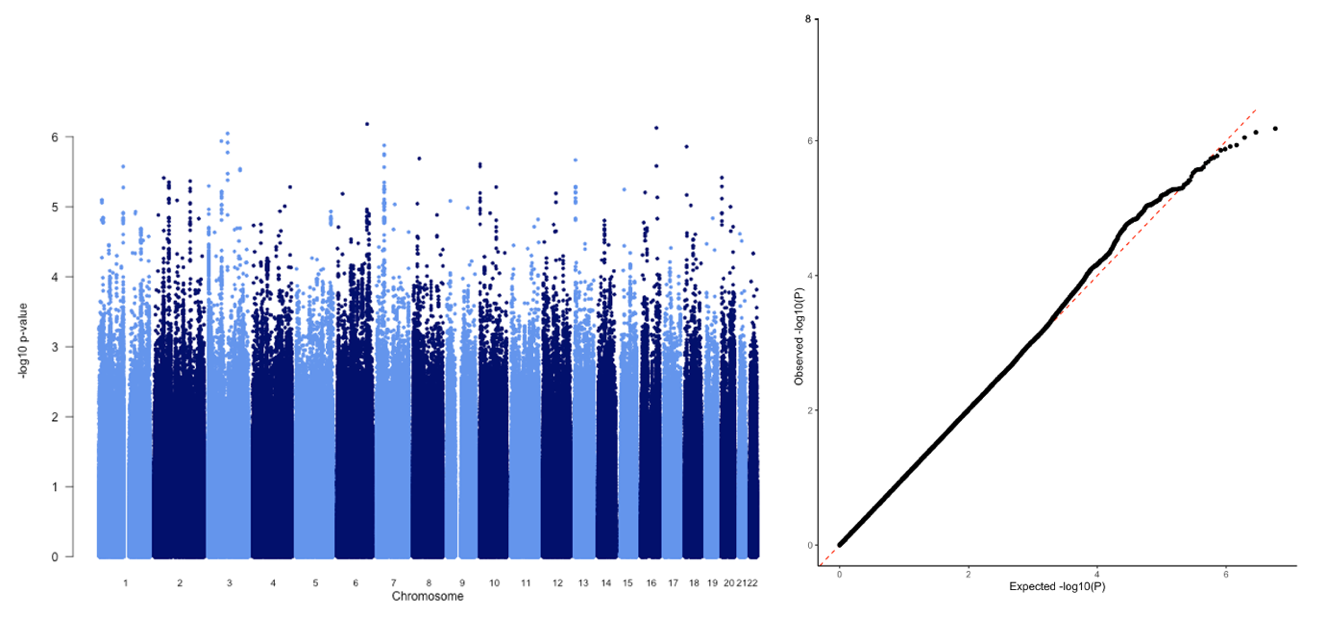

**Supplementary Figure 39.** *Manhattan plot (left) and QQ plot (right) of the results for the fetal adjusted meta-analysis of gestational diabetes mellitus (GDM) conducted using DINGO.* Plot shows fetal effects after adjusting for maternal genotype. Calculated λ=1.014.

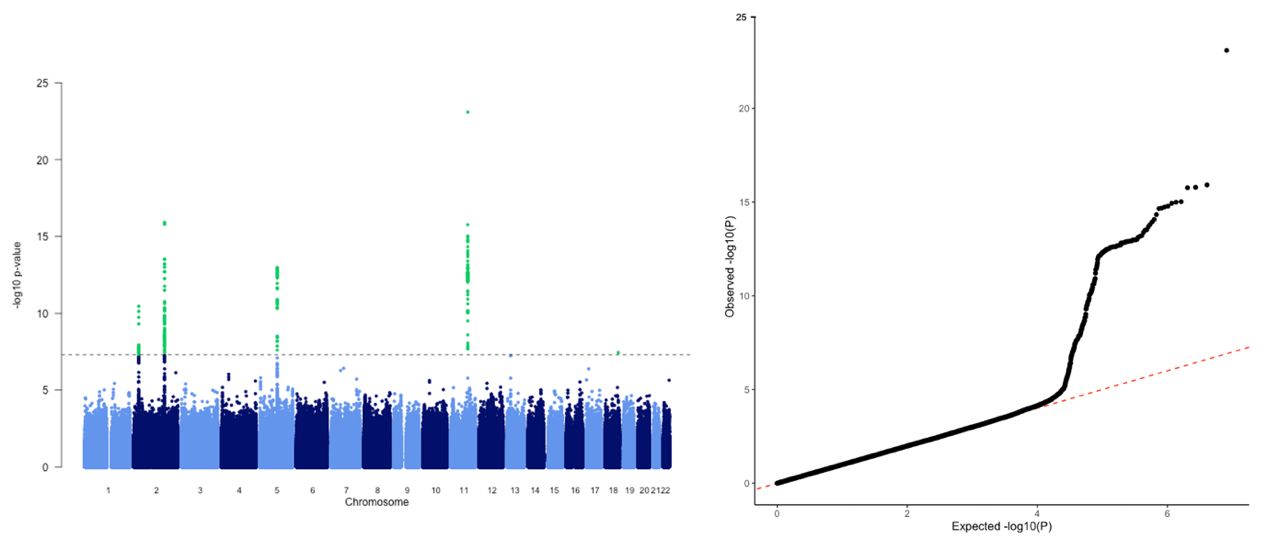

**Supplementary Figure 40.** *Manhattan plot (left) and QQ plot (right) of the results for the maternal adjusted meta-analysis of fasting glucose conducted using DINGO.* Plot shows maternal effects after adjusting for fetal genotype. Calculated λ=1.041.

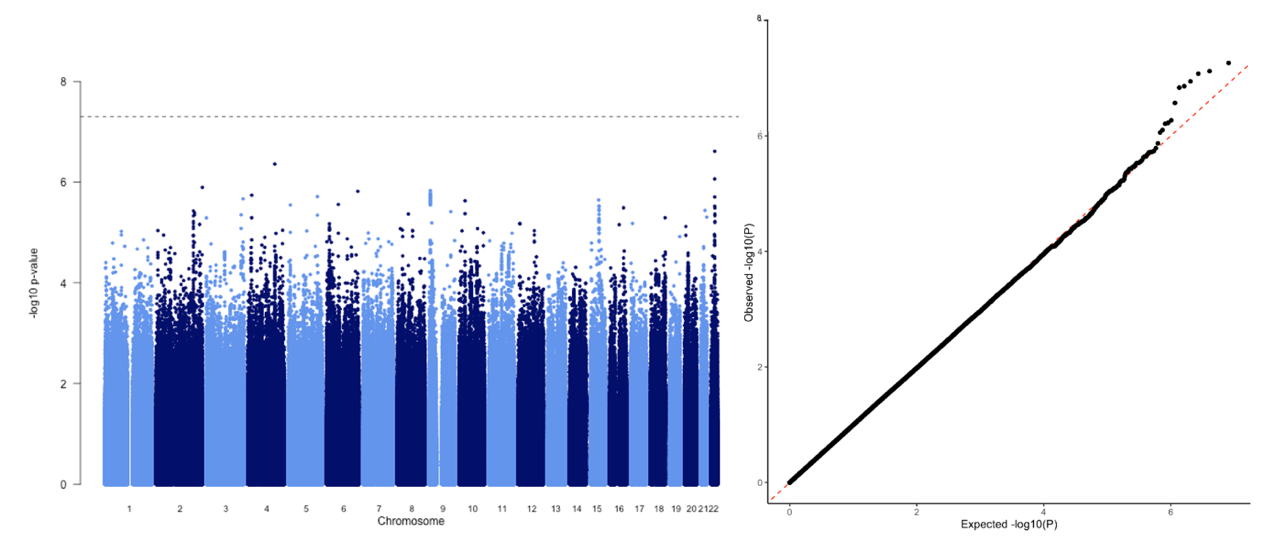

**Supplementary Figure 41.** *Manhattan plot (left) and QQ plot (right) of the results for the fetal adjusted meta-analysis of fasting glucose conducted using DINGO.* Plot shows fetal effects after adjusting for maternal genotype. Calculated λ=0.989.

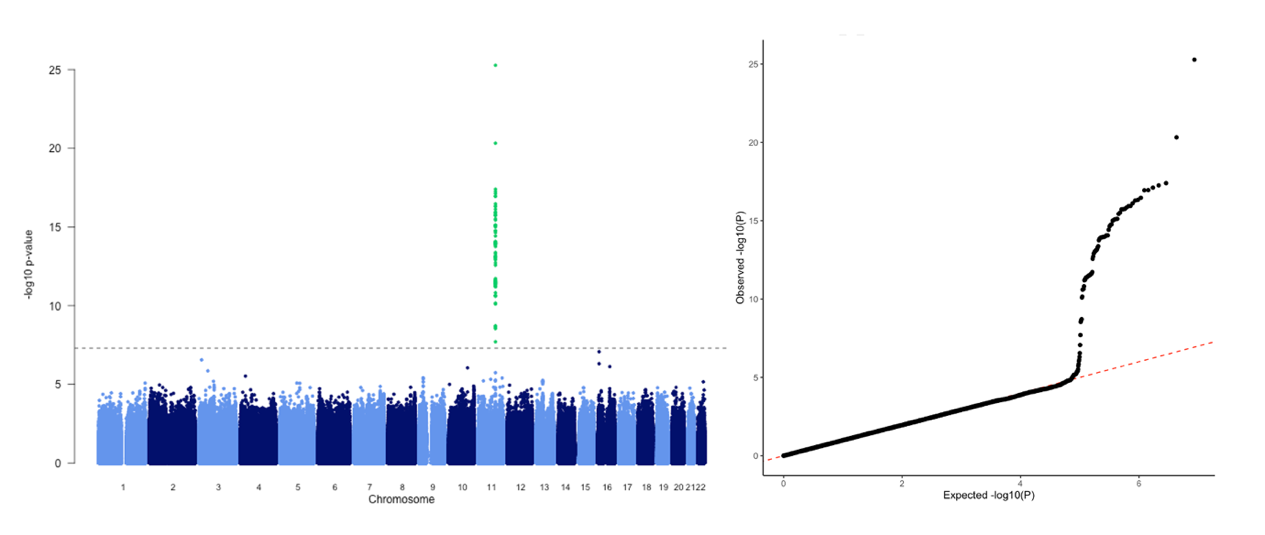

**Supplementary Figure 42.** *Manhattan plot (left) and QQ plot (right) of the results for the maternal adjusted meta-analysis of 1-hour glucose post-OGTT conducted using DINGO.* Plot shows maternal effects after adjusting for fetal genotype. Calculated λ=1.023.

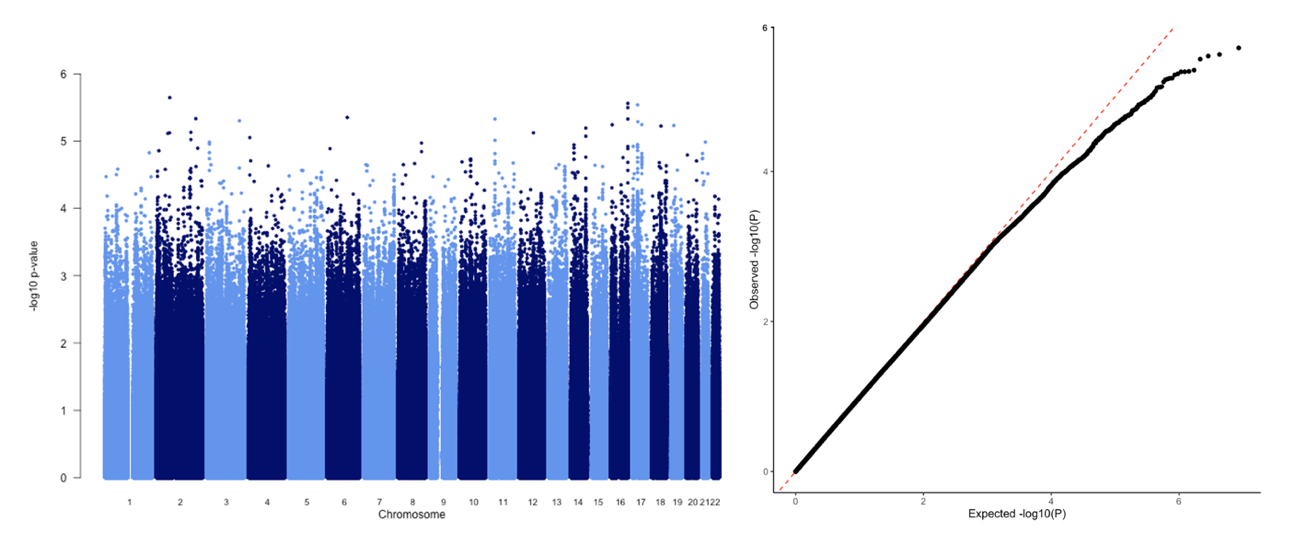

**Supplementary Figure 43.** *Manhattan plot (left) and QQ plot (right) of the results for the fetal adjusted meta-analysis of 1-hour glucose post-OGTT conducted using DINGO.* Plot shows fetal effects after adjusting for maternal genotype. Calculated λ=0.991.

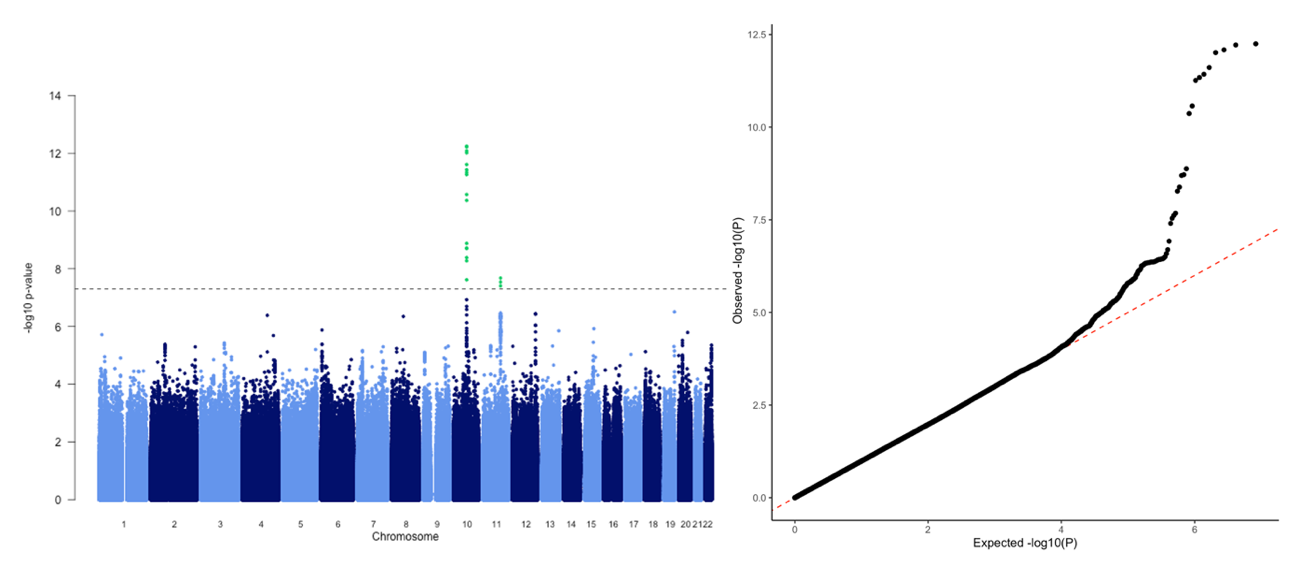

**Supplementary Figure 44.** *Manhattan plot (left) and QQ plot (right) of the results for the maternal adjusted meta-analysis of 2-hour glucose post-OGTT conducted using DINGO.* Plot shows maternal effects after adjusting for fetal genotype. Calculated λ=1.043.

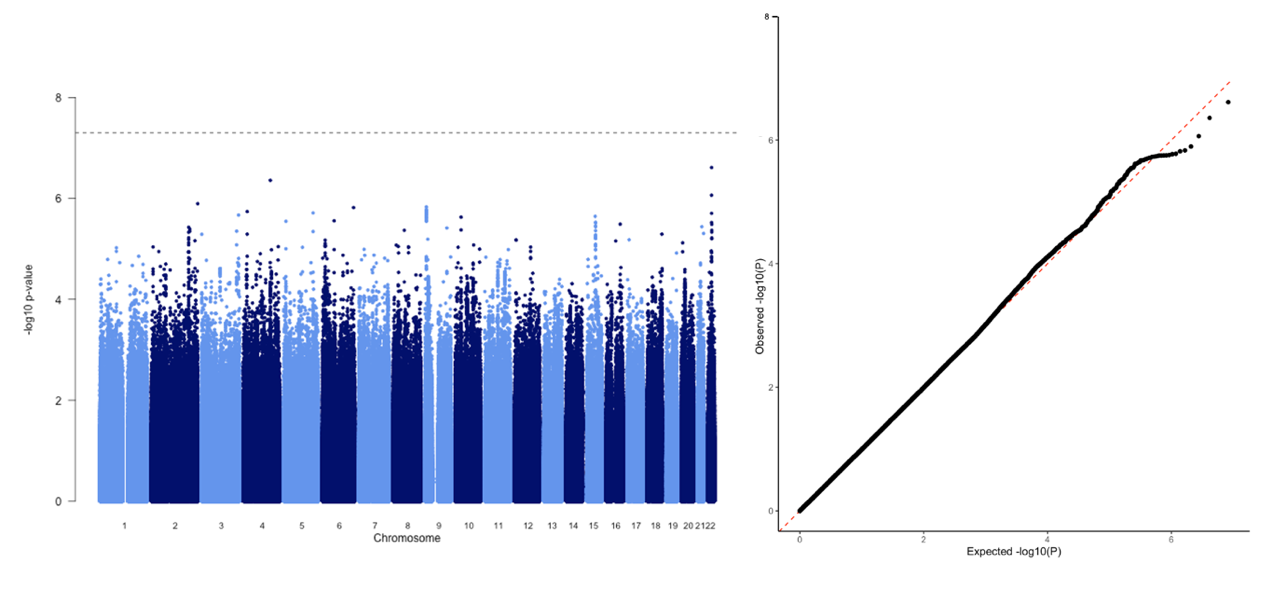

**Supplementary Figure 45.** *Manhattan plot (left) and QQ plot (right) of the results for the fetal adjusted meta-analysis of 2-hour glucose post-OGTT conducted using DINGO.* Plot shows fetal effects after adjusting for maternal genotype. Calculated λ=0.992.

### **3.0 Heterogeneity in allelic effects in the maternal multi-ancestry meta-analyses**

#### 3.1 Ancestry-specific variation in allelic effects

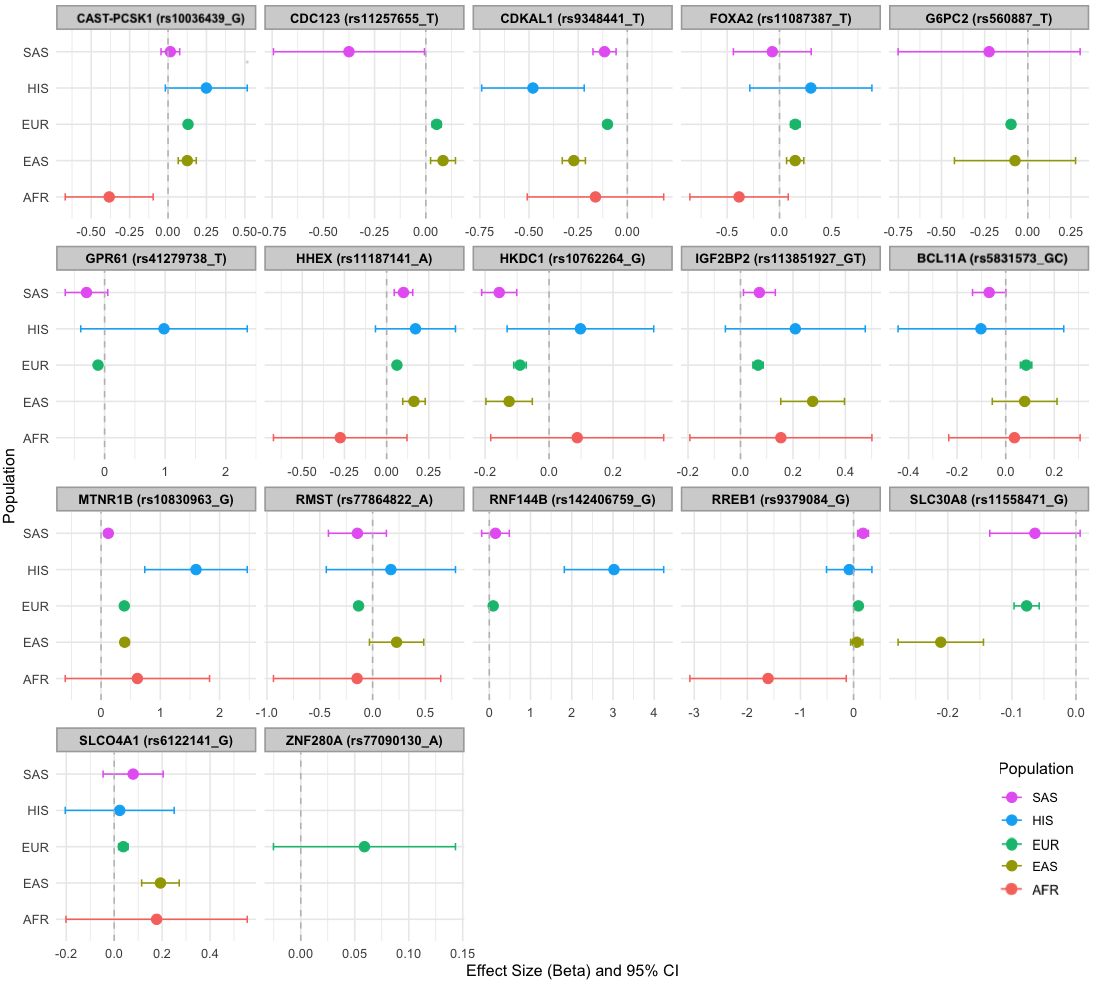

**Supplementary Figure 46.** *Comparison of effect estimates across ancestry groups for lead variants showing significant heterogeneity in allelic effects in the multi-ancestry meta-analysis of maternal GDM.* In MR-MEGA, a test of heterogeneity in effect sizes across ancestries was performed and variants where P<0.05 are plotted in this figure. The putative gene name is also provided for each lead variant (listed in parentheses alongside the effect allele used for plotting). Effect estimates and corresponding 95% confidence intervals (CI) were only plotted for variants if they were present in the ancestry-specific GWAS meta-analysis. The rs77090130 variant was solely added for reference, as it was only present in Europeans. GDM; Gestational Diabetes Mellitus. AFR; African. EAS; East Asian. EUR; European. SAS; South Asian. HIS; Hispanic.

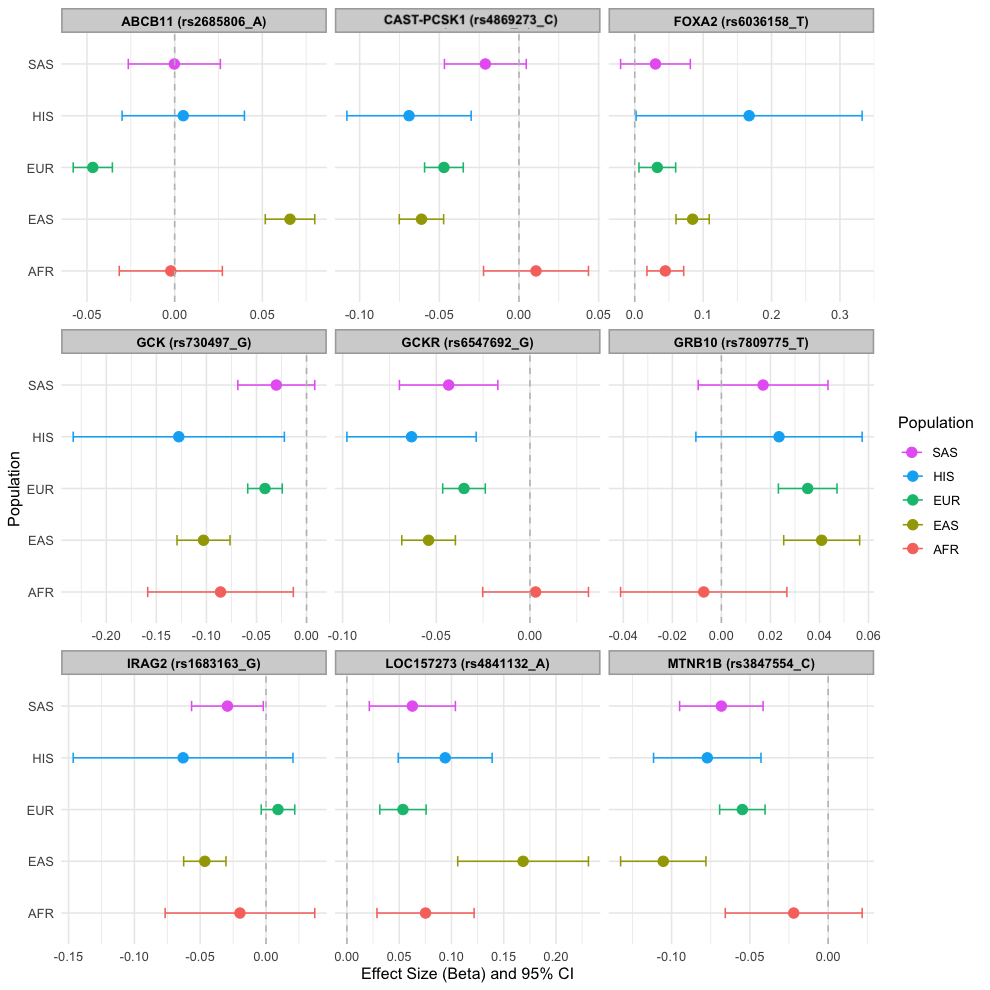

**Supplementary Figure 47.** *Comparison of effect estimates across ancestry groups for lead variants showing significant heterogeneity in allelic effects in the multi-ancestry meta-analysis of maternal FG.* In MR-MEGA, a test of heterogeneity in effect sizes across ancestries was performed and variants where P<0.05 are plotted in this figure. The putative gene name is also provided for each lead variant (listed in parentheses alongside the effect allele used for plotting). Effect estimates and corresponding 95% confidence intervals (CI) were only plotted for variants if they were present in the ancestry-specific GWAS meta-analysis. FG; Fasting Glucose. AFR; African. EAS; East Asian. EUR; European. SAS; South Asian. HIS; Hispanic.

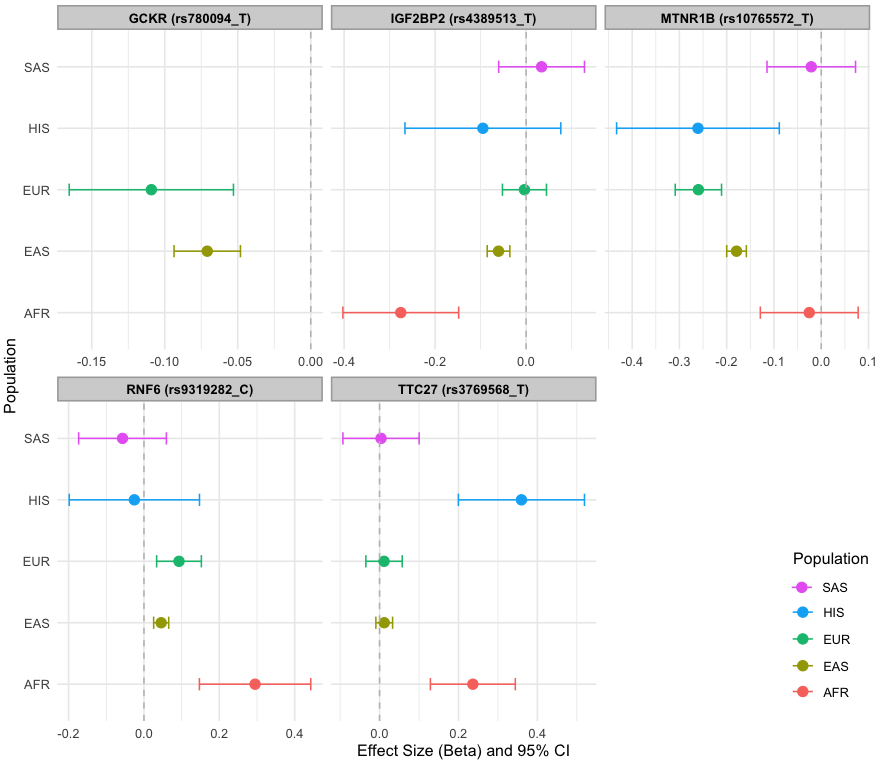

**Supplementary Figure 48.** *Comparison of effect estimates across ancestry groups for lead variants showing significant heterogeneity in allelic effects in the multi-ancestry meta-analysis of maternal 1hG.* In MR-MEGA, a test of heterogeneity in effect sizes across ancestries was performed and variants where P<0.05 are plotted in this figure. The putative gene name is also provided for each lead variant (listed in parentheses alongside the effect allele used for plotting). Effect estimates and corresponding 95% confidence intervals (CI) were only plotted for variants if they were present in the ancestry-specific GWAS meta-analysis. 1hG; 1-hour glucose post-oral glucose tolerance test. AFR; African. EAS; East Asian. EUR; European. SAS; South Asian. HIS; Hispanic.

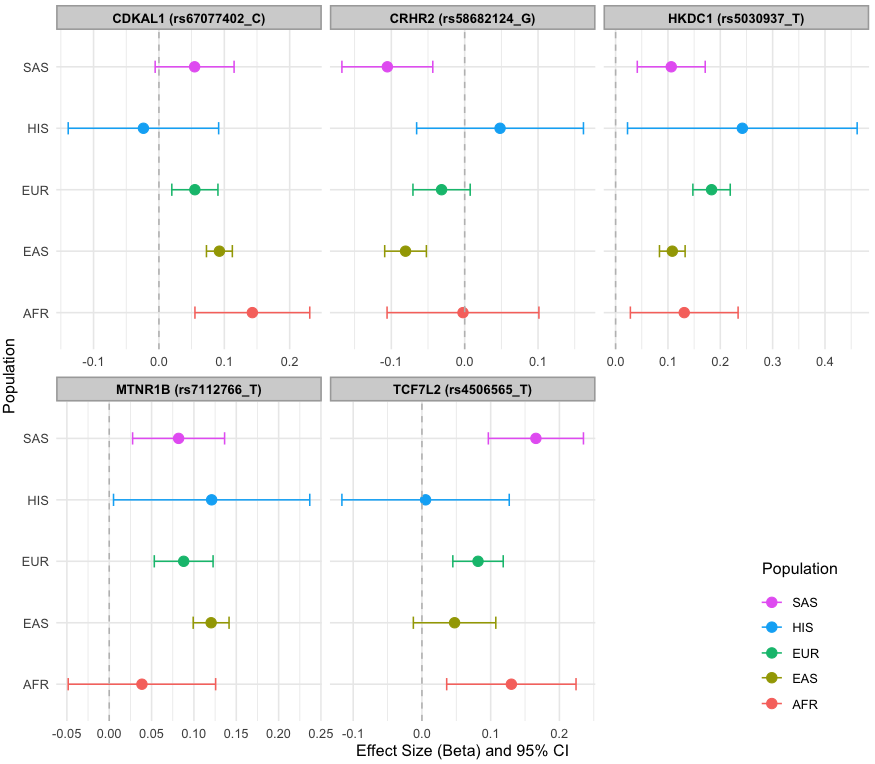

**Supplementary Figure 49.** *Comparison of effect estimates across ancestry groups for lead variants showing significant heterogeneity in allelic effects in the multi-ancestry meta-analysis of maternal 2hG.* In MR-MEGA, a test of heterogeneity in effect sizes across ancestries was performed and variants where P<0.05 are plotted in this figure. The putative gene name is also provided for each lead variant (listed in parentheses alongside the effect allele used for plotting). Effect estimates and corresponding 95% confidence intervals (CI) were only plotted for variants if they were present in the ancestry-specific GWAS meta-analysis. 2hG; 2-hour glucose post-oral glucose tolerance test. AFR; African. EAS; East Asian. EUR; European. SAS; South Asian. HIS; Hispanic.

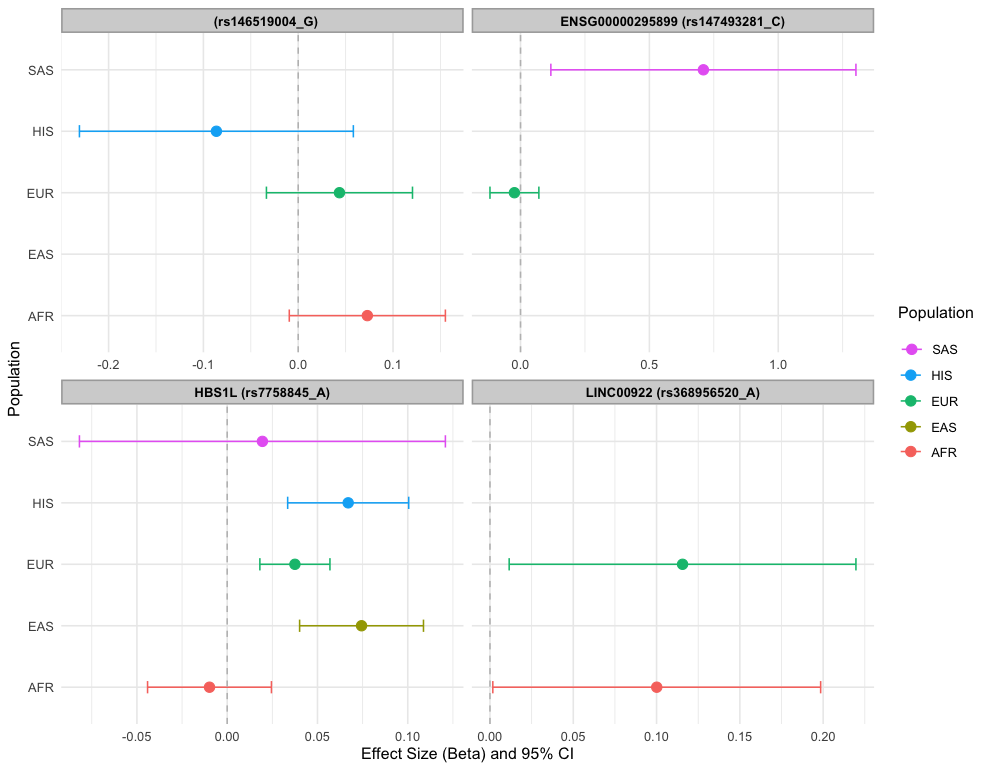

**Supplementary Figure 50.** *Comparison of effect estimates across ancestry groups for lead variants showing significant heterogeneity in allelic effects in the multi-ancestry meta-analysis of maternal HbA1c.* In MR-MEGA, a test of heterogeneity in effect sizes across ancestries was performed and variants where P<0.05 are plotted in this figure. The putative gene name is also provided for each lead variant (listed in parentheses alongside the effect allele used for plotting). Effect estimates and corresponding 95% confidence intervals (CI) were only plotted for variants if they were present in the ancestry-specific GWAS meta-analysis. AFR; African. EAS; East Asian. EUR; European. SAS; South Asian. HIS; Hispanic.

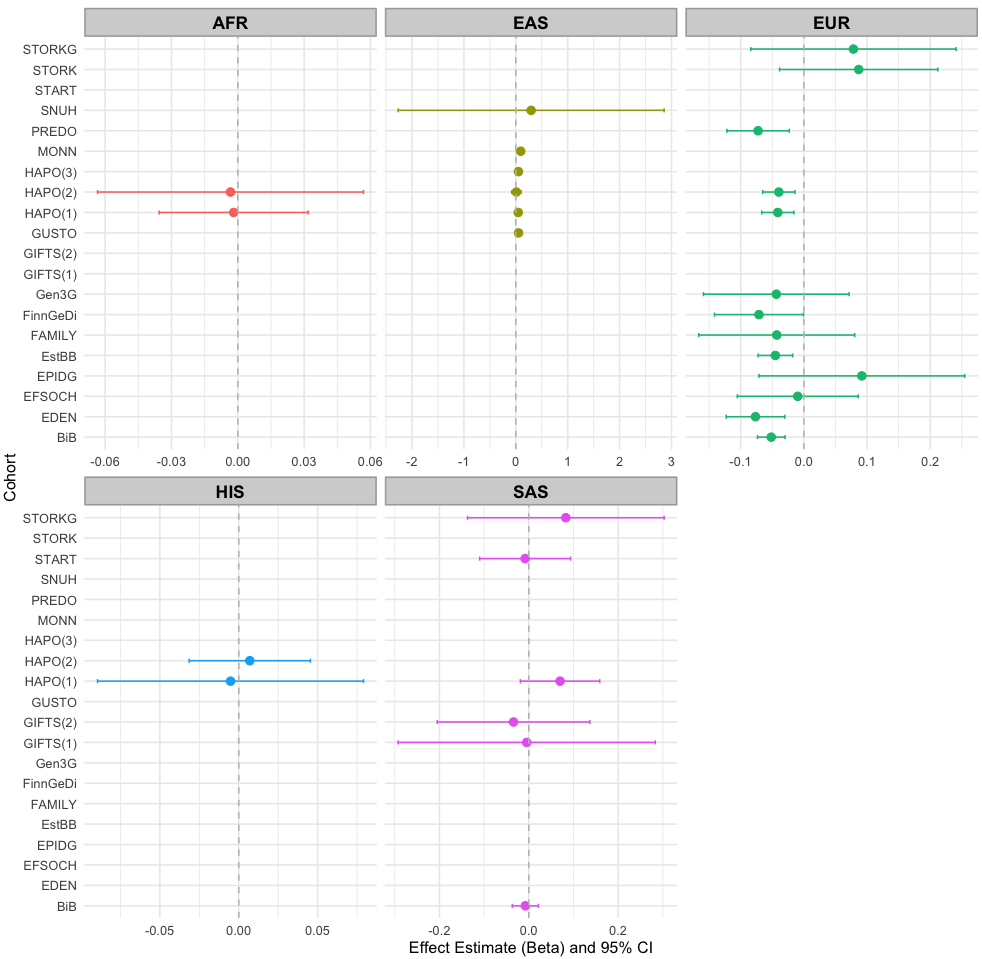

**Supplementary Figure 51.** *Comparison of the effect estimates across populations and cohorts for the variant rs2685806 (ABCB11) detected in the multi-ancestry meta-analysis of FG and showing opposite direction of effects in East Asians (EAS) and Europeans (EUR).* Effects are shown with respect to the A allele. In cases where a cohort contributed multiple files for the same population, bracketed numbers were used to distinguish them for reference purposes. FG; Fasting glucose. AFR; African. EAS; East Asian. EUR; European. SAS; South Asian. HIS; Hispanic.

#### 3.2 Associations driven by study heterogeneity

While MR-MEGA offers advantages for multi-ancestry GWAS by leveraging systematic variation in allelic effects across ancestries, we suspected some false-positive associations in our analyses. MR-MEGA models SNP effect sizes across cohorts using a meta-regression framework in which heterogeneity is decomposed into ancestry-correlated and residual (unmodeled) components. Specifically, principal coordinates derived from a multidimensional scaling (MDS) of genome-wide allele frequency differences across studies are used as covariates in the meta-regression model. These MDS components represent major axes of genetic differentiation between cohorts and are used to account for systematic ancestry-driven heterogeneity in allelic effects. However, if a variant exhibits between-cohort variation in effect size that is coincidentally aligned with one or more ancestry axes - but is not due to a genuine genetic effect - this alignment can inflate the overall test for association. This is particularly problematic for variants with low minor allele frequency or sparse representation across studies, where a small number of outlier cohorts can unduly influence the meta-regression. In such cases, MR-MEGA may identify associations that reach genome-wide significance and show extreme heterogeneity (e.g., significant ancestry-correlated heterogeneity P-values), but do not replicate in ancestry-stratified meta-analyses, suggesting potential type I error due to cohort-specific artefacts or unmodeled confounding. We provide forest plots comparing effect estimates between cohorts and across populations below (Supplementary Figures 52-55) for associations that are likely false positives.

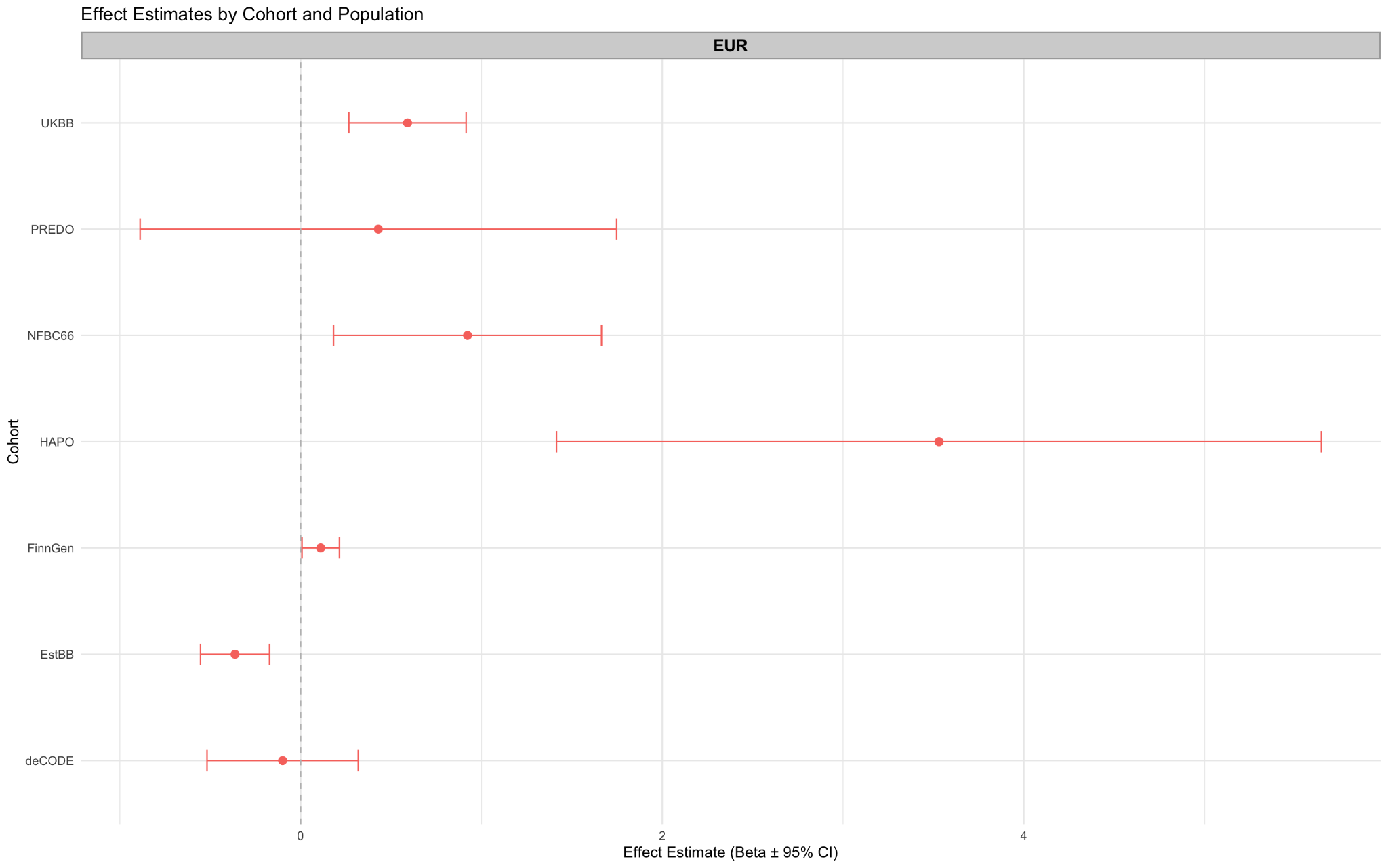

**Supplementary Figure 52.** *Comparison of the effect estimates across populations and cohorts for the variant rs77090130 detected in the multi-ancestry meta-analysis of GDM.* Effects are shown with respect to the A allele. In cases where a cohort contributed multiple files for the same population, bracketed numbers were used to distinguish them for reference purposes. Effect allele frequencies (EAF) for each study were as follows: NFBC66=0.01, PREDO=0.01, UKBB=0.01, EstBB=0.04, deCODE=0.01, HAPO=0.01, FinnGen 0.01.

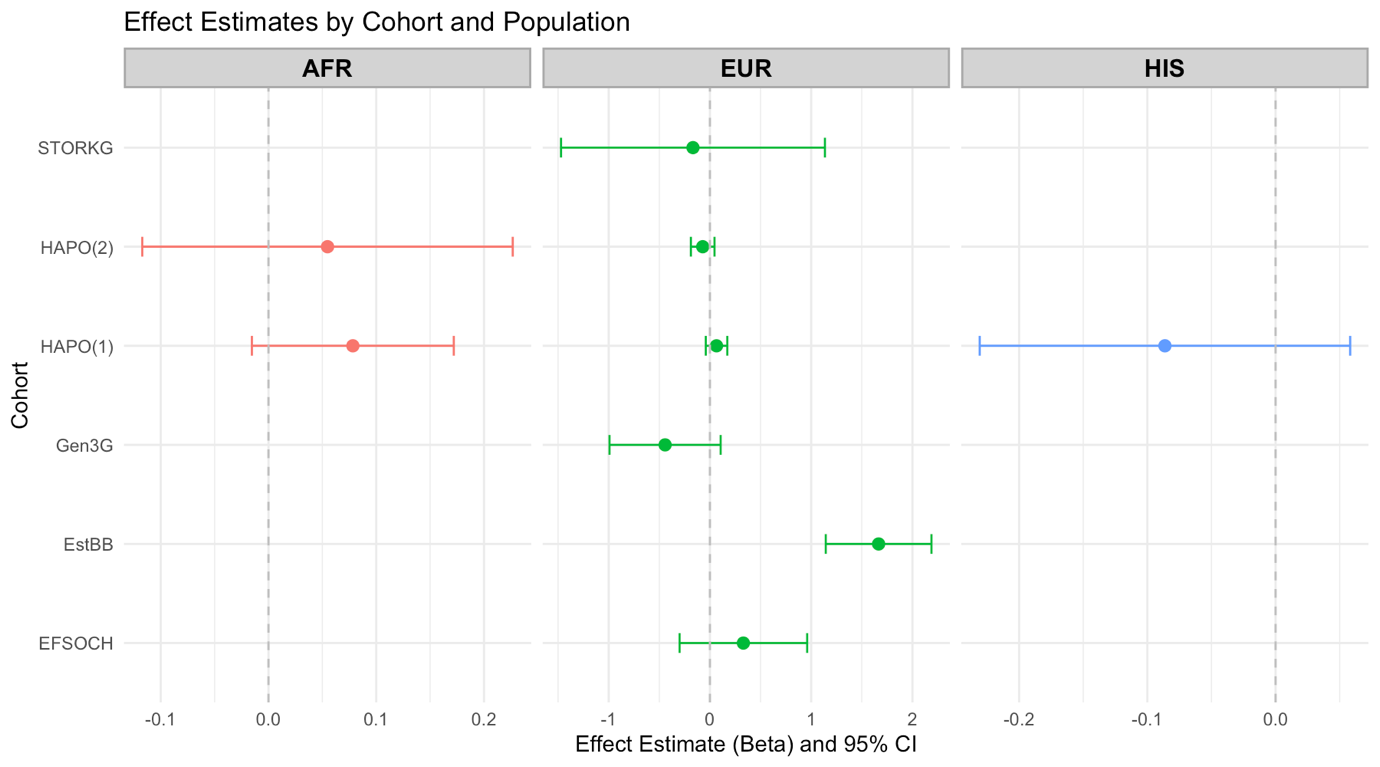

**Supplementary Figure 53.** *Comparison of the effect estimates across populations and cohorts for the variant rs146519004 detected in the multi-ancestry meta-analysis of HbA1c.* Effects are shown with respect to the G allele. In cases where a cohort contributed multiple files for the same population, bracketed numbers were used to distinguish them for reference purposes. Effect allele frequencies (EAF) for each study were as follows: STORKG=0.99, EstBB=0.99, EFSOCH=0.99, Gen3G=0.99, HAPO(1) AFR=0.96, HAPO(2), AFR=0.96, HAPO(1) EUR=0.98, HAPO(1) HIS=0.99 and HAPO(1) EUR=0.99. AFR; African. EAS; East Asian. EUR; European. SAS; South Asian. HIS; Hispanic.

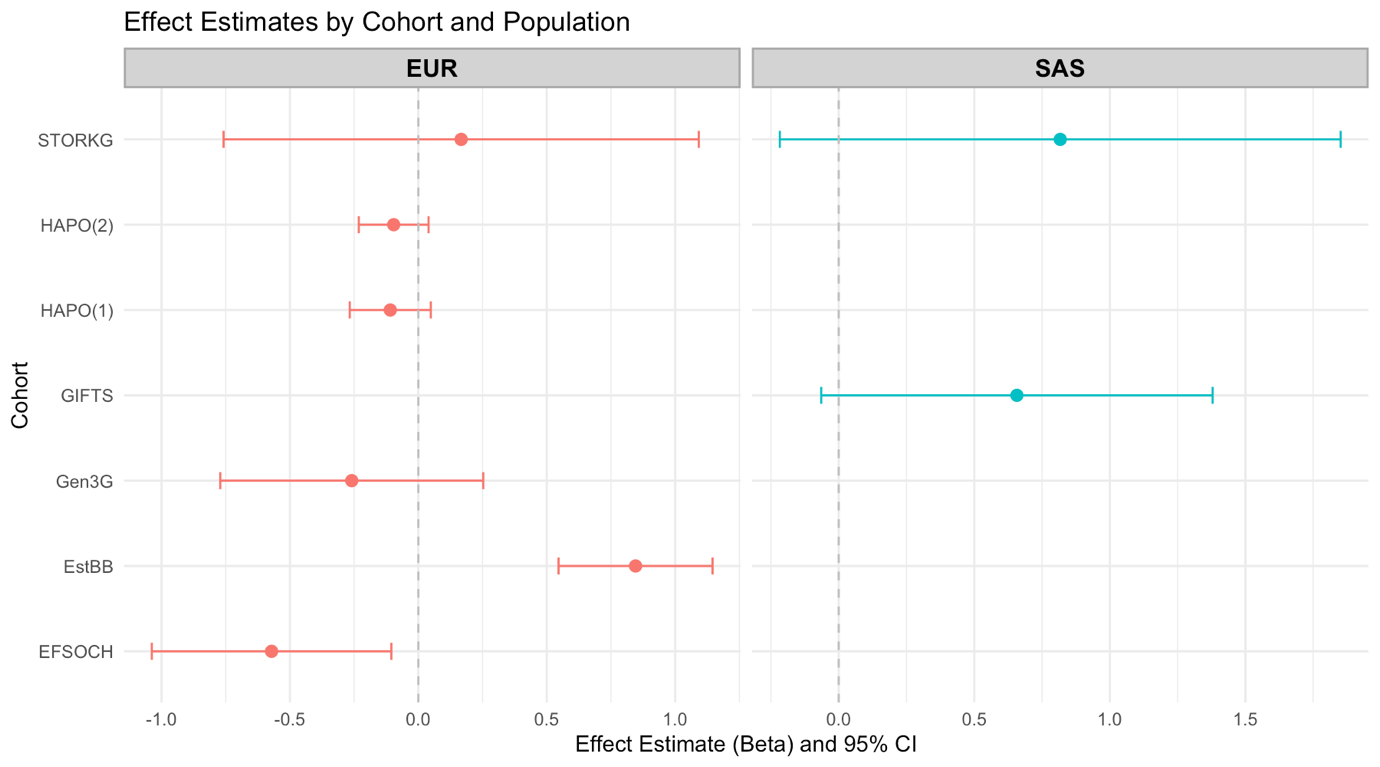

**Supplementary Figure 54.** *Comparison of the effect estimates across populations and cohorts for the variant rs147493281 detected in the multi-ancestry meta-analysis of HbA1c.* Effects are shown with respect to the C allele. In cases where a cohort contributed multiple files for the same population, bracketed numbers were used to distinguish them for reference purposes. Effect allele frequencies (EAF) for each study were as follows: STORKG EUR=0.99, STORKG SAS=0.99, GIFTS SAS=0.98, EstBB EUR=0.99, EFSOCH EUR=0.99, Gen3G EUR=0.99, HAPO(1) EUR=0.99 and HAPO(2) EUR=0.99. AFR; African. EAS; East Asian. EUR; European. SAS; South Asian. HIS; Hispanic.

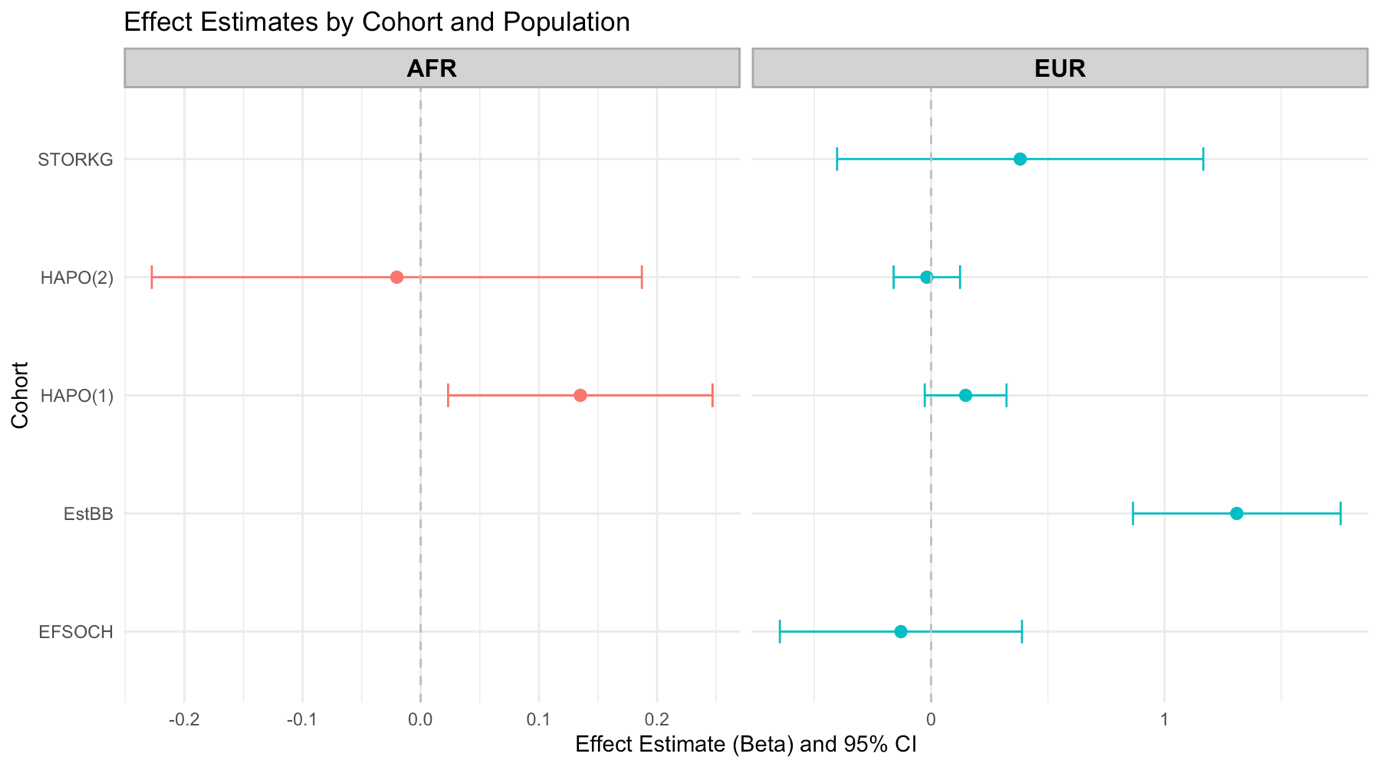

**Supplementary Figure 55.** *Comparison of the effect estimates across populations and cohorts for the variant rs368956520 detected in the multi-ancestry meta-analysis of HbA1c.* Effects are shown with respect to the A allele. In cases where a cohort contributed multiple files for the same population, bracketed numbers were used to distinguish them for reference purposes. Effect allele frequencies (EAF) for each study were as follows: STORKG=0.99, EstBB=0.99, EFSOCH=0.99, HAPO(1) AFR=0.97, HAPO(2) AFR=0.97, HAPO(1) EUR=0.99 and HAPO(2) EUR=0.99. AFR; African. EAS; East Asian. EUR; European. SAS; South Asian. HIS; Hispanic.

#### 3.3 GDM diagnostic test sensitivity analysis

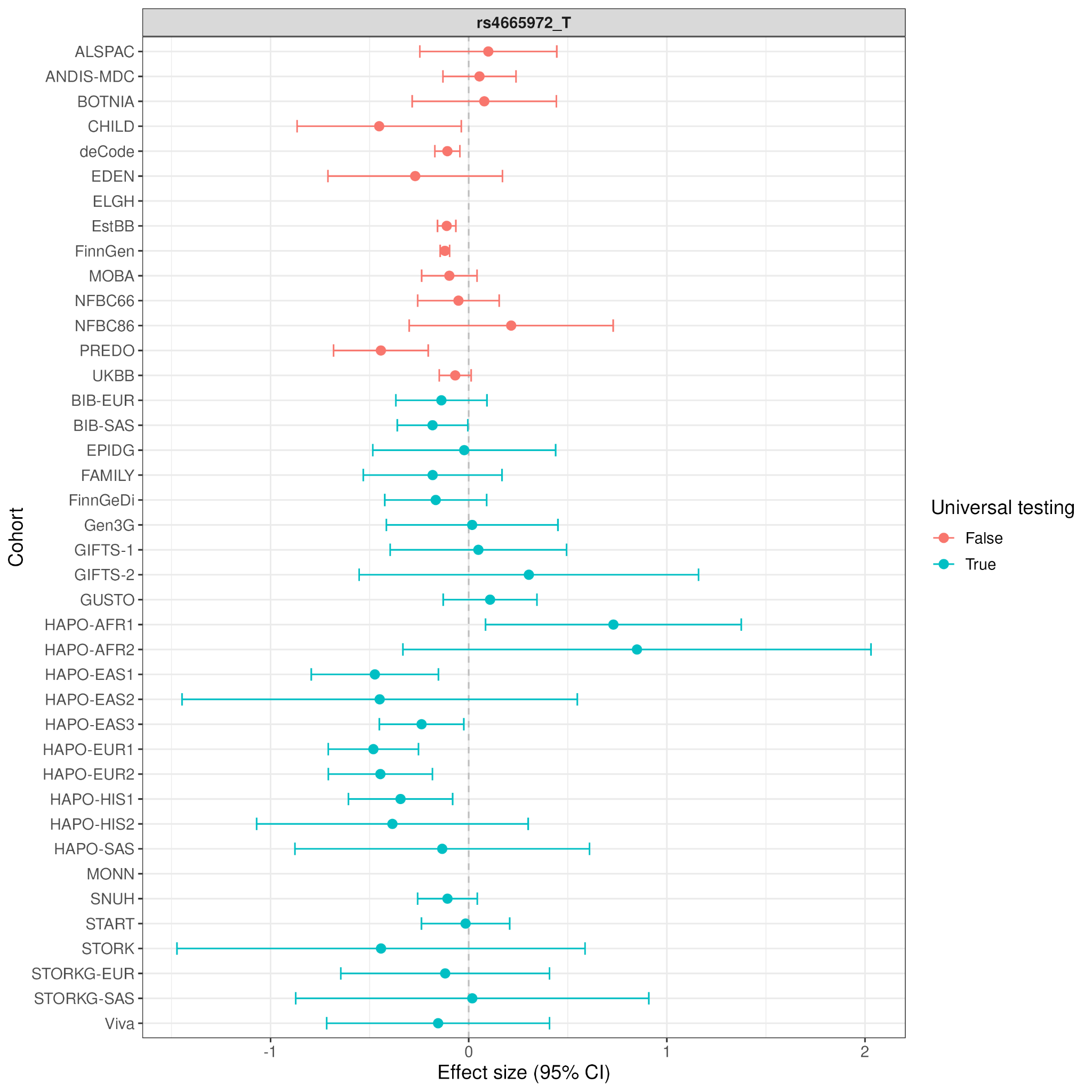

**Supplementary Figure 56.** *Comparison of the effect estimates for maternal multi-ancestry GDM lead variant rs4665972 at the GCKR-SNX17 locus exhibiting evidence of phenotype-correlated heterogeneity.* Effect estimates and corresponding 95% confidence intervals (CI) are obtained from cohort-level summary statistics for maternal GDM provided by 28 contributing cohorts. In case where a cohort contributed multiple files for the same population, numbers were used to distinguish them for reference purposes. Effect estimates (only plotted if present) are shown with respect to the allele indicated next to the marker name. GDM; gestational diabetes mellitus. AFR; African. EAS; East Asian. EUR; European. SAS; South Asian. HIS; Hispanic.

### **4.0 Gene-based test**

#### 4.1 European analyses

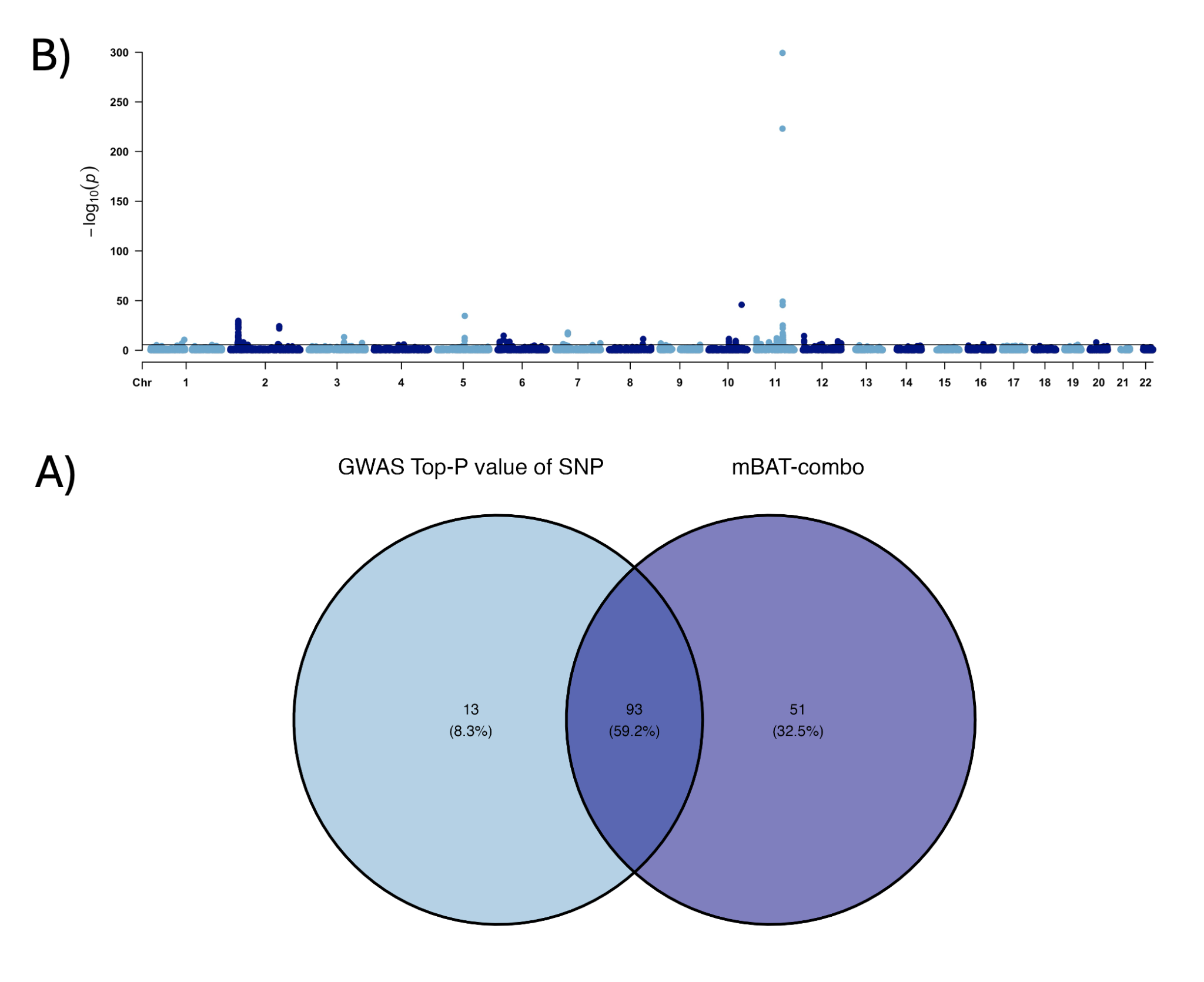

**Supplementary Figure 57.** *Gene-based test results for the maternal gestational diabetes mellitus (GDM) analysis* (A) Manhattan plot displaying gene-based association results using mBAT-combo. The y-axis represents the −log10-transformed p-values, and the x-axis shows the genomic positions of genes across chromosomes. The solid black line indicates the genome-wide significance threshold (P= 2.66$\times$10^-6^; P=0.05/number of genes). **(B)** Venn diagram illustrating the overlap between genes reaching genome-wide significance identified by mBAT-combo (P= 2.66$\times$10^-6^) and those identified using the P-value for the top SNP within the gene (P=0.05/number of SNPs). The analysis reveals that 59.2% of significant genes are detected by both methods, indicating substantial concordance. However, mBAT-combo identifies an additional 51 genes not detected by the GWAS top SNP P-value approach.

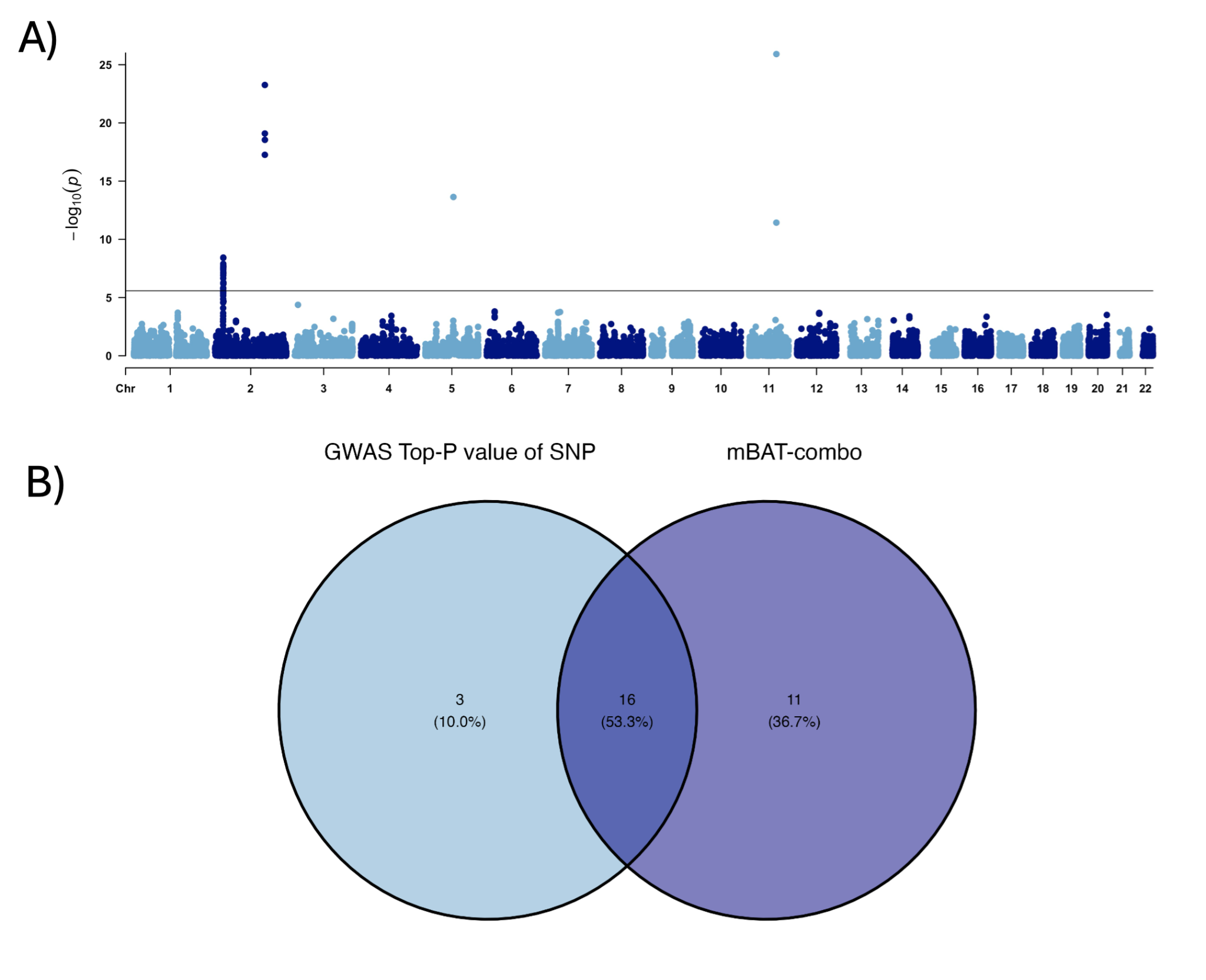

**Supplementary Figure 58.** *Gene-based test results for the maternal fasting glucose analysis.* (A) Manhattan plot displaying gene-based association results using mBAT-combo. The y-axis represents the −log10-transformed p-values, and the x-axis shows the genomic positions of genes across chromosomes. The solid black line indicates the genome-wide significance threshold (P= 2.66$\times$10^-6^; P=0.05/number of genes). **(B)** Venn diagram illustrating the overlap between genes reaching genome-wide significance identified by mBAT-combo (P=2.66$\times$10^-6^) and those identified using the P-value for the top SNP within the gene (P=0.05/number of SNPs). The analysis reveals that 53.3% of significant genes are detected by both methods, indicating substantial concordance. However, mBAT-combo identifies eleven additional genes not detected by the GWAS top SNP P-value approach.

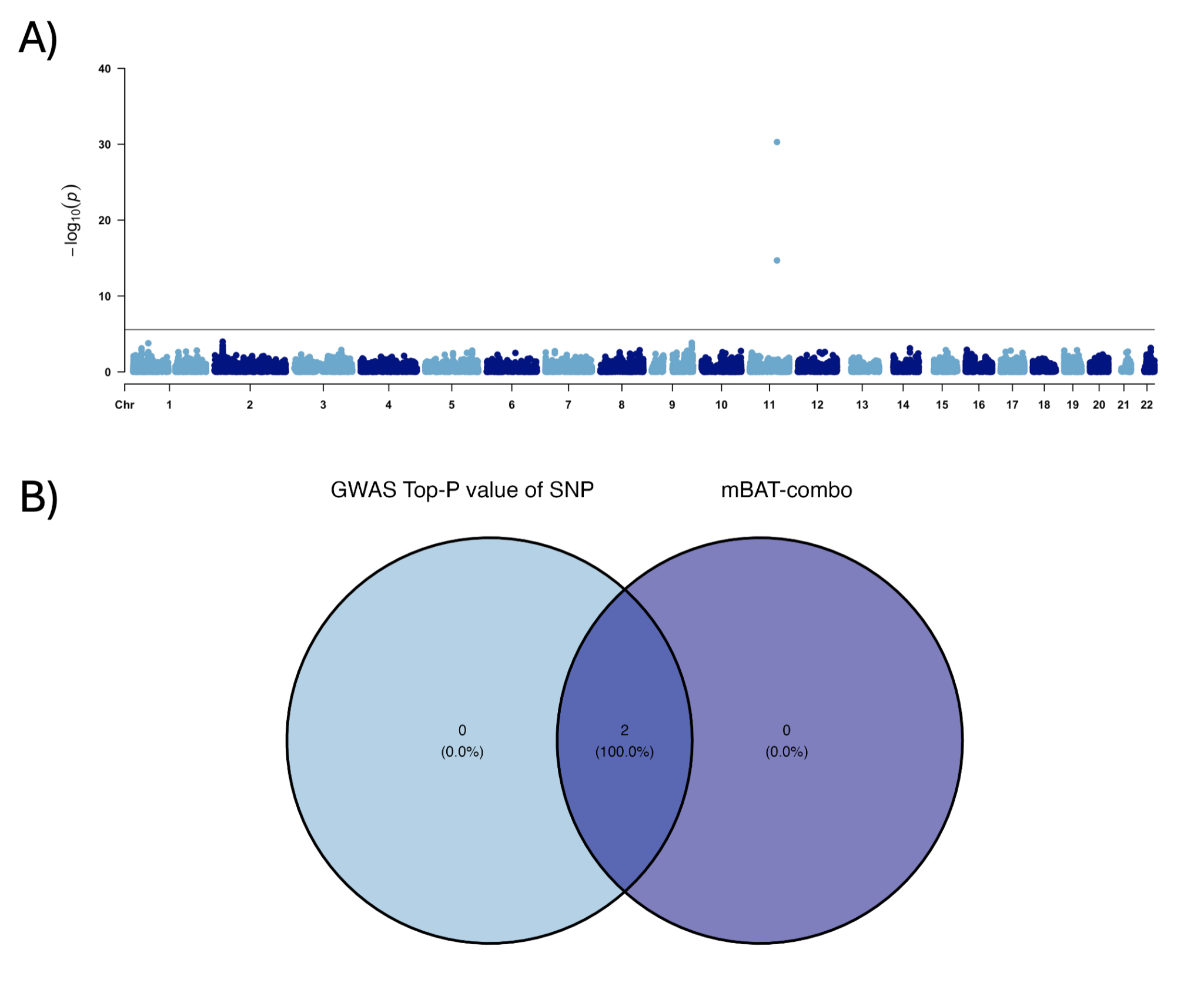

**Supplementary Figure 59.** *Gene-based test results for the maternal 1-hour glucose post-OGTT analysis.* (A) Manhattan plot displaying gene-based association results using mBAT-combo. The y-axis represents the −log10-transformed p-values, and the x-axis shows the genomic positions of genes across chromosomes. The solid black line indicates the genome-wide significance threshold (P= 2.66$\times$10^-6^; P=0.05/number of genes). **(B)** Venn diagram illustrating the overlap between genes reaching genome-wide significance identified by mBAT-combo (P=2.66$\times$10^-6^) and those identified using the P-value for the top SNP within the gene (P=0.05/number of SNPs). The analysis shows that significant genes are detected by both methods, with the gene-based test not providing an advantage over individual SNP testing.

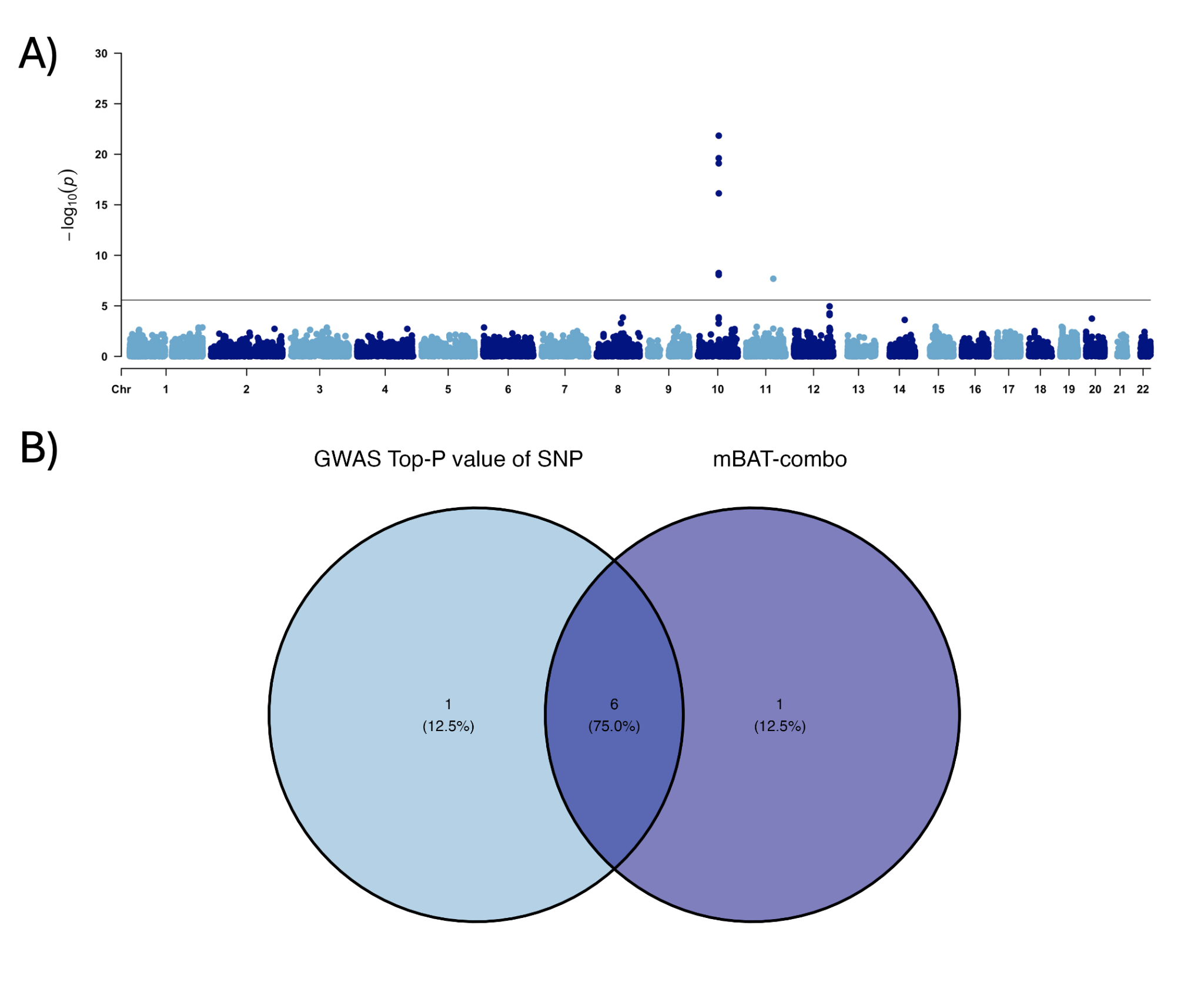

**Supplementary Figure 60.** *Gene-based test results for the maternal 2-hour glucose post-OGTT values analysis* (A) Manhattan plot displaying gene-based association results using mBAT-combo. The y-axis represents the −log10-transformed p-values, and the x-axis shows the genomic positions of genes across chromosomes. The solid black line indicates the genome-wide significance threshold (P= 2.66$\times$10^-6^; P=0.05/number of genes). **(B)** Venn diagram illustrating the overlap between genes reaching genome-wide significance identified by mBAT-combo (P=2.66$\times$10^-6^) and those identified using the P-value for the top SNP within the gene (P=0.05/number of SNPs). The analysis shows that most (75%) significant genes are detected by both methods, with only one additional gene being identified using mBAT-combo.

#### 4.2 East Asian analyses

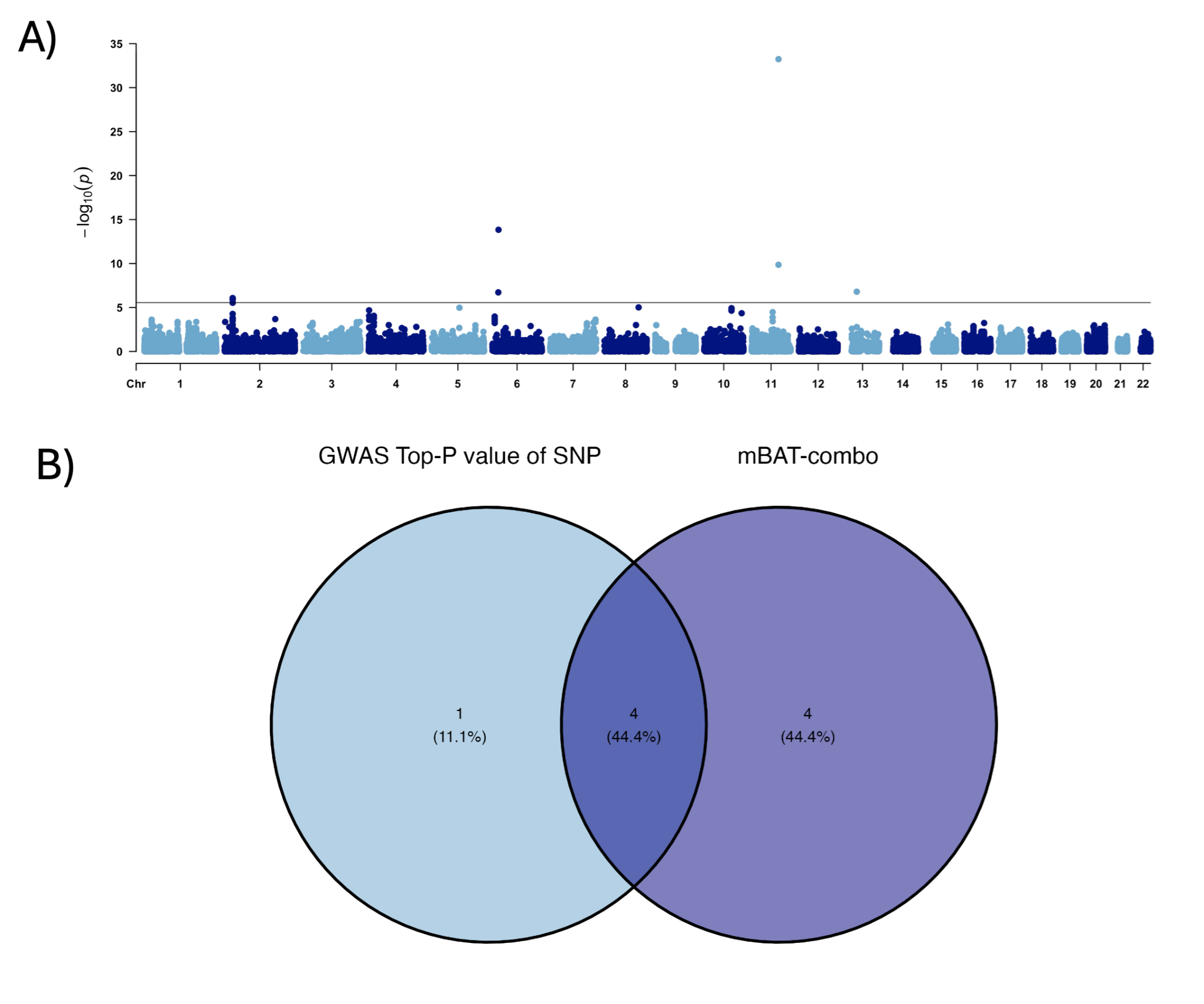

**Supplementary Figure 61.** *Gene-based test results for the maternal gestational diabetes mellitus (GDM) analysis* (A) Manhattan plot displaying gene-based association results using mBAT-combo. The y-axis represents the −log10-transformed p-values, and the x-axis shows the genomic positions of genes across chromosomes. The solid black line indicates the genome-wide significance threshold (P= 2.66$\times$10^-6^; P=0.05/number of genes). **(B)** Venn diagram illustrating the overlap between genes reaching genome-wide significance identified by mBAT-combo (P=2.75$\times$10^-6^) and those identified using the P-value for the top SNP within the gene (P=0.05/number of SNPs). The analysis reveals that 44.4% of significant genes are detected by both methods. However, mBAT-combo identifies four additional genes not detected by the GWAS top SNP P-value approach.

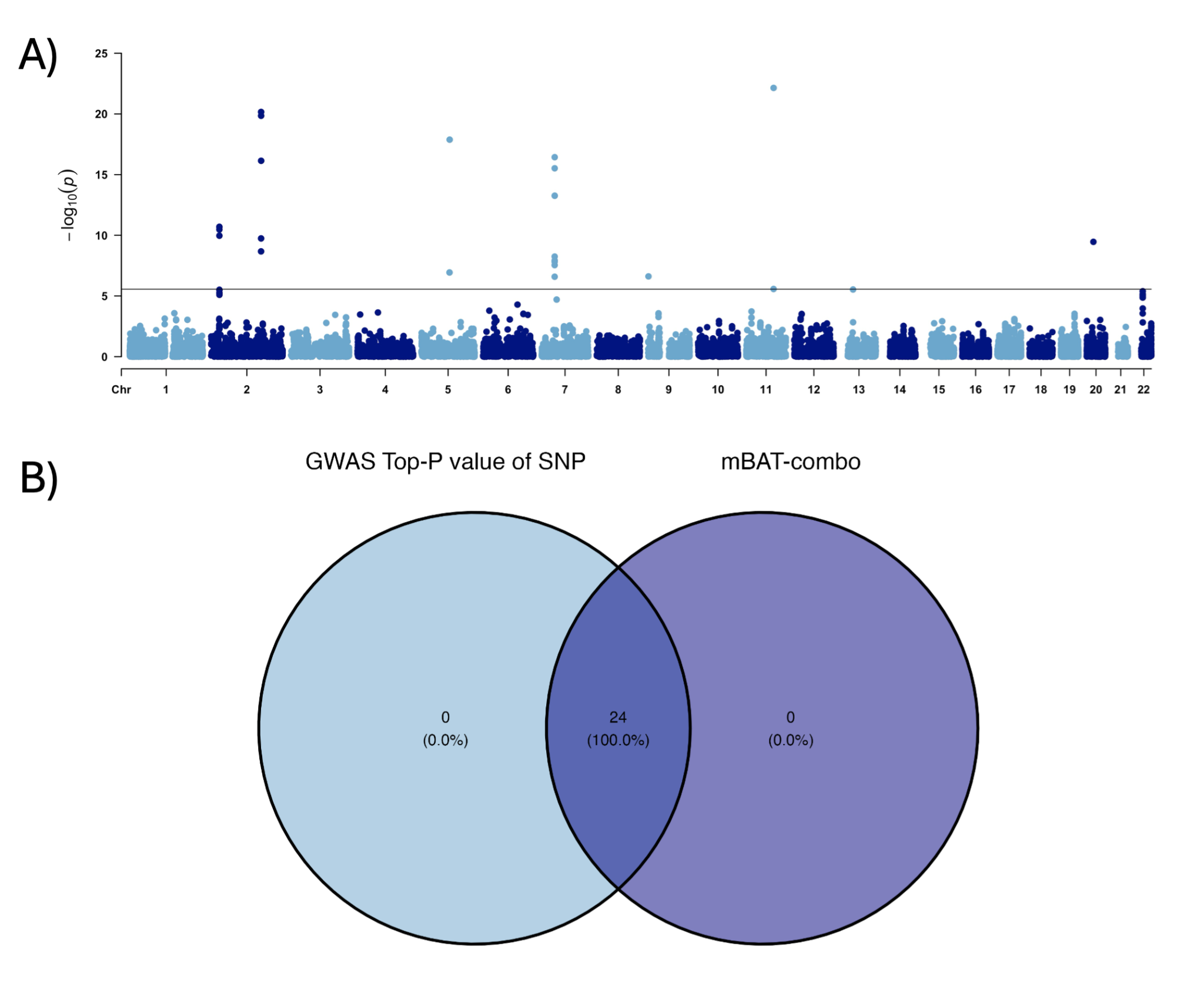

**Supplementary Figure 62.** *Gene-based test results for the maternal fasting glucose analysis.* (A) Manhattan plot displaying gene-based association results using mBAT-combo. The y-axis represents the −log10-transformed p-values, and the x-axis shows the genomic positions of genes across chromosomes. The solid black line indicates the genome-wide significance threshold (P= 2.66$\times$10^-6^; P=0.05/number of genes). **(B)** Venn diagram illustrating the overlap between genes reaching genome-wide significance identified by mBAT-combo (P=2.75$\times$10^-6^) and those identified using the P-value for the top SNP within the gene (P=0.05/number of SNPs). The analysis shows that significant genes are detected by both methods, with the gene-based test not providing an advantage over individual SNP testing.

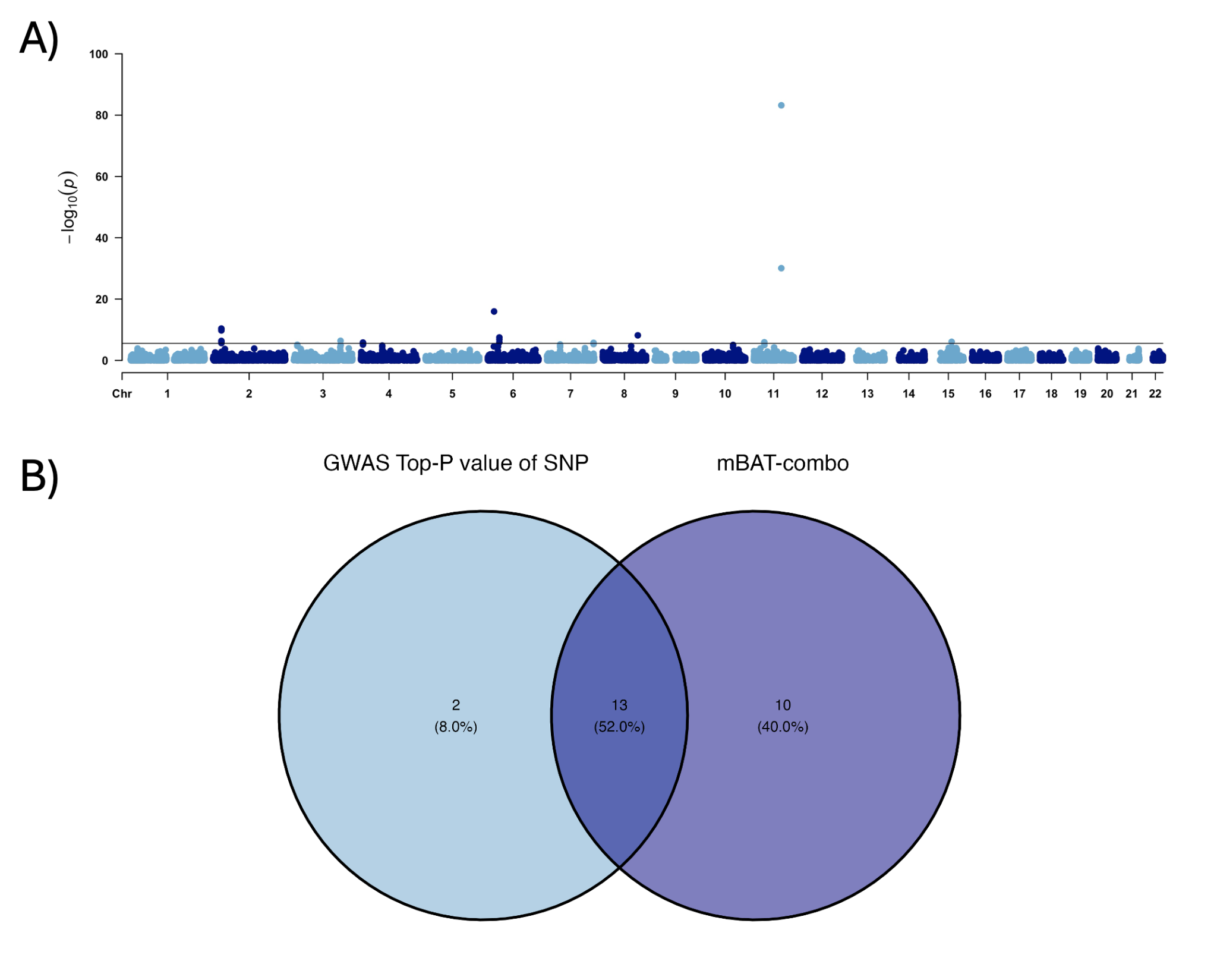

**Supplementary Figure 63.** *Gene-based test results for the maternal 1-hour glucose post-OGTT values analysis.* (A) Manhattan plot displaying gene-based association results using mBAT-combo. The y-axis represents the −log10-transformed p-values, and the x-axis shows the genomic positions of genes across chromosomes. The solid black line indicates the genome-wide significance threshold (P= 2.66$\times$10^-6^; P=0.05/number of genes). **(B)** Venn diagram illustrating the overlap between genes reaching genome-wide significance identified by mBAT-combo (P=2.75$\times$10^-6^) and those identified using the P-value for the top SNP within the gene (P=0.05/number of SNPs). The analysis reveals that 52% of significant genes are detected by both methods. However, mBAT-combo identifies ten additional genes not detected by the GWAS top SNP P-value approach.

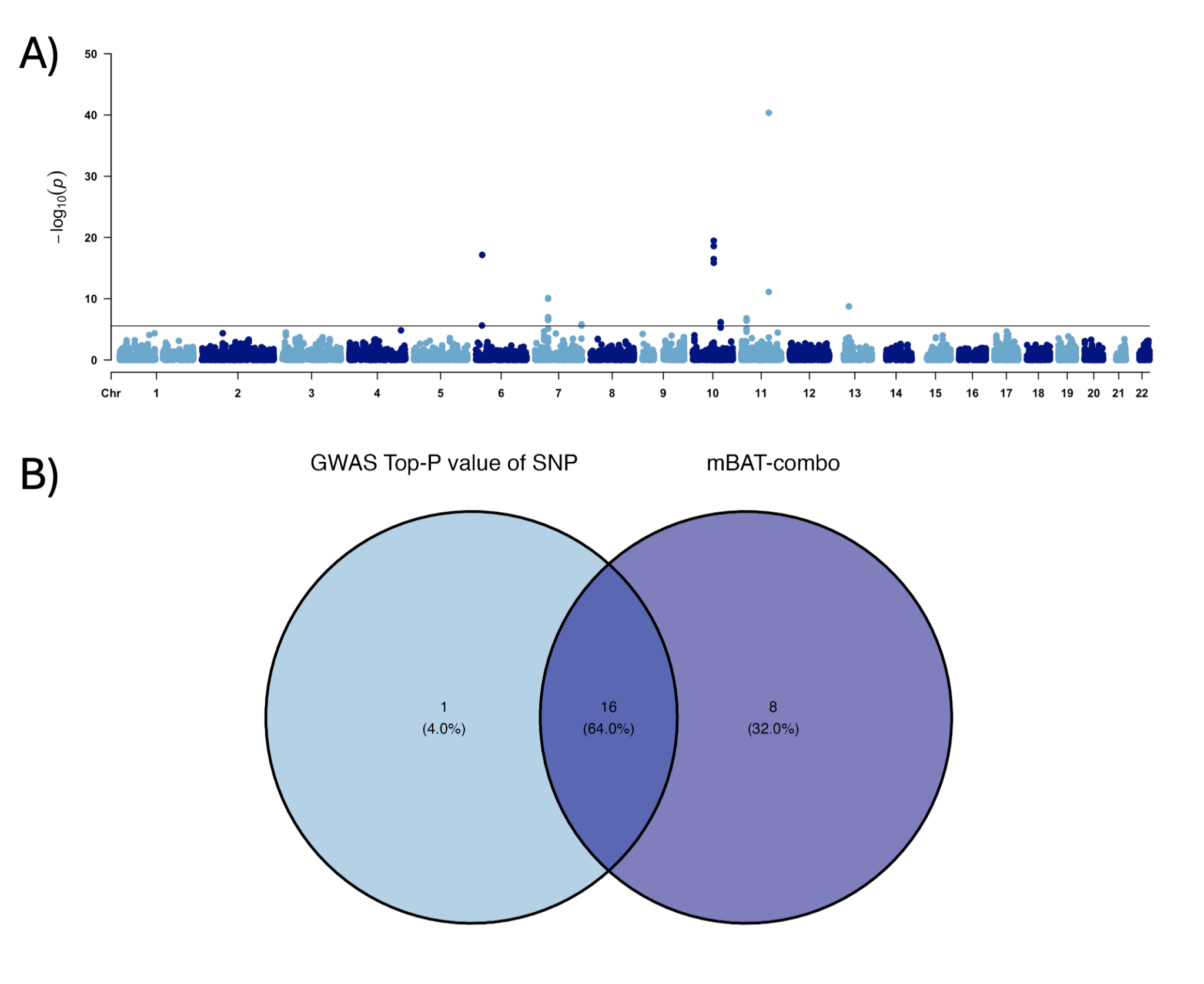

**Supplementary Figure 64.** *Gene-based test results for the maternal 2-hour glucose post-OGTT values analysis.* (A) Manhattan plot displaying gene-based association results using mBAT-combo. The y-axis represents the −log10-transformed p-values, and the x-axis shows the genomic positions of genes across chromosomes. The solid black line indicates the genome-wide significance threshold (P= 2.66$\times$10^-6^; P=0.05/number of genes). **(B)** Venn diagram illustrating the overlap between genes reaching genome-wide significance identified by mBAT-combo (P=2.75$\times$10^-6^) and those identified using the P-value for the top SNP within the gene (P=0.05/number of SNPs). The analysis reveals that 64% of significant genes are detected by both methods. However, mBAT-combo identifies eight additional genes not detected by the GWAS top SNP P-value approach.

#### 4.3 Genetic overlap between phenotypes

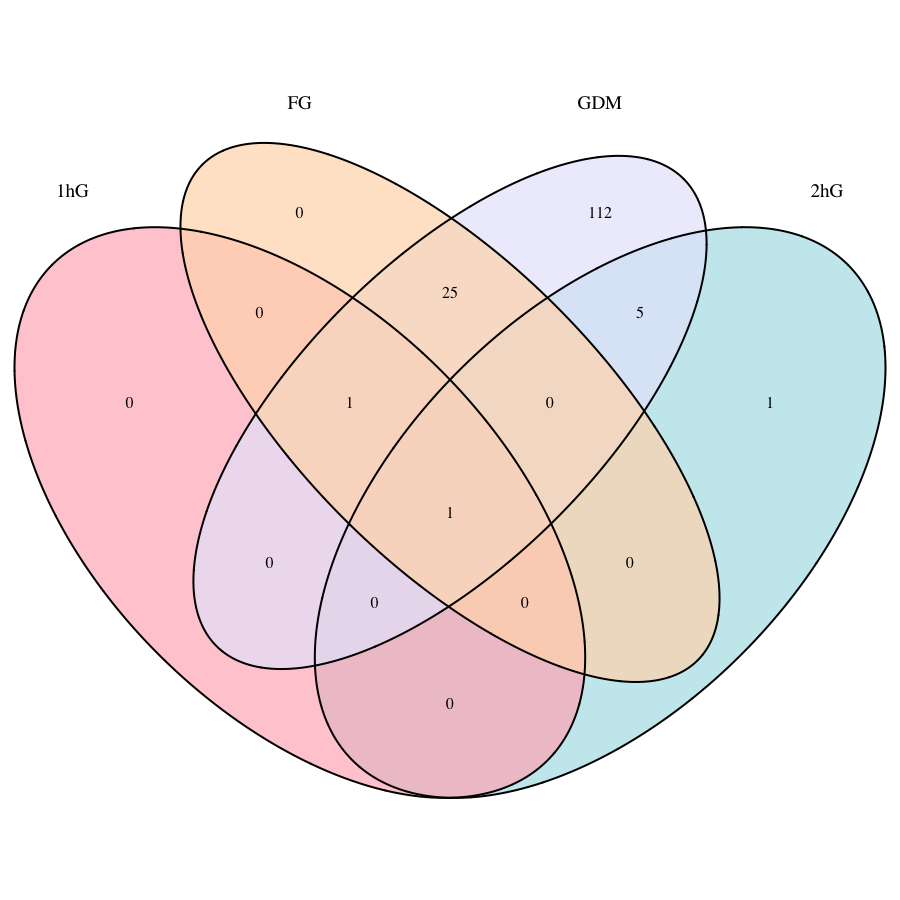

**Supplementary Figure 65.** *Venn diagram illustrating the genetic overlap between the phenotypes in the gene-based test analyses in Europeans.* Genes included in the diagram reached genome-wide significance at the gene-level in the mBAT-combo analyses (P=0.05/number of genes, P=2.66×10^-6^). *MTNR1B* was consistently identified across all phenotypes, while *FAT3* was shared among 1hG, FG, and GDM but absent in 2hG. Five genes (*SRGN, VPS26A, SUPV3L1, HKDC1,* and *HK1*) were exclusive to GDM and 2hG, whereas 25 genes were associated with GDM and FG, but not 1hG or 2hG (these include *ABCB11, GCKR, SLC30A3, MPV17, GTF3C2, EIF2B4, NRBP1, FNDC4, SNX17-GCKR, PPM1G, SUPT7L, IFT172, SPC25, G6PC2, KRTCAP3, NOSTRIN, DNAJC5G, SLC4A1AP, RBKS, CCDC121, GPN1, C2orf16, MRPL33, ZNF512,* and *CAST-PCSK1).* Additionally, *KIFBP* was identified only in 2hG, while 112 genes were exclusive to GDM. GDM; Gestational Diabetes Mellitus. FG; Fasting glucose. 1hG; 1-hour glucose after oral-glucose tolerance test (OGTT). 2hG; 2-hour glucose after oral-glucose tolerance test (OGTT).

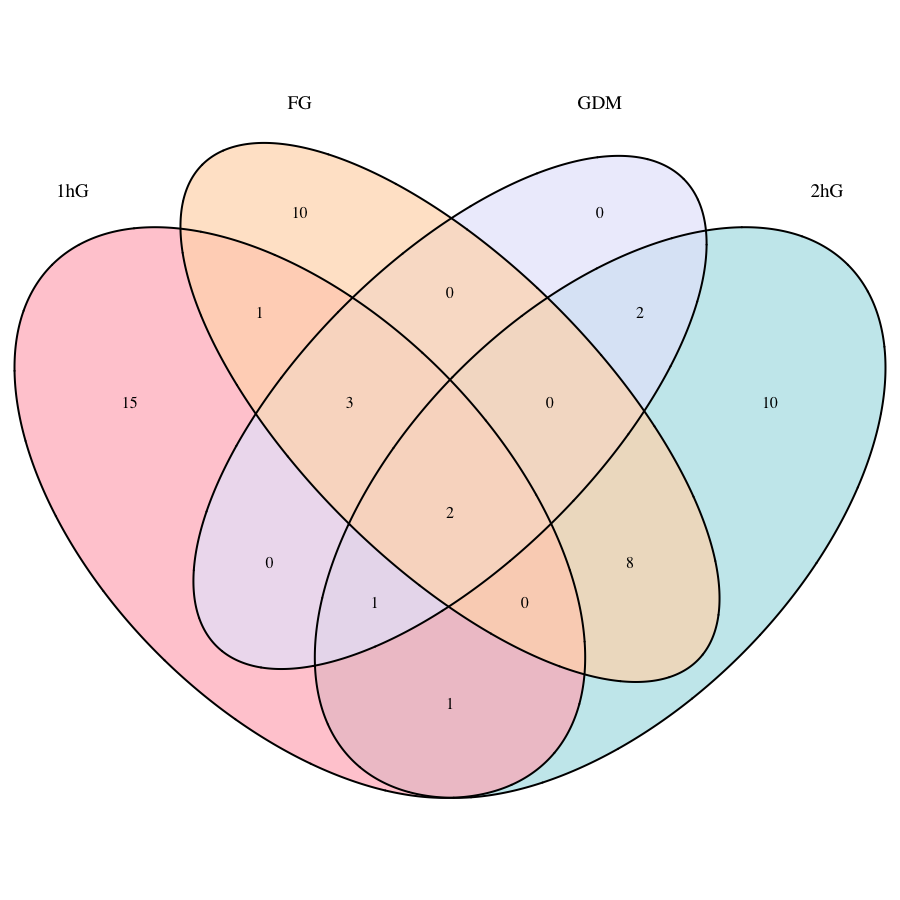

**Supplementary Figure 66.** *Venn diagram illustrating the genetic overlap between the phenotypes in the gene-based test analyses in East Asians.* Genes included in the diagram reached genome-wide significance at the gene-level in the mBAT-combo analyses (P=0.05/number of genes, P= 2.75×10^-6^). *MTNR1B* and *FAT3* were consistently identified across all phenotypes. *GCKR, FNDC4*, and *IFT172* were shared among 1hG, FG, and GDM, but absent in 2hG, while *CDKAL1* was found in 1hG, 2hG, and GDM, but not FG. *C2orf16* was exclusive to 1hG and FG, *MNX1* to 1hG and 2hG, and eight genes (*AEBP1, POLD2, MYL7, GCK, YKT6, POLM, DBNL*, and *PGAM2*) to FG and 2hG. Two genes (*EXOSC8* and *E2F3*) were identified in GDM and 2hG, but not in other analyses. Additionally, 10 genes were exclusive to 2hG, 15 to 1hG, and 10 to FG. GDM; Gestational Diabetes Mellitus. FG; Fasting glucose. 1hG; 1-hour glucose after oral-glucose tolerance test (OGTT). 2hG; 2-hour glucose after oral-glucose tolerance test (OGTT).

### **5.0 Genetic correlation analyses**

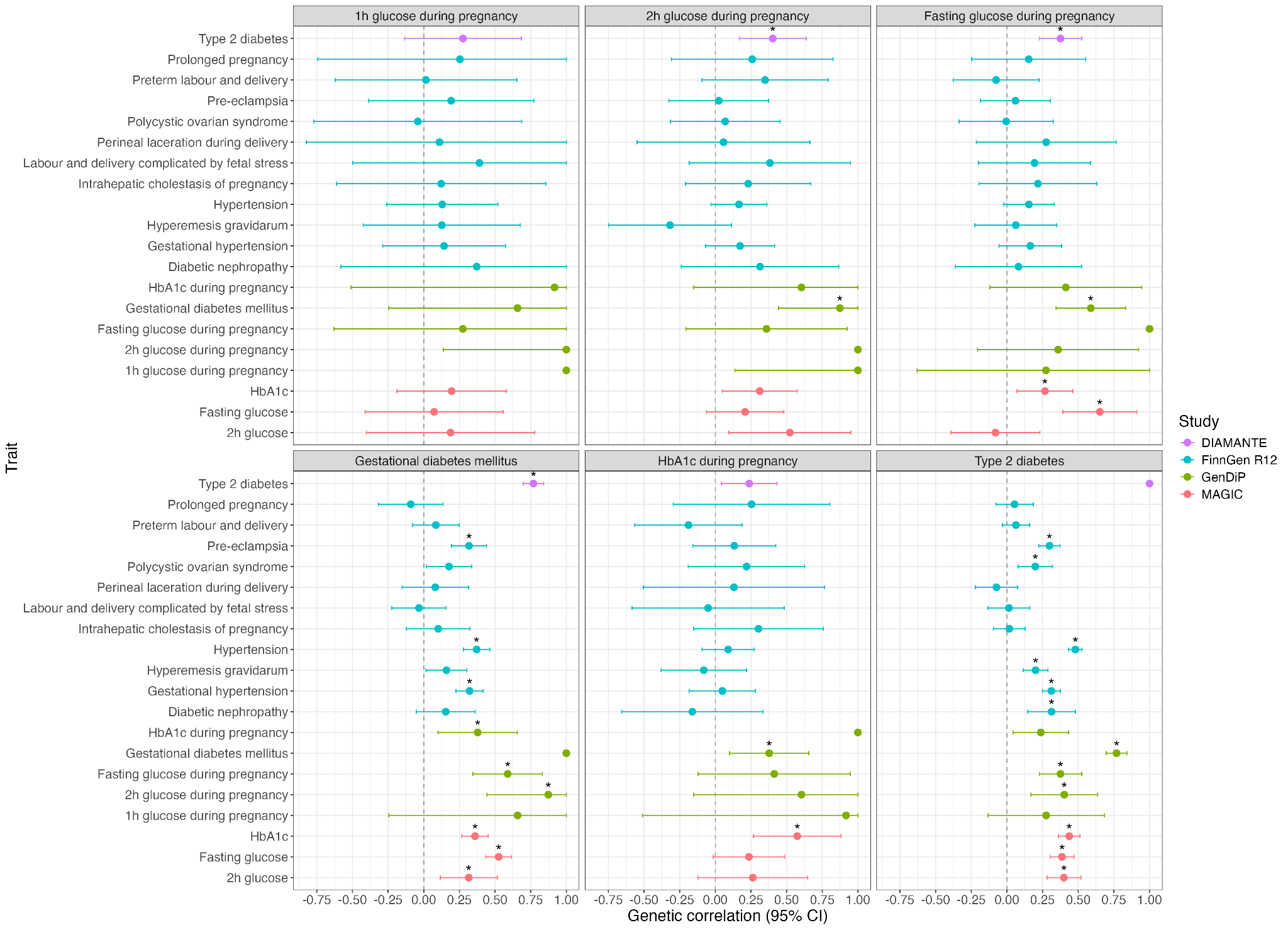

**Supplementary Figure 67***. Pairwise genetic correlation analyses in Europeans.* Panels display trait-specific correlations for diabetes and glycemic traits during pregnancy (1-hour glucose, 2-hour glucose, fasting glucose, GDM and HbA1c) as well as type 2 diabetes (T2DM). Each panel shows correlations with: (i) T2DM (purple; DIAMANTE consortium, PMID: 35551307), (ii) women's reproductive health outcomes and hypertension (blue; FinnGen R12, PMID: 36653562), (iii) pregnancy-specific glycemic traits (green; from our European GWAS meta-analyses), and (iv) glycemic trait measures outside of pregnancy (red; MAGIC consortium, PMID: 34059833). Genetic correlation estimates (rG) are presented as the dots on the forest plots, and error bars indicate 95% confidence intervals (CI). Asterisks indicate statistically significant associations after correction for false discovery rate (FDR < 0.05). The vertical dashed line at rG = 0 represents no genetic correlation. 95% CI were truncated in case they were outside -1 to 1 range.

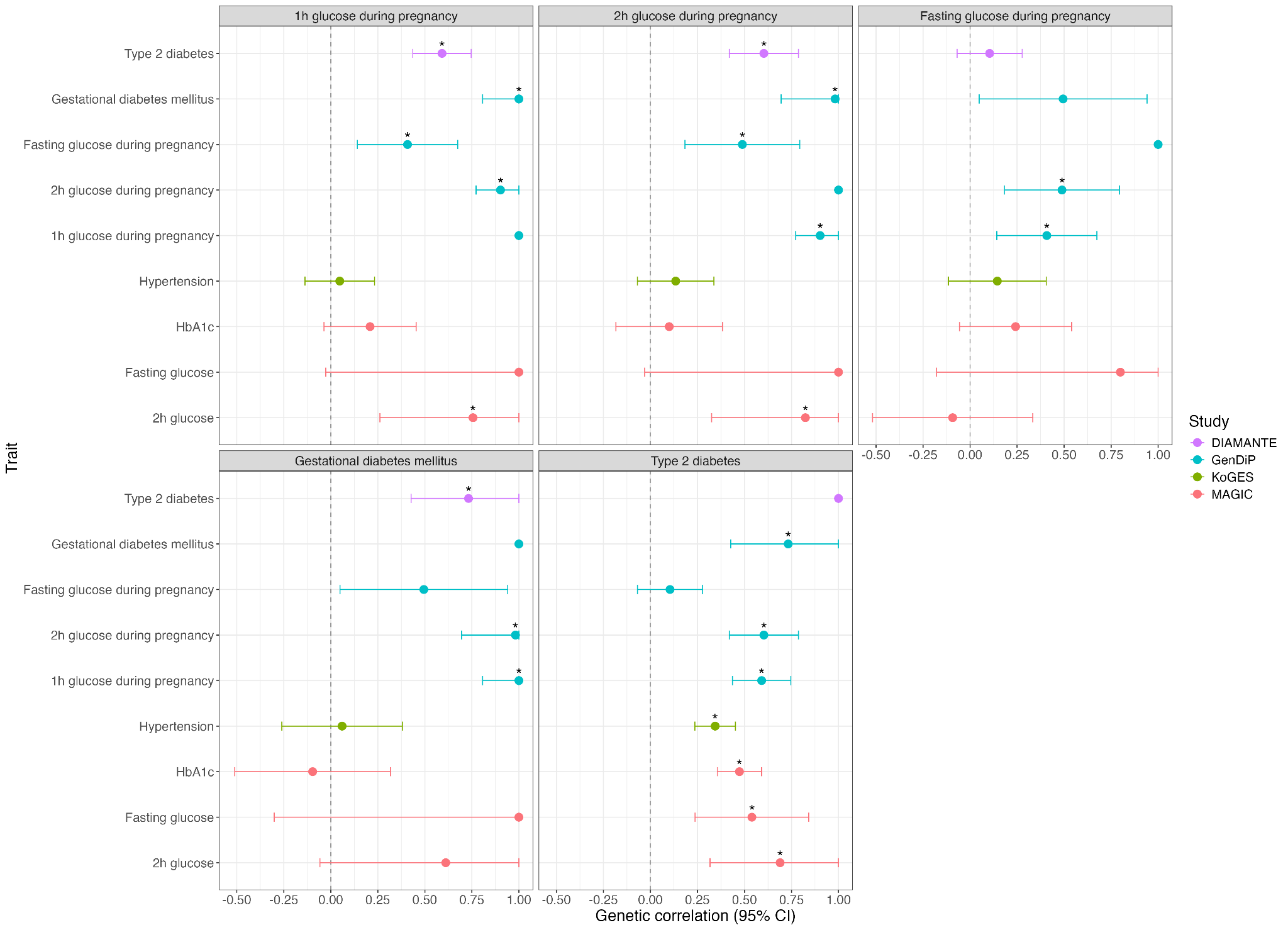

**Supplementary Figure 68***. Pairwise genetic correlation analyses in East Asians.* Panels display trait-specific correlations for diabetes and glycemic traits during pregnancy (1-hour glucose, 2-hour glucose, fasting glucose, and GDM) as well as type 2 diabetes (T2DM). Each panel shows correlations with: (i) T2DM (purple; DIAMANTE consortium, PMID: 35551307), (ii) pregnancy-specific glycemic traits (blue; from our East Asian GWAS meta-analyses), (iii) hypertension (green; KoGES consortium, PMID: 36777999), and (iv) glycemic trait measures outside of pregnancy (red; MAGIC consortium, PMID: 34059833). Genetic correlation estimates (rG) are presented as dots on the forest plots, and error bars indicate 95% confidence intervals (CI). Asterisks indicate statistically significant associations after correction for false discovery rate (FDR < 0.05). The vertical dashed line at rG = 0 represents no genetic correlation. 95% CI were truncated in case they were outside -1 to 1 range.

### **6.0 SCOUTJOY analyses**

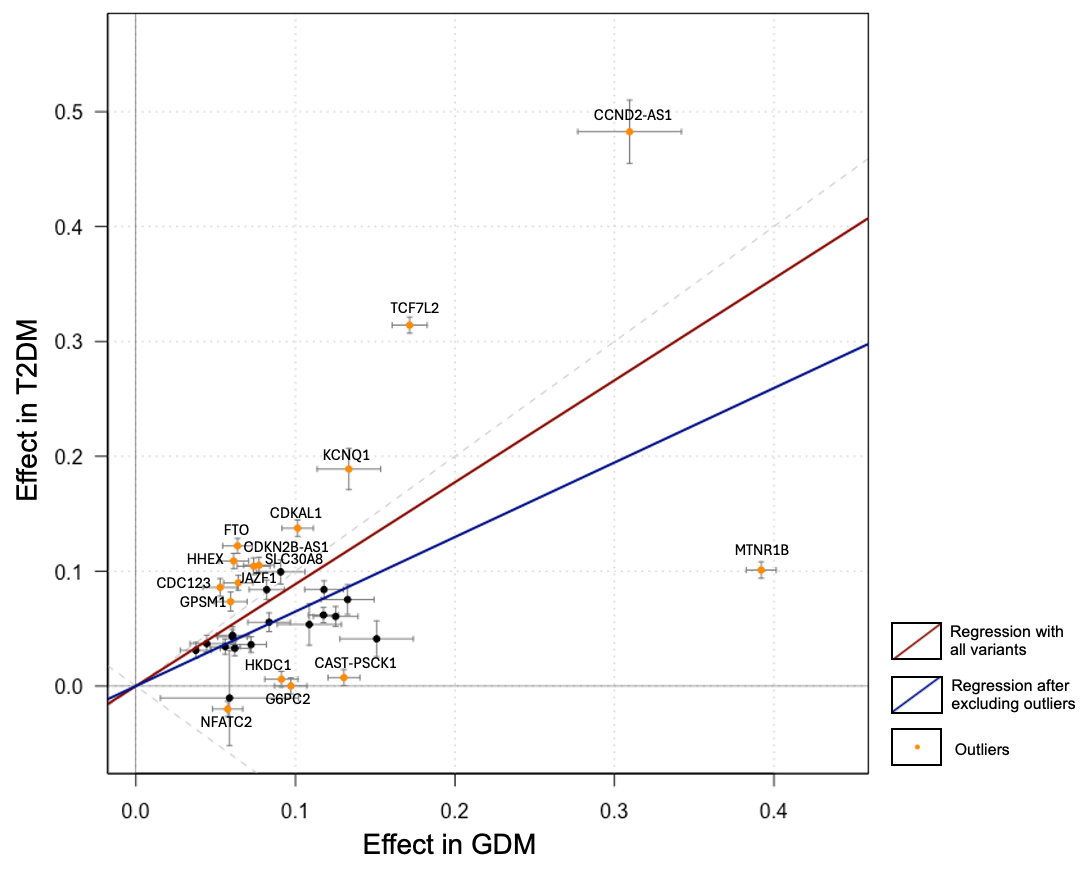

**Supplementary Figure 69***. Comparison of genetic effect estimates for lead SNPs associated with gestational diabetes mellitus (GDM) and their corresponding effects on type 2 diabetes mellitus (T2DM; sex-combined) in Europeans.* Genetic variants analysed were identified in the trans-ancestry GWAS meta-analysis of GDM (38,241 cases and 775,169 controls), with effect estimates and standard errors obtained from the European GDM GWAS meta-analysis (29,625 cases, 736,101 controls). European T2DM GWAS was obtained from Mahajan et al., 2022 (PMID: 35551307), and included 80,154 cases and 853,816 controls (including males and females, unadjusted for BMI). Effect estimates are presented as betas (log odds ratio). Error bars indicate +/- 1 standard error in each GWAS. Orange points labelled with putative names are identified as outliers by SCOUTJOY. Fitted slopes are shown from York regression with all variants (red) and after the removal of identified outliers (blue). Dashed gray reference lines indicate equal absolute effect size. SCOUTJOY analysis suggests a significant positive relationship in genetic effects before and after outlier removal. Analyses also suggest significant heterogeneity, identifying sixteen outliers. Outliers identified included: *CCND2-AS1* (lead variant: rs76895963), *TCF7L2* (rs7903146), *KCNQ1* (rs2299620), *CDKAL1* (rs9348441), *FTO* (rs55872725), *CDKN2B-AS1* (rs7018475), *SLC30A8* (rs11558471), *JAZF1* (rs849134), *HHEX* (rs11187141), *CDC123* (rs11257655), *GPSM1* (rs28642213), *HKDC1* (rs10762264), *NFATC2* (rs6021276), *G6PC2* (rs560887), *CAST-PSCK1* (rs10036439), *MTNR1B* (rs10830963). The following SNPs were not present in the T2DM dataset: rs537224022 (*ESR1*), rs142406759 (*RNF144B*), rs113851927 (*IGF2BP2*) and rs5831573 (*BCL11A*). T2DM; Type-II Diabetes Mellitus. GDM; Gestational Diabetes Mellitus.

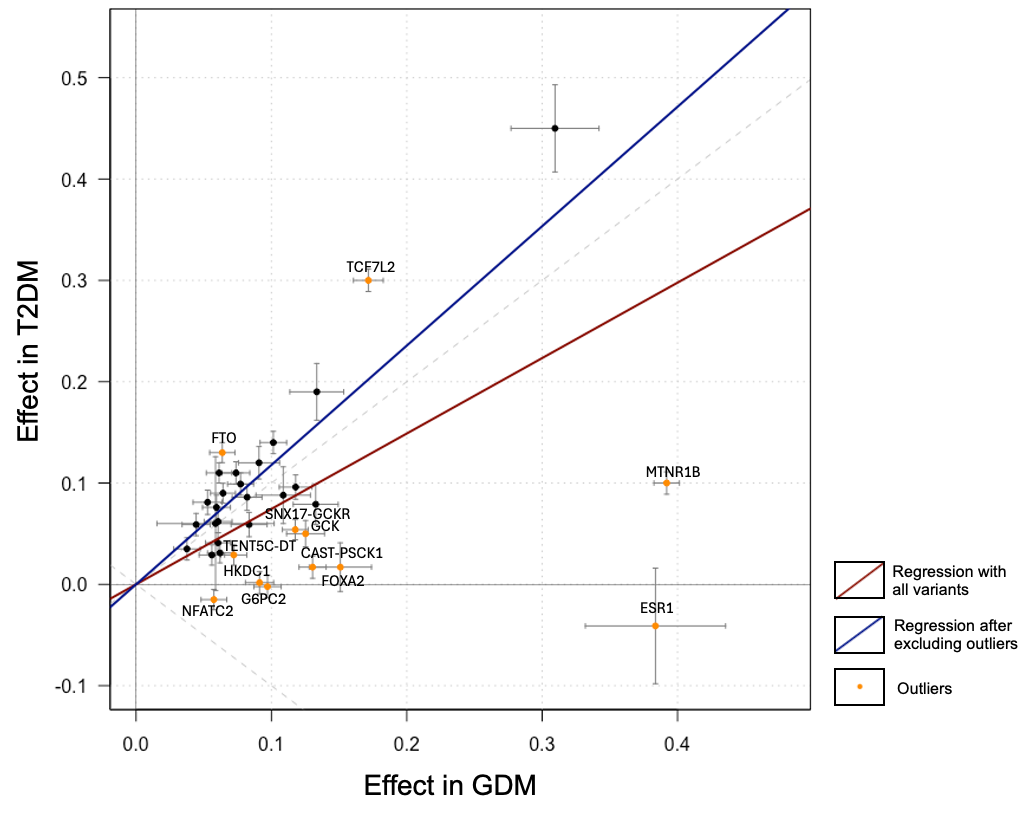

**Supplementary Figure 70*.*** *Comparison of genetic effect estimates for lead SNPs associated with gestational diabetes mellitus (GDM) and their corresponding effects on type 2 diabetes mellitus (T2DM; females only) in Europeans.* Genetic variants analysed were identified in the trans-ancestry GWAS meta-analysis of GDM (38,241 cases and 775,169 controls), with effect estimates and standard errors obtained from the European GDM GWAS meta-analysis (29,625 cases, 736,101 controls). European T2DM GWAS was obtained from Mahajan et al., 2018 (PMID: 30297969), and included 30,053 cases and 434,336 controls (females only, unadjusted for BMI). Effect estimates are presented as betas (log odds ratio). Error bars indicate +/- 1 standard error in each GWAS. Orange points labelled with putative gene names are identified as outliers by SCOUTJOY. Fitted slopes are shown from York regression with all variants (red) and after the removal of identified outliers (blue). Dashed gray reference lines indicate equal absolute effect size. SCOUTJOY analysis suggests a significant positive relationship in genetic effects before and after outlier removal. Analyses also suggest significant heterogeneity, identifying twelve outliers. Outliers identified included: *TCF7L2* (rs7903146), *FTO* (rs55872725), *SNX17-GCKR* (rs4665972), *GCK* (rs6975024), *CAST-PSCK1* (rs10036439), *FOXA2* (rs11087387), *TENT5C-DT* (rs1975283), *HKDC1* (rs10762264), *G6PC2* (rs560887), *NFATC2* (rs6021276), *ESR1* (rs537224022), and *MTNR1B* (rs10830963). The following SNPs were not present in the T2DM dataset: rs142406759 (*RNF144B*), rs113851927 (*IGF2BP2*) and rs5831573 (*BCL11A*). T2DM; Type-II Diabetes Mellitus. GDM; Gestational Diabetes Mellitus.

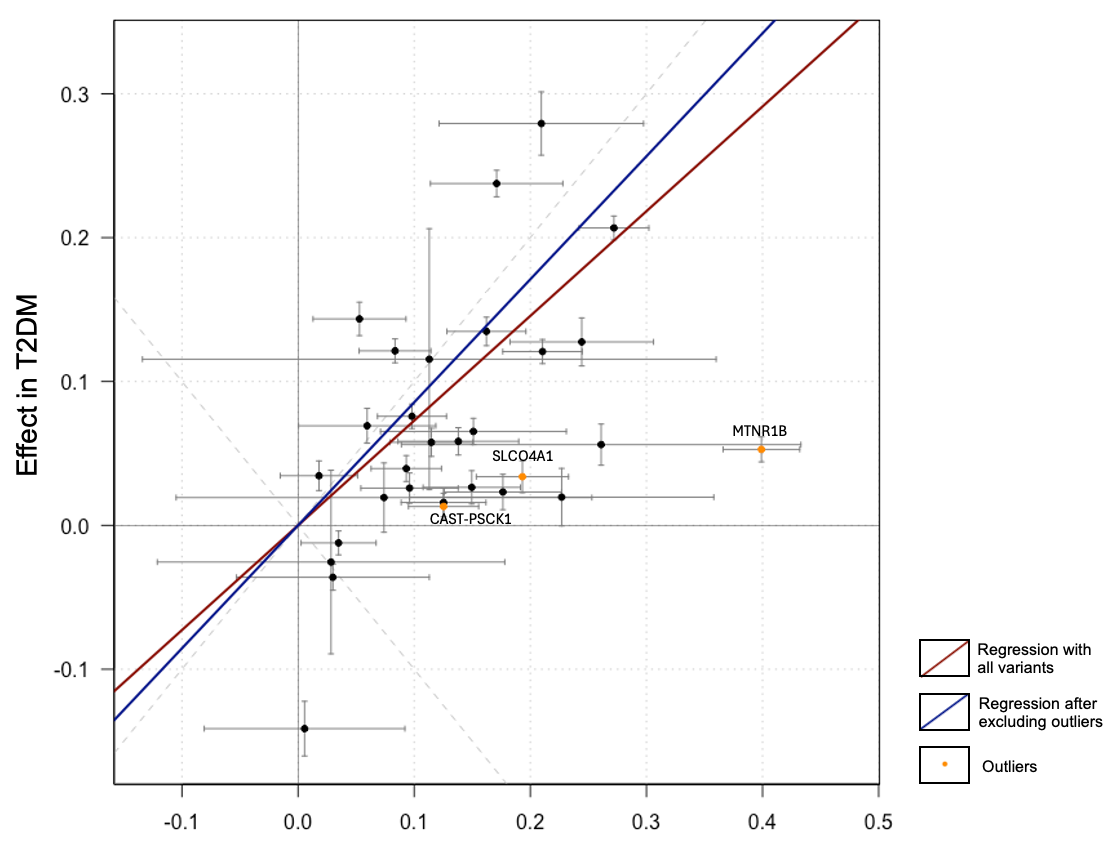

**Supplementary Figure 71*.*** *Comparison of genetic effect estimates for lead SNPs associated with gestational diabetes mellitus (GDM) and their corresponding effects on type 2 diabetes mellitus (T2DM; sex-combined) in East Asians.* Genetic variants analysed were identified in the trans-ancestry GWAS meta-analysis of GDM (38,241 cases and 775,169 controls), with effect estimates and standard errors obtained from the East Asian GDM GWAS meta-analysis (4,346 cases, 23,286 controls). East Asian T2DM GWAS was obtained from Mahajan et al., 2018 (PMID: 30297969), and included 56,268 cases and 227,155 controls (males and females, unadjusted for BMI). Effect estimates are presented as betas (log odds ratio). Error bars indicate +/- 1 standard error in each GWAS. Orange points labelled with putative gene names are identified as outliers by SCOUTJOY. Fitted slopes are shown from York regression with all variants (red) and after the removal of identified outliers (blue). Dashed gray reference lines indicate equal absolute effect size. SCOUTJOY analysis suggests a significant positive relationship in genetic effects before and after outlier removal. Analyses also suggest significant heterogeneity, identifying three outliers. Outliers detected include: *CAST-PSCK1* (rs10036439), *SLCO4A1* (rs6122141), and *MTNR1B* (rs10830963). The following SNPs were not present in the GDM or T2DM dataset: s41279738 (*GPR61*), rs142406759 (*RNF144B*), rs537224022 (*ESR1*), rs76895963 (*CCND2*), and rs77090130 (*PRAME*). T2DM; Type-II Diabetes Mellitus. GDM; Gestational Diabetes Mellitus.

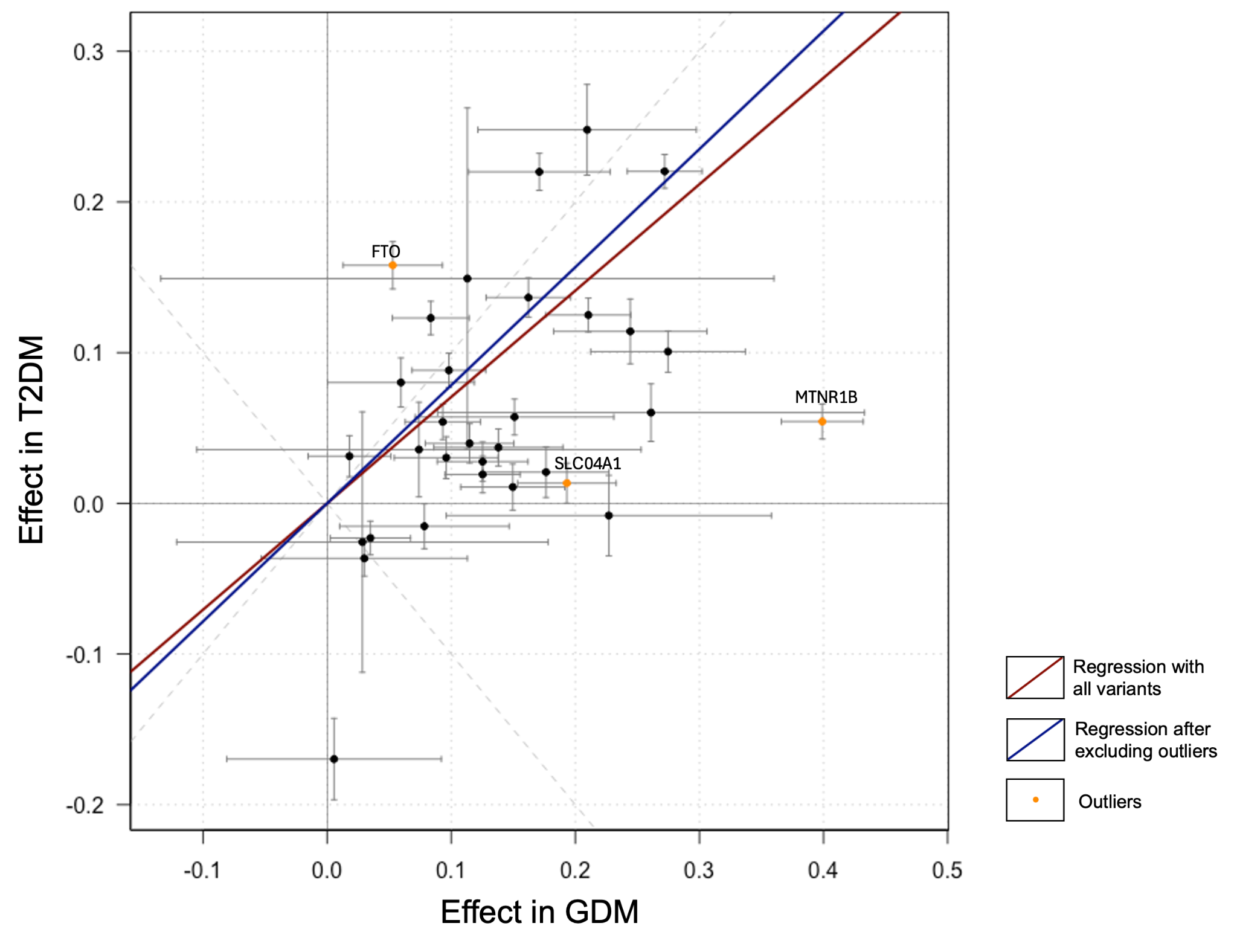

**Supplementary Figure 72*.*** *Comparison of genetic effect estimates for lead SNPs associated with gestational diabetes mellitus (GDM) and their corresponding effects on type 2 diabetes mellitus (T2DM; females only) in East Asians.* Genetic variants analysed were identified in the trans-ancestry GWAS meta-analysis of GDM (38,241 cases and 775,169 controls), with effect estimates and standard errors obtained from the East Asian GDM GWAS meta-analysis (4,346 cases, 23,286 controls). East Asian T2DM GWAS was obtained from Spracklen et al., 2020 (PMID: 32499647), and included 27,370 cases and 135,055 controls (females only, unadjusted for BMI). Effect estimates are presented as betas (log odds ratio). Error bars indicate +/- 1 standard error in each GWAS. Orange points labelled with putative gene names are identified as outliers by SCOUTJOY. Fitted slopes are shown from York regression with all variants (red) and after the removal of identified outliers (blue). Dashed gray reference lines indicate equal absolute effect size. SCOUTJOY analysis suggests a significant positive relationship in genetic effects before and after outlier removal. Analyses also suggest significant heterogeneity, identifying three outliers. Outliers detected include *FTO* (rs55872725), *SLCO4A1* (rs6122141), and *MTNR1B* (rs10830963). The following SNPs were not present in the GDM or T2DM dataset: rs41279738 (*GPR61*), rs142406759 (*RNF144B*), rs537224022 (*ESR1*), rs76895963 (*CCND2*), and rs77090130 (*PRAME*). T2DM; Type-II Diabetes Mellitus. GDM; Gestational Diabetes Mellitus.

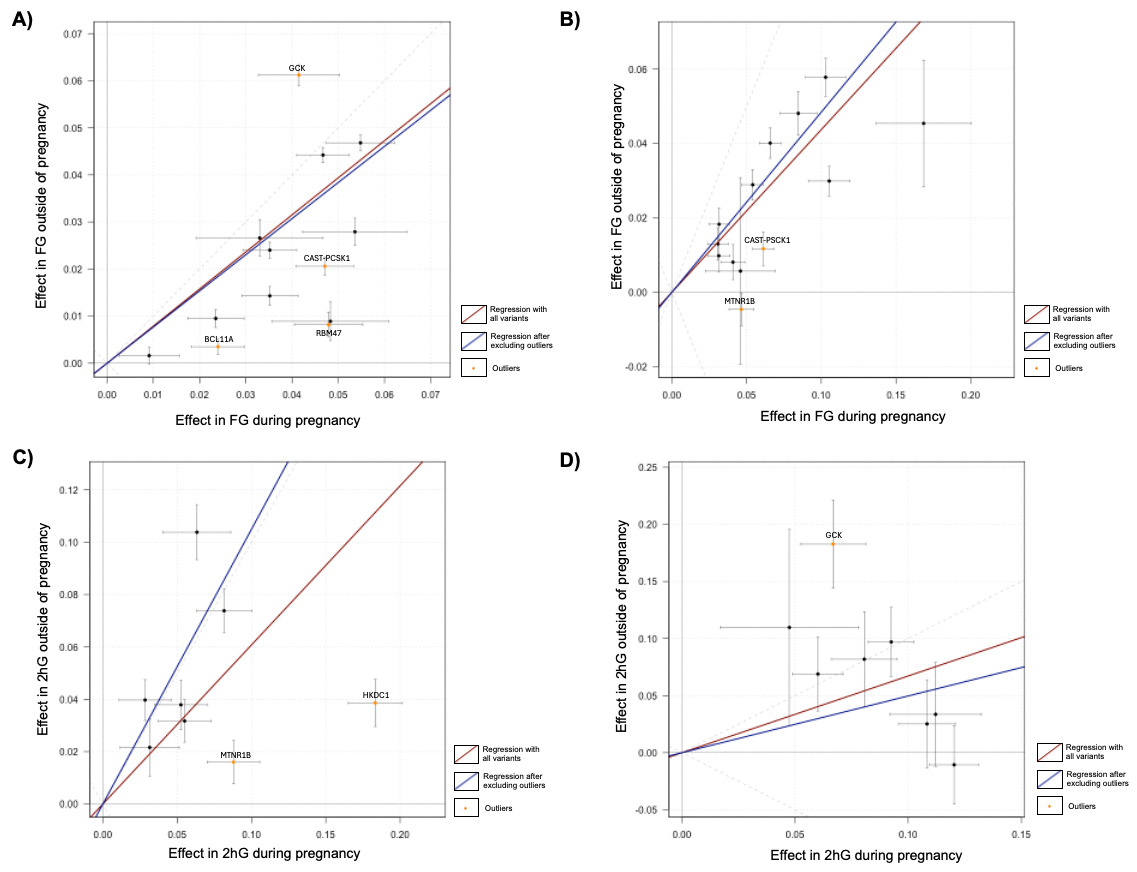

**Supplementary Figure 73.** *SCOUTJOY analyses comparing i) FG during and outside of pregnancy in Europeans (A) and East Asians (B) and ii) 2hG during and outside of pregnancy in Europeans (C) and East Asians (D).* Genetic variants analysed were identified in the trans-ancestry GWAS meta-analyses (overall N for FG being 55,371 and for 2hG 46,401), with effect estimates and standard errors obtained from the ancestry-specific GWAS meta-analyses. European GWAS sample sizes were as follows: FG (N=17,810) and 2hG (N=10,784) during pregnancy; FG (N= 200,622) and 2hG (N= 63,396) outside of pregnancy. East Asian GWAS sample sizes were as follows: FG (N=30,188) and 2hG (N=28,368) during pregnancy; FG (N= 35,619) and 2hG (N= 8,509) outside of pregnancy. GWAS of glycemic traits were conducted by the MAGIC consortium and adjusted for BMI. Error bars indicate +/- 1 standard error in each GWAS. Orange points labelled with variant names are identified as outliers by SCOUTJOY. Fitted slopes are shown from York regression with all variants (red) and after the removal of identified outliers (blue). Dashed gray reference lines indicate equal absolute effect size. SCOUTJOY analysis suggests a significant positive relationship of GDM with all four traits. Four outliers were identified when comparing the effect sizes of FG during and outside of pregnancy in Europeans, i.e. *GCK* (rs730497), *CAST* (rs4869273), *RBM47* (rs3579382), *MIR4432HG* (rs243020), whereas two were detected in the East Asian analysis *CAST-PSCK1* (rs4869273), and *MTNR1B* (rs3847554). Two outliers were identified when comparing the effect sizes of 2hG during and outside of pregnancy in Europeans, i.e., *MTNR1B* (rs7112766) and *HKDC1* (rs5030937), whilst only one was detected in the East Asian analysis i.e. *GCK* (rs2971670). GWAS of glycemic traits outside of pregnancy were obtained from Chen et al., 2021 (PMID: 34059833). FG; Fasting Glucose. 2hG; 2-hour glucose after oral glucose tolerance test (OGTT). BMI; Body Mass Index.

### **7.0 Shared variant analyses**

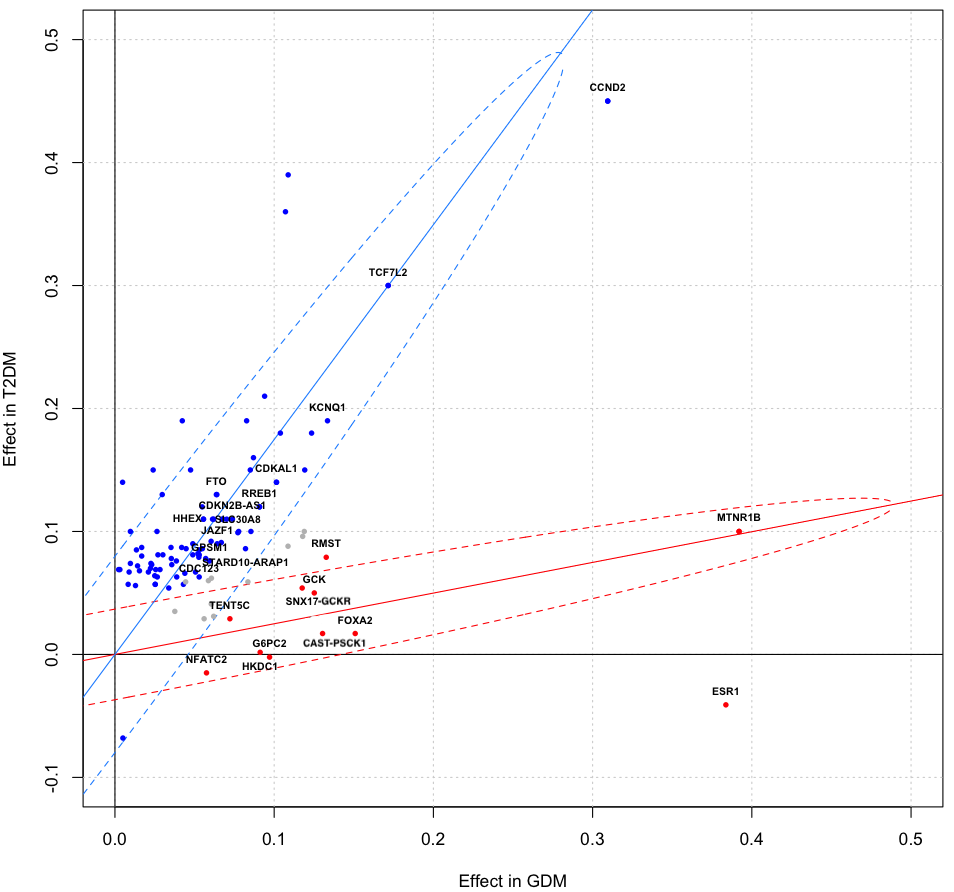

**Supplementary Figure 74.** *Comparison of log odds ratios in the GWAS of GDM (x-axis) and T2DM in European females (y-axis) for top-associated SNPs from GDM (34 SNPs) and T2DM (75 SNPs).* The following two distinct classes of SNP effects were identified by a Bayesian classifier in shared variants analysis: class T (blue) containing SNPs with T2DM-predominant genetic effects and class G (red), with GDM-predominant effects. Gray SNPs were not confidently assigned to either class (posterior probability >95%). Putative gene names are included in the figure for variants assigned to class T or class G. Dotted ellipses indicate the 95% probability regions of the fitted bivariate effect size distributions with each class. T2DM GWAS was obtained from Mahajan et al., 2018 (PMID: 30297969). T2DM; Type-II Diabetes Mellitus. GDM; Gestational Diabetes Mellitus.

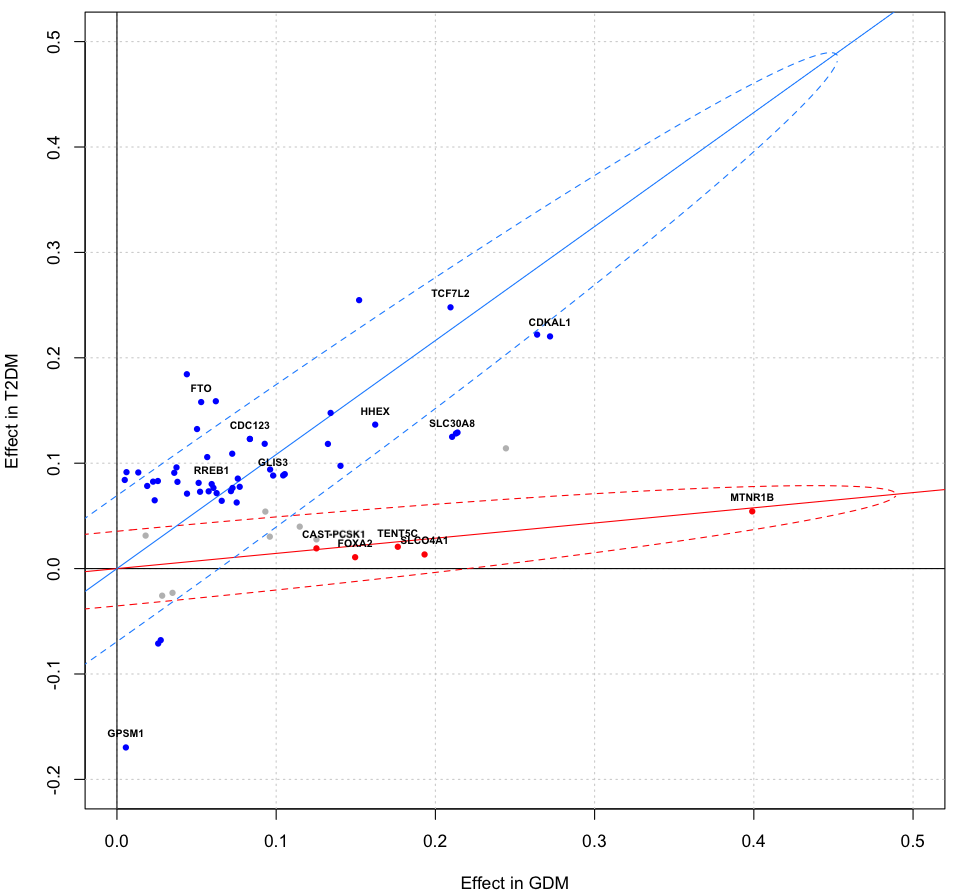

**Supplementary Figure 75.** *Comparison of log odds ratios in GWAS of GDM (x axis) and T2DM in East Asian females (y axis) for top-associated SNPs from GDM (22 SNPs) and T2D (40 SNPs).* The following two distinct classes of SNP effects were identified by a Bayesian classifier in shared variants analysis: class T (blue) containing SNPs with T2DM-predominant genetic effects and class G (red), with GDM-predominant effects. Gray SNPs were not confidently assigned to either class (posterior probability >95%). Putative gene names are included in the figure for variants assigned to class T or class G. Dotted ellipses indicate the 95% probability regions of the fitted bivariate effect size distributions with each class. T2DM GWAS was obtained from Spracklen et al., 2020 (PMID: 32499647). T2DM; Type-II Diabetes Mellitus. GDM; Gestational Diabetes Mellitus.

### **8.0 Colocalization analyses**

#### 8.1 Genetic signal for GDM only

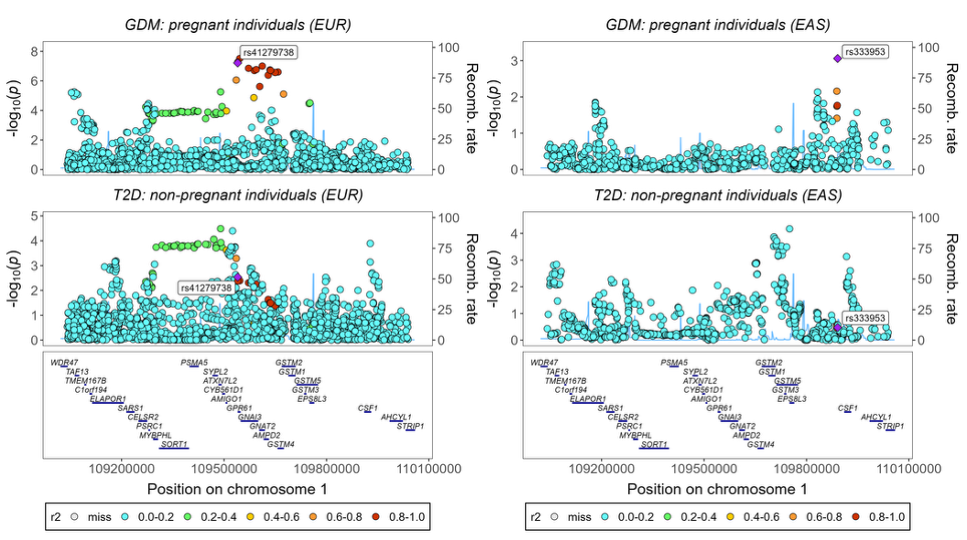

**Supplementary Figure 76.** *Regional association plots showing colocalization between GDM and T2DM in EUR (right) and EAS (left) populations near the GPR61 locus.* In EAS, the GDM lead variant rs41279738 was not present in the GDM and T2DM dataset or reference population; therefore, the variant with the lowest P-value in the GenDiP dataset is shown instead in the GDM and T2DM panels. Each variant's color represents its LD (r2) with the lead variant (purple diamond). The left y-axis shows recombination rate (cM/Mb), while the right y-axis displays -log10(P) values. Gene annotations appear below each plot. Blue shading in the background highlights recombination hotspots. Stacked points represent nearby variants (within ±500kb) with their respective association signals for GDM (top) and T2DM (bottom). LD; Linkage Disequilibrium. EUR; Europeans. EAS; East Asians.

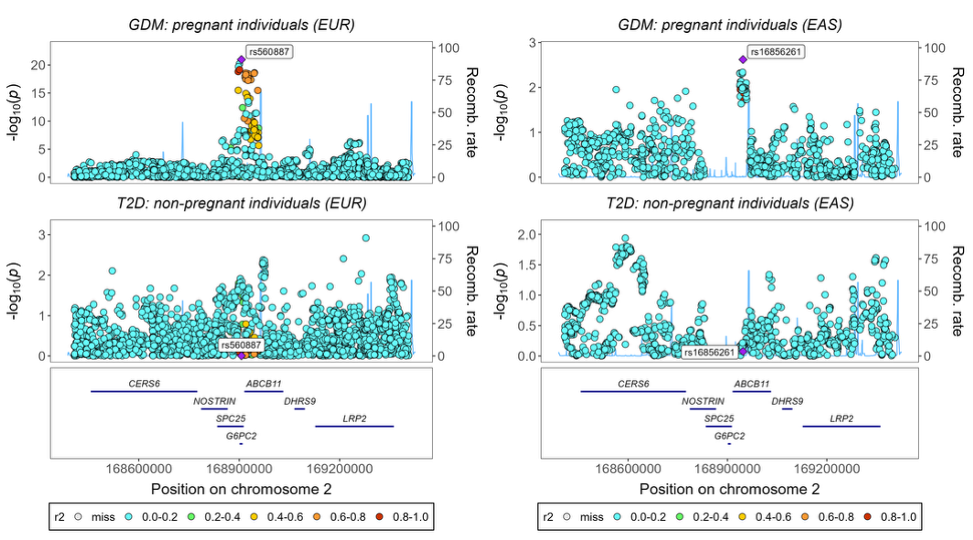

**Supplementary Figure 77.** *Regional association plots showing colocalization between GDM and T2DM in EUR (right) and EAS (left) populations near the G6PC2 locus.* In EUR, the lead variant rs560887 is highlighted (purple diamond). In EAS, rs560887 was not present in the GDM and T2DM dataset or reference population; therefore, the variant with the lowest P-value in the GenDiP dataset is shown instead in the GDM and T2DM panels. Each variant's color represents its LD (r2) with the lead variant (purple diamond). The left y-axis shows recombination rate (cM/Mb), while the right y-axis displays -log10(P) values. Gene annotations appear below each plot. Blue shading in the background highlights recombination hotspots. Stacked points represent nearby variants (within ±500kb) with their respective association signals for GDM (top) and T2DM (bottom). Gaps in the EAS GDM plot reflect variants removed during quality control (MAF <0.01 and sample size <50% of total sample). LD; Linkage Disequilibrium. EUR; Europeans. EAS; East Asians.

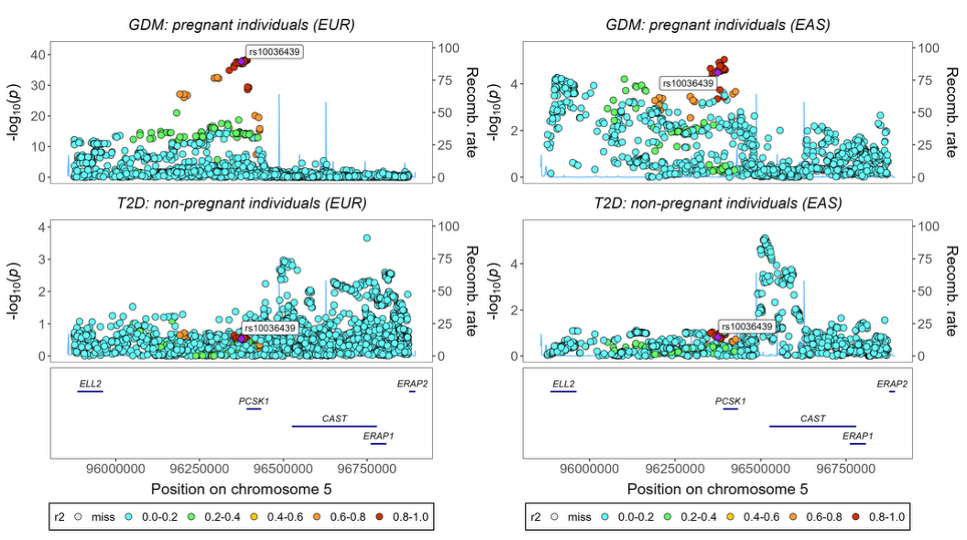

**Supplementary Figure 78.** *Regional association plots showing colocalization between GDM and T2DM in EUR (right) and EAS (left) populations for the lead variant rs10036439 (CAST-PSCK1).* Each variant's color represents its LD (r2) with the lead variant (purple diamond). The left y-axis shows recombination rate (cM/Mb), while the right y-axis displays -log10(P) values. Gene annotations appear below each plot. Blue shading in the background highlights recombination hotspots. Stacked points represent nearby variants (within ±500kb) with their respective association signals for GDM (top) and T2DM (bottom). LD; Linkage Disequilibrium. EUR; Europeans. EAS; East Asians.

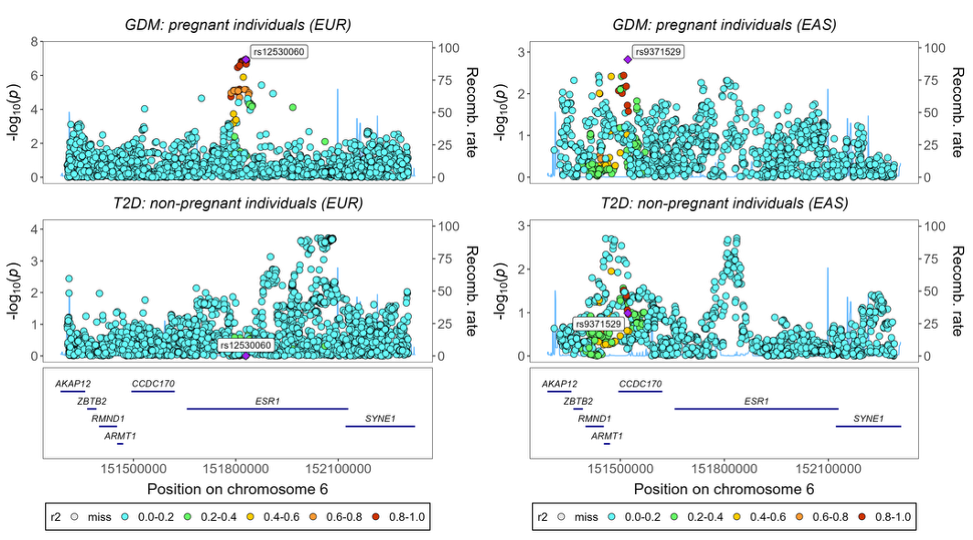

**Supplementary Figure 79.** *Regional association plots showing colocalization between GDM and T2DM in EUR (right) and EAS (left) populations near the ESR1 locus.* The original lead variant rs537224022 was unavailable in different datasets across populations: in EUR, it was absent from the T2DM dataset and reference population; in EAS, it was absent from the GDM dataset, T2DM dataset, and reference population. Therefore, the variant with the lowest P-value from the GenDiP dataset is displayed instead in both the GDM and T2DM panels. Each variant's color represents its LD (r2) with the lead variant (purple diamond). The left y-axis shows recombination rate (cM/Mb), while the right y-axis displays -log10(P) values. Gene annotations appear below each plot. Blue shading in the background highlights recombination hotspots. Stacked points represent nearby variants (within ±500kb) with their respective association signals for GDM (top) and T2DM (bottom). LD; Linkage Disequilibrium. EUR; Europeans. EAS; East Asians.

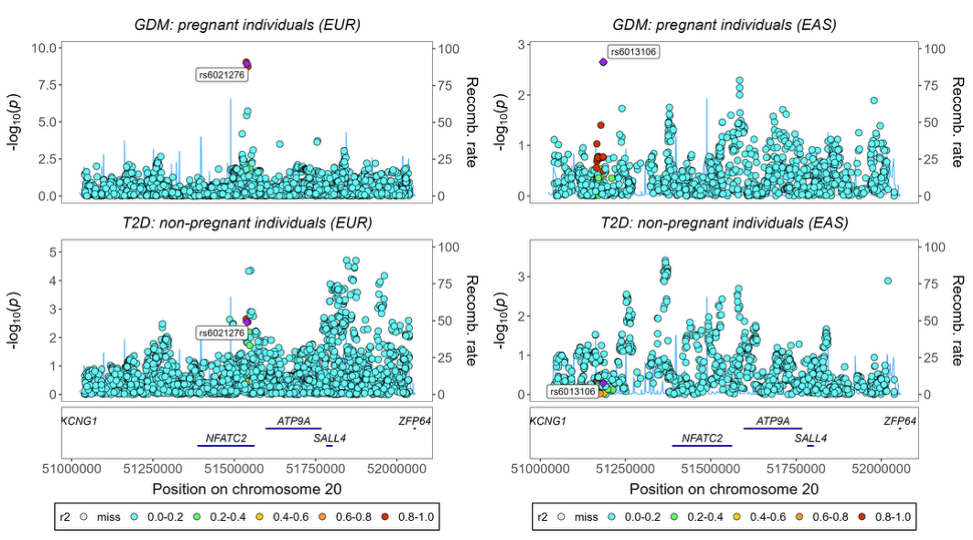

**Supplementary Figure 80.** *Regional association plots showing colocalization between GDM and T2DM in EUR (right) and EAS (left) populations near the NFATC2 locus.* In EUR, the lead variant rs6021276 is highlighted (purple diamond). In EAS, rs6021276 was not present in the GDM and T2DM dataset and reference population; therefore, the variant with the lowest P-value in the GenDiP dataset is shown instead in the GDM and T2DM panels. Each variant's color represents its LD (r2) with the lead variant (purple diamond). The left y-axis shows recombination rate (cM/Mb), while the right y-axis displays -log10(P) values. Gene annotations appear below each plot. Blue shading in the background highlights recombination hotspots. Stacked points represent nearby variants (within ±500kb) with their respective association signals for GDM (top) and T2DM (bottom). LD; Linkage Disequilibrium. EUR; Europeans. EAS; East Asians.

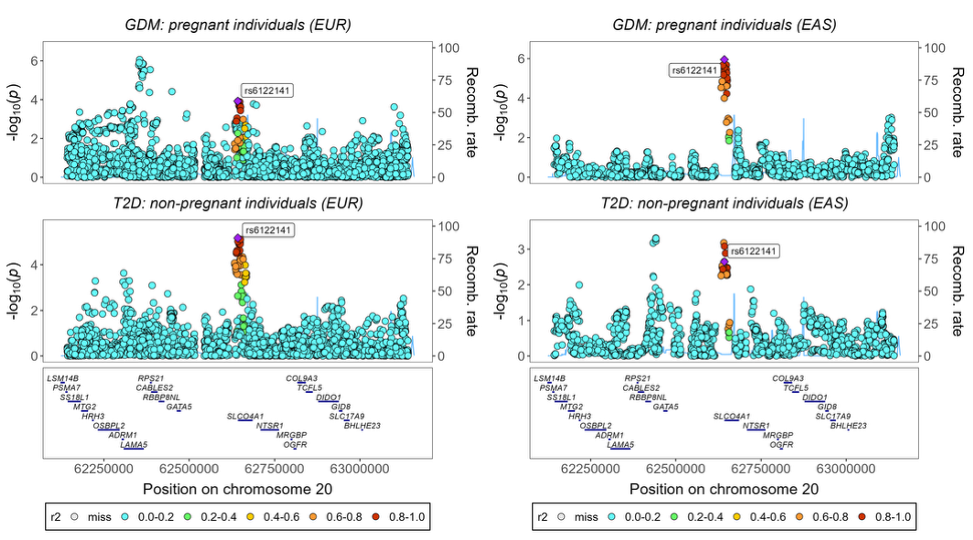

**Supplementary Figure 81.** *Regional association plots showing colocalization between GDM and T2DM in EUR (right) and EAS (left) populations for the lead variant rs6122141 (SLCO4A1).* Each variant's color represents its LD (r2) with the lead variant (purple diamond). The left y-axis shows recombination rate (cM/Mb), while the right y-axis displays -log10(P) values. Gene annotations appear below each plot. Blue shading in the background highlights recombination hotspots. Stacked points represent nearby variants (within ±500kb) with their respective association signals for GDM (top) and T2DM (bottom). LD; Linkage Disequilibrium. EUR; Europeans. EAS; East Asians.

#### 8.2 Different causal variants for GDM and T2DM

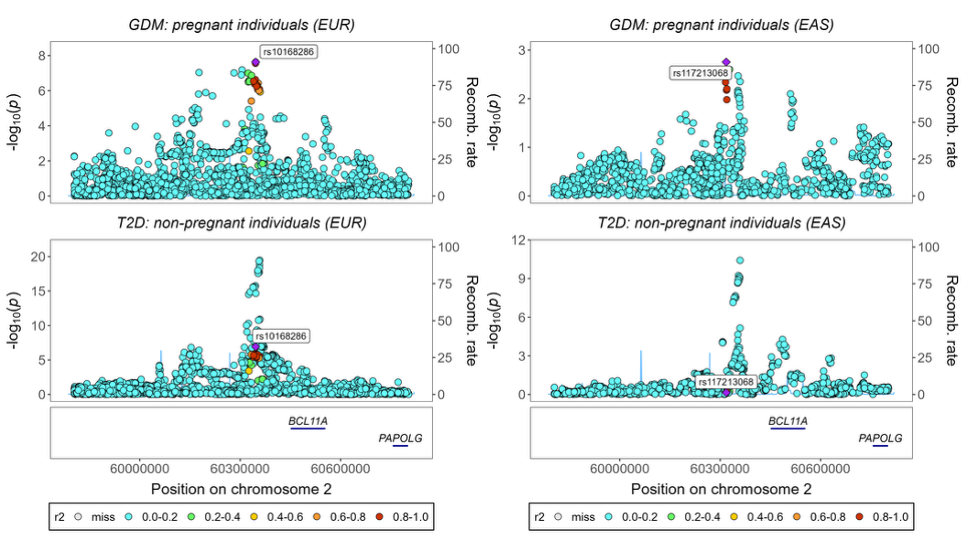

**Supplementary Figure 82.** *Regional association plots showing colocalization between GDM and T2DM in EUR (right) and EAS (left) populations near the BCL11A locus.* The original lead variant rs5831573 was unavailable in different datasets across populations: in EUR, it was absent from the T2DM dataset and reference population; in EAS, it was absent from the GDM dataset, T2DM dataset, and reference population. Therefore, the variant with the lowest P-value from the GenDiP dataset is displayed instead in both the GDM and T2DM panels. Each variant's color represents its LD (r2) with the lead variant (purple diamond). The left y-axis shows recombination rate (cM/Mb), while the right y-axis displays -log10(P) values. Gene annotations appear below each plot. Blue shading in the background highlights recombination hotspots. Stacked points represent nearby variants (within ±500kb) with their respective association signals for GDM (top) and T2DM (bottom). LD; Linkage Disequilibrium. EUR; Europeans. EAS; East Asians.

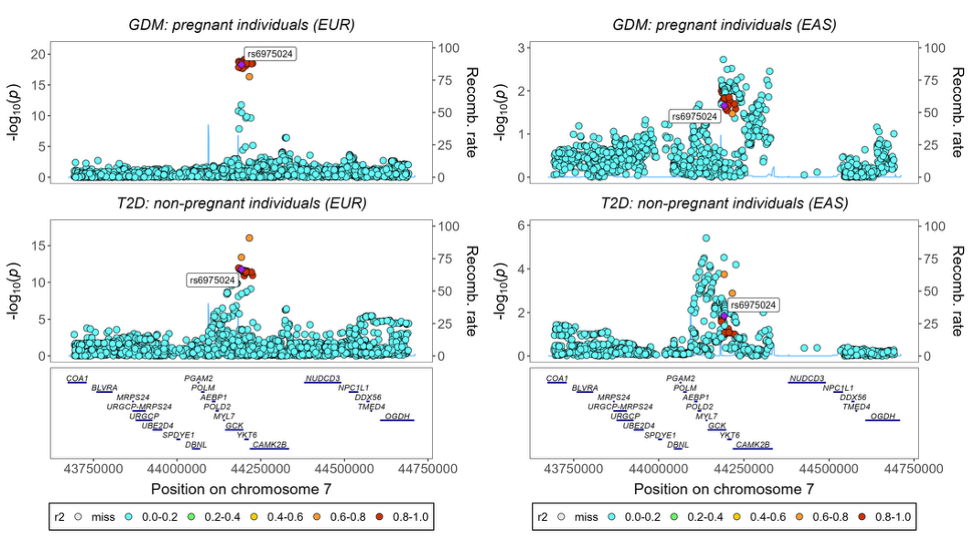

**Supplementary Figure 83.** *Regional association plots showing colocalization between GDM and T2DM in EUR (right) and EAS (left) populations for the lead variant rs6975024 (GCK).* Each variant's color represents its LD (r2) with the lead variant (purple diamond). The left y-axis shows recombination rate (cM/Mb), while the right y-axis displays -log10(P) values. Gene annotations appear below each plot. Blue shading in the background highlights recombination hotspots. Stacked points represent nearby variants (within ±500kb) with their respective association signals for GDM (top) and T2DM (bottom). Gaps in the EAS GDM plot reflect variants removed during quality control (MAF <0.01 and sample size <50% of total sample). LD; Linkage Disequilibrium. EUR; Europeans. EAS; East Asians.

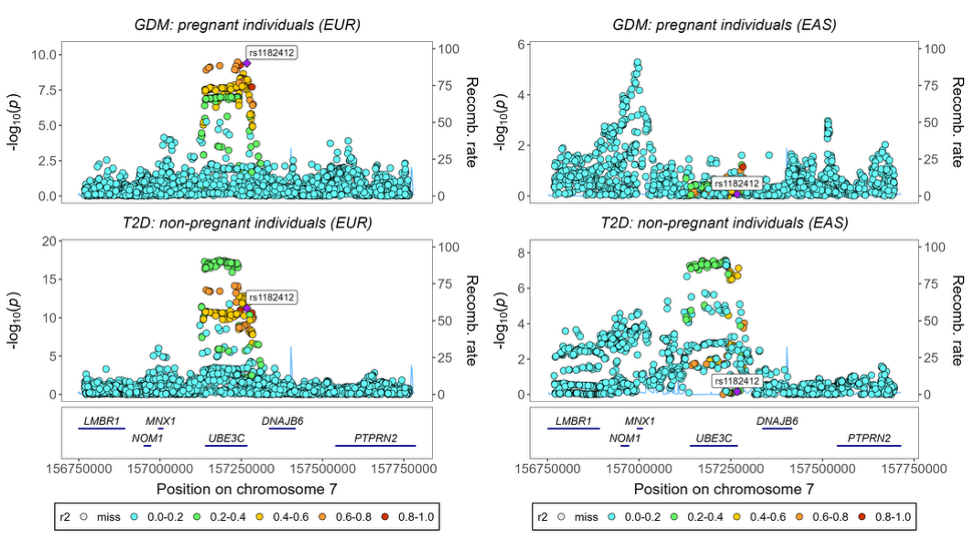

**Supplementary Figure 84.** *Regional association plots showing colocalization between GDM and T2DM in EUR (right) and EAS (left) populations for the lead variant rs1182412 (UBE3C).* Each variant's color represents its LD (r2) with the lead variant (purple diamond). The left y-axis shows recombination rate (cM/Mb), while the right y-axis displays -log10(P) values. Gene annotations appear below each plot. Blue shading in the background highlights recombination hotspots. Stacked points represent nearby variants (within ±500kb) with their respective association signals for GDM (top) and T2DM (bottom). LD; Linkage Disequilibrium. EUR; Europeans. EAS; East Asians.

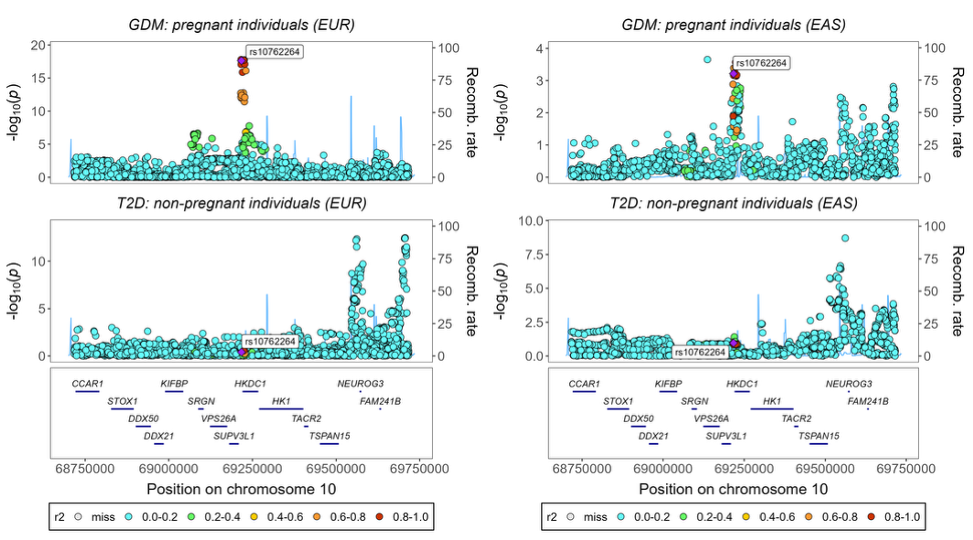

**Supplementary Figure 85.** *Regional association plots showing colocalization between GDM and T2DM in EUR (right) and EAS (left) populations for the lead variant rs10762264 (HKDC1).* Each variant's color represents its LD (r2) with the lead variant (purple diamond). The left y-axis shows recombination rate (cM/Mb), while the right y-axis displays -log10(P) values. Gene annotations appear below each plot. Blue shading in the background highlights recombination hotspots. Stacked points represent nearby variants (within ±500kb) with their respective association signals for GDM (top) and T2DM (bottom). LD; Linkage Disequilibrium. EUR; Europeans. EAS; East Asians.

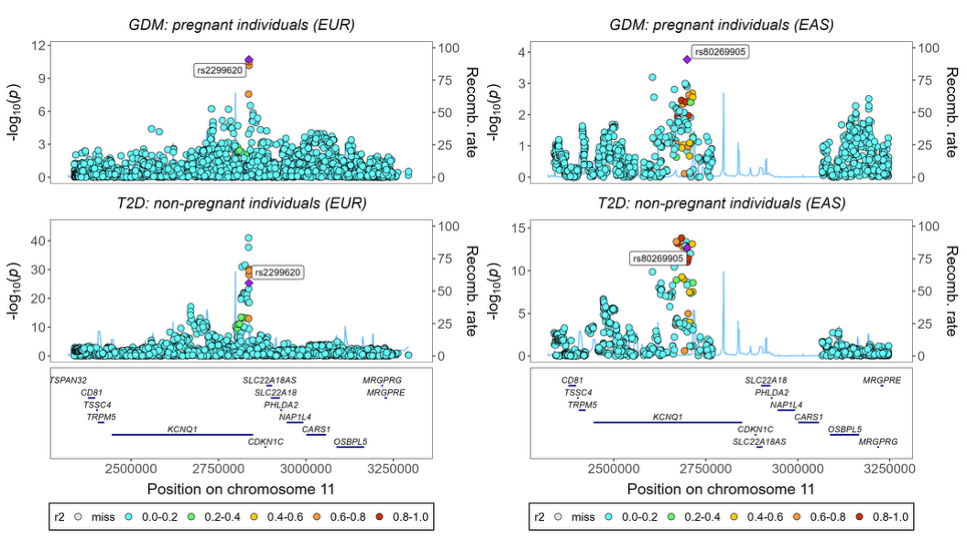

**Supplementary Figure 86.** *Regional association plots showing colocalization between GDM and T2DM in EUR (right) and EAS (left) populations near the KCNQ1 locus.* In EUR, the lead variant rs2299620 is highlighted (purple diamond). In EAS, rs2299620 was not present in the GDM and T2DM dataset and was absent from the reference population; therefore, the variant with the lowest P-value in the GenDiP dataset is shown instead in the GDM and T2DM panels. Each variant's color represents its LD (r2) with the lead variant (purple diamond). The left y-axis shows recombination rate (cM/Mb), while the right y-axis displays -log10(P) values. Gene annotations appear below each plot. Blue shading in the background highlights recombination hotspots. Gaps in the EAS GDM plot reflect variants removed during quality control (MAF <0.01 and sample size <50% of total sample). Stacked points represent nearby variants (within ±500kb) with their respective association signals for GDM (top) and T2DM (bottom). LD; Linkage Disequilibrium. EUR; Europeans. EAS; East Asians.

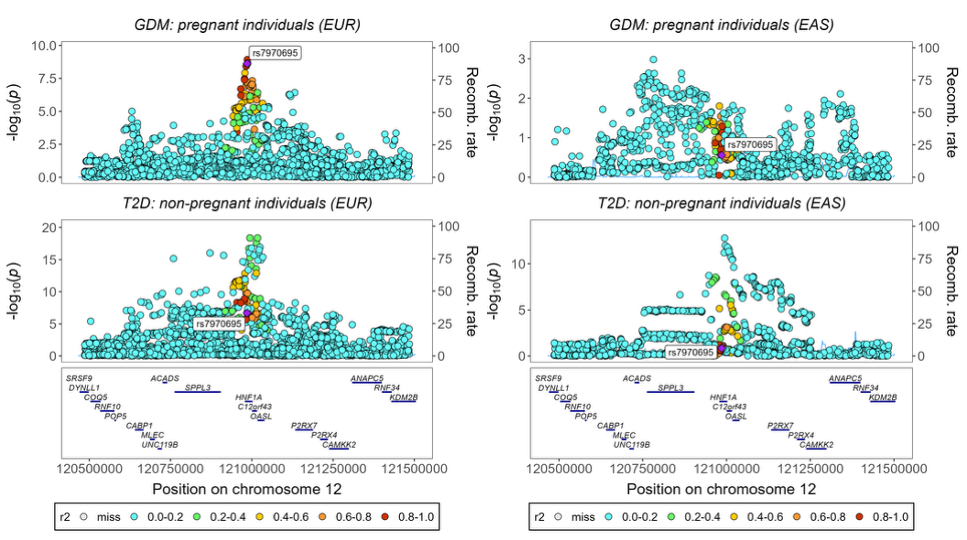

**Supplementary Figure 87.** *Regional association plots showing colocalization between GDM and T2DM in EUR (right) and EAS (left) populations for the lead variant rs7970695 (HNF1A).* Each variant's color represents its LD (r2) with the lead variant (purple diamond). The left y-axis shows recombination rate (cM/Mb), while the right y-axis displays -log10(P) values. Gene annotations appear below each plot. Blue shading in the background highlights recombination hotspots. Stacked points represent nearby variants (within ±500kb) with their respective association signals for GDM (top) and T2DM (bottom). LD; Linkage Disequilibrium. EUR; Europeans. EAS; East Asians.

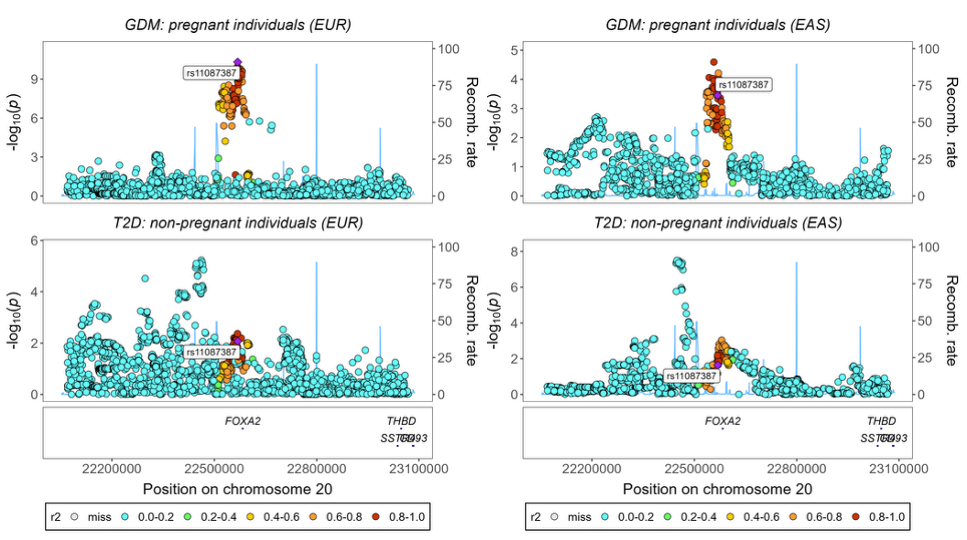

**Supplementary Figure 88.** *Regional association plots showing colocalization between GDM and T2DM in EUR (right) and EAS (left) populations for the lead variant rs11087387 (FOXA2).* Each variant's color represents its LD (r2) with the lead variant (purple diamond). The left y-axis shows recombination rate (cM/Mb), while the right y-axis displays -log10(P) values. Gene annotations appear below each plot. Blue shading in the background highlights recombination hotspots. Stacked points represent nearby variants (within ±500kb) with their respective association signals for GDM (top) and T2DM (bottom). LD; Linkage Disequilibrium. EUR; Europeans. EAS; East Asians.

#### 8.3 Shared causal variants for GDM and T2DM

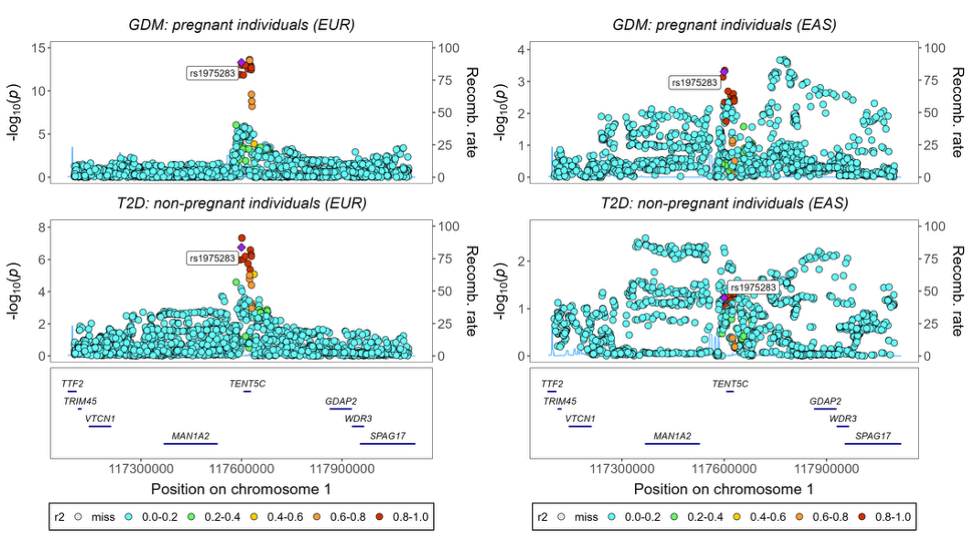

**Supplementary Figure 89.** *Regional association plots showing colocalization between GDM and T2DM in EUR (right) and EAS (left) populations for the lead variant rs1975283 (TENT5C).* Each variant's color represents its LD (r2) with the lead variant (purple diamond). The left y-axis shows recombination rate (cM/Mb), while the right y-axis displays -log10(P) values. Gene annotations appear below each plot. Blue shading in the background highlights recombination hotspots. Stacked points represent nearby variants (within ±500kb) with their respective association signals for GDM (top) and T2DM (bottom). LD; Linkage Disequilibrium. EUR; Europeans. EAS; East Asians.

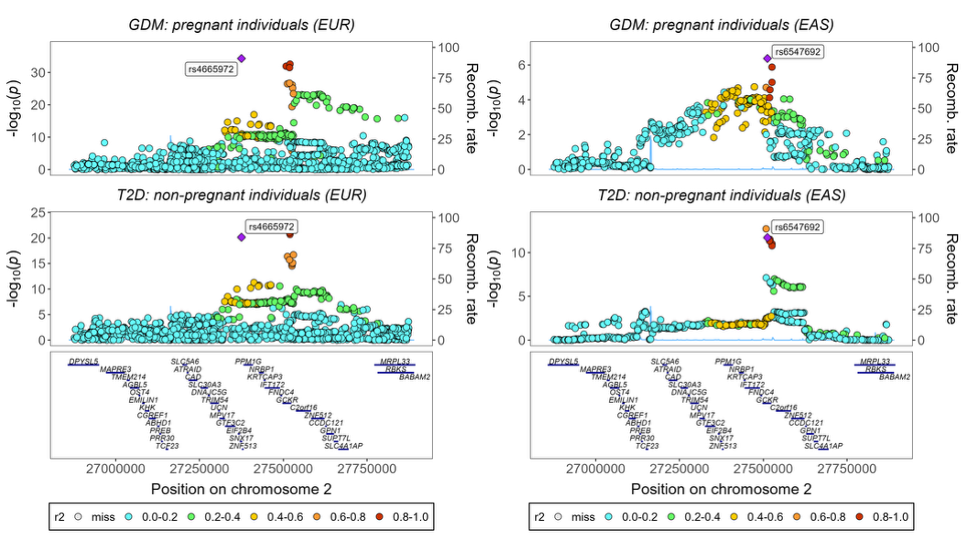

**Supplementary Figure 90.** *Regional association plots showing colocalization between GDM and T2DM in EUR (right) and EAS (left) populations near the SNX17-GCKR locus.* In EUR, the lead variant rs4665972 is highlighted (purple diamond). In EAS, rs4665972 was not present in the GDM and T2DM dataset and was absent from reference population; therefore, the variant with the lowest P-value in the GenDiP dataset is shown instead in the GDM and T2DM panels. Each variant's color represents its LD (r2) with the lead variant (purple diamond). The left y-axis shows recombination rate (cM/Mb), while the right y-axis displays -log10(P) values. Gene annotations appear below each plot. Blue shading in the background highlights recombination hotspots. Stacked points represent nearby variants (within ±500kb) with their respective association signals for GDM (top) and T2DM (bottom). LD; Linkage Disequilibrium. EUR; Europeans. EAS; East Asians.

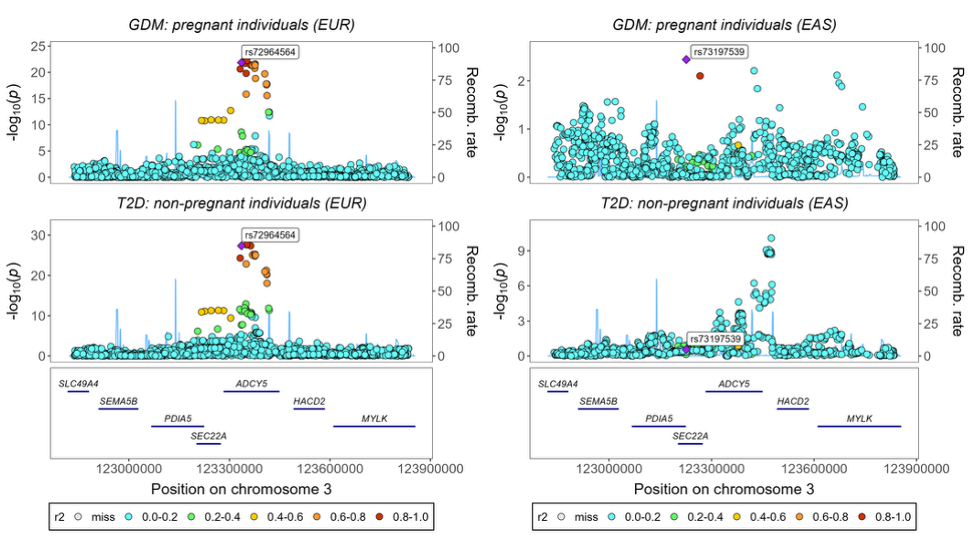

**Supplementary Figure 91.** *Regional association plots showing colocalization between GDM and T2DM in EUR (right) and EAS (left) populations near the ADCY5 locus.* In EUR, the lead variant rs72964564 is highlighted (purple diamond). In EAS, rs72964564 was not present in the GDM and T2DM dataset and was absent from reference population; therefore, the variant with the lowest P-value in the GenDiP dataset is shown instead in the GDM and T2DM panels. Each variant's color represents its LD (r2) with the lead variant (purple diamond). The left y-axis shows recombination rate (cM/Mb), while the right y-axis displays -log10(P) values. Gene annotations appear below each plot. Blue shading in the background highlights recombination hotspots. Stacked points represent nearby variants (within ±500kb) with their respective association signals for GDM (top) and T2DM (bottom). LD; Linkage Disequilibrium. EUR; Europeans. EAS; East Asians.

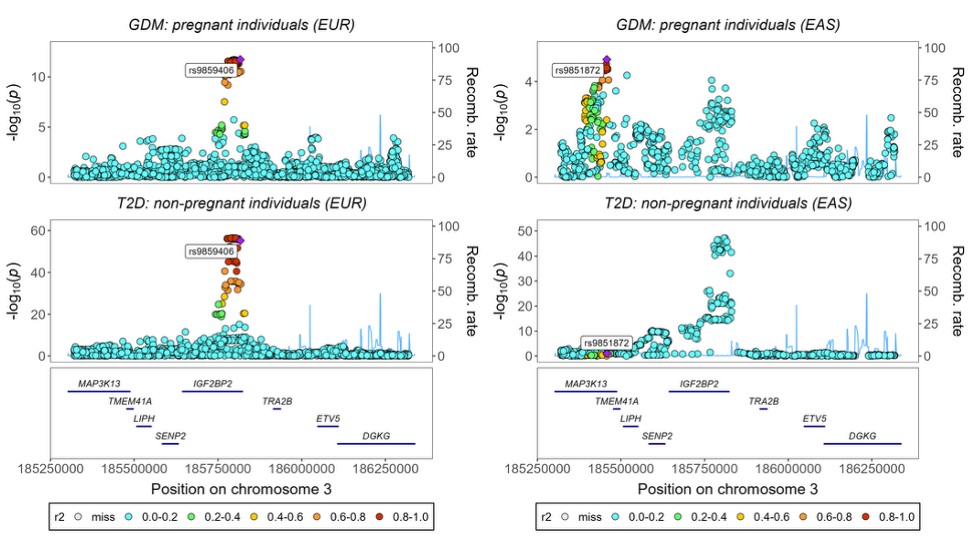

**Supplementary Figure 92.** *Regional association plots showing colocalization between GDM and T2DM in EUR (right) and EAS (left) near the IGF2BP2 locus.* The original lead variant rs113851927 was unavailable in different datasets across populations: in EUR, it was absent from the T2DM dataset and reference population; in EAS, it was absent from the GDM dataset, T2DM dataset, and reference population. Therefore, the variant with the lowest P-value from the GenDiP dataset is displayed instead in both the GDM and T2DM panels. Each variant's color represents its LD (r2) with the lead variant (purple diamond). The left y-axis shows recombination rate (cM/Mb), while the right y-axis displays -log10(P) values. Gene annotations appear below each plot. Blue shading in the background highlights recombination hotspots. Stacked points represent nearby variants (within ±500kb) with their respective association signals for GDM (top) and T2DM (bottom). LD; Linkage Disequilibrium. EUR; Europeans. EAS; East Asians.

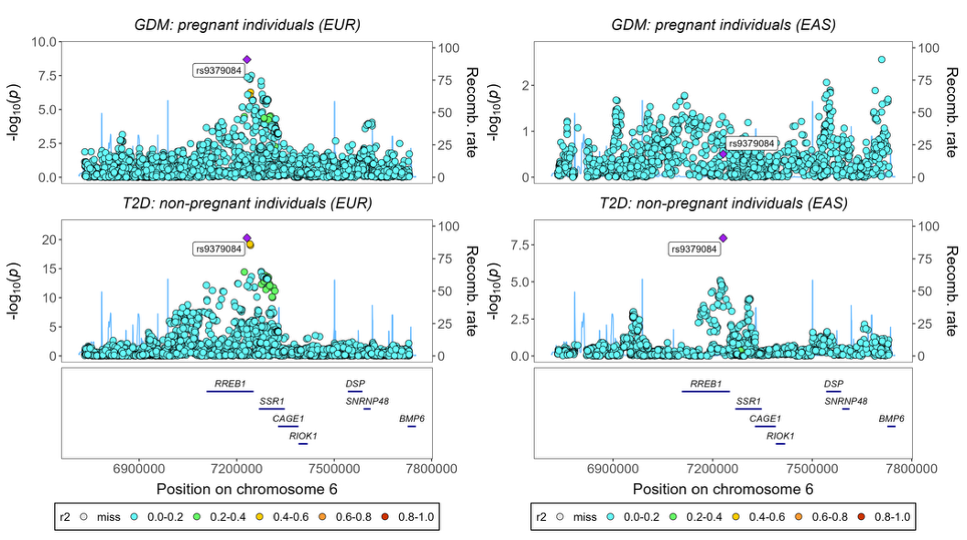

**Supplementary Figure 93.** *Regional association plots showing colocalization between GDM and T2DM in EUR (right) and EAS (left) populations for the lead variant rs9379084 (RREB1).* Each variant's color represents its LD (r2) with the lead variant (purple diamond). The left y-axis shows recombination rate (cM/Mb), while the right y-axis displays -log10(P) values. Gene annotations appear below each plot. Blue shading in the background highlights recombination hotspots. Stacked points represent nearby variants (within ±500kb) with their respective association signals for GDM (top) and T2DM (bottom). LD; Linkage Disequilibrium. EUR; Europeans. EAS; East Asians.

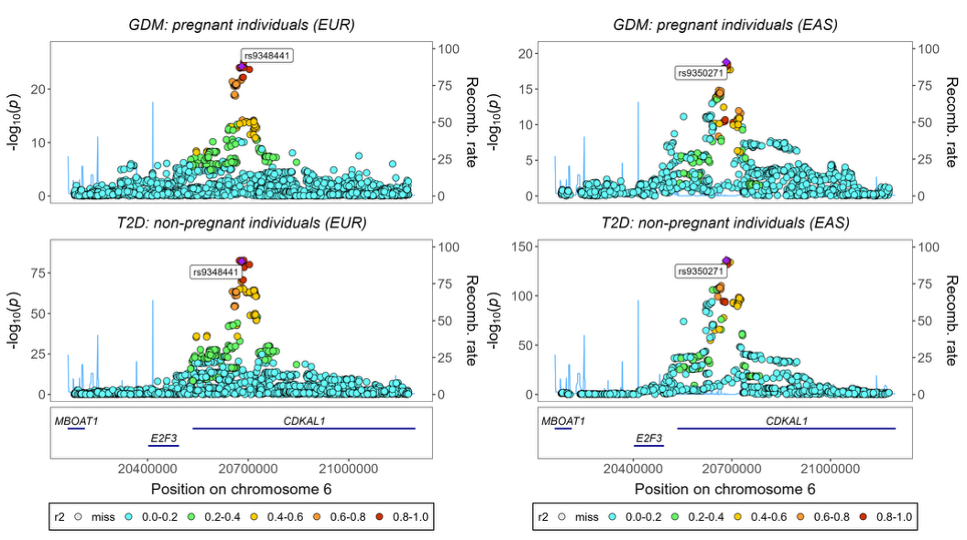

**Supplementary Figure 94.** *Regional association plots showing colocalization between GDM and T2DM in EUR (right) and EAS (left) populations near the CDKAL1 locus.* In EUR, the lead variant rs9348441 is highlighted (purple diamond). In EAS, rs9348441 was not present in the population reference dataset; therefore, the variant with the lowest P-value in the GenDiP dataset is shown instead in the GDM and T2DM panels. Each variant's color represents its LD (r2) with the lead variant (purple diamond). The left y-axis shows recombination rate (cM/Mb), while the right y-axis displays -log10(P) values. Gene annotations appear below each plot. Blue shading in the background highlights recombination hotspots. Stacked points represent nearby variants (within ±500kb) with their respective association signals for GDM (top) and T2DM (bottom). LD; Linkage Disequilibrium. EUR; Europeans. EAS; East Asians.

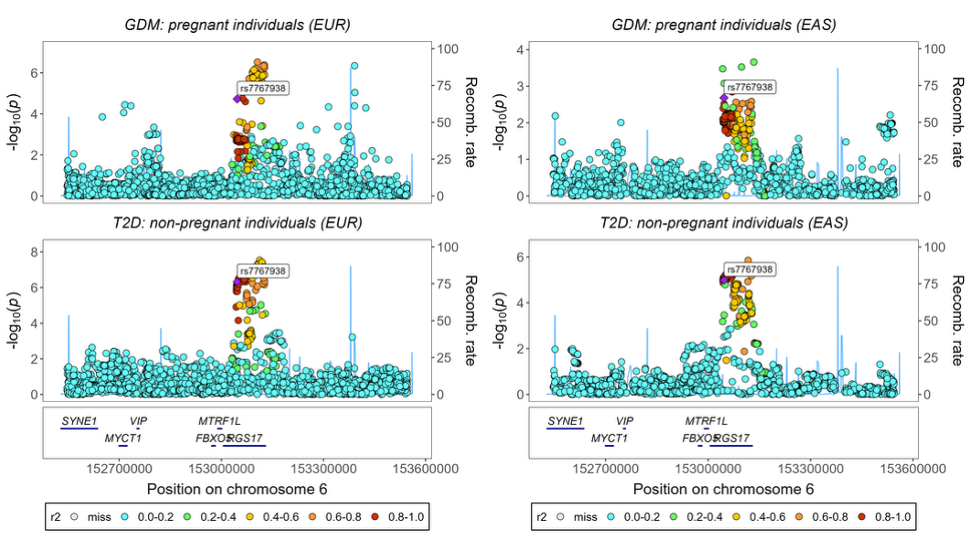

**Supplementary Figure 95.** *Regional association plots showing colocalization between GDM and T2DM in EUR (right) and EAS (left) populations for the lead variant rs7767938 (RGS17).* Each variant's color represents its LD (r2) with the lead variant (purple diamond). The left y-axis shows recombination rate (cM/Mb), while the right y-axis displays -log10(P) values. Gene annotations appear below each plot. Blue shading in the background highlights recombination hotspots. Stacked points represent nearby variants (within ±500kb) with their respective association signals for GDM (top) and T2DM (bottom). LD; Linkage Disequilibrium. EUR; Europeans. EAS; East Asians.

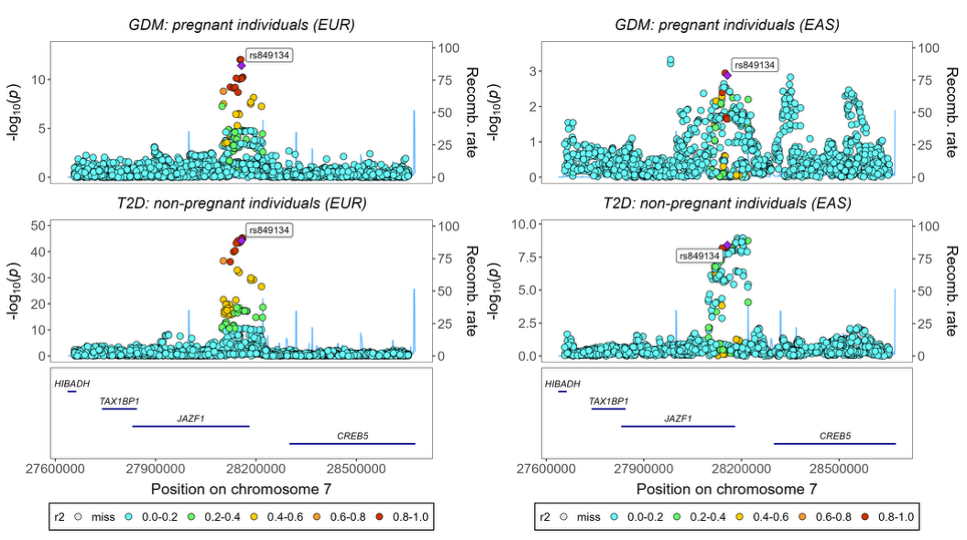

**Supplementary Figure 96.** *Regional association plots showing colocalization between GDM and T2DM in EUR (right) and EAS (left) populations for the lead variant rs849134 (JAZF1).* Each variant's color represents its LD (r2) with the lead variant (purple diamond). The left y-axis shows recombination rate (cM/Mb), while the right y-axis displays -log10(P) values. Gene annotations appear below each plot. Blue shading in the background highlights recombination hotspots. Stacked points represent nearby variants (within ±500kb) with their respective association signals for GDM (top) and T2DM (bottom). LD; Linkage Disequilibrium. EUR; Europeans. EAS; East Asians.

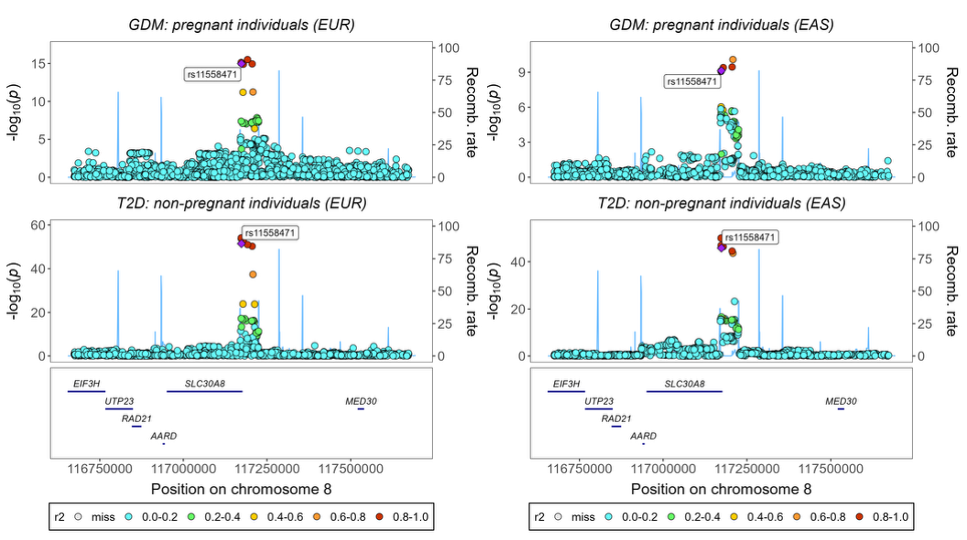

**Supplementary Figure 97.** *Regional association plots showing colocalization between GDM and T2DM in EUR (right) and EAS (left) populations for the lead variant rs11558471 (SLC30A8).* Each variant's color represents its LD (r2) with the lead variant (purple diamond). The left y-axis shows recombination rate (cM/Mb), while the right y-axis displays -log10(P) values. Gene annotations appear below each plot. Blue shading in the background highlights recombination hotspots. Stacked points represent nearby variants (within ±500kb) with their respective association signals for GDM (top) and T2DM (bottom). LD; Linkage Disequilibrium. EUR; Europeans. EAS; East Asians.

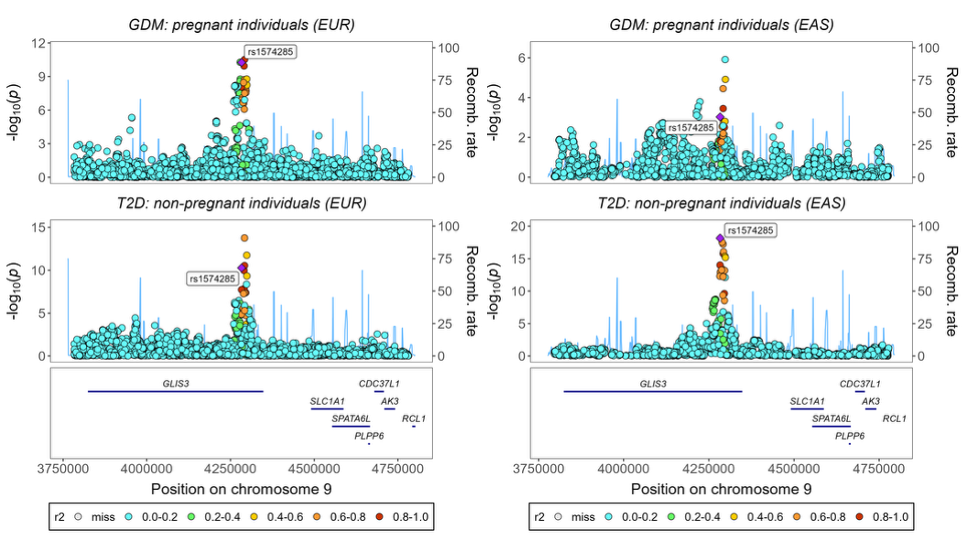

**Supplementary Figure 98.** *Regional association plots showing colocalization between GDM and T2DM in EUR (right) and EAS (left) populations for the lead variant rs1574285 (GLIS3).* Each variant's color represents its LD (r2) with the lead variant (purple diamond). The left y-axis shows recombination rate (cM/Mb), while the right y-axis displays -log10(P) values. Gene annotations appear below each plot. Blue shading in the background highlights recombination hotspots. Stacked points represent nearby variants (within ±500kb) with their respective association signals for GDM (top) and T2DM (bottom). LD; Linkage Disequilibrium. EUR; Europeans. EAS; East Asians.

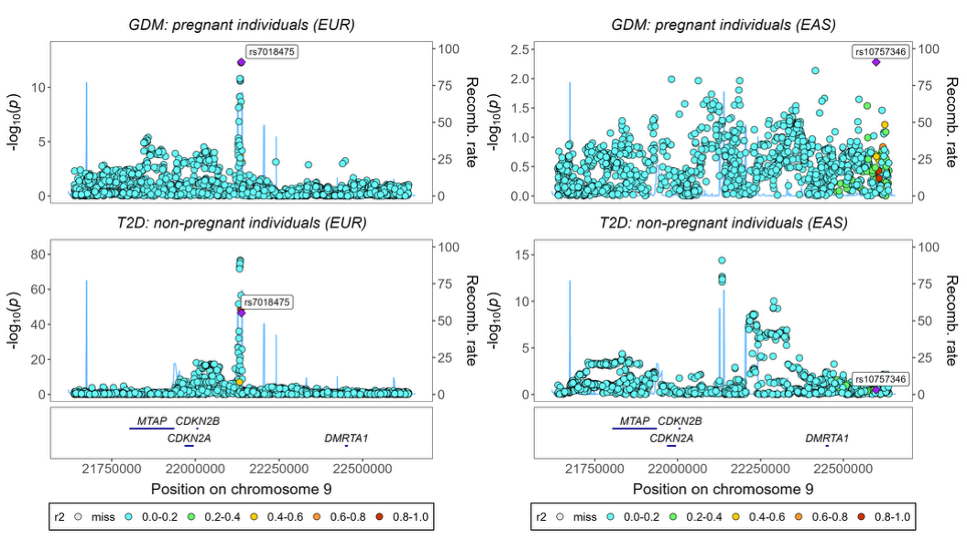

**Supplementary Figure 99.** *Regional association plots showing colocalization between GDM and T2DM in EUR (right) and EAS (left) populations near the CDKN2B-AS1 locus.* In EUR, the lead variant rs7018475 is highlighted (purple diamond). In EAS, rs7018475 was not present in the GDM and T2DM datasets and was absent from the reference population dataset; therefore, the variant with the lowest P-value in the GenDiP dataset is shown instead in the GDM and T2DM panels. Each variant's color represents its LD (r2) with the lead variant (purple diamond). The left y-axis shows recombination rate (cM/Mb), while the right y-axis displays -log10(P) values. Gene annotations appear below each plot. Blue shading in the background highlights recombination hotspots. Stacked points represent nearby variants (within ±500kb) with their respective association signals for GDM (top) and T2DM (bottom). LD; Linkage Disequilibrium. EUR; Europeans. EAS; East Asians.

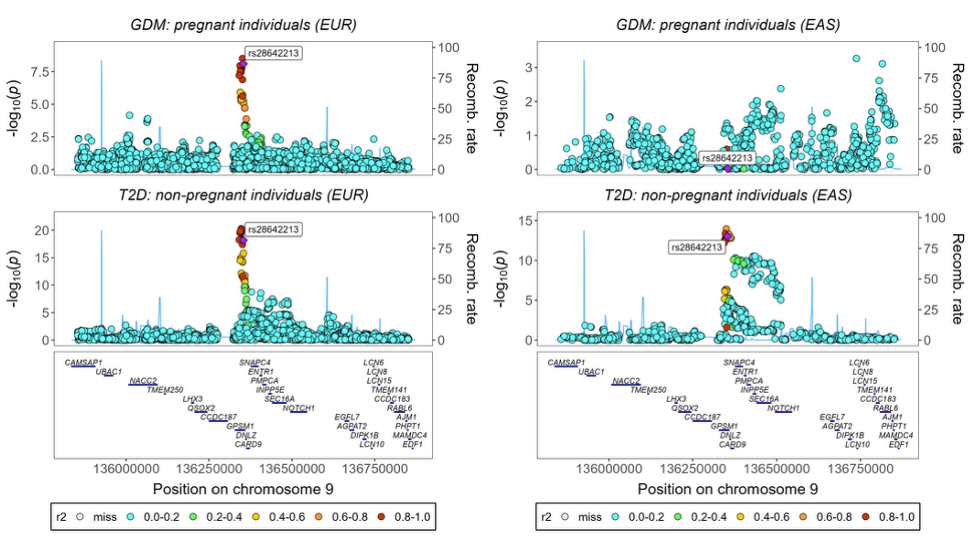

**Supplementary Figure 100.** *Regional association plots showing colocalization between GDM and T2DM in EUR (right) and EAS (left) populations for the lead variant rs28642213 (GPSM1).* Each variant's color represents its LD (r2) with the lead variant (purple diamond). The left y-axis shows recombination rate (cM/Mb), while the right y-axis displays -log10(P) values. Gene annotations appear below each plot. Blue shading in the background highlights recombination hotspots. Stacked points represent nearby variants (within ±500kb) with their respective association signals for GDM (top) and T2DM (bottom). LD; Linkage Disequilibrium. EUR; Europeans. EAS; East Asians.

**Supplementary Figure 101.** *Regional association plots showing colocalization between GDM and T2DM in EUR (right) and EAS (left) populations for the lead variant rs11257655 (CDC123).* Each variant's color represents its LD (r2) with the lead variant (purple diamond). The left y-axis shows recombination rate (cM/Mb), while the right y-axis displays -log10(P) values. Gene annotations appear below each plot. Blue shading in the background highlights recombination hotspots. Stacked points represent nearby variants (within ±500kb) with their respective association signals for GDM (top) and T2DM (bottom). LD; Linkage Disequilibrium. EUR; Europeans. EAS; East Asians.

**Supplementary Figure 102.** *Regional association plots showing colocalization between GDM and T2DM in EUR (right) and EAS (left) populations for the lead variant rs11187141 (HHEX).* Each variant's color represents its LD (r2) with the lead variant (purple diamond). The left y-axis shows recombination rate (cM/Mb), while the right y-axis displays -log10(P) values. Gene annotations appear below each plot. Blue shading in the background highlights recombination hotspots. Stacked points represent nearby variants (within ±500kb) with their respective association signals for GDM (top) and T2DM (bottom). LD; Linkage Disequilibrium. EUR; Europeans. EAS; East Asians.

**Supplementary Figure 103.** *Regional association plots showing colocalization between GDM and T2DM in EUR (right) and EAS (left) populations for the lead variant rs7903146 (TCF7L2).* Each variant's color represents its LD (r2) with the lead variant (purple diamond). The left y-axis shows recombination rate (cM/Mb), while the right y-axis displays -log10(P) values. Gene annotations appear below each plot. Blue shading in the background highlights recombination hotspots. Stacked points represent nearby variants (within ±500kb) with their respective association signals for GDM (top) and T2DM (bottom). LD; Linkage Disequilibrium. EUR; Europeans. EAS; East Asians.

**Supplementary Figure 104.** *Regional association plots showing colocalization between GDM and T2DM in EUR (right) and EAS (left) populations near the CRY2 locus.* In EUR, rs7933420 was not present in population reference datatset; therefore, the variant with the lowest P-value in the GenDiP dataset is shown instead in the GDM and T2DM panels. Each variant's color represents its LD (r2) with the lead variant (purple diamond). The left y-axis shows recombination rate (cM/Mb), while the right y-axis displays -log10(P) values. Gene annotations appear below each plot. Blue shading in the background highlights recombination hotspots. Stacked points represent nearby variants (within ±500kb) with their respective association signals for GDM (top) and T2DM (bottom). LD; Linkage Disequilibrium. EUR; Europeans. EAS; East Asians.

**Supplementary Figure 105.** *Regional association plots showing colocalization between GDM and T2DM in EUR (right) and EAS (left) populations for the lead variant rs613937 (STARD10-ARAP1).* Each variant's color represents its LD (r2) with the lead variant (purple diamond). The left y-axis shows recombination rate (cM/Mb), while the right y-axis displays -log10(P) values. Gene annotations appear below each plot. Blue shading in the background highlights recombination hotspots. Stacked points represent nearby variants (within ±500kb) with their respective association signals for GDM (top) and T2DM (bottom). LD; Linkage Disequilibrium. EUR; Europeans. EAS; East Asians.

**Supplementary Figure 106.** *Regional association plots showing colocalization between GDM and T2DM in EUR (right) and EAS (left) populations near the MTNR1B locus.* In EUR, the lead variant rs10830963 is highlighted (purple diamond). In EAS, rs10830963 was not present in the reference dataset; therefore, the variant with the lowest P-value in the GenDiP dataset is shown instead in the GDM and T2DM panels. Each variant's color represents its LD (r2) with the lead variant (purple diamond). The left y-axis shows recombination rate (cM/Mb), while the right y-axis displays -log10(P) values. Gene annotations appear below each plot. Blue shading in the background highlights recombination hotspots. Stacked points represent nearby variants (within ±500kb) with their respective association signals for GDM (top) and T2DM (bottom). LD; Linkage Disequilibrium. EUR; Europeans. EAS; East Asians.

**Supplementary Figure 107.** *Regional association plots showing colocalization between GDM and T2DM in EUR (right) and EAS (left) populations near the CCND2 locus.* In EUR, the lead variant rs76895963 is highlighted (purple diamond). In EAS, rs76895963 was not present in the GDM and T2DM dataset and was absent from the population reference dataset; therefore, the variant with the lowest P-value in the GenDiP dataset is shown instead in the GDM and T2DM panels. Each variant's color represents its LD (r2) with the lead variant (purple diamond). The left y-axis shows recombination rate (cM/Mb), while the right y-axis displays -log10(P) values. Gene annotations appear below each plot. Blue shading in the background highlights recombination hotspots. Stacked points represent nearby variants (within ±500kb) with their respective association signals for GDM (top) and T2DM (bottom). LD; Linkage Disequilibrium. EUR; Europeans. EAS; East Asians.

**Supplementary Figure 108.** *Regional association plots showing colocalization between GDM and T2DM in EUR (right) and EAS (left) populations near the RMST locus.* In EUR, the lead variant rs77864822 is highlighted (purple diamond). In EAS, rs77864822 was not present in the GDM and T2DM dataset and was absent from the population reference dataset; therefore, the variant with the lowest P-value in the GenDiP dataset is shown instead in the GDM and T2DM panels. Each variant's color represents its LD (r2) with the lead variant (purple diamond). The left y-axis shows recombination rate (cM/Mb), while the right y-axis displays -log10(P) values. Gene annotations appear below each plot. Blue shading in the background highlights recombination hotspots. Stacked points represent nearby variants (within ±500kb) with their respective association signals for GDM (top) and T2DM (bottom). LD; Linkage Disequilibrium. EUR; Europeans. EAS; East Asians.

**Supplementary Figure 109.** *Regional association plots showing colocalization between GDM and T2DM in EUR (right) and EAS (left) populations near the FBRSL1 locus.* In EUR, the lead variant rs3751297 is highlighted (purple diamond). In EAS, rs3751297 was not present in the GDM and T2DM datasets and was absent from the population reference dataset; therefore, the variant with the lowest P-value in the GenDiP dataset is shown instead in the GDM and T2DM panels. Each variant's color represents its LD (r2) with the lead variant (purple diamond). The left y-axis shows recombination rate (cM/Mb), while the right y-axis displays -log10(P) values. Gene annotations appear below each plot. Blue shading in the background highlights recombination hotspots. Stacked points represent nearby variants (within ±500kb) with their respective association signals for GDM (top) and T2DM (bottom). LD; Linkage Disequilibrium. EUR; Europeans. EAS; East Asians.

**Supplementary Figure 110.** *Regional association plots showing colocalization between GDM and T2DM in EUR (right) and EAS (left) populations for the lead variant rs55872725 (FTO).* Each variant's color represents its LD (r2) with the lead variant (purple diamond). The left y-axis shows recombination rate (cM/Mb), while the right y-axis displays -log10(P) values. Gene annotations appear below each plot. Blue shading in the background highlights recombination hotspots. Stacked points represent nearby variants (within ±500kb) with their respective association signals for GDM (top) and T2DM (bottom). LD; Linkage Disequilibrium. EUR; Europeans. EAS; East Asians.

#### 8.4 No association with GDM or T2DM

**Supplementary Figure 111.** *Regional association plots showing colocalization between GDM and T2DM in EUR (right) and EAS (left) populations near the RNF144B locus.* The original lead variant rs142406759 was unavailable in different datasets across populations: in EUR, it was absent from the T2DM dataset and reference population; in EAS, it was absent from the GDM dataset, T2DM dataset, and reference population. Therefore, the variant with the lowest P-value from the GenDiP dataset is displayed instead in both the GDM and T2DM panels. Each variant's color represents its LD (r2) with the lead variant (purple diamond). The left y-axis shows recombination rate (cM/Mb), while the right y-axis displays -log10(P) values. Gene annotations appear below each plot. Blue shading in the background highlights recombination hotspots. Stacked points represent nearby variants (within ±500kb) with their respective association signals for GDM (top) and T2DM (bottom). LD; Linkage Disequilibrium. EUR; Europeans. EAS; East Asians.

**Supplementary Figure 112.** *Regional association plots showing colocalization between GDM and T2DM in EUR (right) and EAS (left) populations near the PRAME locus.* The original lead variant rs77090130 was unavailable in different datasets across populations: in EUR, it was absent from the reference population dataset; in EAS, it was absent from the GDM dataset, T2DM dataset, and reference population. Therefore, the variant with the lowest P-value from the GenDiP dataset is displayed instead in both the GDM and T2DM panels. Each variant's color represents its LD (r2) with the lead variant (purple diamond). The left y-axis shows recombination rate (cM/Mb), while the right y-axis displays -log10(P) values. Gene annotations appear below each plot. Blue shading in the background highlights recombination hotspots. Gaps in the EAS GDM plot reflect variants removed during quality control (MAF <0.01 and sample size <50% of total sample). Stacked points represent nearby variants (within ±500kb) with their respective association signals for GDM (top) and T2DM (bottom). LD; Linkage Disequilibrium. EUR; Europeans. EAS; East Asians.

#### 8.5 Shared causal variants for glycemic traits during and outside of pregnancy

**Supplementary Figure 113.** *Regional association plots showing colocalization between FG during and outside of pregnancy in EUR (right) and EAS (left) populations for the lead variant rs6547692 (GCKR).* Each variant's color represents its LD (r2) with the lead variant (purple diamond). The left y-axis shows recombination rate (cM/Mb), while the right y-axis displays -log10(P) values. Gene annotations appear below each plot. Blue shading in the background highlights recombination hotspots. Stacked points represent nearby variants (within ±500kb) with their respective association signals for FG during pregnancy (top) and FG outside of pregnancy (bottom). LD; Linkage Disequilibrium. FG; Fasting Glucose. EUR; Europeans. EAS; East Asians.

**Supplementary Figure 114.** *Regional association plots showing colocalization between FG during and outside of pregnancy in EUR (right) and EAS (left) populations for the lead variant rs1761879 (RFX6).* Each variant's color represents its LD (r2) with the lead variant (purple diamond). The left y-axis shows recombination rate (cM/Mb), while the right y-axis displays -log10(P) values. Gene annotations appear below each plot. Blue shading in the background highlights recombination hotspots. Stacked points represent nearby variants (within ±500kb) with their respective association signals for FG during pregnancy (top) and FG outside of pregnancy (bottom). LD; Linkage Disequilibrium. FG; Fasting Glucose. EUR; Europeans. EAS; East Asians.

**Supplementary Figure 115.** *Regional association plots showing colocalization between FG during and outside of pregnancy in EUR (right) and EAS (left) populations for the lead variant rs730497 (GCK).* Each variant's color represents its LD (r2) with the lead variant (purple diamond). The left y-axis shows recombination rate (cM/Mb), while the right y-axis displays -log10(P) values. Gene annotations appear below each plot. Blue shading in the background highlights recombination hotspots. Stacked points represent nearby variants (within ±500kb) with their respective association signals for FG during pregnancy (top) and FG outside of pregnancy (bottom). LD; Linkage Disequilibrium. FG; Fasting Glucose. EUR; Europeans. EAS; East Asians.

**Supplementary Figure 116.** *Regional association plots showing colocalization between FG during and outside of pregnancy in EUR (right) and EAS (left) populations for the lead variant rs3847554 (MTNR1B).* Each variant's color represents its LD (r2) with the lead variant (purple diamond). The left y-axis shows recombination rate (cM/Mb), while the right y-axis displays -log10(P) values. Gene annotations appear below each plot. Blue shading in the background highlights recombination hotspots. Stacked points represent nearby variants (within ±500kb) with their respective association signals for FG during pregnancy (top) and FG outside of pregnancy (bottom). LD; Linkage Disequilibrium. FG; Fasting Glucose. EUR; Europeans. EAS; East Asians.

**Supplementary Figure 117.** *Regional association plots showing colocalization between FG during and outside of pregnancy in EUR (right) and EAS (left) populations for the lead variant rs60397071 (KANK1).* Each variant's color represents its LD (r2) with the lead variant (purple diamond). The left y-axis shows recombination rate (cM/Mb), while the right y-axis displays -log10(P) values. Gene annotations appear below each plot. Blue shading in the background highlights recombination hotspots. Stacked points represent nearby variants (within ±500kb) with their respective association signals for FG during pregnancy (top) and FG outside of pregnancy (bottom). LD; Linkage Disequilibrium. FG; Fasting Glucose. EUR; Europeans. EAS; East Asians.

**Supplementary Figure 118.** *Regional association plots showing colocalization between FG during and outside of pregnancy in EUR (right) and EAS (left) populations for the lead variant rs6036158 (FOXA2).* Each variant's color represents its LD (r2) with the lead variant (purple diamond). The left y-axis shows recombination rate (cM/Mb), while the right y-axis displays -log10(P) values. Gene annotations appear below each plot. Blue shading in the background highlights recombination hotspots. Stacked points represent nearby variants (within ±500kb) with their respective association signals for FG during pregnancy (top) and FG outside of pregnancy (bottom). LD; Linkage Disequilibrium. FG; Fasting Glucose. EUR; Europeans. EAS; East Asians.

**Supplementary Figure 119.** *Regional association plots showing colocalization between 2hG during and outside of pregnancy in EUR (right) and EAS (left) populations for the lead variant rs67077402 (CDKAL1).* Each variant's color represents its LD (r2) with the lead variant (purple diamond). The left y-axis shows recombination rate (cM/Mb), while the right y-axis displays -log10(P) values. Gene annotations appear below each plot. Blue shading in the background highlights recombination hotspots. Stacked points represent nearby variants (within ±500kb) with their respective association signals for 2hG during pregnancy (top) and FG outside of pregnancy (bottom). LD; Linkage Disequilibrium. 2hG; 2-hour glucose after oral-glucose tolerance test (OGTT). EUR; Europeans. EAS; East Asians.

**Supplementary Figure 120.** *Regional association plots showing colocalization between 2hG during and outside of pregnancy in EUR (right) and EAS (left) populations for the lead variant rs4506565 (TCF7L2).* Each variant's color represents its LD (r2) with the lead variant (purple diamond). The left y-axis shows recombination rate (cM/Mb), while the right y-axis displays -log10(P) values. Gene annotations appear below each plot. Blue shading in the background highlights recombination hotspots. Stacked points represent nearby variants (within ±500kb) with their respective association signals for 2hG during pregnancy (top) and FG outside of pregnancy (bottom). LD; Linkage Disequilibrium. 2hG; 2-hour glucose after oral-glucose tolerance test (OGTT). EUR; Europeans. EAS; East Asians.

**Supplementary Figure 121.** *Regional association plots showing colocalization between HbA1c during and outside of pregnancy in EUR (right) and EAS (left) populations for the lead variant rs7758845 (HBS1L).* Each variant's color represents its LD (r2) with the lead variant (purple diamond). The left y-axis shows recombination rate (cM/Mb), while the right y-axis displays -log10(P) values. Gene annotations appear below each plot. Blue shading in the background highlights recombination hotspots. Stacked points represent nearby variants (within ±500kb) with their respective association signals for HbA1c during pregnancy (top) and FG outside of pregnancy (bottom). LD; Linkage Disequilibrium. EUR; Europeans. EAS; East Asians.

#### 8.6 Different causal variants for glycemic traits during and outside of pregnancy

**Supplementary Figure 122.** *Regional association plots showing colocalization between FG during and outside of pregnancy in EUR (right) and EAS (left) populations for the lead variant rs2685806 (ABCB11).* Each variant's color represents its LD (r2) with the lead variant (purple diamond). The left y-axis shows recombination rate (cM/Mb), while the right y-axis displays -log10(P) values. Gene annotations appear below each plot. Blue shading in the background highlights recombination hotspots. Gaps in the EAS FG during pregnancy plot reflect variants removed during quality control (MAF <0.01 and sample size <50% of total sample). Stacked points represent nearby variants (within ±500kb) with their respective association signals for FG during pregnancy (top) and FG outside of pregnancy (bottom). LD; Linkage Disequilibrium. FG; Fasting Glucose. EUR; Europeans. EAS; East Asians.

**Supplementary Figure 123.** *Regional association plots showing colocalization between 2hG during and outside of pregnancy in EUR (right) and EAS (left) populations for the lead variant rs2971670 (GCK).* Each variant's color represents its LD (r2) with the lead variant (purple diamond). The left y-axis shows recombination rate (cM/Mb), while the right y-axis displays -log10(P) values. Gene annotations appear below each plot. Blue shading in the background highlights recombination hotspots. Stacked points represent nearby variants (within ±500kb) with their respective association signals for 2hG during pregnancy (top) and FG outside of pregnancy (bottom). LD; Linkage Disequilibrium. 2hG; 2-hour glucose after oral-glucose tolerance test (OGTT). EUR; Europeans. EAS; East Asians.

#### 8.7 Association with glycemic traits only in pregnancy

**Supplementary Figure 124.** *Regional association plots showing colocalization between FG during and outside of pregnancy in EUR (right) and EAS (left) populations near the RBM47 locus.* In EUR, the lead variant rs35793823 is highlighted (purple diamond). In EAS, rs35793823 was not present in the GDM and T2DM datasets and was absent from the reference population dataset; therefore, the variant with the lowest P-value in the GenDiP dataset is shown instead in the FG during and outside of pregnancy panels. Each variant's color represents its LD (r2) with the lead variant (purple diamond). The left y-axis shows recombination rate (cM/Mb), while the right y-axis displays -log10(P) values. Gene annotations appear below each plot. Blue shading in the background highlights recombination hotspots. Stacked points represent nearby variants (within ±500kb) with their respective association signals for FG during pregnancy (top) and FG outside of pregnancy (bottom). LD; Linkage Disequilibrium. FG; Fasting Glucose. EUR; Europeans. EAS; East Asians.

**Supplementary Figure 125.** *Regional association plots showing colocalization between FG during and outside of pregnancy in EUR (right) and EAS (left) populations for the lead variant rs1683163 (IRAG2).* Each variant's color represents its LD (r2) with the lead variant (purple diamond). The left y-axis shows recombination rate (cM/Mb), while the right y-axis displays -log10(P) values. Gene annotations appear below each plot. Blue shading in the background highlights recombination hotspots. Stacked points represent nearby variants (within ±500kb) with their respective association signals for FG during pregnancy (top) and FG outside of pregnancy (bottom). LD; Linkage Disequilibrium. FG; Fasting Glucose. EUR; Europeans. EAS; East Asians.

**Supplementary Figure 126.** *Regional association plots showing colocalization between 2hG during and outside of pregnancy in EUR (right) and EAS (left) populations for the lead variant rs320370 (TENT5C).* Each variant's color represents its LD (r2) with the lead variant (purple diamond). The left y-axis shows recombination rate (cM/Mb), while the right y-axis displays -log10(P) values. Gene annotations appear below each plot. Blue shading in the background highlights recombination hotspots. Stacked points represent nearby variants (within ±500kb) with their respective association signals for 2hG during pregnancy (top) and FG outside of pregnancy (bottom). LD; Linkage Disequilibrium. 2hG; 2-hour glucose after oral-glucose tolerance test (OGTT). EUR; Europeans. EAS; East Asians.

**Supplementary Figure 127.** *Regional association plots showing colocalization between 2hG during and outside of pregnancy in EUR (right) and EAS (left) populations for the lead variant rs58682124 (CRHR2).* Each variant's color represents its LD (r2) with the lead variant (purple diamond). The left y-axis shows recombination rate (cM/Mb), while the right y-axis displays -log10(P) values. Gene annotations appear below each plot. Blue shading in the background highlights recombination hotspots. Stacked points represent nearby variants (within ±500kb) with their respective association signals for 2hG during pregnancy (top) and FG outside of pregnancy (bottom). LD; Linkage Disequilibrium. 2hG; 2-hour glucose after oral-glucose tolerance test (OGTT). EUR; Europeans. EAS; East Asians.

**Supplementary Figure 128.** *Regional association plots showing colocalization between 2hG during and outside of pregnancy in EUR (right) and EAS (left) populations for the lead variant rs4148646 (ABCC8).* Each variant's color represents its LD (r2) with the lead variant (purple diamond). The left y-axis shows recombination rate (cM/Mb), while the right y-axis displays -log10(P) values. Gene annotations appear below each plot. Blue shading in the background highlights recombination hotspots. Stacked points represent nearby variants (within ±500kb) with their respective association signals for 2hG during pregnancy (top) and FG outside of pregnancy (bottom). LD; Linkage Disequilibrium. 2hG; 2-hour glucose after oral-glucose tolerance test (OGTT). EUR; Europeans. EAS; East Asians.

#### 8.8 Different colocalization results in EAS and EUR

**Supplementary Figure 129.** *Regional association plots showing colocalization between FG during and outside of pregnancy in EUR (right) and EAS (left) populations for the lead variant rs4841132 (LOC157273).* Results from colocalization analyses suggest different causal variants for FG during and outside of pregnancy in Europeans, whilst an association only with FG during pregnancy was detected in the analyses with East Asians. Each variant's color represents its LD (r2) with the lead variant (purple diamond). The left y-axis shows recombination rate (cM/Mb), while the right y-axis displays -log10(P) values. Gene annotations appear below each plot. Blue shading in the background highlights recombination hotspots. Stacked points represent nearby variants (within ±500kb) with their respective association signals for FG during pregnancy (top) and FG outside of pregnancy (bottom). LD; Linkage Disequilibrium. FG; Fasting Glucose. EUR; Europeans. EAS; East Asians.

**Supplementary Figure 130.** *Regional association plots showing colocalization between FG during and outside of pregnancy in EUR (right) and EAS (left) populations for the lead variant rs4869273 (CAST-PSCK1).* Results from colocalization analyses suggest different causal variants for FG during and outside of pregnancy in Europeans, whilst an association only with FG during pregnancy was detected in the analyses with East Asians. Each variant's color represents its LD (r2) with the lead variant (purple diamond). The left y-axis shows recombination rate (cM/Mb), while the right y-axis displays -log10(P) values. Gene annotations appear below each plot. Blue shading in the background highlights recombination hotspots. Stacked points represent nearby variants (within ±500kb) with their respective association signals for FG during pregnancy (top) and FG outside of pregnancy (bottom). LD; Linkage Disequilibrium. FG; Fasting Glucose. EUR; Europeans. EAS; East Asians.

**Supplementary Figure 131.** *Regional association plots showing colocalization between FG during and outside of pregnancy in EUR (right) and EAS (left) populations for the lead variant rs7809775 (GRB10).* Results from colocalization analyses suggest different causal variants for FG during and outside of pregnancy in Europeans, whilst an association only with FG during pregnancy was detected in the analyses with East Asians. Each variant's color represents its LD (r2) with the lead variant (purple diamond). The left y-axis shows recombination rate (cM/Mb), while the right y-axis displays -log10(P) values. Gene annotations appear below each plot. Blue shading in the background highlights recombination hotspots. Stacked points represent nearby variants (within ±500kb) with their respective association signals for FG during pregnancy (top) and FG outside of pregnancy (bottom). LD; Linkage Disequilibrium. FG; Fasting Glucose. EUR; Europeans. EAS; East Asians.

**Supplementary Figure 132.** *Regional association plots showing colocalization between 2hG during and outside of pregnancy in EUR (right) and EAS (left) populations for the lead variant rs5030937 (HKDC1).* Results from colocalization analyses suggest different causal variants for FG during and outside of pregnancy in Europeans, whilst an association only with FG during pregnancy was detected in the analyses with East Asians. Each variant's color represents its LD (r2) with the lead variant (purple diamond). The left y-axis shows recombination rate (cM/Mb), while the right y-axis displays -log10(P) values. Gene annotations appear below each plot. Blue shading in the background highlights recombination hotspots. Stacked points represent nearby variants (within ±500kb) with their respective association signals for 2hG during pregnancy (top) and FG outside of pregnancy (bottom). LD; Linkage Disequilibrium. 2hG; 2-hour glucose after oral-glucose tolerance test (OGTT). EUR; Europeans. EAS; East Asians.

#### 8.9 No association with trait during or outside of pregnancy

**Supplementary Figure 133.** *Regional association plots showing colocalization between FG during and outside of pregnancy in EUR (right) and EAS (left) populations for the lead variant rs243020 (BCL11A).* Each variant's color represents its LD (r2) with the lead variant (purple diamond). The left y-axis shows recombination rate (cM/Mb), while the right y-axis displays -log10(P) values. Gene annotations appear below each plot. Blue shading in the background highlights recombination hotspots. Stacked points represent nearby variants (within ±500kb) with their respective association signals for FG during pregnancy (top) and FG outside of pregnancy (bottom). LD; Linkage Disequilibrium. FG; Fasting Glucose. EUR; Europeans. EAS; East Asians.

**Supplementary Figure 134.** *Regional association plots showing colocalization between 2hG during and outside of pregnancy in EUR (right) and EAS (left) populations for the lead variant rs4148646 (ABCC8).* Each variant's color represents its LD (r2) with the lead variant (purple diamond). The left y-axis shows recombination rate (cM/Mb), while the right y-axis displays -log10(P) values. Gene annotations appear below each plot. Blue shading in the background highlights recombination hotspots. Stacked points represent nearby variants (within ±500kb) with their respective association signals for 2hG during pregnancy (top) and FG outside of pregnancy (bottom). LD; Linkage Disequilibrium. 2hG; 2-hour glucose after oral-glucose tolerance test (OGTT). EUR; Europeans. EAS; East Asians.

**Supplementary Figure 135.** *Regional association plots showing colocalization between HbA1c during and outside of pregnancy in EUR (right) and EAS (left) populations near the lead variant rs147493281.* The original lead variant rs147493281 was unavailable in different datasets across populations: in both EUR and EAS, it was absent from the GDM dataset (passing the post-meta-analysis quality control step when including all ancestries, but removed after ancestry-specific meta-analyses as it was not present in more than half of the sample), T2DM dataset, and reference population. Therefore, the variant with the lowest P-value from the GenDiP dataset is displayed instead in both the GDM and T2DM panels. Each variant's color represents its LD (r2) with the lead variant (purple diamond). The left y-axis shows recombination rate (cM/Mb), while the right y-axis displays -log10(P) values. Gene annotations were not available for this region. Blue shading in the background highlights recombination hotspots. Stacked points represent nearby variants (within ±500kb) with their respective association signals for HbA1c during pregnancy (top) and FG outside of pregnancy (bottom). LD; Linkage Disequilibrium. EUR; Europeans. EAS; East Asians.

**Supplementary Figure 136.** *Regional association plots showing colocalization between HbA1c during and outside of pregnancy in EUR (right) and EAS (left) populations near the locus LINC00922.* The original lead variant rs368956520 was unavailable in different datasets across populations: in EUR, it was absent from the HbA1c dataset outside of pregnancy and reference population; in EAS, it was absent from the GDM and T2DM dataset, and reference population. Therefore, the variant with the lowest P-value from the GenDiP dataset is displayed instead in the panels. Each variant's color represents its LD (r2) with the lead variant (purple diamond). The left y-axis shows recombination rate (cM/Mb), while the right y-axis displays -log10(P) values. Gene annotations appear below each plot, *LINC00922* was not present in the gene list used. Blue shading in the background highlights recombination hotspots. Stacked points represent nearby variants (within ±500kb) with their respective association signals for HbA1c during pregnancy (top) and FG outside of pregnancy (bottom). LD; Linkage Disequilibrium. EUR; Europeans. EAS; East Asians.

**Supplementary Figure 137.** *Regional association plots showing colocalization between HbA1c during and outside of pregnancy in EUR (right) and EAS (left) populations for the lead variant rs146519004.* The original lead variant rs146519004 was unavailable in different datasets across populations: in EUR, it was absent from the reference population dataset; in EAS, it was absent from the GDM and T2DM dataset, and reference population. Therefore, the variant with the lowest P-value from the GenDiP dataset is displayed instead in the panels. Each variant's color represents its LD (r2) with the lead variant (purple diamond). The left y-axis shows recombination rate (cM/Mb), while the right y-axis displays -log10(P) values. Gene annotations appear below each plot. Blue shading in the background highlights recombination hotspots. Stacked points represent nearby variants (within ±500kb) with their respective association signals for HbA1c during pregnancy (top) and FG outside of pregnancy (bottom). LD; Linkage Disequilibrium. EUR; Europeans. EAS; East Asians.

### **9.0 GDM GRS associations with glucose levels during and after pregnancy**

**Supplementary Figure 138.** *Forest plot of genetic risk score (GRS) associations with fasting glucose levels (mmol/L) during (FgPreg) and after pregnancy (FgoutPreg).* Three GRS categories were analyzed: (1) GDM-predominant (G-GRS: 11 Class G variants), (2) T2DM-predominant (T-GRS: 13 Class T variants), and (3) combined (All-GRS: 37 variants including unclassified SNPs). All models were adjusted for principal components. Note: Not all cohorts had complete data for every SNP included in the GRS, nor data for every outcome. SNP; Single-nucleotide polymorphism. GRS; Genetic Risk Score. GDM; Gestational Diabetes Mellitus. T2DM; Type-II diabetes mellitus. AFR: African. EAS: East Asian. EUR: European. SAS: South Asian. AMR: Hispanic.

**Supplementary Figure 139.** *Forest plot of genetic risk score (GRS) associations with 2-hour glucose levels (mmol/L) during (2HgPreg) and after pregnancy (2HgoutPreg).* Three GRS categories were analyzed: (1) GDM-predominant (G-GRS: 11 Class G variants), (2) T2DM-predominant (T-GRS: 13 Class T variants), and (3) combined (All-GRS: 37 variants including unclassified SNPs). All models were adjusted for principal components. Note: Not all cohorts had complete data for every SNP included in the GRS, nor data for every outcome. SNP; Single-nucleotide polymorphism. GRS; Genetic Risk Score. GDM; Gestational Diabetes Mellitus. T2DM; Type-II diabetes mellitus. AFR: African. EAS: East Asian. EUR: European. SAS: South Asian. AMR: Hispanic.
