## Supplementary material for "Multi-ancestry, trans-generational GWAS meta-analysis of gestational diabetes and glycaemic traits during pregnancy reveals limited evidence of pregnancy-specific genetic effects": Cohort funding, approval and acknowledgents

### Cohort funding and acknowledgments

**ALSPAC**

We are extremely grateful to all the families who took part in this study, the midwives for their help in recruiting them, and the whole ALSPAC team, which includes interviewers, computer and laboratory technicians, clerical workers, research scientists, volunteers, managers, receptionists and nurses.

The UK Medical Research Council and Wellcome (Grant ref: 217065/Z/19/Z) and the University of Bristol provide core support for ALSPAC. A comprehensive list of grant funding is available on the ALSPAC website (<http://www.bristol.ac.uk/alspac/external/documents/grant-acknowledgements.pdf>). This publication is the work of the authors, and Gunn-Helen Moen and Caroline Brito Nunes will serve as guarantors for the contents of this paper. Genome-wide genotyping data were generated by Sample Logistics and Genotyping Facilities at Wellcome Sanger Institute and LabCorp (Laboratory Corporation of America) using support from 23andMe.

Please note that the study website contains details of all the data that is available through a fully searchable data dictionary and variable search tool (http://www.bristol.ac.uk/alspac/researchers/our-data/).

Ethical approval for the study was obtained from the ALSPAC Law and Ethics committee and local research ethics committees (NHS Haydock REC: 10/H1010/70). Consent for biological samples has been collected in accordance with the Human Tissue Act (2004). Informed consent for the use of data collected via questionnaires and clinics was obtained from participants following the recommendations of the ALSPAC Ethics and Law Committee at the time.

**ANDIS-MDC**

We thank the participants of the ANDIS-MDC study. The study was financed by Swedish governmental funding of clinical research (ALF), the Swedish Research Council project grant nos. 2020-02191, 2015-2558, infrastructure grant nos. 2010-5983, 2012-5538, 2014-6395, Linnaeus grant no. 349-2006-237, and strategic research grant nos. 2009-1039 (EXODIAB) and from the Swedish Foundation for Strategic Research IRC15-0067 (LUDC-IRC))..The study is supported by the EFSD/Lilly European Diabetes Research Programme and the Hjelt Foundation.

**BIB**

Our thanks to all the children (and parents), teachers and school staff who were involved in the research, and staff at the Bristol Bioresource Laboratory who manage the BiB biobank, including cataloguing and shipping samples. Thanks also to the Born in Bradford (BiB) Research Assistants for data collection. Born in Bradford is only possible because of the enthusiasm and commitment of the Children and Parents in BiB. We are grateful to all the participants, practitioners and researchers who have made Born in Bradford happen.

BiB is supported by a Wellcome Longitudinal Population Study Grant (223601/Z/21/Z); a joint grant from the UK Medical Research Council (MRC) and UK Economic and Social Science Research Council (ESRC) (MR/N024391/1); the British Heart Foundation (CS/16/4/32482); a Wellcome Infrastructure Grant (WT101597MA); the National Institute for Health Research under its Applied Research Collaboration for Yorkshire and Humber (NIHR200166). The National Institute for Health Research Clinical Research Network provided research delivery support for this study. Funding for the metabolomics analyses has been provided by the US National Institutes of Health [R01 DK10324], the European Research Council (ERC) under the European Union’s Seventh Framework Programme [FP7/2007-2013] / ERC grant agreement no 669545 and the MRC via the MRC Integrative Epidemiology Unit Programme to D.A.L [MC_UU_00011/6] provided funds for DNA extraction and genotyping, which was undertaken in the Bristol Bioresource laboratories, University of Bristol. The views expressed in this publication are those of the authors and not necessarily those of the National Institute for Health Research or the Department of Health and Social Care.

**BOTNIA**

The Botnia Study (L.G., T.T.) have been financially supported by grants from Folkhälsan Research Foundation, the Sigrid Juselius Foundation, The Academy of Finland (grant nos 263401, 267882, 312063, 336822 to L.G.; 312072 and 336826 to T.T.), University of Helsinki, Nordic Center of Excellence in Disease Genetics, EU (EXGENESIS), MOSAIC FP7-600914, Ollqvist Foundation, Swedish Cultural Foundation in Finland, Finnish Diabetes Research Foundation, Foundation for Life and Health in Finland, Signe and Ane Gyllenberg Foundation, Finnish Medical Society, Paavo Nurmi Foundation, State Research Funding via the Helsinki University Hospital, Perklén Foundation, Närpes Health Care Foundation and Ahokas Foundation. The study has also been supported by the Ministry of Education in Finland, Municipal Heath Care Center and Hospital in Jakobstad and Health Care Centers in Vasa, Närpes and Korsholm. The research leading to these results has received funding from the European Research Council under the European Union’s Seventh Framework Programme (FP7/2007-2013)/ERC grant agreement (n° 269045). The study is supported by the EFSD/Lilly European Diabetes Research Programme and the Hjelt foundation.

**CHILD, FAMILY and START cohort**

We express our sincere gratitude to all the participating families of the CHILD, FAMILY and START studies for their contribution, dedication, and commitment to advancing health research. We also thank all the study teams, including interviewers, nurses, computer and laboratory technicians, clerical workers, research scientists, volunteers, managers, and receptionists who made this work possible. We thank and acknowledge the members of the Nutrigen Alliance who are a founding co-PI of our studies or helped acquiring and/or working with the data including: Stephanie A. Atkinson; Meghan Azad; Allan B. Becker; Jeffrey Brook; Judah A Denburg; Dipika Desai; Russell J. de Souza; Milan K. Gupta; Michael Kobor; Diana L. Lefebvre; Wendy Lou; Piushkumar J. Mandhane; Sarah McDonald; Andrew Mente; David Meyre; Theo J. Moraes; Katherine Morrison; Guillaume Paré; Malcolm R. Sears; Elinor Simons; Padmaja Subbarao; Stuart E. Turvey; Julie Wilson; Salim Yusuf; Gita Wahi.

This work was supported by the Canadian Institutes for Health Research Team (CIHR) Grant [#MWG-146332], the Natural Sciences and Engineering Research Council of Canada and Genome Canada. The CHILD cohort study received base funding from the Allergy, Genes and Environment Network of Centres of Excellence (AllerGen NCE) and CIHR), Women's and Children's Health Research Institute provided core support for establishing the CHILD Study. The FAMILY cohort study was funded by CIHR, the Population Health Research Institute, and the McMaster Children’s Hospital Foundation. Dr. Anand is supported by a Tier 1 Canada Research Chair in Ethnicity and CVD and Michael G. DeGroote and Heart, Stroke Foundation Chair in Population Health, a grant from the Canadian Partnership Against Cancer, Heart and Stroke Foundation of Canada and Canadian Institutes of Health Research. Dr. Subbarao is supported by a Tier 1 Chair in Pediatric Asthma and Lung Health.

**deCODE**

The study was funded by deCODE genetics.

**EDEN**

We are extremely grateful to all the families who took part in this study, the midwives and psychologists for recruiting and following them, and the whole EDEN team, including research scientists, engineers, technicians and managers for their commitment and their role in the success of the study. We also express our sincere thanks to the participants of the EDEN study.

The EDEN study was supported by Foundation for medical research (FRM), National Agency for Research (ANR), National Institute for Research in Public health (IRESP: TGIR cohorte santé 2008 program), French Ministry of Health (DGS), French Ministry of Research, INSERM Bone and Joint Diseases National Research (PRO-A), and Human Nutrition National Research Programs, Paris-Sud University, Nestlé, French National Institute for Population Health Surveillance (InVS), French National Institute for Health Education (INPES), the European Union FP7 programmes (FP7/2007–2013, HELIX, ESCAPE, ENRIECO, Medall projects), Diabetes National Research Program (through a collaboration with the French Association of Diabetic Patients (AFD)), French Agency for Environme ntal Health Safety (now ANSES), Mutuelle Générale de l’Education Nationale a complementary health insurance (MGEN), French national agency for food security, French-speaking association for the study of diabetes and metabolism (ALFEDIAM).

The study received approval on 12 December 2002 from the ethics committee (CCPPRB, N°02-70) of Kremlin Bicêtre and from the Commission Nationale Informatique et Liberté (CNIL, n°902267), the French data privacy institution. Written informed consent was obtained twice from parents, once at enrolment and once after the child’s birth.

**EFSOCH**

The Exeter Family Study of Childhood Health (EFSOCH) was supported by South West NHS Research and Development, Exeter NHS Research and Development, the Darlington Trust and the National Institute for Health and Care Research Exeter Clinical Research Facility. This work was also supported by the National Institute for Health and Care Research Exeter Biomedical Research Centre. The views expressed are those of the authors and not necessarily those of the NIHR or the Department of Health and Social Care. Genotyping of the EFSOCH study samples was funded by the Wellcome Trust and Royal Society grant 104150/Z/14/Z.

**ELGH**

Genes & Health is/has recently been core-funded by Wellcome (WT102627, WT210561), the Medical Research Council (UK) (M009017, MR/X009777/1, MR/X009920/1), Higher Education Funding Council for England Catalyst, Barts Charity (845/1796), Health Data Research UK (for London substantive site), and research delivery support from the NHS National Institute for Health Research Clinical Research Network (North Thames). We acknowledge the support of the National Institute for Health and Care Research Barts Biomedical Research Centre (NIHR203330); a delivery partnership of Barts Health NHS Trust, Queen Mary University of London, St George’s University Hospitals NHS Foundation Trust and St George’s University of London

Genes & Health is/has recently been funded by Alnylam Pharmaceuticals, Genomics PLC; and a Life Sciences Industry Consortium of AstraZeneca PLC, Bristol-Myers Squibb Company, GlaxoSmithKline Research and Development Limited, Maze Therapeutics Inc, Merck Sharp & Dohme LLC, Novo Nordisk A/S, Pfizer Inc, Takeda Development Centre Americas Inc.

We thank Social Action for Health, Centre of The Cell, members of our Community Advisory Group, and staff who have recruited and collected data from volunteers. We thank the NIHR National Biosample Centre (UK Biocentre), the Social Genetic & Developmental Psychiatry Centre (King’s College London), Wellcome Sanger Institute, and Broad Institute for sample processing, genotyping, sequencing and variant annotation. This work uses data provided by patients and collected by the NHS as part of their care and support. This research utilised Queen Mary University of London’s Apocrita HPC facility, supported by QMUL Research-IT, http://doi.org/10.5281/zenodo.438045

We thank: Barts Health NHS Trust, NHS Clinical Commissioning Groups (City and Hackney, Waltham Forest, Tower Hamlets, Newham, Redbridge, Havering, Barking and Dagenham), East London NHS Foundation Trust, Bradford Teaching Hospitals NHS Foundation Trust, Public Health England (especially David Wyllie), Discovery Data Service/Endeavour Health Charitable Trust (especially David Stables), Voror Health Technologies Ltd (especially Sophie Don), NHS England (for what was NHS Digital) - for GDPR-compliant data sharing backed by individual written informed consent.

Most of all, we thank all of the volunteers participating in Genes & Health.

A favourable ethical opinion for the main Genes & Health research study was granted by NRES Committee London - South East (reference 14/LO/1240) on 16 Sept 2014. Queen Mary University of London is the Sponsor and Data Controller.

**EPIDG**

The authors thank all the women who participated in the EPIDG study. We are grateful to all researchers who have participated in the study, especially to MJ Picón-César, head of the Diabetes and Pregnancy Unit, for her invaluable contribution to patient recruitment and data collection, and to Teresa Linares-Pineda for all the laboratory work. The author gratefully acknowledges the IBIMA-BIONAND Platform's Bioinformatics Unit, as well as the computer resources (Picasso Supercomputer), technical expertise, and assistance provided by the SCBI (Supercomputing and Bioinformatics) Center of the University of Malaga.

EPIDG study has been supported by the Instituto de Salud Carlos III (ISCIII) through the projects PI18/01175, PI21/01864, and co-funded by the European Union; Servicio Andaluz de Salud, Junta de Andalucía (PI-0283-2018, PI-0419-2019). In addition, this study was supported by the Nicolas Monardes Program from the “Servicio Andaluz de Salud, Junta de Andalucía”, Spain (RC-0008-2021 to SM).

**EstBB**

The activities of the EstBB are regulated by the Human Genes Research Act, which was adopted in 2000 specifically for the operations of the EstBB. Individual-level data analysis in the EstBB was carried out under ethical approval 1.1-12/624 from the Estonian Committee on Bioethics and Human Research (Estonian Ministry of Social Affairs), using data according to release applications 3-10/GI/7064 and 3-10/GI/10790 from the Estonian Biobank. Data analysis was carried out in part in the High-Performance Computing Center of the University of Tartu.

This work was supported by the Estonian Research Council grant (PRG1911) and by the Ministry of Education and Research Centres of Excellence grant TK214 Centre of Excellence for Personalised Medicine. The project has received funding from the European Union’s Horizon Europe research and innovation programme under grant agreement No 101060011. Views and opinions expressed are, however, those of the author(s) only and do not necessarily reflect those of the European Union or European Research Executive Agency. Neither the European Union nor the granting authority can be held responsible for them.

**FinnGeDi**

The FinnGeDi cohort have received funding from: Research Council of Finland, Sigrid Jusélius Foundation, Emil Aaltonen Foundation, Juho Vainio Foundation, Foundation for Diabetes Research, Signe and Ane Gyllenberg Foundation, Yrjö Jahnsson Foundation, Novo Nordisk Foundation, Päivikki and Sakari Sohlberg Foundation, The Finnish Medical Foundation, Agence Nationale de la Recherche, France, Diabetes UK, United Kingdom, Suorsa Foundation, Otto A. Malm foundation and The Finnish Foundation for Cardiovascular Research.

**FinnGen**

We would like to thank all participants and investigators involved in the FinnGen project.

The FinnGen project has received funding from two Business Finland grants (HUS 4685/31/2016 and UH 4386/31/2016), as well as from a consortium of industry partners. These include AbbVie Inc., AstraZeneca UK Ltd, Biogen MA Inc., Bristol Myers Squibb (including Celgene Corporation and Celgene International II Sàrl), Genentech Inc., Merck Sharp & Dohme Corp., Pfizer Inc., GlaxoSmithKline Intellectual Property Development Ltd., Sanofi US Services Inc., Maze Therapeutics Inc., Janssen Biotech Inc., Novartis AG, and Boehringer Ingelheim International GmbH.

The project also acknowledges the contributions of the following biobanks for providing samples: Auria Biobank, THL Biobank, Helsinki Biobank, Biobank Borealis of Northern Finland, Finnish Clinical Biobank Tampere, Biobank of Eastern Finland, Central Finland Biobank, Finnish Red Cross Blood Service Biobank, Terveystalo Biobank, and Arctic Biobank. All these biobanks are part of the BBMRI.fi infrastructure, coordinated nationally by the Finnish Biobank Cooperative – FINBB. Access to Finnish biobank data is facilitated through the Fingenious® services platform, managed by FINBB.

Additional support for this research includes: A.E. was supported as a Sarnoff Cardiovascular Research Foundation Scholar, and E.W. received support through the Academy of Finland (grants #352796 and #363016).

**Gen3G**

Gen3G was funded by the Fonds de recherche du Québec—Santé (FRQS) operating grant (to M‑FH, grant #20697); the Canadian Institute of Health Research (CIHR) operating grants (to M‑FH grant #MOP 115071 and to LB #PJT152989, #IGH155183 and #PJT190076), Diabète Québec, Internal funding supports from le Centre de recherche du CHUS and l’Université de Sherbrooke, and grants from the National Institute of Health (NIH) (to MFH grant #R01HD094150) and from American Diabetes Association (to MFH #1-15-ACE-26).

**GIFTS**

The GIFTS and London Bangladeshi cohorts and genotyping were supported by the Diabetes Association of Bangladesh, MRC [Clinical Research Training Fellowship (G0800441)] and the European Union (FP7 EU grant: 83599025). Akhtar Hussain (deceased) and Abdul Kalam Azad Khan; Diabetes Association of Bangladesh. The authors are indebted to all the participants from Dhaka (Bangladesh) and London (UK).

**GUSTO**

We thank all cohort members and researchers who have participated in the study. This study is supported by the Singapore National Research Foundation under its Translational and Clinical Research (TCR) Flagship Program and administered by the Singapore Ministry of Health’s National Medical Research Council (NMRC), Singapore—NMRC/TCR/004-NUS/2008; NMRC/TCR/012-NUHS/2014. Additional funding is provided by the Institute for Human Development and Potential (IHDP) – Agency for Science, Technology and Research (A*STAR), Singapore.

**HAPO**

The HAPO Study and subsequent analyses were funded by NIH grants DK095963, DK117491, HD34242, HD34243, HG-004415 and R03CA211318.

**MoBa**

We thank the Norwegian Institute of Public Health (NIPH) for maintaining and distributing MoBa data. The work was supported by the Research Council of Norway through its Centres of Excellence funding scheme, project number 262700. This research is part of the HARVEST collaboration, supported by the Research Council of Norway (#229624). We also thank the NORMENT Centre for providing genotype data, funded by the Research Council of Norway (#223273 and #229624), South East Norway Health Authorities and Stiftelsen Kristian Gerhard Jebsen. We further thank the Center for Diabetes Research, funded by the University of Bergen, the European Research Council (AdG #293574), Stiftelsen Kristian Gerhard Jebsen, Trond Mohn Foundation, the Research Council of Norway (#240413 and #301178), the Novo Nordisk Foundation (grant #54741), the Western Norway Health Authorities (grant #912250). The Norwegian Mother, Father and Child Cohort Study is supported by the Norwegian Ministry of Health and Care Services and the Ministry of Education and Research, NIH/NIEHS (contract no N01-ES-75558), NIH/NINDS (grant no.1 UO1 NS 047537-01 and grant no.2 UO1 NS 047537-06A1). The participating families who contributed with data and biological material are gratefully acknowledged.

Computation:

Analyses on MoBa were performed using digital laboratories in HUNT Cloud at the Norwegian University of Science and Technology, Trondheim, Norway. We are grateful for outstanding support from the HUNT Cloud community.

Ethical agreements:

The establishment of MoBa and initial data collection was based on a license from the Norwegian Data Protection Agency and approval from The Regional Committees for Medical and Health Research Ethics. The MoBa cohort is currently regulated by the Norwegian Health Registry Act. The current study was approved by The Regional Committees for Medical and Health Research Ethics (West committee number 2012/67).

Data availability:

Data from the Norwegian Mother, Father and Child Cohort Study and the Medical Birth Registry of Norway used in this study are managed by the national health register holders in Norway (Norwegian Institute of public health) and can be made available to researchers, provided approval from the Regional Committees for Medical and Health Research Ethics (REK), compliance with the EU General Data Protection Regulation (GDPR) and approval from the data owners. The consent given by the participants does not open for storage of data on an individual level in repositories or journals. Researchers who want access to data sets for replication should apply through helsedata.no. Access to data sets requires approval from The Regional Committee for Medical and Health Research Ethics in Norway and an agreement with MoBa.

**MONN**

We wish to thank all participants and cohort members. The MONN study was supported by the National Natural Science Foundation of China (grants 32470642, 31900487), the Guangdong Natural Science Foundation (grant 2022B1515120080), and the Shenzhen Science and Technology Program (grant 20220818100717002 and ZDSYS20230626091203007).

**NFBC66**

We wish to thank all cohort members, researchers and NFBC project center personnel who participated in the NFBC data collections. NFBC1966 46y follow-up study received financial support from University of Oulu Grant no. 24000692, Oulu University Hospital Grant no. 24301140, ERDF European Regional Development Fund Grant no. 539/2010 A31592. Ethics: Northern Ostrobothnia Hospital District Ethical Committee 94/2011 (12.12.2011).

**NFBC86**

We wish to thank all cohort members, researchers and NFBC project center personnel who participated in the NFBC data collections. The follow-up study received financial support from the University of Oulu (Strategic funding from donations) and Oulu University Hospital (K65760). The oral health study was supported in part by the Research Council of Finland (former Academy of Finland, Grant no. 326189).

Ethics: Northern Ostrobothnia Hospital District Ethical Committee 108/2017 (15.1.2018)

**NFBC66&86**

NFBC data are available from the University of Oulu, Infrastructure for Population Studies. Permission to use the data can be applied for research purposes via an electronic material request portal. In the use of data, we follow the EU General Data Protection Regulation (679/2016) and the Finnish Data Protection Act. The use of personal data is based on a cohort participant’s written informed consent in their latest follow-up study, which may cause limitations to its use. Please, contact the NFBC project center (NFBCprojectcenter(at)oulu.fi) and visit the cohort website (www.oulu.fi/nfbc) for more information.

**PREDO**

The PREDO Study has been funded by the Academy of Finland (JL: 311617 and 269925, KR: 1312670 ja 128789 1287891), EraNet Neuron, EVO (a special state subsidy for health science research), University of Helsinki Research Funds, the Signe and Ane Gyllenberg foundation, the Emil Aaltonen Foundation, the Finnish Medical Foundation, the Jane and Aatos Erkko Foundation, the Novo Nordisk Foundation, the Päivikki and Sakari Sohlberg Foundation, Juho Vainio foundation, Yrjö Jahnsson foundation, The Finnish Society of Sciences and Letters, Jalmari and Rauha Ahokas foundation, Sigrid Juselius Foundation granted to members of the Predo study board. Methylation assays were funded by the Academy of Finland (269925). Dr. Lahti has received research support from the Strategic Research Council (SRC) established within the Academy of Finland (decision number: 352700).

The PREDO study would not have been possible without the dedicated contribution of the PREDO study group members: E Hamäläinen, E Kajantie, H Laivuori, PM Villa, A-K Pesonen, A Aitokallio-Tallberg, A-M Henry, VK Hiilesmaa, T Karipohja, R Meri, S Sainio, T Saisto, S Suomalainen-Konig, V-M Ulander, T Vaitilo (Department of Obstetrics and Gynaecology, University of Helsinki and Helsinki University Central Hospital, Helsinki, Finland), L Keski-Nisula, Maija-Riitta Orden (Kuopio University Hospital, Kuopio Finland), E Koistinen, T Walle, R Solja (Northern Karelia Central Hospital, Joensuu, Finland), M Kurkinen (Päijät-Häme Central Hospital, Lahti, Finland), P.Taipale. P Staven (Iisalmi Hospital, Iisalmi, Finland), J Uotila (Tampere University Hospital, Tampere, Finland). We thank all the PREDO children and their parents for their enthusiastic participation. We also thank all the research nurses, research assistants, and laboratory personnel involved in the Predo study.

**Roskilde**

Cohort was supported by a research grant from the Danish Diabetes Academy, supported by the Novo Nordisk Foundation. The study was approved by the Ethics Committee for Region Zealand (SJ-55 and SJ-347) and the Danish Data Protection Agency and was registered in ClinicalTrials.gov (Identifier: NCT00836524). The cohort would not have been possible without the dedicated work by Dorte Gybel-Brask (Department of Obstetrics, Roskilde hospital) and Estrid Hoegdall (Department of Pathology, Herlev hospital).

**SNUH**

Cohort was supported by a research grant (00-PJ3-PG6-GN07-001) from the Korea Health 21 R&D Project, Ministry of Health and Welfare, Republic of Korea. The study was supported by a National Research Foundation of Korea grant (RS-2023-00262002) funded by the Korean Ministry of Science and ICT.

**STORK**

GHM had a PhD grant from the South-Eastern Norway Regional Health Authority. The STORK study received additional funding from the Norwegian Diabetes Association, Oslo Diabetes Research Centre, the Norwegian Odd Fellow Research Fund, Johan Selmer Kvanes’ Endowment for Research in Diabetes, University of Oslo, Thematic research: Perinatal nutrition, Oslo University Hospital, Department of Obstetrics, the Norwegian Health Association and the Norwegian Extra Foundation for Health and Rehabilitation.

**STORK Groruddalen**

Data collection for the Stork Groruddalen study was funded by The Research Council of Norway, The South-Eastern Norway Regional Health Authority and the public Child Health Clinics in Stovner, Grorud and Bjerke districts in Oslo.

**UKBB**

This research has been conducted using the UK Biobank resource (Reference 53641). The UKBB has ethical approval from the North West Multi-Centre Research Ethics Committee (MREC), which covers the UK, and all participants provided written informed consent. This project received ethical approval from the Institutional Human Research Ethics Committee, University of Queensland (Approval Number 2019002705).

**Project Viva**

Project Viva has been funded by the National Institutes of Health (R01 HD034568). In addition, Project Viva acknowledges the contributions of Dr. Steven Zeisel, whose laboratory provided support for Project Viva genotyping.

### Individual funding and acknowledgments

GHM is the recipient of an Australian Research Council Discovery Early Career Award (Project number: DE220101226) funded by the Australian Government and supported by the Research Council of Norway (Project grant: 325640).

DME is supported by a National Health and Medical Research Council Investigator Award (2017942).

SL was supported by the National Natural Science Foundation of China (grants 32470642, 31900487), the Guangdong Natural Science Foundation (grant 2022B1515120080), and the Shenzhen Science and Technology Program (grant 20220818100717002)- the same funding for the MONN study.

SES was funded by the NNF Copenhagen Bioscience PhD Program (grant no. NNF18CC0033668). Novo Nordisk Foundation for supporting the Novo Nordisk Foundation Center for Basic Metabolic Research (grant nos. NNF18CC0034900 and NNF23SA0084103).

SK & JC are supported by the National Human Genome Research Institute (U01HG011723).

SM was supported by the Programa Nicolas Monardes del Servicio Andaluz de Salud, Junta de Andalucia, Spain (RC-0008-2021). MM-V was supported by Programa Juan Rodés from Instituto de Salud Carlos III (JR20-00040).

DAL, MCB and NM work in a unit supported by the University of Bristol and UK Medical Research Council (MC_UU_00032/5) and the British Heart Foundation (AA/18/7/34219). DAL’s contribution is also supported by the British Heart Foundation (CH/F/20/90003) and European Research Council under the European Union’s Horizon 2020 research and innovation program (grant agreements No 101021566).

JR was supported by the European Union’s Horizon 2020 (grant number 874739, LongITools).

EK is funded by the Research Council of Finland, Päivikki and Sakari Sohlberg Foundation.

MV is funded by Foundation for Diabetes Research, Yrjö Jahnsson Foundation, Signe and Ane Gyllenberg Foundation, Suorsa Foundation, The Finnish Foundation for Cardiovascular Research.

LB is a member of the CR-CHUS, a FRQS-funded Research Center.

PÉJ is a senior research scholar from the FRQS and a member of the CR-CHUS, a FRQS-funded Research Center.

AHC, RNB and RMF were supported by a Wellcome Senior Research Fellowship (WT220390). This research was funded in part by the Wellcome Trust [Grant number WT2209390]. For the purpose of open access, the authors have applied a CC BY public copyright licence to any Author Accepted Manuscript version arising from this submission.

AEH was supported by an NIHR-funded Academic Clinical Fellowship.

MV is supported by the Research Council of Norway and the University of Bergen (project #301178).

SJ is supported by grants Helse Vest's Open Research Grant (grants #912250 and F-12144), the Novo Nordisk Foundation (grant NNF19OC0057445), the Research Council of Norway (grant #315599), and the Medical Faculty at the University of Bergen.

PRN was funded by the ERC AdG project SELECTionPREDISPOSED #293574, Stiftelsen Kristian Gerhard Jebsen, Trond Mohn Foundation TMS2022TMT01, the RCN #240413, the Novo Nordisk Foundation #NNF18OC0054741, the University of Bergen, and the Western Norway Regional Health Authority.

AE received funding from Sarnoff Cardiovascular Research Foundation.

EW received funding from The Academy of Finland Center of Excellence program (352796).

SS, VK, AH work was supported by funding from the European Union under Horizon Europe projects STAGE (Grant agreement: 101137146); IHEN (Grant agreement: N°101137317, OBCT (Grant agreement: 101080250); OBELISK (Grant agreement: 101080465), TRIGGER (Grant agreement: 101057739); Horizon 2020 EARLYCAUSE (Grant agreement: 848158) and LongITools (Grant agreement: 356888) and the Research Council of Finland (Profi6 RCF 336449 and RCF 356888).

APM was supported by the Medical Research Council (MR/W029626/1), Versus Arthritis (grant 21754) and the NIHR Manchester Biomedical Research Centre. The views expressed are those of the authors and not necessarily those of the NIHR.

LC work was supported by funding from the European Union’s Horizon 2020 (grant number 874739, LongITools).

### Conflicts of Interest

KIB has received research support from AstraZeneca, Bayer, Boehringer Ingelheim, Lilly, MSD, Novo Nordisk, Roche, Sanofi and Sysmex Norway.

GT, VS and KS are employees of deCODE genetics, a subsidiary of Amgen.

SF and DvH receive research funding for Genes & Health from MRC, NIHR, Alnylam Pharmaceuticals, Takeda, Glaxo Smith Kline, Merck, Pfizer, NovoNordisk, Maze Pharmaceuticals, Bristol Myers Squibb.

The authors declare no competing interests.
